# Seroprevalence, prevalence, and genomic surveillance: monitoring the initial phases of the SARS-CoV-2 pandemic in Betim, Brazil

**DOI:** 10.1101/2021.10.21.21265140

**Authors:** Ana Valesca Fernandes Gilson Silva, Diego Menezes, Filipe Romero Rebello Moreira, Octavio Alcântara Torres, Paula Luize Camargos Fonseca, Rennan Garcias Moreira, Hugo José Alves, Vivian Ribeiro Alves, Tania Maria de Resende Amaral, Adriano Neves Coelho, Júlia Maria Saraiva Duarte, Augusto Viana da Rocha, Luiz Gonzaga Paula de Almeida, João Locke Ferreira de Araújo, Hilton Soares de Oliveira, Nova Jersey Claudio de Oliveira, Camila Zolini de Sá, Jôsy Hubner de Sousa, Elizângela Gonçalves de Souza, Rafael Marques de Souza, Luciana de Lima Ferreira, Alexandra Lehmkuhl Gerber, Ana Paula de Campos Guimarães, Paulo Henrique Silva Maia, Fernanda Martins Marim, Lucyene Miguita, Cristiane Campos Monteiro, Tuffi Saliba Neto, Fabricia Soares Freire Pugêdo, Daniel Costa Queiroz, Damares Nigia Alborguetti Cuzzuol Queiroz, Luciana Cunha Resende-Moreira, Franciele Martins Santos, Erika Fernanda Carlos Souza, Carolina Moreira Voloch, Ana Tereza Vasconcelos, Renato Santana de Aguiar, Renan Pedra de Souza

## Abstract

The Covid-19 pandemic has created an unprecedented need for epidemiological monitoring using diverse strategies. We conducted a project combining prevalence, seroprevalence, and genomic surveillance approaches to describe the initial pandemic stages in Betim City, Brazil. We collected 3239 subjects in a population-based age-, sex- and neighbourhood-stratified, household, prospective; cross-sectional study divided into three surveys 21 days apart sampling the same geographical area. In the first survey, overall prevalence (participants positive in serological or molecular tests) reached 0.46% (90% CI 0.12% – 0.80%), followed by 2.69% (90% CI 1.88% – 3.49%) in the second survey and 6.67% (90% CI 5.42% - 7.92%) in the third. The underreporting reached 11, 19.6, and 20.4 times in each survey, respectively. We observed increased odds to test positive in females compared to males (OR 1.88 95% CI 1.25 – 2.82), while the single best predictor for positivity was ageusia/ anosmia (OR 8.12, 95% CI 4.72 – 13.98). Thirty-five SARS-CoV-2 genomes were sequenced, of which 18 were classified as lineage B.1.1.28, while 17 were B.1.1.33. Multiple independent viral introductions were observed. Integration of multiple epidemiological strategies was able to describe Covid-19 dispersion in the city adequately. Presented results have helped local government authorities to guide pandemic management.

## 1. Introduction

Since its emergence in December 2019, the new human coronavirus has had a tremendous impact on humanity due to the pandemic nature of its infection, called Covid-19 [1]. The SARS-CoV-2 pathogen was described on January 24, 2020. In Brazil, the first case of Covid-19 was reported on February 26, 2020, in the city of São Paulo [2]. The virus spread rapidly, and the country had the highest number of cases and deaths in Latin America, experiencing its first peak wave in late July 2020. Although most cases were identified in the most prominent Brazilian cities, São Paulo and Rio de Janeiro, dispersion to other municipalities were quickly reported. Betim, a town located in the Minas Gerais State in Brazil with an estimated population of 439,340 in 2019, had its first reported SARS-CoV-2 case on March 23, 2020, in two patients returning from Europe. Two months later, on May 23, 2020, only 73 confirmed cases had been reported, although 4380 suspected cases were identified in public databases indicating limited testing availability.

Brazilian public healthcare system has prioritized testing subjects with symptoms due to scarce diagnostic tests, particularly in the early days of the pandemic. Since data suggest that symptomatic cases represent a fraction of persons infected with SARS-CoV-2, official statistics were expected to be underestimated [3]. Epidemiological surveillance using prevalence studies is needed to evaluate the true extent of SARS-CoV-2 dispersion, significantly extending testing to asymptomatic subjects. Combining serological and molecular tests may be a more robust strategy to uncover viral diffusion in a territory, avoiding each test’s kinetic detection limitations. Valid prevalence and seroprevalence estimates for a population rely on two major factors: (i) a representative population sample and (ii) accurate diagnostic testing [4].

While the epidemiological investigation is essential for controlling Covid-19, genomic surveillance is equally crucial. Robust SARS-CoV-2 variant monitoring can track viral evolution, detect new variants, describe patterns and clusters of transmission, outbreak tracking, among others. Therefore, it can provide actionable information on implementing a more targeted public health strategy that addresses local priorities through stakeholder engagement and mitigation efforts [5]. Here, we conducted a study combining seroprevalence, prevalence, and genomic surveillance approaches to understand the SARS-CoV-2 epidemic spread in Betim city.

## 2 Materials and Methods

### 2.1 Seroprevalence and prevalence

The Research Ethics Committee approved the present experiment under protocol CAAE 31459220.2.0000.5651. We conducted a population-based age-, sex- and neighbourhood-stratified, household, prospective; cross-sectional study repeated every 21 days in the same geographic area to determine the extent of SARS-CoV-2 transmission in Betim, Minas Gerais, Brazil. Three surveys were held: June 3-5, June 23-25, and July 13-15, 2020. The sample size (n = 1,080 each survey) was estimated considering dichotomous outcome (positive or negative), the population of 439,340 inhabitants, the confidence level of 90%, the maximum margin of error of 2.5%, and lack of a priori information on the prevalence of SARS-COV-2 in the municipality’s population (the latter represented by p = q = 0.5) and using the equation below:

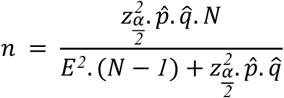

Random sampling was employed to ensure representativeness of the population, stratified by sex, age (0 to 5; 6 to 19; 20 to 39; 40 to 59 and 60 years or older) and city neighbourhoods (Centro, Alterosas, Imbiruçu, Norte, Teresópolis, PTB, Citrolândia, Vianópolis, Icaivera, and Petrovale). Every census tract (population stratum created by Governmental agencies) was sampled with at least one address. In case of refusal or closed households, the closest home was selected. Thirty-six teams (one driver, one nurse, and one community health worker) worked on active sampling subjects in 1080 addresses during three days. Clinical and epidemiological data were obtained using a questionnaire during interviews with participants or their legal guardians who signed the Informed Consent. Biological samples were collected using a nasal swab to conduct RT-PCR and capillary blood obtained by fingerstick for the serological test.

RT-PCR to detect SARS-CoV-2 RNA was initially conducted in pools of ten samples [6]. Whenever pools were positive, individual samples were examined. Molecular diagnosis was established according to the CDC 2019-Novel Coronavirus Real-Time RT-PCR Diagnostic Panel (N1, N2 and RNP primers). Serological tests were conducted using the SARS-CoV-2 Antibody Test (Guangzhou Wondfo Biotech Co., Ltd.) that detects IgM/IgG antibodies. The same test was used in a previous study in Brazil [7]. Reported sensitivity is 86.43% (95% CI: 82.41% ∼ 89.58%) and specificity 99.57% (95% CI: 97.63% ∼ 99.92%). We have validated antibody tests using serum samples from subjects who were SARS-CoV-2 positive confirmed with RT-PCR.

Associations of each variable of interest with surveys (Table 1) and positive status (Table 2) were assessed using chi-square tests. Odds ratios were estimated using logistic regression with the *glm* function. Spatial geostatistical modelling and prediction were carried out using the *gstat* and *predict* functions from the *gstat* package. All analyses were carried out in R software (version 4.1.1).

**Table 1:** Clinical and epidemiological data obtained from participants. Bolded p values indicate p < 0.05.

| Variable | Level | Overall n (%) | First survey n (%) | Second survey n (%) | Third survey n (%) | p-value |
| --- | --- | --- | --- | --- | --- | --- |
| Administrative Regions | Alterosas | 634 (19.6%) | 198 (18.4%) | 218 (20.2%) | 218 (20.2%) | 0.9584 |
|  | Citolândia | 219 (6.8%) | 83 (7.7%) | 68 (6.3%) | 68 (6.3%) |  |
|  | Icaivera | 62 (1.9%) | 20 (1.9%) | 21 (1.9%) | 21 (1.9%) |  |
|  | Imbiruçu | 565 (17.4%) | 183 (17.0%) | 191 (17.7%) | 191 (17.7%) |  |
|  | Norte | 333 (10.3%) | 111 (10.3%) | 111 (10.3%) | 111 (10.3%) |  |
|  | Petrovale | 41 (1.3%) | 13 (1.2%) | 14 (1.3%) | 14 (1.3%) |  |
|  | PTB | 290 (9.0%) | 108 (10.0%) | 91 (8.4%) | 91 (8.4%) |  |
|  | Sede | 583 (18.0%) | 201 (18.6%) | 191 (17.7%) | 191 (17.7%) |  |
|  | Terezópolis | 319 (9.8%) | 109 (10.1%) | 105 (9.7%) | 105 (9.7%) |  |
|  | Vianópolis | 193 (6.0%) | 53 (4.9%) | 70 (6.5%) | 70 (6.5%) |  |
| Sex | Female | 1628 (50.3%) | 548 (50.8%) | 536 (49.6%) | 544 (50.4%) | 0.8619 |
| Age range | 0 - 5 | 217 (6.7%) | 71 (6.6%) | 73 (6.8%) | 73 (6.8%) | 1.0000 |
|  | 6 - 19 | 650 (20.1%) | 218 (20.2%) | 217 (20.1%) | 215 (19.9%) |  |
|  | 20-39 | 1067 (32.9%) | 354 (32.8%) | 355 (32.9%) | 358 (33.1%) |  |
|  | 40-59 | 871 (26.9%) | 291 (27.0%) | 289 (26.8%) | 291 (26.9%) |  |
|  | Above 60 | 434 (13.4%) | 145 (13.4%) | 146 (13.5%) | 143 (13.2%) |  |
| Pneumopathy | Yes | 30 (0.9%) | 7 (0.6%) | 13 (1.2%) | 10 (0.9%) | 0.4042 |
| Chronic neurological disease | Yes | 39 (1.2%) | 16 (1.5%) | 10 (0.9%) | 13 (1.2%) | 0.4948 |
| Pregnant | Yes | 28 (0.9%) | 10 (0.9%) | 11 (1.0%) | 7 (0.6%) | 0.6257 |
| Postpartum | Yes | 9 (0.3%) | 2 (0.2%) | 3 (0.3%) | 4 (0.4%) | 0.7165 |
| Chronic cardiovascular disease | Yes | 96 (3.0%) | 34 (3.2%) | 39 (3.6%) | 23 (2.1%) | 0.1154 |
| Chronic kidney disease | Yes | 50 (1.5%) | 24 (2.2%) | 12 (1.1%) | 14 (1.3%) | 0.0799 |
| Obesity | Yes | 105 (3.2%) | 33 (3.1%) | 37 (3.4%) | 35 (3.2%) | 0.8903 |
| Asthma | Yes | 173 (5.3%) | 65 (6.0%) | 58 (5.4%) | 50 (4.6%) | 0.3537 |
| Immunodepression | Yes | 22 (0.7%) | 9 (0.8%) | 5 (0.5%) | 8 (0.7%) | 0.5507 |
| Chronic liver disease | Yes | 15 (0.5%) | 4 (0.4%) | 7 (0.6%) | 4 (0.4%) | 0.5478 |
| Diabetes | Yes | 228 (7.0%) | 78 (7.2%) | 74 (6.9%) | 76 (7.0%) | 0.9430 |
| Hypertension | Yes | 563 (17.4%) | 190 (17.6%) | 186 (17.2%) | 187 (17.3%) | 0.9698 |
| Transplanted | Yes | 4 (0.1%) | 2 (0.2%) | 1 (0.1%) | 1 (0.1%) | 0.7780 |
| Cancer | Yes | 23 (0.7%) | 10 (0.9%) | 8 (0.7%) | 5 (0.5%) | 0.4342 |
| Any comorbidity | Yes | 955 (29.5%) | 327 (30.3%) | 320 (29.6%) | 308 (28.5%) | 0.6552 |
| Fever | Yes | 224 (6.9%) | 66 (6.1%) | 70 (6.5%) | 88 (8.1%) | 0.1398 |
| Cough | Yes | 648 (20.0%) | 185 (17.1%) | 213 (19.7%) | 250 (23.1%) | **0.0022** |
| Sore throat | Yes | 397 (12.3%) | 112 (10.4%) | 125 (11.6%) | 160 (14.8%) | **0.0051** |
| Dyspnoea | Yes | 141 (4.4%) | 49 (4.5%) | 46 (4.3%) | 46 (4.3%) | 0.9336 |
| Myalgia | Yes | 284 (8.8%) | 74 (6.9%) | 99 (9.2%) | 111 (10.3%) | **0.0165** |
| Rhinorrhea | Yes | 717 (22.1%) | 205 (19.0%) | 240 (22.2%) | 272 (25.2%) | **0.0025** |
| Respiratory discomfort | Yes | 188 (5.8%) | 63 (5.8%) | 58 (5.4%) | 67 (6.2%) | 0.7084 |
| Nausea/ vomit | Yes | 120 (3.7%) | 37 (3.4%) | 39 (3.6%) | 44 (4.1%) | 0.7156 |
| Headache | Yes | 790 (24.4%) | 244 (22.6%) | 259 (24.0%) | 287 (26.6%) | 0.0936 |
| Prostration | Yes | 188 (5.8%) | 60 (5.6%) | 51 (4.7%) | 77 (7.1%) | 0.0523 |
| Diarrhea | Yes | 211 (6.5%) | 59 (5.5%) | 76 (7.0%) | 76 (7.0%) | 0.2336 |
| Conjunctivitis | Yes | 32 (1.0%) | 13 (1.2%) | 11 (1.0%) | 8 (0.7%) | 0.5478 |
| Ageusia/ anosmia | Yes | 101 (3.1%) | 30 (2.8%) | 30 (2.8%) | 41 (3.8%) | 0.2914 |
| Loss of voice | Yes | 56 (1.7%) | 18 (1.7%) | 13 (1.2%) | 25 (2.3%) | 0.1381 |
| Sought health assistance | Hospital | 138 (4.3%) | 41 (3.8%) | 41 (3.8%) | 56 (5.2%) | 0.1492 |
|  | Basic Health Unit | 129 (4.0%) | 42 (3.9%) | 41 (3.8%) | 46 (4.3%) |  |
|  | Emergency Care Unit | 127 (3.9%) | 38 (3.5%) | 35 (3.2%) | 54 (5.0%) |  |
|  | None | 2845 (87.8%) | 958 (88.8%) | 963 (89.2%) | 924 (85.6%) |  |
| Admitted to a health institution | Yes | 38 (1.2%) | 11 (1.0%) | 12 (1.1%) | 15 (1.4%) | 0.7085 |
| International travel | Yes | 14 (0.4%) | 10 (0.9%) | 4 (0.4%) | 0 (0.0%) | **0.0043** |
| Household contact with symptomatic person | Yes | 640 (19.8%) | 157 (14.6%) | 193 (17.9%) | 290 (26.9%) | **< 0.0001** |
| Sorological test | Reactive | 39 (1.2%) | 3 (0.3%) | 8 (0.7%) | 28 (2.6%) | **< 0.0001** |
|  | Non-reactive | 3200 (98.8%) | 1076 (99.7%) | 1072 (99.3%) | 1052 (97.4%) |  |
| PCR test | Detected | 84 (2.6%) | 2 (0.2%) | 22 (2.0%) | 60 (5.6%) | **< 0.0001** |
|  | Undetected | 3112 (96.1%) | 1035 (95.9%) | 1057 (98.0%) | 1020 (94.4%) |  |
|  | Indeterminate | 42 (1.3%) | 42 (3.9%) | 0 (0.0%) | 0 (0.0%) |  |
| Prevalence | Sorological reactive and/or PCR detected | 106 (3.3%) | 5 (0.5%) | 29 (2.7%) | 72 ( 6.7%) | **< 0.0001** |

**Table 2:**
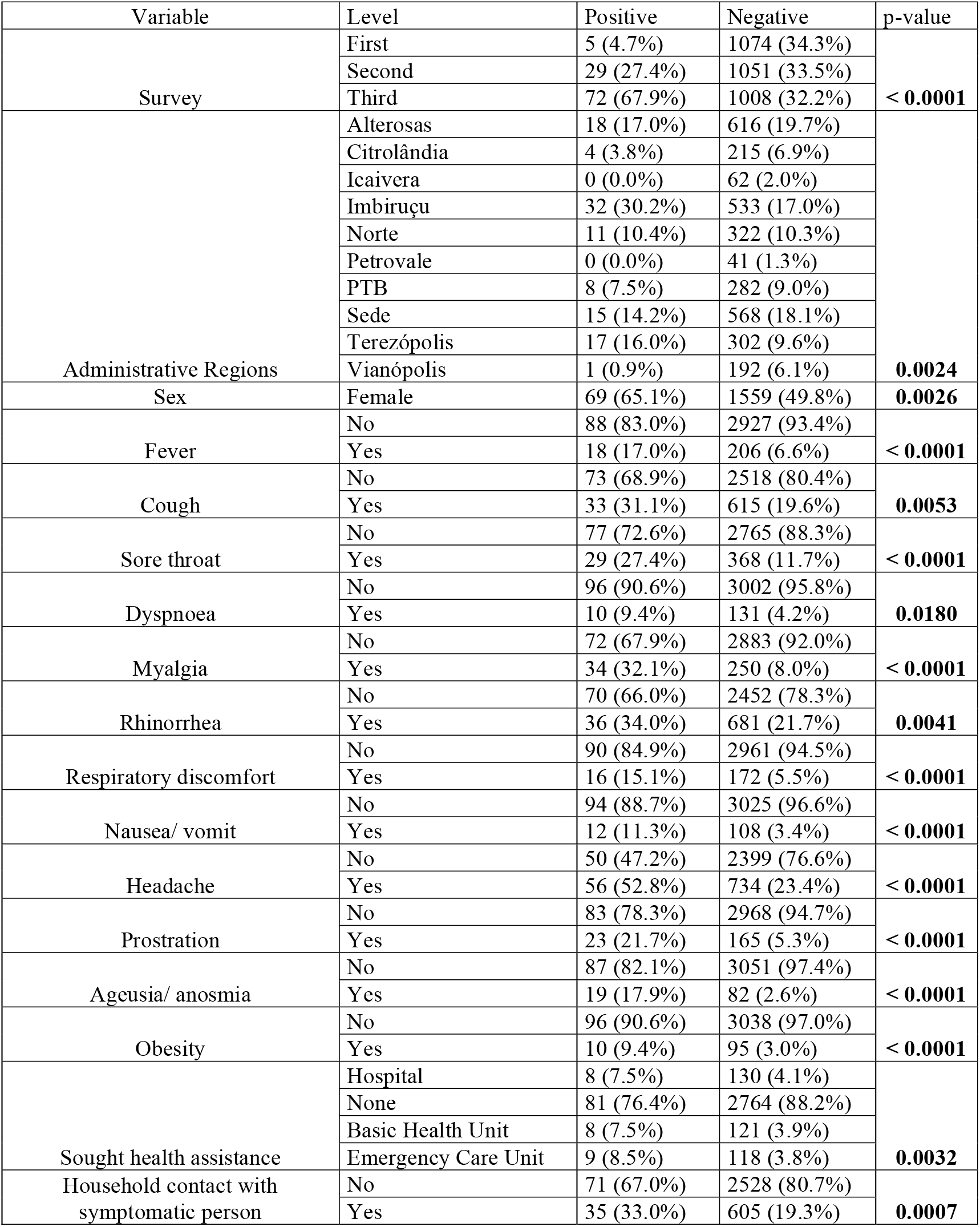
Significant associations of clinical and epidemiological data with positive test (serological or molecular). Non-significant associations are presented in Table S1. Bolded p values indicate p < 0.05.

| Variable | Level | Positive | Negative | p-value |
| --- | --- | --- | --- | --- |
| Survey | First | 5 (4.7%) | 1074 (34.3%) | **< 0.0001** |
|  | Second | 29 (27.4%) | 1051 (33.5%) |  |
|  | Third | 72 (67.9%) | 1008 (32.2%) |  |
| Administrative Regions | Alterosas | 18 (17.0%) | 616 (19.7%) | **0.0024** |
|  | Citrolândia | 4 (3.8%) | 215 (6.9%) |  |
|  | Icaivera | 0 (0.0%) | 62 (2.0%) |  |
|  | Imbiruçu | 32 (30.2%) | 533 (17.0%) |  |
|  | Norte | 11 (10.4%) | 322 (10.3%) |  |
|  | Petrovale | 0 (0.0%) | 41 (1.3%) |  |
|  | PTB | 8 (7.5%) | 282 (9.0%) |  |
|  | Sede | 15 (14.2%) | 568 (18.1%) |  |
|  | Terezópolis | 17 (16.0%) | 302 (9.6%) |  |
|  | Vianópolis | 1 (0.9%) | 192 (6.1%) |  |
| Sex | Female | 69 (65.1%) | 1559 (49.8%) | **0.0026** |
| Fever | No | 88 (83.0%) | 2927 (93.4%) | **< 0.0001** |
|  | Yes | 18 (17.0%) | 206 (6.6%) |  |
| Cough | No | 73 (68.9%) | 2518 (80.4%) | **0.0053** |
|  | Yes | 33 (31.1%) | 615 (19.6%) |  |
| Sore throat | No | 77 (72.6%) | 2765 (88.3%) | **< 0.0001** |
|  | Yes | 29 (27.4%) | 368 (11.7%) |  |
| Dyspnoea | No | 96 (90.6%) | 3002 (95.8%) | **0.0180** |
|  | Yes | 10 (9.4%) | 131 (4.2%) |  |
| Myalgia | No | 72 (67.9%) | 2883 (92.0%) | **< 0.0001** |
|  | Yes | 34 (32.1%) | 250 (8.0%) |  |
| Rhinorrhea | No | 70 (66.0%) | 2452 (78.3%) | **0.0041** |
|  | Yes | 36 (34.0%) | 681 (21.7%) |  |
| Respiratory discomfort | No | 90 (84.9%) | 2961 (94.5%) | **< 0.0001** |
|  | Yes | 16 (15.1%) | 172 (5.5%) |  |
| Nausea/ vomit | No | 94 (88.7%) | 3025 (96.6%) | **< 0.0001** |
|  | Yes | 12 (11.3%) | 108 (3.4%) |  |
| Headache | No | 50 (47.2%) | 2399 (76.6%) | **< 0.0001** |
|  | Yes | 56 (52.8%) | 734 (23.4%) |  |
| Prostration | No | 83 (78.3%) | 2968 (94.7%) | **< 0.0001** |
|  | Yes | 23 (21.7%) | 165 (5.3%) |  |
| Ageusia/ anosmia | No | 87 (82.1%) | 3051 (97.4%) | **< 0.0001** |
|  | Yes | 19 (17.9%) | 82 (2.6%) |  |
| Obesity | No | 96 (90.6%) | 3038 (97.0%) | **< 0.0001** |
|  | Yes | 10 (9.4%) | 95 (3.0%) |  |
| Sought health assistance | Hospital | 8 (7.5%) | 130 (4.1%) | **0.0032** |
|  | None | 81 (76.4%) | 2764 (88.2%) |  |
|  | Basic Health Unit | 8 (7.5%) | 121 (3.9%) |  |
|  | Emergency Care Unit | 9 (8.5%) | 118 (3.8%) |  |
| Household contact with symptomatic person | No | 71 (67.0%) | 2528 (80.7%) | **0.0007** |
|  | Yes | 35 (33.0%) | 605 (19.3%) |  |

### 2.2 Genomic surveillance

Whole viral genome amplification and DNA library preparation was carried out as described elsewhere [8]. Briefly, QIAseq SARS-CoV-2 Primer Panel - QIAGEN kit was used to amplify positive samples, following manufacturer instructions. In total, 39 of the 84 detectable samples were eligible for library preparation based on their CTs ≤ 30. Library concentration was measured using the QIAseq Library Quant Assay - QIAGEN kit, and the fragment integrity and size were evaluated using Bioanalyzer (Agilent Technologies, Waldbronn, DE). Sequencing was carried out on a MiSeq (Illumina, San Diego, CA, USA).

The raw data generated were filtered by Trimmomatic v0.39 [9], which trimmed low-quality bases (Phred score < 30) and removed short reads (<50 nucleotides) as well as adapters and primer sequences. Reads were then mapped against the SARS-CoV-2 reference genome (accession number: NC_045512.2) with Bowtie2 [10]. The resulting BAM files were manipulated with SAMtools, BCFtools [11], and BEDtools [12] to generate consensus genome sequences. Bases with less than 10x sequencing depth were masked. In total, 35 of the 39 genome sequences presented coverage greater than 79% and average sequencing depth greater than 200x. Sequencing metadata is available in **Table S1**. The 35 consensus genome sequences were submitted to the PANGOLIN 2.0 lineage classification tool (database version February 2, 2021) [13].

To confirm the PANGOLIN identification and further contextualize the diversity of lineages circulating in Betim, we performed a set of phylogenetic analyses. First, a global dataset was assembled from a subset of high-quality data available on GISAID and the newly generated genomes (*n* = 3,814). This dataset contained all Brazilian sequences and one per week for each country, as available on GISAID until January 12, 2021. These sequences were aligned with MAFFT v7.475[14], and a maximum likelihood tree was inferred on IQ-Tree 2 [15], under the GTR+F+I+G4 model [16], [17]. Shimoidara-Hasegawa approximate likelihood ratio test (SH-aLRT) was used to assess branches’ statistical support [18].

Two subsets of the previous dataset were assembled to explore the temporal dynamics of introduction and circulation of SARS-CoV-2 in Betim, comprehending sequences belonging to lineages B.1.1.28 (*n* = 258) and B.1.1.33 (*n* = 284). The parameterization of the phylogeographic model was set to be primarily informative concerning introductions of SARS-CoV-2 in Betim. Therefore, we set the model with six discrete categories: Betim City, Minas Gerais State, Rio de Janeiro State, São Paulo State, other Brazilian States, and foreign sequences. These locations were represented by 18, 2, 22, 71, 79, and 66 sequences in dataset B.1.1.28 while B.1.1.33 dataset composition was 17, 20, 53, 52, 73, and 69 sequences from each region, respectively.

Maximum likelihood trees were inferred from these datasets, and their temporal signal was evaluated with tempest v1.5.3 [19]. Time scaled phylogenies were then inferred from these datasets with BEAST v1.10.4 [20], using: (***i***) the HKY+I+G4 nucleotide substitution model [17], (***ii***) the strict molecular clock model, (***iii***) the non-parametric coalescent skygrid tree prior [21] and (***iv***) a symmetric discrete phylogeographic model [22]. A normal prior distribution (mean = 1.13 × 10^−3^; std= 5.1 × 10^−4^) on clock rate was assumed, based on a previous estimate [23]. The cutoff values of the skygrid tree prior were set based on the previously estimated dates for the emergence of each lineage [23]. The number of grids of the tree priors was set to match the approximate number of weeks comprehended between the estimated dates for lineages’ emergence and the dates of the most recently sampled sequences (41 weeks, both datasets). Two and three independent chains of 200 million generations sampling every 10,000 states were performed for datasets B.1.1.33 and B.1.1.28, respectively. Tracer v1.7.1 [24] was used to verify mixing and convergence of chains (effective sample size > 200 for all parameters), which were then combined with logcombiner v1.10.4 after 10% burning removal. Maximum clade credibility trees were generated with treeannotator v1.10.4. All logs and trees are available in https://github.com/LBI-lab/SARS-CoV-2_phylogenies.git.

## 3 Results

### 3.1 Seroprevalence and prevalence

**Table 1** presents clinical and epidemiological data obtained from participants. No significant difference was observed for the presence of any prior health condition across surveys (pneumopathy, chronic neurological disease, pregnant, postpartum, chronic cardiovascular disease, chronic kidney disease, obesity, asthma, immunodepression, chronic liver disease, diabetes, hypertension, transplanted, cancer or any comorbidity) indicating proper sampling was conducted since there was no reason to find significant differences in the period. Four symptoms (cough, sore throat, myalgia, and rhinorrhea) and contact with a symptomatic person increased while international travel decreased. Prevalence and seroprevalence increased across surveys.

Pandemic progression in Betim city is presented in **Figure 1**. Confirmed cases underestimation was found in all three surveys. In the first survey, overall prevalence (participants positive in serological or molecular tests) reached 0.46% (90% CI 0.12% – 0.80%), followed by 2.69% (90% CI 1.88% – 3.49%) in the second survey and 6.67% (90% CI 5.42% - 7.92%) in the third. The underreporting was obtained by the difference between survey prevalence and official data, and its magnitude reached 11, 19.6, and 20.4 times (distance between black dots and red curve in Figure 1B). Active transmission areas (RT-PCR positive participants) were observed increasing across time (**Figure 1C-E**). By the third survey, almost all populated city areas were likely to have viral circulation (**Figure 1E**). The same pattern of increase was observed in overall prevalence for most administrative regions (**Figure 1F-G**).

**Figure 1:**
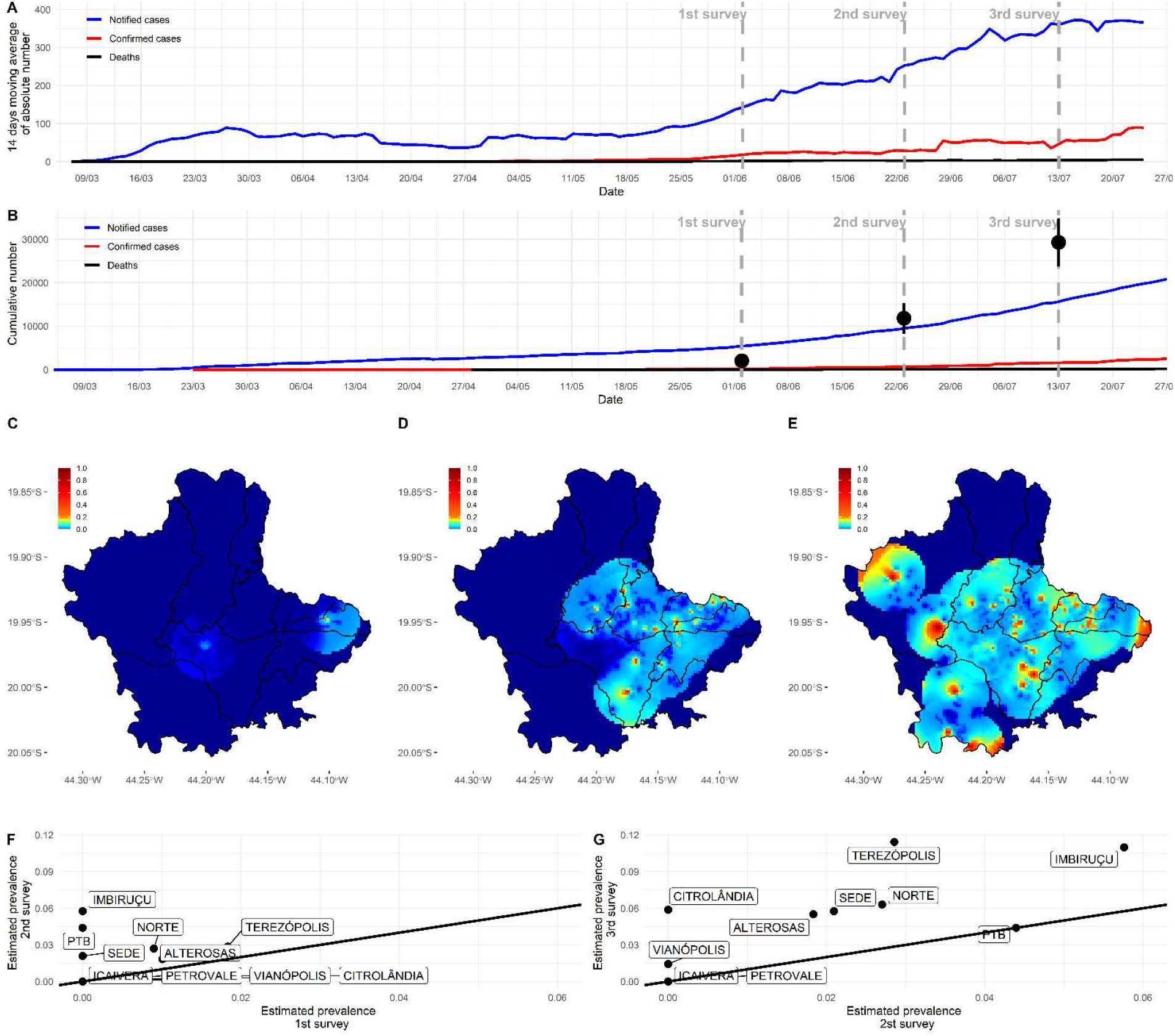
Covid-19 pandemic progression in Betim. (A) Absolute number of new cases according to official city statistics. (B) Cumulative number of cases according to official city statistics. Black dots indicate estimated overall prevalence (immunological and molecular tests) in the current study with its 95% confidence interval. Distance from black dots and red curve represent underreporting. (C-E) Dispersion of positive molecular tests across each survey. In the third survey (panel E), most populated areas of the city already had a non-null probability of presenting residents with a positive molecular test. (F-G) Overall prevalence (immunological and molecular tests) comparison in each of the ten administrative regions of the city across successive surveys. An increase was observed in most areas from the first to the second survey and, more substantially, from the second to the third survey.

We have also evaluated whether clinical and epidemiological variables were associated with molecular or serological test positivity (**Table 2**). Several significant results were observed, mostly with reported symptoms (fever, cough, sore throat, dyspnoea, myalgia, rhinorrhea, respiratory discomfort, nausea/ vomit, headache, prostration, ageusia/ anosmia). We also observed increased odds to test positive in females compared to males (OR 1.88 95% CI 1.25 – 2.82) and clear enrichment of positive cases in certain city regions (e.g., Imbiruçu and Terezópolis). Surprisingly, people with obesity were more likely to be positive (OR 3.33, 95% CI 1.68 – 6.59). The single best predictor for positivity was ageusia/ anosmia (OR 8.12, 95% CI 4.72 – 13.98). Non-significant results can be found in **Table S2**.

### 3.2 Genomic viral surveillance

In total, 35 novel SARS-CoV-2 genome sequences were obtained (GISAID EPI_ISL_5416087-5416121). The sequences were classified by PANGOLIN 2.0 to assess the genetic diversity of SARS-CoV-2 circulating in Betim. 18 of the 35 genomes were classified as lineage B.1.1.28, while 17 were B.1.1.33 (*Probability* = 1.0). Further, a maximum likelihood tree was inferred from the global dataset GISAID [25].

The analysis supported these results, revealing sequences from the Betim cluster within several clades of these lineages confirming the circulation of (B.1.1.28 and B.1.1.33 during the first wave of COVID-19 pandemics in the city (**Figure 2**). The spread of Betim sequences across the tree suggests multiple independent introductions occurred in the town. Further, eight clades majorly composed by Betim sequences were inferred with variable degrees of statistical support (median SH-aLRT = 82.75, range: 0 - 100), suggesting the occurrence of local transmission in the city after initial introduction events. In addition to these clusters, nine introductions supported by single sequences have also been detected. Most Betim sequences or clusters are closely related to sequences from Rio de Janeiro and São Paulo, two neighbouring States connected by highways to Minas Gerais. To formally assess the dynamics of introduction and spread of SARS-CoV-2 in Betim, separated datasets for lineages B.1.1.28 and B.1.1.33 were evaluated. Regression between sampling times and genetic distances revealed both datasets had moderate temporal signal (B.1.1.28: R^2^ = 0.49; B.1.1.33: R^2^ = 0.58), justifying molecular clock analysis.

**Figure 2:**
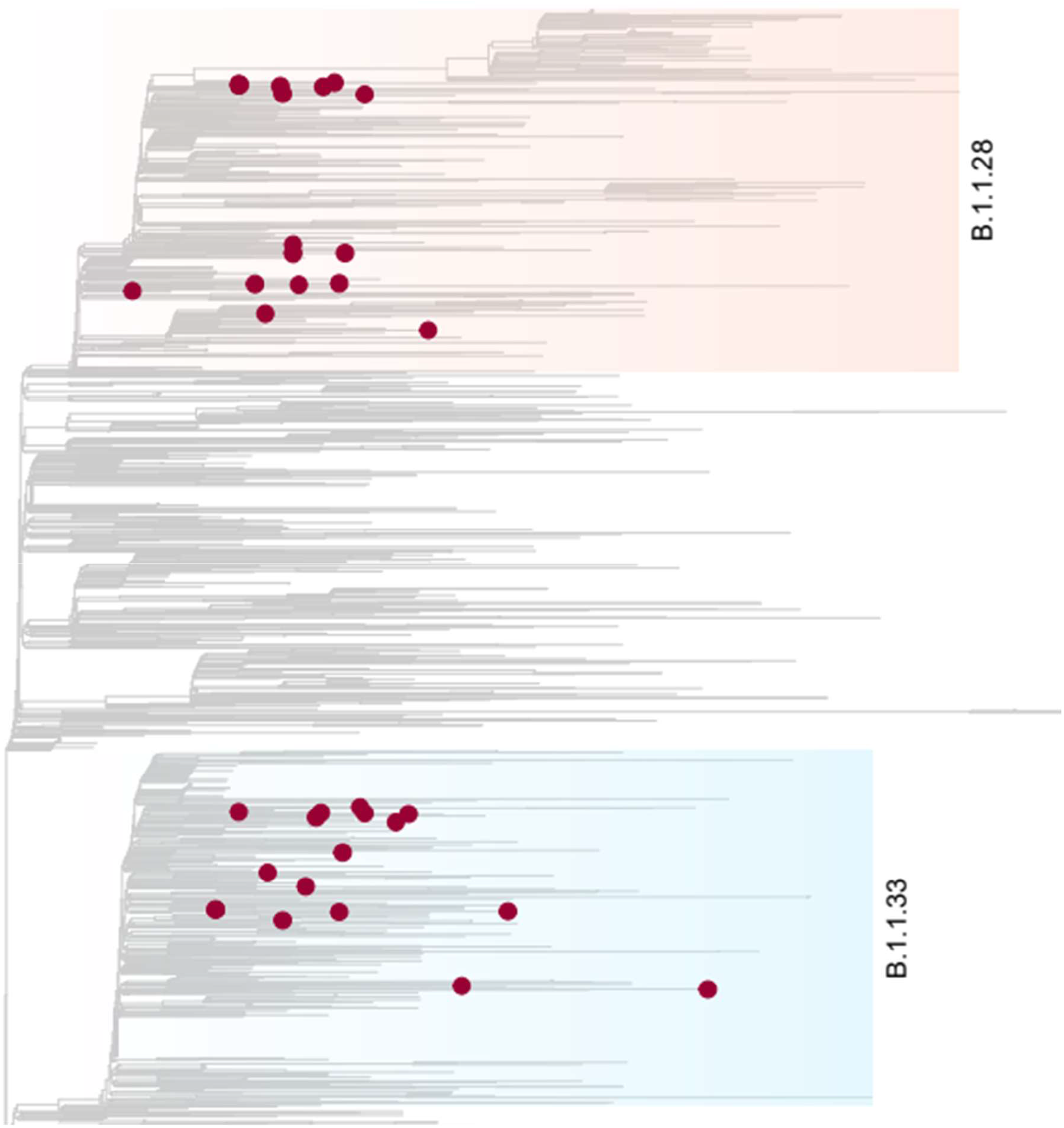
Phylogenetic characterization of SARS-CoV-2 genomes characterized in Betim. A maximum-likelihood tree was inferred on IQ-Tree under the GTR+F+I+G4 model with a comprehensive reference dataset, encompassing all Brazilian sequences plus one international sequence per country per week, from late 2019 to January 12 2021 (*n* = 3,814). The phylogeny depicted exhibits a subtree of 2,023 tips that harbours all relevant diversity considered for this study, mainly lineages B.1.1.28 (light salmon) and B.1.1.33 (light blue) where the novel genome sequences sparsely clustered. Tip shapes mark sequences characterized in this study. The scale bar indicates average nucleotide substitutions per site.

The time-scaled phylogeographic analysis performed with dataset B.1.1.28 suggests this lineage emerged on February 22, 2020, in São Paulo (95% highest posterior density, HPD: February 11, 2020 - March 05, 2020; geographic model posterior probability, PP = 0.91), later spreading to other Brazilian states (**Figure 3A**). The phylogeny reveals that two introduction events, dated between April 19, 2020 (95% HPD: April 17, 2020 - May 11, 2020) and April 22, 2020 (95% HPD: April 20, 2020 – May 27, 2020), led to the emergence of Betim clusters (harbouring between two and six sequences). Additionally, four introductions related to single sequences have been detected. The phylogeographic model suggests that three introductions occurred from another Brazilian region to Betim, in addition to other single events from RJ, another one from SP, and another from foreign sequences. All events presented high statistical support (PP > 92% for all events).

**Figure 3:**
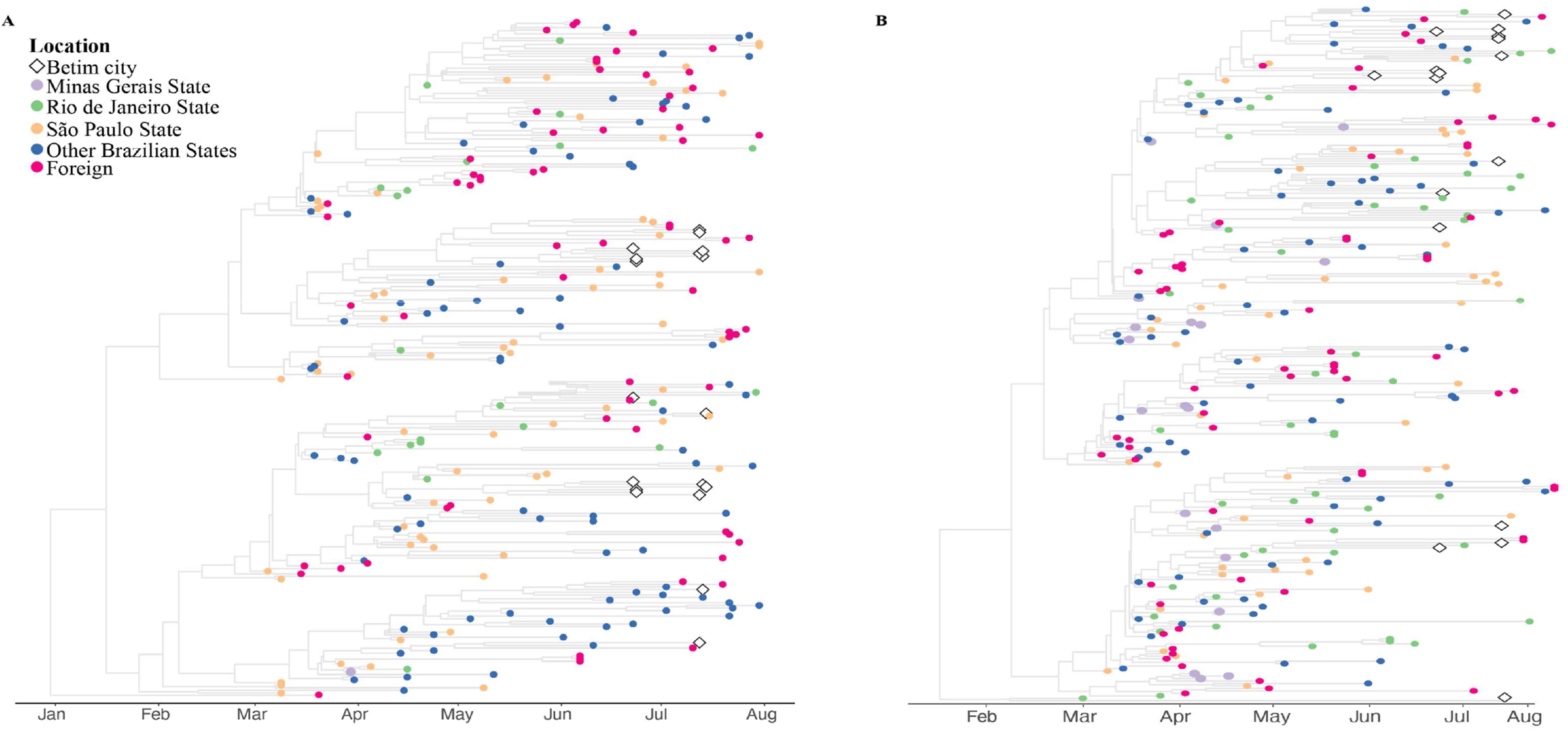
Spread of B.1.1.28 and B.1.1.33 lineages in Betim city. **(A)** Time-resolved maximum clade credibility phylogeny from a dataset comprehending 240 publicly available B.1.1.28 sequences and the 18 genomes generated in this study. **(B)** Time-resolved maximum clade credibility phylogeny from a dataset including 267 publicly available B.1.1.33 sequences and the 17 genomes generated in this study. For both analyses, the HKY+I+G4 nucleotide substitution model was used. The diamond indicates sequences from Betim city obtained in this study. The trees inferred are available on https://github.com/LBI-lab/SARS-CoV-2_phylogenies.git.

The phylogeographic reconstruction performed for dataset B.1.1.33 infers the origin of this lineage on February 06, 2020, in Rio de Janeiro (95% HPD: January 14, 2020 – February 25, 2020, PP = 0.78). The model supports the occurrence of many Betim clusters. One cluster comprises four sequences, dating to May 27, 2020 (95% HPD: May 01, 2020 - June 03, 2020) grouped with other sequences from other Brazilian regions and foreign. The model has also estimated eight introductions supported by single sequences. According to our phylogeny, the B.1.1.33 introductions came from different locations, such as the states of Rio de Janeiro, São Paulo, Minas Gerais, other Brazilian regions, and foreign sequences (PP > 0.81 for all events) (**Figure 3B**). The patterns reconstructed by both phylogeographic inferences are consistent, indicating the emergence of lineages B.1.1.28 and B.1.1.33 was followed by multiple importation events to diverse regions within the country, likely driven by human mobility. Additionally, evolutionary rate estimates also differed between datasets (B.1.1.28: 8.6372 × 10^−4^, 95% HPD: 7.8379 × 10^−4^ - 9.4559 × 10^−4^; B.1.1.33: 6.8743 × 10^−4^, 95% HPD 6.1784 × 10^−4^ - 7.5446 × 10^−4^).

## 4 Discussion

Betim is a medium-sized Brazilian city (439,340 inhabitants, 343 thousand square kilometres) crossed by national roads connecting major Brazilian cities and serving as a local hub for the Brazilian Public Health System. Understanding its pandemic dynamic may provide relevant information for municipalities with similar features. Here, we estimated the overall prevalence of active infections, seroprevalence and conducted genomic surveillance before the first pandemic wave in Betim. Brazilian molecular diagnostic capacity was insufficient in the first months of the pandemic [26]. Therefore, Covid-19 cases may have been included in the official statistics as severe acute respiratory infection cases with unknown aetiology. Data until May 2020 indicated a positive association between higher per-capita income and molecular Covid-19 diagnosis, while the severe acute respiratory infection cases with unknown aetiology were associated with lower per-capita income, suggesting a possible diagnosis bias related to economic status [27]. Inadequate diagnosis availability may lead to underreporting [28]. Our data estimated underreporting rates up to 20 times.

No studies have been conducted in Brazil evaluating active infection prevalence using adequate sampling. Our study design was inspired by previous research conducted in Santa Clara, USA, using pooled samples [29]. Pooled PCR tests were initially suggested to be used in asymptomatic people [6] and later were recommended for surveillance studies in populations with low infection prevalence [30]. Seroprevalence studies were conducted during the first wave in Brazil that peaked in July 2020. Two of the highest city seroprevalences reported during the period were Boa Vista (25.4% in June 2020) [7] and São Luiz (40.4% between the end of July and August 2020) [31], both in the northern area of the country. A nationwide survey carried out in May and June 2020 presented seroprevalence lower than two per cent during both surveys in all sampled cities neighbouring Betim (less than 200km), corroborating our findings [7]. Furthermore, seroprevalences higher than ten per cent were solely found in towns in the North Region [7]. In December 2020, Manaus, the largest city in the North Region, experienced a resurgence of Covid-19 [32] despite high seroprevalence [33], likely due to the gamma variant [34].

Previous seroprevalence studies have indicated ethnic and socioeconomic bias for SARS-CoV-2 infection in Brazil since the pandemic’s beginning [35], [36]. Results from Rio de Janeiro in April 2020 indicated that younger blood donors with lower education levels were more likely to test positive for SARS-CoV-2 antibodies [35]. A nationwide study revealed that the poorest quintile was 2.16 times more likely to test positive with the lowest risks among white, educated, and wealthy individuals [36]. Likewise, we found one of the highest prevalences in the poorest neighbourhood, Terezópolis, that include the largest slum of the city where more than 23 thousand people live.

Further modelling results showed higher infection rates among young adults, lower socioeconomic status, and people without healthcare access in the less developed North and Northeast areas until August 2020 [37]. Betim also presents most of its inhabitants with less than 59 years (90.7%), but no age effect was observed in the infectivity rates. Increased female infection odds were observed, although previous reports indicated a gender predisposition towards death in some Brazilian regions with higher male risk [38]. One possible explanation could be that 70% of the global health workforce are women [39] and a gender bias of pandemic perception and attitude [40].

Covid-19 diffusion presents strong socio-spatial determinants. Relocation diffusion from more- to less-developed regions and hierarchical diffusion from countries with higher population and density were relevant since early 2020 [41]. Data indicated a similar pattern in the São Paulo State with contiguous diffusion from the capital metropolitan area and hierarchical with long-distance spread through major highways that connects São Paulo city with cities of regional relevance [42]. Modelling results revealed that São Paulo city may have accounted for more than 85% of the initial case spread in the entire country [43]. Betim is directly connected to São Paulo city by a main national highway which may have contributed to Covid-19 diffusion.

Genomic surveillance is a powerful tool to elucidate viral dispersion patterns. The first sequencing work conducted in Brazil evaluated the first six positive individuals and reported the same predominant lineages found in Italy [44]. Later, a study with samples collected until late April 2020 from different country areas showed the dominance of clade B-derived lineages. At the national level, the respective frequency of these clades was seen in a 98.98%/1.02% ratio [23]. In Minas Gerais State, A lineages represented 2.5% of the infections, B.1 appeared in 92.5% of the samples, and B was responsible for 5% of the cases [45]. The exclusivity of lineages B.1.1.28 and B.1.1.33 circulating in Betim-MG from June to July 2020, given that multiple introductions from different country regions were demonstrated, is representative of the extent of these lineages’ dominance in the Brazilian scenario at the moment. Independent introductions also emphasize the importance of inter-state mobility barriers as a measure to control the epidemic.

Our study presents some limitations. First, the household survey is less likely to sample severe cases, thus underestimating symptomatic Covid-19. Second, all clinical data were self-reported, which may lead to reporting bias [46]. Third, we could not sequence all PCR positive samples due to the low viral load and sequencing technology employed. Nevertheless, our study shows the potential to integrate different epidemiological inquiries (prevalence, seroprevalence, and genomic surveillance) to describe pandemic dispersion adequately. Moreover, our findings present original and relevant evidence that has helped local government authorities to guide pandemic management.

## Data Availability

All data produced are available online at GISAID database

## Acknowledgement

We want to thank nurses, community health workers, drivers and management personnel who collaborated in this project. We also thank Mr. Guilherme Carvalho da Paixão for his support. We gratefully acknowledge the authors from the originating laboratories responsible for obtaining the specimens and the submitting laboratories where genetic sequence data were generated and shared via the GISAID Initiative, on which this research is based (**Table S3**).

---

**Table S1:**
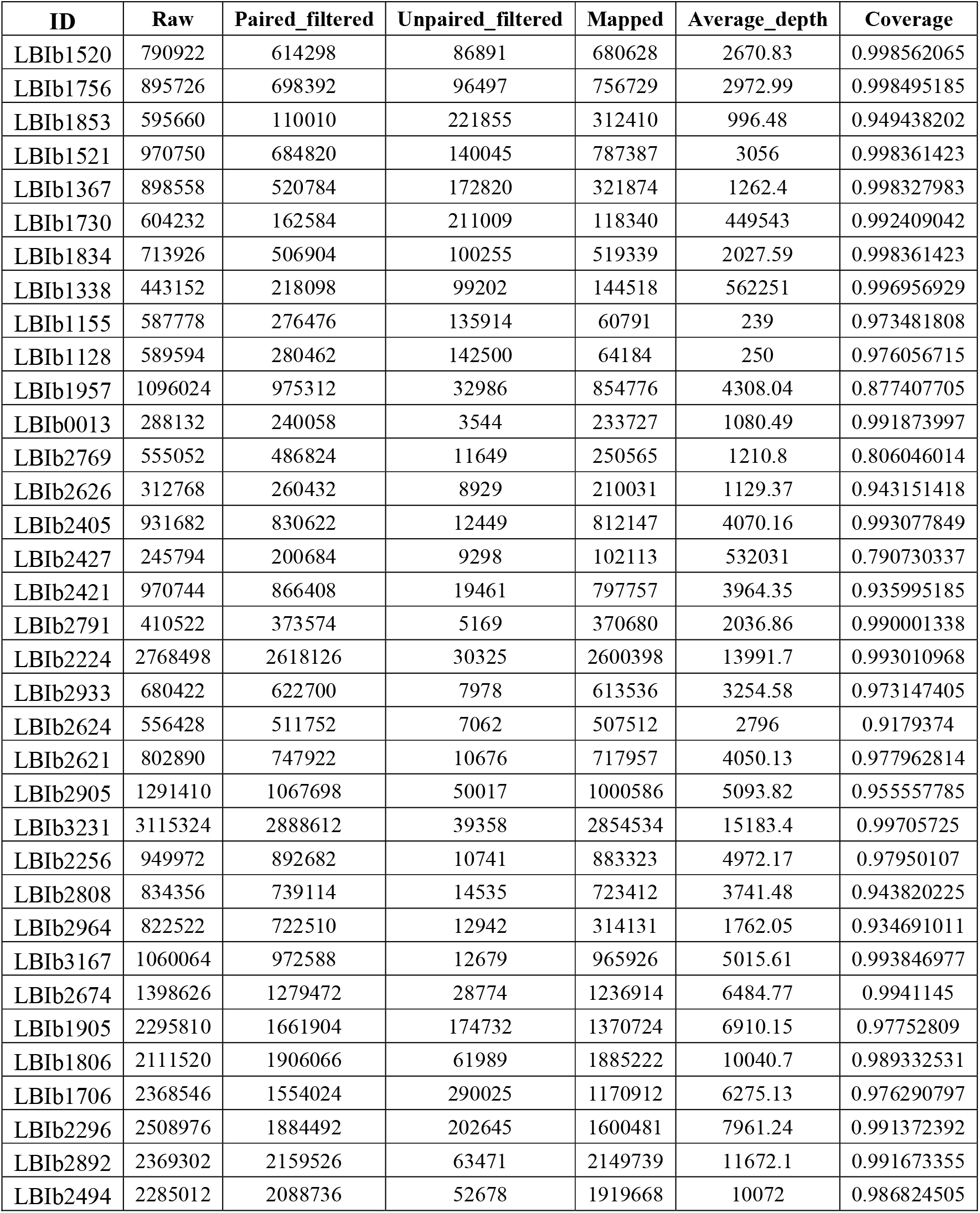
Sequencing statistics.

**Table S2:**
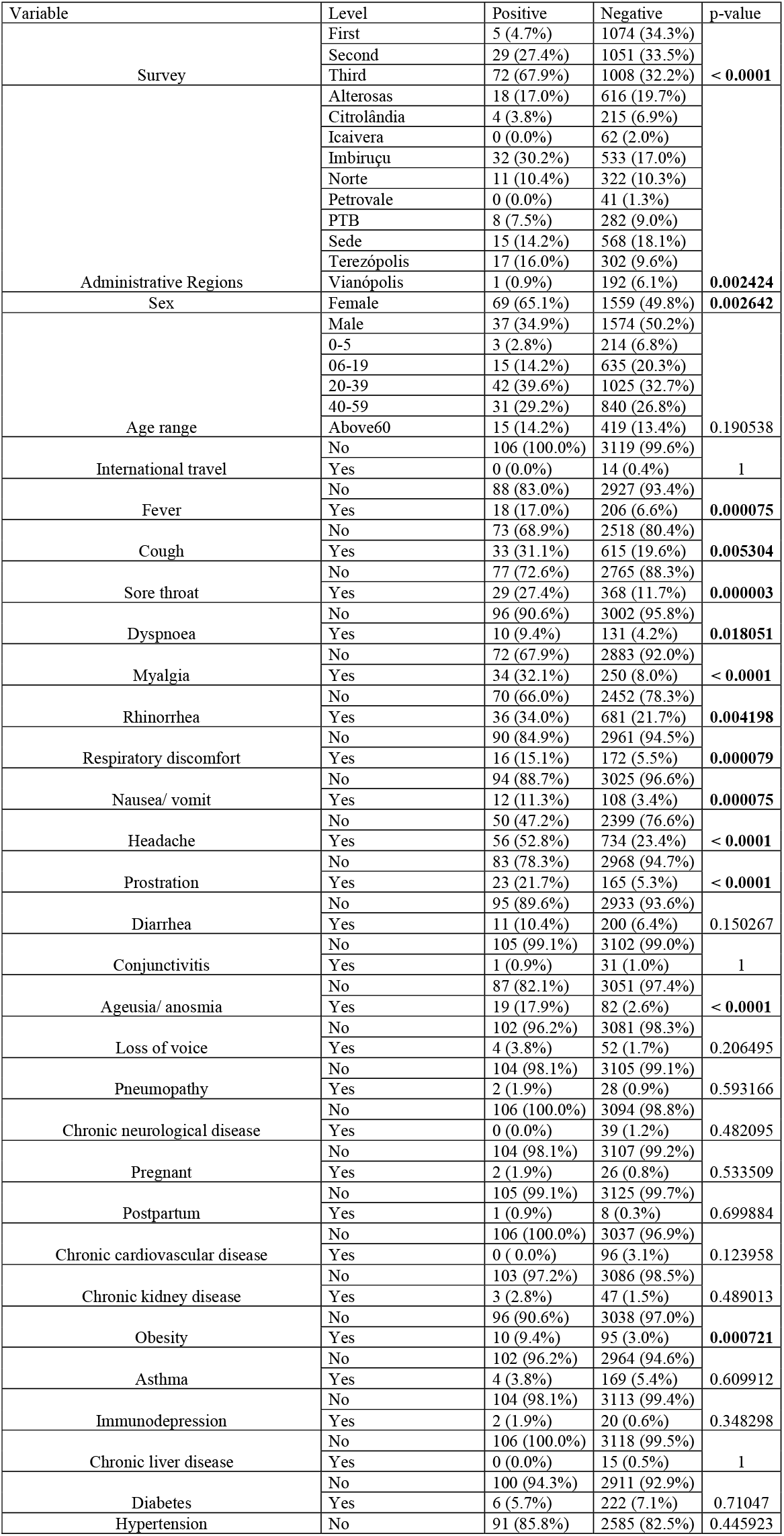
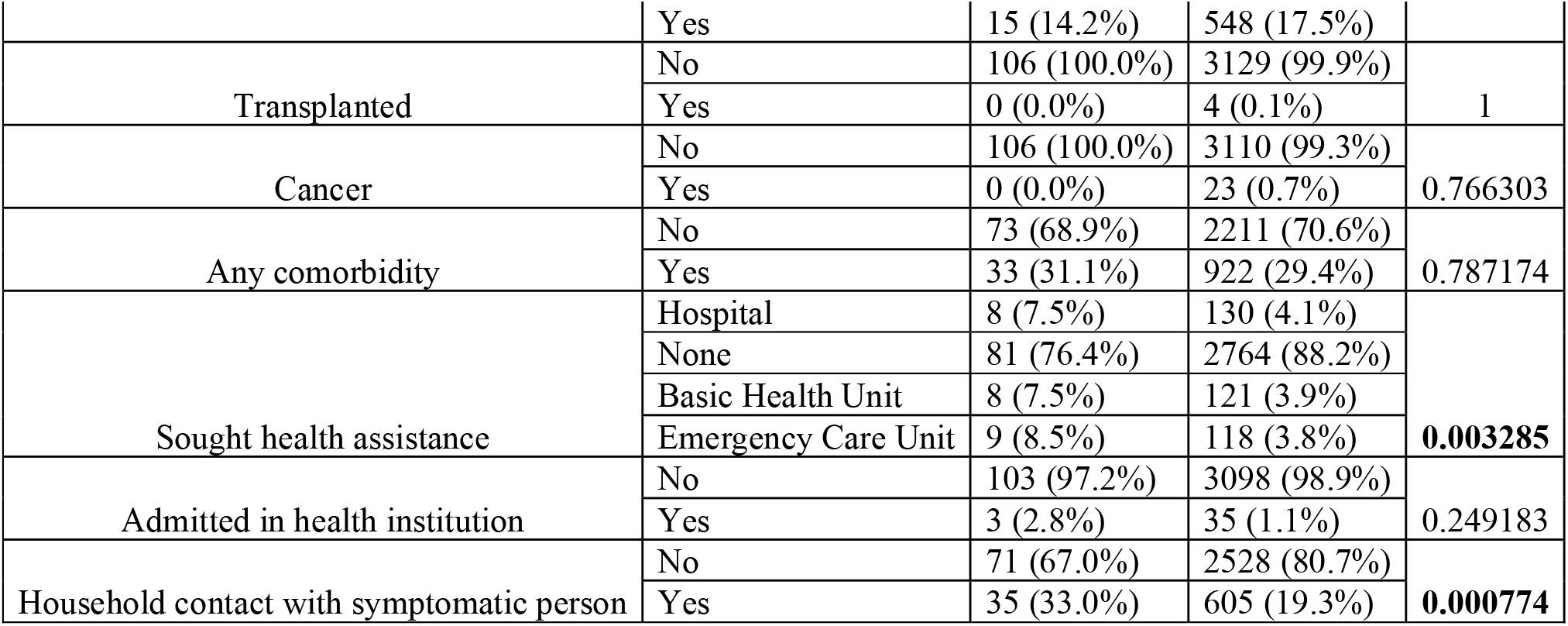
Association of clinical and epidemiological data with a positive test (serological or molecular). Bolded p values indicate p < 0.05.

**Table S3: Acknowledgement to sequences obtained from GSAID.**

