## Supplementary material for "Seroprevalence, prevalence, and genomic surveillance: monitoring the initial phases of the SARS-CoV-2 pandemic in Betim, Brazil": Table S3

We gratefully acknowledge the following Authors from the Originating laboratories responsible for obtaining the specimens, as well as the Submitting laboratories where the genome data were generated and shared via GISAID, on which this research is based.

All Submitters of data may be contacted directly via www.gisaid.org

Authors are sorted alphabetically.

| Accession ID | Originating Laboratory | Submitting Laboratory | Authors |
|---|---|---|---|
| EPI_ISL_683329, EPI_ISL_699657, EPI_ISL_707699, EPI_ISL_707791, EPI_ISL_707793, EPI_ISL_733499, EPI_ISL_733500, EPI_ISL_794736 | | | |
| see above | 1-Laboratory of Microbiology, National Reference Lab, Charles Nicolle Hospital; 2-University of Tunis ElManar, Faculty of Medicine of Tunis, LR99ES09, Tunis, Tunisia | 1-Clinical and Experimental Pharmacology Lab, LR16SP02, National Center of Pharmacovigilance, University of Tunis El Manar, Tunis, Tunisia. 2-Neurodegenerative diseases and psychiatric troubles, LR18SP03, Razi Hospital, University of Tunis El Manar, Tunis, Tunisia. 3- Ministry of Health, National Observatory of New and Emerging Diseases, 1006, Tunis, Tunisia | Alia Ben Kahla; Alia BenKahla; Asma Ferjani; Awatef El MOussi; Gaies Emna; Guedi Ali Barreh; Guedi Berrabeh; Habiba Ben Romdhane; Hanen El Jebari; Hanen ElJebari; Ilhem Boutiba-Ben Boubaker; Ines Mdini; Jalila Ben Khelil; Maher Kharrat; Mouna Ben Sassi; Mouna Safer; Nissaf Ben Alaya; Riadh Daghfous; Riadh Gouider.; Rouaa Ben Othman; Salma Abid; Salwa Mrabet; Sameh Trabelsi; Sana Ferjani; Sarra Chamman; Souissi Amira; Zaineb Hamzaoui |
| EPI_ISL_455444, EPI_ISL_455446, EPI_ISL_492069 | 1. ViroGenetics - BSL3 Laboratory of Virology, Małopolska Centre of Biotechnology, Jagiellonian University; 2. II Department of Internal Medicine, Faculty of Medicine, Jagiellonian University Medical College; 3. Narodowy Instytut Zdrowia Publicznego – Państwowy Zakład Higieny (NIZP-PZH) | 1. ViroGenetics - BSL3 Laboratory of Virology, Małopolska Centre of Biotechnology, Jagiellonian University; 2. II Department of Internal Medicine, Faculty of Medicine, Jagiellonian University Medical College; 3. Narodowy Instytut Zdrowia Publicznego – Państwowy Zakład Higieny (NIZP-PZH). | Agnieszka Kołakowska-Kulesza; Aleksandra A. Zasada; Aleksandra Milewska; Ewelina Hallman-Szelińska; Katarzyna Owczarek; Katarzyna Pancer; Katarzyna Zacharczuk; Krzysztof Pyrć; Magdalena Rzeczkowska; Marek Sanak; Natalia Wolaniuk; Paweł P Łabaj; Tomasz Wołkowicz; Wojciech Branicki |
| EPI_ISL_640078 | 2 Military Hospital wc MAA | NHLS/UCT | Arash Iranzadeh; Bruna Galvao; Carolyn Williamson; Deelan Doolabh; Diana Hardie; Innocent Mudau; Kruger Marais; Lynn Tyers; Marvin Hsiao; Stephen Korsman |
| EPI_ISL_729431, EPI_ISL_729463, EPI_ISL_729498, EPI_ISL_729562, EPI_ISL_729563, EPI_ISL_729566, EPI_ISL_729576, EPI_ISL_729585, EPI_ISL_729594, EPI_ISL_729679, EPI_ISL_729681 | | | |
| see above | A. Krumbholz, Labor Dr. Krause und Kollegen MVZ GmbH, Kiel | Charité Universitätsmedizin Berlin, Institut für Virologie | Barbara Mühlemann; Christian Drosten; Julia Schneider; Jörn Beheim-Schwarzbach; Talitha Veith; Terry Jones; Victor M Corman |
| EPI_ISL_775550, EPI_ISL_788995 | ABC | The Public Health Agency of Sweden | Department of Microbiology; The Public Health Agency of Sweden |
| EPI_ISL_801517, EPI_ISL_801519 | ABC Algarve | Instituto Nacional de Saude (INSA) | Borges et al |
| EPI_ISL_498523 | ACT Pathology | Schwessinger Lab | Ashley Jones; Benjamin Schwessinger; Craig Kennedy; Karina Kennedy; Kevin Murray; Megan McDonald; Ming-Dao Chia; Robert Lanfear; Robyn N Hall |
| EPI_ISL_768770, EPI_ISL_768784 | AIID | Irish Coronavirus Sequencing Consortium - National Virus Reference Laboratory | Alejandro Abner Garcia Leon; Gabriel Gonzalez; Michael Carr; Patrick Mallon |
| EPI_ISL_778829 | AIID | Irish Coronavirus Sequencing Consortium-Teagasc Grange | Aljandro Abner Garcia Leon; Calum Walsh; Fiona Crispie; Gabriel Gonzalez; John Kenny; Matthew McCabe; Michael Carr; Patrick Mallon; Paul Cotter |
| EPI_ISL_471546 | AMA DR Jose Soares Hungria | Instituto Adolfo Lutz, Interdiciplinary Procedures Center, Strategic Laboratory | Claudia Regina Gonçalves; Claudio Tavares Sacchi; Erica Valessa Ramos Gomes |
| EPI_ISL_523989 | AMA Jardim Joamar | Instituto Adolfo Lutz, Interdiciplinary Procedures Center, Strategic Laboratory | Claudia Regina Gonçalves; Claudio Tavares Sacchi; Erica Valessa Ramos Gomes |
| EPI_ISL_523990 | AMA Jardim Peri | Instituto Adolfo Lutz, Interdiciplinary Procedures Center, Strategic Laboratory | Claudia Regina Gonçalves; Claudio Tavares Sacchi; Erica Valessa Ramos Gomes |
| EPI_ISL_467467 | AMPATH-DBN | KRISP, KZN Research Innovation and Sequencing Platform | Chimukangara B; Giandhari J; Khan S; Lessells R; Mdlalose K; Pillay S; Tegally H; Wilkinson E; York D; de Oliveira T |
| EPI_ISL_458150 | ANOUAL | ANOUAL | Azami Nawfel; Benhida Rachid; Chenaoui Mohamed; El Aliani Aissam; El Ansari Fatima Zahra; Fekkak Jamal; Jouali Farah; Kasmi Yassine; Kitane Driss Lahlou; Loukman Salma; Marchoudi Nabila |
| EPI_ISL_682235 | AREA DE SALUD ALAJUELA NORTE - CLINICA DR. MARCIAL RODRIGUEZ | Inciensa, Instituto Costarricense de Investigación y Enseñanza en Nutrición y Salud | Adriana Godinez & Melany Calderon; Claudio Soto-Garita; Estela Cordero; Francisco Duarte; Hebleen Porras |
| EPI_ISL_682256 | AREA DE SALUD CORREDORES | Inciensa, Instituto Costarricense de Investigación y Enseñanza en Nutrición y Salud | Adriana Godinez; Claudio Soto-Garita; Estela Cordero; Francisco Duarte; Hebleen Porras; Melany Calderon & Mariel López |
| EPI_ISL_796764, EPI_ISL_796780 | ARS Algarve - Laboratorio Laura Ayres | Instituto Nacional de Saude (INSA) | Borges et al |
| EPI_ISL_542175, EPI_ISL_542178, EPI_ISL_542240, EPI_ISL_542438 | ASST GOM Niguarda | Dep. Of Oncology and Hemato-Oncology University of Milan | Antonio Piralla; Carlo Federico Perno; Chiara Vismara; Claudia Alteri; Elisa Matarazzo; Fausto Baldanti; Federica Giardina; Federica Novazzi; Luna Colagrossi; Maria Antonello; Massimo Puoti; Monica Tallarita; Oscar Massimiliano Epis; Roberto Fumagalli; Silvia Renica; Stefano Gaiarsa; Valentino Costabile; Valeria Cento |
| EPI_ISL_735501 | Abdul Malek Ukil Medical College, Noakhali | Central Biological Research Laboratory and Department of Biochemistry and Molecular Biology | H. M. Abdullah Al Masud; Imam Hossen; Md. Arif Hossain; Md. Imranul Hoq; Md. Khondakar Raziur Rahman; Md. Omer Faruq; Mohammad Omar Faruque; Robiul Hasan Bhuiyan; Sajib Rudra; Shanta Paul |
| EPI_ISL_517616, EPI_ISL_517621, EPI_ISL_517623, EPI_ISL_517625, EPI_ISL_517657, EPI_ISL_517659, EPI_ISL_518816 | | | |
| see above | Academic Hospital Paramaribo | Erasmus Medical Center | Bas Oude Munnink; Dion Gajadin; Ed Ijzerman; Emmanuelle Munger; Gary Gummels; Ingrid Krishnadath; Lycke Woittiez; Marion Koopmans; Mireille Van de Veer; Princes Wongsowidjojo; Radjesh Ori; Rohma Banwari; Stephen Vreden |
| EPI_ISL_528948 | Agenzia di Tutela della Salute di Bergamo | Istitituto Zooprofilattico Sperimentale dell'Abruzzo e Molise "G.Caporale" | Ancora M; Cammà C; Curini V; Di Domenico M; Di Pasquale A; Lorusso A; Mangone I; Marcacci M; Puglia I; Rinaldi A; Savini G. |
| EPI_ISL_420139, EPI_ISL_500782, EPI_ISL_549084, EPI_ISL_549171, EPI_ISL_590891 | Akershus University Hospital, Department for Microbiology and Infectious Disease Control | Norwegian Institute of Public Health, Department of Virology | Hilde Elshaug; Hilde Synnøve Vollan; Hilde Vollan; Kamilla Heddeland Instefjord; Karoline Bragstad; Kathrine Stene-Johansen; Olav Hungnes; Rasmus Riis Kopperud |
| EPI_ISL_661272 | Al-Quds Nutrition and Health Research Institute, Al-Quds University | Al-Quds Nutrition and Health Research Institute, Al-Quds University | A. and Al-Jawabreh, A.; Ereqat, S.; Nasereddin |
| EPI_ISL_769937 | Albany Medical Center Hospital Clinical Laboratories | Wadsworth Center, New York State Department.of Health | Alexis Russel; Daryl M. Lamson; Erasmus Schneider; Erica Lasek-Nesselquist; John Kelly; Jonathan Plitnick; Kirsten St. George; Matthew Shudt; Melissa A Leisner; Navjot Singh; Sara Griesemer |
| EPI_ISL_528696, EPI_ISL_528697, EPI_ISL_528707, EPI_ISL_528708, EPI_ISL_528715 | Alsafar - Khalifa University Abu Dhabi | Alsafar - Khalifa University Abu Dhabi | Andreas Henschel; Ernesto Damiani; Gihan Daw Elbait; Guan Tay; Habiba Alsafar; Rifat Hamoudi; Samuel Feng |
| EPI_ISL_515544, EPI_ISL_523984, EPI_ISL_523986 | Ama Dr Jose Soares Hungria | Instituto Adolfo Lutz, Interdiciplinary Procedures Center, Strategic Laboratory | Claudia Regina Gonçalves; Claudio Tavares Sacchi; Erica Valessa Ramos Gomes |
| EPI_ISL_569878 | Amedeo di savoia | Crosetto lab, Karolinska Institutet, SciLifeLab | Anna Sapino; Luuk Harbers; Maria Grazia Milia; Michele Simonetti; Nicola Crosetto; Ning Zhang; Valeria Ghisetti |
| EPI_ISL_717714 | Animal Health Centre, British Columbia Ministry of Agriculture | National Centre for Foreign Animal Disease, Canadian Food Inspection Agency | Asma Sultana; Brad Pickering; Oliver Lung; Peter Kruczkiewicz; Tomy Joseph |
| EPI_ISL_770030 | Area De Salud Buenos Aires | Inciensa, Instituto Costarricense de Investigación y Enseñanza en Nutrición y Salud | Adriana Godínez; Claudio Soto-Garita; Estela Cordero; Francisco Duarte; Hebleen Porras; Melany Calderón & Mariel López |
| EPI_ISL_769994, EPI_ISL_770026 | Area De Salud Catedral Noreste | Inciensa, Instituto Costarricense de Investigación y Enseñanza en Nutrición y Salud | Adriana Godínez; Claudio Soto-Garita; Estela Cordero; Francisco Duarte; Hebleen Porras; Melany Calderón & Mariel López |
| EPI_ISL_770008, EPI_ISL_770029 | Area De Salud Corredores | Inciensa, Instituto Costarricense de Investigación y Enseñanza en Nutrición y Salud | Adriana Godínez; Claudio Soto-Garita; Estela Cordero; Francisco Duarte; Hebleen Porras; Melany Calderón & Mariel López |
| EPI_ISL_512659 | Area De Salud Fortuna | Inciensa, Instituto Costarricense de Investigación y Enseñanza en Nutrición y Salud | Adriana Godinez & Melany Calderon; Claudio Soto-Garita; Estela Cordero; Francisco Duarte; Hebleen Porras |
| EPI_ISL_512667, EPI_ISL_770002 | Area De Salud La Cruz | Inciensa, Instituto Costarricense de Investigación y Enseñanza en Nutrición y Salud | Adriana Godinez & Melany Calderon; Adriana Godínez; Claudio Soto-Garita; Estela Cordero; Francisco Duarte; Hebleen Porras; Melany Calderón & Mariel López |
| EPI_ISL_512661 | Area De Salud Los Chiles | Inciensa, Instituto Costarricense de Investigación y Enseñanza en Nutrición y Salud | Adriana Godinez & Melany Calderon; Claudio Soto-Garita; Estela Cordero; Francisco Duarte; Hebleen Porras |
| EPI_ISL_770005 | Area De Salud Moravia | Inciensa, Instituto Costarricense de Investigación y Enseñanza en Nutrición y Salud | Adriana Godínez; Claudio Soto-Garita; Estela Cordero; Francisco Duarte; Hebleen Porras; Melany Calderón & Mariel López |
| EPI_ISL_770023 | Area De Salud San Francisco-San Antonio (Coopesana) | Inciensa, Instituto Costarricense de Investigación y Enseñanza en Nutrición y Salud | Adriana Godínez; Claudio Soto-Garita; Estela Cordero; Francisco Duarte; Hebleen Porras; Melany Calderón & Mariel López |
| EPI_ISL_770025 | Area De Salud San Juan-San Diego-Concepcion 2 | Inciensa, Instituto Costarricense de Investigación y Enseñanza en Nutrición y Salud | Adriana Godínez; Claudio Soto-Garita; Estela Cordero; Francisco Duarte; Hebleen Porras; Melany Calderón & Mariel López |
| EPI_ISL_491457 | Area de Salud Los Santos | Inciensa, Instituto Costarricense de Investigación y Enseñanza en Nutrición y Salud | Adriana Godinez & Melany Calderon; Claudio Soto-Garita; Estela Cordero; Francisco Duarte; Hebleen Brenes |
| EPI_ISL_678306, EPI_ISL_678365, EPI_ISL_678377 | Area of Virology, Serology and Virology Division (SAViD), New South Wales Health Pathology Randwick | Virology Research Laboratory; Area of Virology, Serology and Virology Division (SAViD), New South Wales Health Pathology Randwick | Au, J.; Bull, R.; Deveson, I.; Foster, C.; Rawlinson, W.; Ruiz Silva, M.; Van Hal, S. |
| EPI_ISL_539811, EPI_ISL_539821 | Asiaworld Expo Command Post | Hong Kong Department of Health | Alan K.L. Tsang; Dominic N.C. Tsang; Edman T.K. Lam; Peter C.W. Yip; Rickjason C.W. Chan |
| EPI_ISL_416519 | Auckland Hospital | Institute of Environmental Science and Research (ESR) | Erasmus Smit; Gary McAuliffe; Joep de Ligt; Lauren Jelly; Matt Storey; Matthew Blakiston; Sally Roberts; Xiaoyun Ren |

| | | | |
|---|---|---|---|
| EPI_ISL_583654, EPI_ISL_583674, EPI_ISL_583883, EPI_ISL_583885, EPI_ISL_583889 | Austrian Agency for Health and Food Safety (AGES) | Bergthaler laboratory, CeMM Research Center for Molecular Medicine of the Austrian Academy of Sciences | Adi Steinrigl; Alexander Lercher; Alexandra Popa; Andreas Bergthaler; Benedikt Agerer; Christian Paar; Christoph Bock; Daniela Schmid; Dorothee von Laer; Elisabeth Puchhammer-Stoeckl; Franz Allerberger; Gernot Walder; Gregor Hörmann; Guenter Weiss; Gunther Vogl; Henrique Colaco; Jakob-Wendelin Genger; Jan Laine; Judith Aberle; Kinga Rigler-Hohenwarter; Lukas Endler; Manfred Nairz; Mark Smyth; Martin Senekowitsch; Michael Schuster; Peter Hufnagl; Peter Obrist; Rainer Gattringer; Sabine Sussitz-Rack; Stephan Aberle; Thomas Penz; Wegene Borena |
| EPI_ISL_447038 | B.J. Medical College and Civil hospital | Gujarat Biotechnology Research Centre | Akanksha Verma; Amit Kanani; Ankit Hinsu; Apurvasinh Puvar; Armi Chaudhari; Bhavesh Modi; Binita Aring; Chaitanya Joshi; Dinesh Kumar; Dipa Kinariwala; Disha Patel; Gaurishankar Shrimali; Geeta Vaghela; Janvi Raval; Kairavi Joshi; Kamlesh J Upadhyay; Madhvi Joshi; Maharshi Pandya; Monika Gandhi; Neeta Khandelwal; Nidhi Sood; Nitin Savaliya; Pinal Trivedi; Pranay Shah; Pritesh Sabara; R D Dixit; Raghawendra Kumar; Ramesh Pandit; Snehal Bagatharia; Sonia Barve; Tejas Shah; Zuber Saiyed |
| EPI_ISL_803904 | BBMP Urban PHC | Department of Neurovirology, National Institute of Mental Health and Neurosciences (NIMHANS) | Anita S Desai; Chitra Pattabiraman; Pramada Prasad; V Ravi |
| EPI_ISL_412965 | BCCDC Public Health Laboratory | BCCDC Public Health Laboratory | Choi; Gilmour; Harrigan; Hoang; Kamelian; Krajden; Lapointe; Lee; Levett; Li; Loman; Prystajecky; Quick; Sekirov; Tyson |
| EPI_ISL_574619 | BTKLPP Kelas I Manado | Eijkman Institute for Molecular Biology, Ministry of Research and Technology/National Agency for Research and Innovation | Amin Soebandrio; David H Muljono; Edison Johar; Frilasita A Yudhaputri; Herawati Sudoyo; Hidayat Trimarsanto; Iskandar A Adnan; Khin Saw Myint; Safarina G Malik; Willy Agustine |
| EPI_ISL_791978 | Balai Labkes Lampung | National Institute of Health Research and Development | AA; HA; HD; Ikawati; KD; KNA; L; N; Nugraha; Pangesti; Pawestri; Puspa; Puspandari; Setiawaty; Soekarso; Subangkit; T; V; Yurina |
| EPI_ISL_775594 | Balitvet Lampung | National Institute of Health Research and Development | AA; E; EA; HA; HD; Ikawati; KD; KNA; N; Nugraha; Pangesti; Pawestri; Puspa; Puspandari; Saswiyanti; Setiawaty; Soekarso; Srihanto; Subangkit; T; V |
| EPI_ISL_403963 | Bamrasnaradura Hospital | 1. Department of Medical Sciences, Ministry of Public Health, Thailand 2. Thai Red Cross Emerging Infectious Diseases - Health Science Centre 3. Department of Disease Control, Ministry of Public Health, Thailand | Buathong; Chittaganpitch; Malinee; Mekha; Nanthawan; Okada; Parnmen; Phuygun; Pilailuk; Rome; Siripaporn; Sittiporn; Sunthareeya; Supaporn; Thanadachakul; Thanutsapa; Wacharapluesadee; Waicharoen; Warawan; Wongboot |
| EPI_ISL_434693 | Bamrasnaradura hospital | National Institute of Health. Department of medical Sciences, Ministry of Public Health, Thailand | Chittaganpitch; Malinee; Okada; Parnmen; Phuygun; Pilailuk; Siripaporn; Sittiporn; Sunthareeya; Thanadachakul; Thanutsapa; Waicharoen; Warawan; Wongboot |
| EPI_ISL_529216 | Beijing Institute of Microbiology and Epidemiology | Beijing Institute of Microbiology and Epidemiology | Cui, Y.; Fan; Guo, Y.; Hang; Hou, J.; Li, B.; Mi, Z.; Mu, J.; Qin, E.; Song; Teng; Wu, Y.; Xu, Z.; Yajun.; Yang, R.; Yong, Y.; Yue; Zhang, X. |
| EPI_ISL_509711, EPI_ISL_509712, EPI_ISL_509713, EPI_ISL_509714 | Belize Ministry of Health | Pathogen Discovery, Respiratory Viruses Branch, Division of Viral Diseases, Centers for Disease Control and Prevention | Anna Uehara; Clinton Paden; Haibin Wang; Jing Zhang; Krista Queen; Suxiang Tong; Yan Li; Ying Tao |
| EPI_ISL_780385, EPI_ISL_780391, EPI_ISL_780398, EPI_ISL_780399 | Bermuda Government Molecular Diagnostics Laboratory (MDL) | Respiratory Virus Unit, National Infection Service, Public Health England | Dr Ayoola Oyinloye (Bermuda); Dr Carika Weldon (Bermuda); PHE Covid Sequencing Team |
| EPI_ISL_735495 | Bhashabir M A Wadud RT-PCR Lab, Chandpur | Central Biological Research Laboratory and Department of Biochemistry and Molecular Biology | H. M. Abdullah Al Masud; Imam Hossen; Md. Arif Hossain; Md. Imranul Hoq; Md. Khondakar Raziur Rahman; Md. Omer Faruq; Mohammad Omar Faruque; Robiul Hasan Bhuiyan; Sajib Rudra; Shanta Paul |
| EPI_ISL_430000, EPI_ISL_430006, EPI_ISL_434516, EPI_ISL_450187, EPI_ISL_450188, EPI_ISL_635777, EPI_ISL_635778, EPI_ISL_635779, EPI_ISL_635780, EPI_ISL_635781, EPI_ISL_636315, EPI_ISL_730377, EPI_ISL_730399, EPI_ISL_730440, EPI_ISL_730544, EPI_ISL_730557, EPI_ISL_755233, EPI_ISL_755234, EPI_ISL_755236, EPI_ISL_755241, EPI_ISL_755256 | | | |
| see above | Biolab Diagnostic Laboratories | Andersen lab at Scripps Research | Ahmad Tibi; Amid Abdelnour with SEARCH Alliance San Diego; Issa Abu-Dayyeh; Lama Hussein; Lina Mohammad; Zein Naber |
| EPI_ISL_527007 | Biological Prevention, Army | Biological Prevention, Army | A.E.; A.F.; A.M.; Ageez; B.E.; Elhoseiny; Gad; Harty; M.D.; M.F.; M.G.; Seadawy; Shabaan; Shamel |
| EPI_ISL_526992 | Biological prevention, army | Biological prevention, army | A.F.; B.E.; Elhosieny; Gad; Harty; M.D.; M.F.; M.G.; Seadawy; Shamel |
| EPI_ISL_582030 | Biology Department, College of Science, Al-Muthanna University | International Centre for Genetic Engineering and Biotechnology (ICGEB) and ARGO Open Lab Platform | Alessandro Marcello; Danilo Licastro; Nihad Al-Rashedi; Simeone Dal Monego; Sreejith Rajasekharan |
| EPI_ISL_794634 | Biology, MCL | Biology, MCL | A.F.; A.S.; B.S. and EL-Safty; EL-harty; EL-hosieny; Gad; M.D.; M.F.; M.G.; Seadawy; Shamel |
| EPI_ISL_458287 | Biosafety Department PCL3 | Biosafety Department PCL3 | A. and El Kabbaj, S.; Lemriss, S.; Souiri |
| EPI_ISL_747238 | Bogor Public Health | West Java Health Laboratory; School of Life Sciences and Technology, Institut Teknologi Bandung | Azzania Fibriani; Cut Nur Cinthia Alamanda; Ema Rahmawati; Isak Solihin; Karimatu Khoirunnisa; Miftahul Faridl; Rifky Waluyajati Rachman; Rini Robiani; Ryan Bayusantika Ristandi |
| EPI_ISL_596453 | Booali laboratory, Qom, Iran | Genetics Research Center, University of Social Welfare and Rehabilitation Sciences | Ali Jafarpour; Azam Ghaziasadi; Hossein Najmabadi; Khadijeh Jalalvand; Kimia Kahrizi; Marzieh Mohseni; Mohammad Khazeni; Seyed Amir Momeni; Seyed Mohammad Jazayeri; Seyedeh elham Mortazavi; Zohreh Fattahi |
| EPI_ISL_718167, EPI_ISL_718197 | Borneo Medical Centre | Institute of Health and Community Medicine | Chan Chia Jui; Chua Hock Hin; David Perera; Ooi Mong How; Tonnii Sia Loong Loong; Wong Jyn Shan; Wong Kiing Aik |
| EPI_ISL_414520, EPI_ISL_732559 | Bundeswehr Institute of Microbiology | Bundeswehr Institute of Microbiology | Alexandra Rehn; Elham Khatamzas; Enrico Georgi; Markus Antwerpen; Markus H Antwerpen and Roman Wölfel; Mathias C Walter; Mathias Walter; Michael von Bergwelt-Baildon; Roman Wölfel; Sabine Zange |
| EPI_ISL_539531 | C.H.U Nuestra Señora de Candelaria | Instituto de Salud Carlos III | A. Monzón; F. Casas; I; I. Jiménez; Iglesias-Caballero; M. Camarero; M. Cuesta; M. González-Esguevillas; M. Molinero Calamita; M. Zaballos; O. Díez; P. Jiménez; S. Juliá; S. Pozo; S. Varona |
| EPI_ISL_693545, EPI_ISL_731946 | CEDOC_B | Instituto Nacional de Saude (INSA) | Borges et al |
| EPI_ISL_693550 | CEDOC_F | Instituto Nacional de Saude (INSA) | Borges et al |
| EPI_ISL_693552 | CEDOC_H | Instituto Nacional de Saude (INSA) | Borges et al |
| EPI_ISL_605780 | CEIRS Data Processing and Coordinating Center, St. Jude Center of Excellence for Influenza Research and Surveillance (CEIRS) | CEIRS Data Processing and Coordinating Center, St. Jude Center of Excellence for Influenza Research and Surveillance (CEIRS) | A.E.; Ali; El-Guindy; El-Sayes, M.; El-Shesheny, R.; El-Taweel, A.; Gomaa, M.; Kamel; Kandeil, A.; Kayali, G.; Kayed; Kutkat, O.; M.A.; M.N.; Mahmoud; Mahrous, N.; Moatasim, Y.; Mostafa, A.; N.M.; Naguib, A.; Roshdy; S.H.; Shehata, M.; Showky, S.; W.H.; Webby, R. |
| EPI_ISL_644252 | CEPHR / Mater Hospital | Irish Coronavirus Sequencing Consortium - National Virus Reference Laboratory | Alejandro Abner Garcia Leon; Gabriel Gonzalez; Michael Carr; Patrick Mallon |
| EPI_ISL_644280 | CEPHR / Vincent's Hospital | Irish Coronavirus Sequencing Consortium - National Virus Reference Laboratory | Alejandro Abner Garcia Leon; Gabriel Gonzalez; Michael Carr; Patrick Mallon |
| EPI_ISL_445316 | CESFAM BALMACEDA DE RENCA | Instituto de Salud Publica de Chile | Alejandra Acevedo; Andrés E Castillo; Bárbara Parra; Carolina Tambley; Gabriel Leal; Jaime Lagos; Jorge Fernandez; Loredana Arata; Patricia Bustos; Paz Tapia; Rodrigo Fasce; Winston Andrade |
| EPI_ISL_733569 | CHAI WAN FAMILIES CLINIC | Hong Kong Department of Health | Alan K.L. Tsang; Dominic N.C. Tsang; Edman T.K. Lam; Peter C.W. Yip; Rickjason C.W. Chan |
| EPI_ISL_791998 | CHU - Hôpital Cavale Blanche | National Reference Center for Viruses of Respiratory Infections, Institut Pasteur, Paris | Angela Brisebarre; Camille Capel; Etienne Simon-Lorière; Léa Pilorgé; Marion Barbet; Maud Vanpeene; Méline Bizard; Sylvie Behillil; Sylvie van der Werf; Vincent Enouf |
| EPI_ISL_641548 | CHU Clermont-Ferrand | CNR Virus des Infections Respiratoires - France SUD | Amélie Brebion; Antonin Bal; Audrey Mirand; Bruno Lina; Christel Regagnon; Christine Archimbaud; Cécile Henquell; Gregory Destras; Gwendolyne Burfin; Hadrien Règue; Hélène Chabrolles; Laurence Josset; Martine Chambon; Martine Valette; Maxime Bisseux; Patricia Combes; Quentin Semanas |
| EPI_ISL_591542, EPI_ISL_593869, EPI_ISL_593891, EPI_ISL_593899, EPI_ISL_751484 | CHU Purpan - Laboratoire de Virologie - Institut Fédératif de Biologie | CHU Purpan - Laboratoire de Virologie - Institut Fédératif de Biologie | Boyer P.; Carcenac R.; Dubois M.; Harter A.; Izopet J.; Latour J.; Ranger N.; Tremeaux P. |
| EPI_ISL_482883 | CHU Purpan - Laboratoire de Virologie - Institut Fédératif de Biologie | Laboratoire de virologie - École Nationale Vétérinaire de Toulouse | Guillaume Croville; Jacques Izopet; Jean-Luc Guérin |
| EPI_ISL_417987 | CHU Sao Joao, Porto | Instituto Nacional de Saude (INSA) | Guiomar et al |
| EPI_ISL_645185, EPI_ISL_645186, EPI_ISL_660335 | CHU de Saint-Étienne Hôpital Nord | CNR Virus des Infections Respiratoires - France SUD | Antonin Bal; Bruno Lina; Bruno Pozzetto; Gregory Destras; Gwendolyne Burfin; Hadrien Règue; Issam Bechri; Laurence Josset; Manon Vogrig; Marine Delorme; Martine Valette; Quentin Semanas; Sylvie Gonzalo; Sylvie Pillet; Thomas Bourlet |
| EPI_ISL_417993 | CHULC - H D Estefania | Instituto Nacional de Saude (INSA) | Guiomar et al |
| EPI_ISL_731978 | CHULN - H Santa Maria | Instituto Nacional de Saude (INSA) | Borges et al |
| EPI_ISL_683835 | CICM | Malaria Research and Training Center (MRTC-Parasito) | Abdoulaye Djimde; Antoine Dara |
| EPI_ISL_487448, EPI_ISL_487465 | CICM-Mali | Bundeswehr Institut of Microbiology | Antwerpen; Bestehorn-Willmann; Dürr; Heitzer; Kouriba; Maiga; Quedraogo; Rehn; Sangaré; Sogodogo; Traoré; Walter; Wölfel; Zimmermann |
| EPI_ISL_410486, EPI_ISL_508928, EPI_ISL_508964, EPI_ISL_640003, EPI_ISL_683364, EPI_ISL_732682, EPI_ISL_732689, EPI_ISL_779811 | | | |
| see above | CNR Virus des Infections Respiratoires - France SUD | CNR Virus des Infections Respiratoires - France SUD | Alexandre; Alexandre Gaymard; Antonin; Antonin Bal; Bal; Bouscambert-Duchamp; Brengel-Pesce; Bruno Lina; Bruno.; Carine Moustaud; Cheynet; Claudia Gonzalez; Destras; Florence; Florence Morfin-Sherpa; Gaymard; Gregory; Gregory Destras; Gwendolyne Burfin; Hadrien Règue; Josset; Karen; Laurence; Laurence Josset; Lina; Martine; Martine Valette; Maude; Maude Bouscambert-Duchamp; Morfin-Sherpa; Quentin Semanas; Raphaelle Lamy; Solenne Brun; Valette; Valérie |
| EPI_ISL_735408 | COVID 19 Centro de Combate ao Coronavirus CCC Jandira | Instituto Adolfo Lutz, Interdiciplinary Procedures Center, Strategic Laboratory | Claudia Regina Gonçalves; Claudio Tavares Sacchi; Erica Valessa Ramos Gomes; Karoline Rodrigues Campos |
| EPI_ISL_450843 | COVID-19 Laboratory | DNA Solution Ltd. | ABM Khademul Islam; AHM Nurun Nabi; Abu Sufian; Gazi Nurun Nahar; Habibul Bari Shozib; Haseena Khan; Imran Khan; Latiful Bari; M Anwar Hossain.; MA Malek; Mamun Ahmed; Md Imdadul Hoque; Md Ismail Hosen; Md Mizanur Rahman; Mohammad Riazul Islam; Nazmul Ahsan; Richard Malo; Sabita Rezwana Rahman; Sabrina Moriom Elius; Shahryar Nabi; Sharif Akhteruzzaman; Zeba Islam Seraj |
| EPI_ISL_574593, EPI_ISL_574596, EPI_ISL_583494 | CS II Dr. Antonio Vicoso Moreira de Rezende Sumare | Instituto Adolfo Lutz, Interdiciplinary Procedures Center, Strategic Laboratory | Claudia Regina Gonçalves; Claudio Tavares Sacchi; Erica Valessa Ramos Gomes; Karoline Rodrigues Campos |
| EPI_ISL_471585, EPI_ISL_528834, EPI_ISL_539758 | CSIR-Centre for Cellular and Molecular Biology | CSIR-Centre for Cellular and Molecular Biology | Archana Bharadwaj Siva; Dhiviya Vedagiri; Disha Nanda; Divya Das; Divya Gupta; Divya Tej Sowpati; G. Aditya Kumar; Jotin Gogoi; Karthik Bharadwaj Tallapaka; Koushick Sivakumar; Krishnan Harinivas Harshan; Lamuk Zaveri; M Soujanya Reddy; Manish Bhattacharjee; Namami Gaur; Nikhil Hajirnis; Onkar Kulkarni; Payel Mukherjee; Peddapuvala Sai Uday Kiran Rakesh K Mishra; Pratheusa Maccha; Preethi Jampala; Priya Singh; Purushotham Vodnala; Rakesh K Mishra; Ravi Prasad Mukku; Sakshi Shambhavi; Santosh Kumar Kuncha; Shagufta Khan; Sharada Ravi Iyer; Sofia |

| | | | |
|---|---|---|---|
| | | | Banu; Sulagana Mukherjee; Swetha Sundar; Tulasi Nagabandi; Vishal Sah |
| EPI_ISL_661305 | CSIR-Indian Institute of Chemical Biology, MEDICA Supercpecialty Hospital Kolkata | CSIR-Indian Institute of Chemical Biology, MEDICA Supercpecialty Hospital Kolkata | Abhishake Lahiri; Debaleena Bhowmik; Dr. Arpita Ghosh Mitra; Dr. Aviral Roy; Dr. Partha Chakrabarti; Dr. Rajesh Pandey; Dr. Saikat Chakrabarti; Dr. Sandip Paul; Dr. Soumen Saha; Priyanka Mallick; Sujay Krishna Maity |
| EPI_ISL_468314, EPI_ISL_583503 | CTA Centro de Testagem e Aconselhamento | Instituto Adolfo Lutz, Interdiciplinary Procedures Center, Strategic Laboratory | Claudia Regina Gonçalves; Claudio Tavares Sacchi; Erica Valessa Ramos Gomes; Karoline Rodrigues Campos |
| EPI_ISL_535749 | CUSM-Site Glen-LAB Microbiologie | Laboratoire de santé publique du Québec | Guillaume Bourque; Ioannis Ragoussis; Jesse Shapiro; Mark Lathrop and Michel Roger; Sandrine Moreira |
| EPI_ISL_582267, EPI_ISL_582318, EPI_ISL_582322, EPI_ISL_582372, EPI_ISL_582373, EPI_ISL_582427, EPI_ISL_582442, EPI_ISL_582494 | | | |
| see above | Cadham Provincial Laboratory | National Microbiology Laboratory (NML) | Anna Majer; Anneliese Landgraff; CanCOGeN's metadata curation team; Darian Hole; David Alexander; Elsie Grudeski; Gary Van Domselaar; Grace Seo; Jared Bullard; Jennifer Tanner; Kerry Dust; Madison Chapel; Morag Graham; Natalie Knox; Nathalie Bastien; Paul Van Caeseele; Philip Mabon; Public Health Agency of Canada CanCOGeN team; Rhiannon Huzarewich; Russell Mandes; Shari Tyson; Timothy Booth; Yan Li |
| EPI_ISL_411955 | California Department of Public Health | Pathogen Discovery, Respiratory Viruses Branch, Division of Viral Diseases, Centers for Dieases Control and Prevention | Anna Uehara; Brett L. Whitaker; Brian Lynch; Clinton R. Paden; Haibin Wang; Janna' R. Murray; Jing Zhang; Krista Queen; Lijuan Wang; Senthil Kumar K. Sakthivel; Shifaq Kamili; Stephen Lindstrom; Susan I. Gerber; Suxiang Tong; Xiaoyan Lu; Yan Li; Ying Tao |
| EPI_ISL_548140, EPI_ISL_579481, EPI_ISL_794627 | Canterbury Health Laboratories | Institute of Environmental Science and Research (ESR) | Anja Werno; Antje van der Linden; Arlo Upton; Chris Mansell; David Hammer; Dragana Drinkovic; Erasmus Smit; Gary McAuliffe; Hana Sofia Andersson; Hermes Perez; James Ussher; Jill Sherwood; Jing Wang; Joep de Ligt; Josh Freeman; Julia Howard; Juliet Elvy; Lauren Jelly; Mary DeAlmeida; Matt Blakiston; Matt Storey; Matthew Rogers; Max Bloomfield; Michael Addidle; Michelle Balm; Muhammad Faisal; Nikki Freed; Olin Silander; Sally Roberts; Sarah Jefferies; Sharmini Muttaiyah; Susan Morpeth; Susan Taylor; Timothy Blackmore; Vani Sathyendran; Veronica Playle; Virginia Hope; Xiaoyun Ren |
| EPI_ISL_583504, EPI_ISL_583505 | Casa de Saude Stella Maris | Instituto Adolfo Lutz, Interdiciplinary Procedures Center, Strategic Laboratory | Claudia Regina Gonçalves; Claudio Tavares Sacchi; Erica Valessa Ramos Gomes; Karoline Rodrigues Campos |
| EPI_ISL_693235 | Casmi Centro Atendimento Saude da Mulher e Infancia | Instituto Adolfo Lutz, Interdiciplinary Procedures Center, Strategic Laboratory | Claudia Regina Gonçalves; Claudio Tavares Sacchi; Erica Valessa Ramos Gomes; Karoline Rodrigues Campos |
| EPI_ISL_756293, EPI_ISL_756294 | Center for Biotechnology and Cell Therapy, São Rafael Hospital, Salvador, Brazil | Center for Biotechnology and Cell Therapy, São Rafael Hospital, Salvador, Brazil | Ana Verena Almeida Mendes; Bruno Solano de Freitas Souza; Carolina Kymie Vasques Nonaka; Marta Giovanetti; Marília Miranda Franco; Renato Santana de Aguiar; Tiago Gräf |
| EPI_ISL_522515, EPI_ISL_526738, EPI_ISL_526746 | Center for Laboratory Control of Infectious Diseases, Korea Centers for Diseases Control and Prevention | Center for Laboratory Control of Infectious Diseases, Korea Centers for Diseases Control and Prevention | Ae Kyung Park; Eunkyung Shin; Heui Man Kim; Jeong-Min Kim; Jin Sun No; Junyoung Kim; Myung Guk Han; Yoon-Seok Chung |
| EPI_ISL_419669, EPI_ISL_437995, EPI_ISL_438019, EPI_ISL_438093, EPI_ISL_583704, EPI_ISL_583706 | Center for Virology, Medical University of Vienna | Bergthaler laboratory, CeMM Research Center for Molecular Medicine of the Austrian Academy of Sciences | Adi Steinrigl; Alexander Lercher; Alexandra Popa; Andreas Bergthaler; Benedikt Agerer; Christian Paar; Christoph Bock; Daniela Schmid; Dorothee von Laer; Elisabeth Puchhammer-Stoeckl; Elisabeth Puchhammer-Stöckl; Franz Allerberger; Gernot Walder; Gregor Hörmann; Guenter Weiss; Gunther Vogl; Henrique Colaco; Jakob-Wendelin Genger; Jan Laine; Judith Aberle; Kinga Rigler-Hohenwarter; Lukas Endler; Manfred Nairz; Mark Smyth; Martin Senekowitsch; Michael Schuster; Peter Hufnagl; Peter Obrist; Rainer Gattringer; Sabine Sussitz-Rack; Stephan Aberle; Thomas Penz; Wegene Borena |
| EPI_ISL_755574 | Center of Advanced Studies and Technology, CAST | Center of Advanced Studies and Technology, CAST | De Fabritiis, S.; Ferrante, R.; Mandatori, D. |
| EPI_ISL_414497, EPI_ISL_779953 | Center of Medical Microbiology, Virology, and Hospital Hygiene, University of Duesseldorf | Center of Medical Microbiology, Virology, and Hospital Hygiene, University of Duesseldorf | Alexander Dilthey; Andreas Walker; Ashley-Jane Duplessis; Björn-Erik Jensen; Daniel Strelow; Detlef Kindgen-Milles; Jessica Nicolai; Jörg Timm; Klaus Pfeffer; Lisanna Hülse; Malte Kohns Vasconcelos; Marcel Andree; Maximilian Damagnez; Nadine Lübke; Ortwin Adams; Sandra Hauka; Teresa Tamayo; Tina Senff; Tobias Wienemann; Torsten Feldt; Torsten Houwaart |
| EPI_ISL_430820 | Center of Scientific Excellence for Influenza Viruses, National Research Centre (NRC), Egypt. | Center of Scientific Excellence for Influenza Viruses, National Research Centre (NRC), Egypt. | Abo Shama; Ahmed E Kayed; Ahmed El-Taweel; Ahmed Kandeil; Ahmed Mostafa; Amal Naguib; M Noura; Mahmoud Shehata; Mina Kamel; Mohamed Ahmed Ali; Mohamed El Sayes; Mokhtar Gomaa; Nancy M. El Guindy; Omnia Kutkat; Rabeh El-Shesheny; Sara Mahmoud; Shymaa Showky Ahmed; Wael Roshdy; Yassmin Moatasim |
| EPI_ISL_429854, EPI_ISL_495459 | Centers for Disease Control and Prevention of Lishui | Department of Inspection , Centers for Disease Control and Prevention of Lishui | Ji Jiansong; Ji Qiaoying; Wang Xiaoguang; Ye Bifeng; Ye Ling |
| EPI_ISL_428489 | Centers for Disease Control, R.O.C. (Taiwan) | Centers for Disease Control, R.O.C. (Taiwan) | Ji-Rong Yang; Jung-Jung Mu; Ming-Tsan Liu; Yu-Chi Lin |
| EPI_ISL_529032 | Central Molecular Microbiology Laboratory and Next Generation Sequencing Reference Laboratory, Clinical and Chemical Pathology Department, Faculty of Medicine, CAIRO UNIVERSITY | Next Generation Sequencing Reference Laboratory, Faculty of Medicine, CAIRO UNIVERSITY and The Center for Genome and Microbiome Research, Faculty of Pharmacy, CAIRO UNIVERSITY | May Abdelfattah; May Sherif Soliman; Ramy Karam Aziz |
| EPI_ISL_693473, EPI_ISL_693476, EPI_ISL_693477, EPI_ISL_693480, EPI_ISL_693481, EPI_ISL_693483 | Central Public Health Laboratory | National Public Health Laboratory, National Centre for Infectious Diseases | Esorom Daoni; Lin Cui; Raymond Tzer Pin Lin; Sophie Octavia; Theresa Palou; Tze Minn Mak; Zhenyang Zhou |
| EPI_ISL_429667, EPI_ISL_429669, EPI_ISL_429671, EPI_ISL_429674, EPI_ISL_429676, EPI_ISL_429681, EPI_ISL_429687, EPI_ISL_429688, EPI_ISL_429689, EPI_ISL_429695, EPI_ISL_429702 | | | |
| see above | Central Public Health Laboratory/Octávio Magalhães Institute (IOM) from the Ezequiel Dias Foundation (FUNED) | Instituto Octávio Magalhães / Fundação Ezequiel Dias (IOM/Funed) | Joilson Xavier; Luiz Carlos Junior Alcantara; Marcos Vinícius Silva; Marluce Aparecida Assunção Oliveira; Marta Giovanetti; Talita Adelino; Vagner Fonseca |
| EPI_ISL_419211 | Central Virology Laboratory | Israel Institute for Biological Research | Adi Beth-Din; Anat Zvi; Boaz Politi; Dana Stein; Einat Vitner; Gadi Segal; Gili Regev-Yochay; Hadas Tamir; Hagit Achdout; Inbar Cohen-Gihon; Lilach Cherry; Michal Mandelboim; Nir Paran; Ofir Israeli; Ohad Shifman; Oran Erster; Orly Laskar; Sharon Melamed; Shay Weiss; Shmuel C. Shapira; Shmuel Yitzhaki; Tomer Israely; Yfat Yahalom Ronen |
| EPI_ISL_486410 | Centrala laboratorija | Latvian Biomedical Research and Study Centre | Ivars Silamiķelis; Jana Ošīte; Jānis Kloviņš; Kaspars Megnis; Marta Priedīte; Monta Ustinova; Stella Lapiņa; Uga Dumpis; Vita Rovīte; Ņikita Zrelovs |
| EPI_ISL_458000 | Centre For Biotechnology Research and Development | Centre For Biotechnology Research and Development | C.N. and Michuki; D.K.; G.N.; J.O.; Kimotho, J.; Matoke-Muhia; Muuo; Ochwoto, M.; S.L.; S.N.; Symeker; Waruhiu; Zablon |
| EPI_ISL_416495 | Centre Hospitalier Compiègne Laboratoire de Biologie | National Reference Center for Viruses of Respiratory Infections, Institut Pasteur, Paris | Angela Brisebarre; Etienne Simon-Lorière; Flora Donati; Marion Barbet; Maud Vanpeene; Méline Bizard; Mélnie Albert; Raulin Olivia; Sylvie Behillil; Sylvie van der Werf; Vincent Enouf |
| EPI_ISL_582121 | Centre Hospitalier de Bourg en Bresse | CNR Virus des Infections Respiratoires - France SUD | Alexandre Gaymard; Antonin Bal; Bruno Lina; Florence Morfin-Sherpa; Gregory Destras; Gwendolyne Burfin; Hadrien Règue; Laurence Josset; Martine Valette; Maude Bouscambert-Duchamp |
| EPI_ISL_788936, EPI_ISL_788938 | Centre de Recherche et de Formation en Infectiologie Guinée | TransVIHMI, IRD/INSERM/Monpellier University | Abdoul Karim SOUMAH; Abdoulaye TOURE; Ahidjo AYOUBA; Alimou CAMARA; Alpha Kabinet KEITA; Bouna Yatassaye; Christelle BUTEL; Eric DELAPORTE; Jean-louis MONEMOU; Joel KOIVOGUI; Kaba KOUROUMA; Laetitia SERRANO; Mamadou Bhoye KEITA; Mamadou Saliou BAH; Mamadou Saliou SOW; Mandiou DIAKITE; Martine PEETERS; Moriba POVOGUI; Penda Malhado DIALLO; Sakoba KEITA |
| EPI_ISL_539573, EPI_ISL_539576 | Centre de Recherches Medicales de Lambarene (CERMEL) | Department of Emerging Infectious Diseases, Institute of Tropical Medicine, Nagasaki University | Akim A. Adegnika; Bertrand Lell; Haruka Abe; Jiro Yasuda; Rodrigue Bikangui; Yuri Ushijima |
| EPI_ISL_535724 | Centre de SSS La Pommeraie | Laboratoire de santé publique du Québec | Guillaume Bourque; Ioannis Ragoussis; Jesse Shapiro; Mark Lathrop and Michel Roger on behalf of the CoVSeQ research group; Sandrine Moreira |
| EPI_ISL_471457, EPI_ISL_471458 | Centre de Virologie des Maladies Tropicales | Functional Genomic Platform/Service Analyses Biologique/UATRS/ Centre National Pour la Recherche Scientifique Et Technique (CNRST) | Abdelillah LARAQUI; Abdelkader LAATIRIS; Ahmed REGGAD; Elmostafa EL FAHIME; Farida HILALI; Hicham EL ANNAZ; Idriss-Amine LAHLOU; Khalid ENNIBI; Marouane MELLOUL; Mly Abdelaziz ELALAOUI; Mostafa ELOUENNASS; Nadia TOUIL; Rachid ABI; Rida TAGAJDID; Safae ELKOCHRI; Sanaa ALAOUI-Amine; Tahar BAJJOU; Yassine SEKHSOKH; Youssef AKHOUAD; Zohour KASMY |
| EPI_ISL_428670, EPI_ISL_428671, EPI_ISL_525476, EPI_ISL_525478, EPI_ISL_525479, EPI_ISL_525481, EPI_ISL_525489 | | | |
| see above | Centre for Dengue Research | Centre for Dengue Research | Ananda Wijewickrama; Chandima Jeewandara; Damayanthi Idampitiya; Damayanthi Tdampitiya; Deshni Jayathilaka; Dinuka Ariyaratne; Dinuka Ariyatane; Dinuka Guruge; Diyanath Ranasinghe; Eranga Narangoda; Gathsaurie Neelika Malavige; Laksiri Gomes; Malika Karunaratne; Neelika Malaige; Neelika Malavige; Ruwan Wijayamuni |
| EPI_ISL_525474 | Centre for Dengue Research | Centre for Dengue Research, USJ, SL | Chandima Jeewandara; Deshni Jayathilaka; Dinuka Ariyaratne; Dinuka Guruge; Diyanath Ranasinghe; Gathsaurie Neelika Malavige; Laksiri Gomes; Ruwan Wijayamuni |
| EPI_ISL_602564 | Centre for Dengue Research, Department of Immunology and Molecular Medicine | Centre for Dengue Research | Ananda Wijewickrama; Chandima Jeewandara; Deshni Jayathilaka; Dinuka Ariyaratne; Diyanath Ranasinghe; Gathsaurie Neelika Malavige; Laksiri Gomes; Malika Karunaratne |
| EPI_ISL_668454 | Centre for Dengue Research, Department of Immunology and Molecular Medicine | Centre for Dengue Research, Department of Immunology and Molecular Medicine | Chandima Jeewandara; Deshni Jayathilaka; Dinuka Ariyaratne; Diyanath Ranasinghe; Gathsaurie Neelika Malavige; Laksiri Gomes |
| EPI_ISL_733568 | Centre for Health Protection | Hong Kong Department of Health | Alan K.L. Tsang; Dominic N.C. Tsang; Edman T.K. Lam; Peter C.W. Yip; Rickjason C.W. Chan |
| EPI_ISL_417383 | Centre for Infectious Diseases and Microbiology Public Health | NSW Health Pathology - Institute of Clinical Pathology and Medical Research; Westmead Hospital; University of Sydney | Arnott A; Carter I; Chen SC; Eden J-S; Gall M; Gray K; Holmes EC; Kok J and Dwyer DE for the 2019-nCoV Study Group; Lam C; Maddocks S; O'Sullivan MV; Rahman H; Rockett R; Sadsad R; Sintchenko V; Timms V |
| EPI_ISL_639739 | Centre of Nanotechnologies, INCD IMT-Bucuresti (National Institute for Research and Development in Microtechnologies - Bucharest) | Centre of Nanotechnologies, INCD IMT-Bucuresti (National Institute for Research and Development in Microtechnologies - Bucharest) | Gogianu; L. and Baisan, M.; Salceanu, A. |
| EPI_ISL_515547 | Centro Medico da Policia Militar do Estado de Sao Paulo | Instituto Adolfo Lutz, Interdiciplinary Procedures Center, Strategic Laboratory | Claudia Regina Gonçalves; Claudio Tavares Sacchi; Erica Valessa Ramos Gomes |
| EPI_ISL_693248 | Centro Municipal de Epidemiologia e Imunizações | Instituto Adolfo Lutz, Interdiciplinary Procedures Center, Strategic Laboratory | Claudia Regina Gonçalves; Claudio Tavares Sacchi; Erica Valessa Ramos Gomes; Karoline Rodrigues Campos |
| EPI_ISL_693244 | Centro Médico da Polícia Militar do Estado de Sao Paulo | Instituto Adolfo Lutz, Interdiciplinary Procedures Center, Strategic Laboratory | Claudia Regina Gonçalves; Claudio Tavares Sacchi; Erica Valessa Ramos Gomes; Karoline Rodrigues Campos |
| EPI_ISL_629021 | Centro de Biotecnología Vegetal, Universidad Andrés Bello, Center for Genome Regulation | Center for Mathematical Modeling and Center for Genome Regulation. Santiago, Chile | Allende ML; Arriagada G; Bastias M; Bustos F; Castro E; González M; M; Maass A; Meneses C.; Montecino; Orellana A; Sanhueza D; Travisany D |
| EPI_ISL_483065, EPI_ISL_509430, EPI_ISL_509431, EPI_ISL_509432, EPI_ISL_509433, EPI_ISL_509434, EPI_ISL_509435, EPI_ISL_510536, EPI_ISL_529139 | | | |
| see above | Centro de Desenvolvimento Tecnologico em Saude, Fundacao Oswaldo Cruz | Centro de Desenvolvimento Tecnologico em Saude, Fundacao Oswaldo Cruz | A.D.; Barroso; C.Q.; C.Q. and Medeiros; De Paula; F.B.; Ferreira; Fintelman-Rodrigues, N.; Gregorio; J.S.; M.A.; M.L.; Medeiros; Oliveira; S.P.; Sacramento; Saraiva; Souza; T.M.; Tschoeke, D. |
| EPI_ISL_510541, EPI_ISL_529140 | Centro de Desenvolvimento Tecnologico em Saude, Fundacao Oswaldo Cruz | Centro de Desenvolvimento Tecnologico em Saude, Fundacao Oswaldo Cruz | A.D.; C.Q.; C.Q. and Medeiros; De Paula; F.B.; Ferreira; Fintelman-Rodrigues, N.; M.A.; Medeiros; Sacramento; Saraiva; Souza; T.M. |
| EPI_ISL_635486, EPI_ISL_635496, EPI_ISL_635510, EPI_ISL_635535, EPI_ISL_635540, EPI_ISL_635563, EPI_ISL_635574, EPI_ISL_635576 | | | |

| | | | |
|---|---|---|---|
| see above | Centro de Diagn√≥stico COVID-19 UABC Tijuana | Andersen lab at Scripps Research | Germán Ibarra; Jonathan Vincent Baena; Jorge Luis Jiménez Niebla; Manuel Sánchez Alavez; Oscar Efrén Zazueta Fierro; SEARCH Alliance San Diego with Idanya Rubí Serafín Higuera |
| EPI_ISL_491934, EPI_ISL_491935 | Centro de Investigaciones, Universidad de Especialidades Espíritu Santo | Institute of Microbiology, Universidad San Francisco de Quito | Belén Prado-Vivar; Bernardo Gutiérrez; Derly Andrade; Edith Lopez; Fernando Espinoza; Gabriel Morey; Gabriel Trueba; Jose Pedro Barberan; Juan Carlos Fernandez; Juan José Guadalupe; Michelle Grunauer; Monica Becerra-Wong; Patricio Rojas-Silva; Paúl Cárdenas; Ruben Armas; Sully Márquez; Verónica Barragán |
| EPI_ISL_732779 | Centro de Investigación Biomédica de La Rioja - Hospital San Pedro Logroño | SeqCOVID-SPAIN consortium/IBV(CSIC) | José Manuel Azcona Gutiérrez; María Pilar Bea Escudero; María de Toro; Miriam Blasco Alberdi and SeqCOVID-SPAIN consortium |
| EPI_ISL_583491 | Centro de Saude Esf IV Zona Rual Domingos de SJ Rio Pardo | Instituto Adolfo Lutz, Interdiciplinary Procedures Center, Strategic Laboratory | Claudia Regina Gonçalves; Claudio Tavares Sacchi; Erica Valessa Ramos Gomes; Karoline Rodrigues Campos |
| EPI_ISL_471543, EPI_ISL_583500 | Centro de Saude I Tacito Leite de Carvalho e Silva | Instituto Adolfo Lutz, Interdiciplinary Procedures Center, Strategic Laboratory | Claudia Regina Gonçalves; Claudio Tavares Sacchi; Erica Valessa Ramos Gomes; Karoline Rodrigues Campos |
| EPI_ISL_735416 | Centro de Saude II Dr Jose Paione Mococa | Instituto Adolfo Lutz, Interdiciplinary Procedures Center, Strategic Laboratory | Claudia Regina Gonçalves; Claudio Tavares Sacchi; Erica Valessa Ramos Gomes; Karoline Rodrigues Campos |
| EPI_ISL_468305, EPI_ISL_468307, EPI_ISL_735411, EPI_ISL_735423, EPI_ISL_735424, EPI_ISL_735426 | Centro de Vigilancia a Saude de Diadema | Instituto Adolfo Lutz, Interdiciplinary Procedures Center, Strategic Laboratory | Claudia Regina Gonçalves; Claudio Tavares Sacchi; Erica Valessa Ramos Gomes; Karoline Rodrigues Campos |
| EPI_ISL_693240, EPI_ISL_693242 | Centro de Vigilância a Saude de Diadema | Instituto Adolfo Lutz, Interdiciplinary Procedures Center, Strategic Laboratory | Claudia Regina Gonçalves; Claudio Tavares Sacchi; Erica Valessa Ramos Gomes; Karoline Rodrigues Campos |
| EPI_ISL_450521, EPI_ISL_450524, EPI_ISL_639656, EPI_ISL_770052 | Centrālā Laboratorija | Latvian Biomedical Research and Study Centre | Ivars Silamiķelis; Jana Osīte; Jānis Kloviņš; Jānis Pjalkovskis; Kaspars Megnis; Marta Priedīte; Monta Ustinova; Stella Lapiņa; Uga Dumpis; Vita Rovīte; Ņikita Zrelovs |
| EPI_ISL_486419, EPI_ISL_492988, EPI_ISL_492989, EPI_ISL_501287, EPI_ISL_501808, EPI_ISL_501922, EPI_ISL_534210 | | | |
| see above | Centrālā laboratorija | Latvian Biomedical Research and Study Centre | Ivars Silamiķelis; Jana Osīte; Jānis Kloviņš; Jānis Pjalkovskis; Kaspars Megnis; Marta Priedīte; Monta Ustinova; Stella Lapiņa; Uga Dumpis; Vita Rovīte; Ņikita Zrelovs |
| EPI_ISL_729599, EPI_ISL_753820, EPI_ISL_753858, EPI_ISL_753875, EPI_ISL_753953, EPI_ISL_753981, EPI_ISL_753997, EPI_ISL_754191 | | | |
| see above | Charité Universitätsmedizin Berlin, Institut für Virologie/Labor Berlin | Charité Universitätsmedizin Berlin, Institut für Virologie | Barbara Mühlemann; Christian Drosten; Julia Schneider; Jörn Beheim-Schwarzbach; Talitha Veith; Terry Jones; Victor M Corman |
| EPI_ISL_468070, EPI_ISL_468075, EPI_ISL_468077, EPI_ISL_477132, EPI_ISL_625474, EPI_ISL_700336, EPI_ISL_700340, EPI_ISL_700342, EPI_ISL_700343, EPI_ISL_700345, EPI_ISL_700347 | | | |
| see above | Child Health Research Foundation | Child Health Research Foundation | Afroza Akter Tanni; Arif Mohammad Tanmoy; Hafizur Rahman; Maksuda Islam; Md Hafizur Rahman; Md Saiful Islam Sajib; Nikkon Sarkar; Roly Malaker; Samir K Saha; Senjuti Saha; Sharmistha Goswami; Syed Muktadir Al Sium |
| EPI_ISL_430736, EPI_ISL_430737, EPI_ISL_430738, EPI_ISL_430746 | Chinese PLA Institute for Disease Control and Prevention | Chinese PLA Institute for Disease Control and Prevention | Lizhong Li; Peng Li , Jinhui Li |
| EPI_ISL_414579 | Clinica Alemana de Santiago, Chile | Instituto de Salud Publica de Chile | Alejandra Acevedo; Andrés E. Castillo; Bárbara Parra; Carolina Tambley; Gabriel Leal; Gisselle Barra; Jaime Lagos; Javier Tognarelli; Jorge Fernández.; Loredana Arata; Patricia Bustos; Paz Tapia; Rodrigo Fasce; Soledad Ulloa; Winston Andrade |
| EPI_ISL_462450, EPI_ISL_462456, EPI_ISL_462461, EPI_ISL_462466, EPI_ISL_462470, EPI_ISL_462472, EPI_ISL_462476 | | | |
| see above | Clinical Center, University of Sarajevo | Charite Universitatsmedizin Berlin, Institute of Virology | Almedina Hadzihasanovic-Moro; Amela Dedeic-Ljubovic; Barbara Muehlemann; Christian Drosten; Irma Salimovic-Besic; Jorn Beheim-Schwarzbach; Julia Schneider; Selma Mutevelic; Suzana Arapcic; Talitha Veith; Terry Jones; Victor M Corman |
| EPI_ISL_677715, EPI_ISL_677718 | Clinical Hospital - Bitola | Research Center for Genetic Engineering and Biotechnology "Georgi D. Efremov" , Macedonian Academy of Sciences and Arts | RCGEB - MASA |
| EPI_ISL_677677, EPI_ISL_678250 | Clinical Hospital - Shtip | Research Center for Genetic Engineering and Biotechnology "Georgi D. Efremov" , Macedonian Academy of Sciences and Arts | RCGEB - MASA |
| EPI_ISL_424352 | Clinical Laboratory, Fuyang City Center for Disease Control and Prevention | Clinical Laboratory, Fuyang City Center for Disease Control and Prevention | Ge, B. |
| EPI_ISL_450506 | Clinical Laboratory, Hospital Israelita Albert Einstein | Clinical Laboratory, Hospital Israelita Albert Einstein | Amgarten, D.; Araujo; C.L.P.; D.B.; D.B.L.; Durigon; E.L. and Pinho; J.R.R.; Machado; Malta, F.; Mangueira; R.A.F.; R.R.G.; Santana; de Oliveira |
| EPI_ISL_416432 | Clinical Microbiology Lab | Infectious Disease Research Department, King Abdullah International Medical Research Center (KAIMRC) | Abdulrahman Alswaji; Liliane Okdah; Majed Alghoribi; Michel Doumith; Sadeem Alhayli; Sameera Al Johani |
| EPI_ISL_447465 | Clinical Microbiology Laboratory, Sheba Medical Center | Stern Lab | Stern Lab |
| EPI_ISL_490204 | Clinical Microbiology Laboratory- Basurto University Hospital | Biocruces-Bizkaia | Ana Belén Belén de la Hoz; Estibaliz Ugalde-Zarraga; José Luis Díaz de Tuesta del Arco; Matxalen Vidal-García; Mikel J. Urrutikoetxea-Gutierrez; Mº Carmen Nieto Toboso |
| EPI_ISL_447327 | Clinical Virology Laboratory, Soroka Medical Center and the Faculty of Health Sciences, Ben-Gurion University of the Negev | Stern Lab | Stern Lab |
| EPI_ISL_447353, EPI_ISL_447410 | Clinical Virology Unit, Hadassah Hebrew University Medical Center | Stern Lab | Stern Lab |
| EPI_ISL_632261, EPI_ISL_632262, EPI_ISL_632263, EPI_ISL_632284, EPI_ISL_632285, EPI_ISL_632904, EPI_ISL_681301, EPI_ISL_681302, EPI_ISL_681310, EPI_ISL_682299, EPI_ISL_682303, EPI_ISL_682304, EPI_ISL_682305, EPI_ISL_682319, EPI_ISL_682321 | | | |
| see above | Communicable Disease Laboratory, Public Health Directorate | Communicable Disease Laboratory, Public Health Directorate | AlAbbas, Z.; AlHujairi, Z.; AlTaif, Z.; AlWasti, H.; Altaif, Z.; Alwasti, H. |
| EPI_ISL_474921 | Complejo Hospitalario Universitario de Albacete | SeqCOVID-SPAIN consortium/IBV(CSIC) | Caridad Sainz de Baranda Camino and SeqCOVID-SPAIN consortium; Encarnacion Simarro Córdoba; Julia Lozano Serra; Lorena Robles Fonseca; Monica Parra Grandes |
| EPI_ISL_537380 | Complejo Hospitalario Universitario de Vigo | SeqCOVID-SPAIN consortium/IBV(CSIC) | Benito Regueiro and SeqCOVID-SPAIN consortium |
| EPI_ISL_583497 | Complexo Hospitalar Ouro Verde de Campinas | Instituto Adolfo Lutz, Interdiciplinary Procedures Center, Strategic Laboratory | Claudia Regina Gonçalves; Claudio Tavares Sacchi; Erica Valessa Ramos Gomes; Karoline Rodrigues Campos |
| EPI_ISL_523970 | Conjunto Hospitalar do Mandaqui | Instituto Adolfo Lutz, Interdiciplinary Procedures Center, Strategic Laboratory | Claudia Regina Gonçalves; Claudio Tavares Sacchi; Erica Valessa Ramos Gomes |
| EPI_ISL_696492 | Conville CDC wc CVC & NHLS/UCT | KRISP, KZN Research Innovation and Sequencing Platform | Arash Iranzadeh; Bruna Galvao; Carolyn Williamson; Deelan Doolabh; Diana Hardie; Emanuel James San; Houriiyah Tegally; Innocent Mudau; Jennifer Giandhari; Kruger Marais; Lynn Tyers; Marvin Hsiao; Stephen Korsman; Sureshnee Pillay; Tulio de Oliveira |
| EPI_ISL_735493 | Cox's Bazar Medicila College | Central Biological Research Laboratory and Department of Biochemistry and Molecular Biology Central Biological Research Laboratory and Department of Biochemistry and Molecular Biology | H. M. Abdullah Al Masud; Imam Hossen; Md. Arif Hossain; Md. Imranul Hoq; Md. Khondakar Raziur Rahman; Md. Omer Faruq; Mohammad Omar Faruque; Robiul Hasan Bhuiyan; Sajib Rudra; Shanta Paul |
| EPI_ISL_454602 | Croatian Institute of Public Health | University of Zagreb, Centre for research and knowledge transfer in biotechnology | Anamarija Slovic; Irena Tabain; Jelena Ivancic Jelecki; Tatjana Vilibic-Cavlek |
| EPI_ISL_693207 | Cs II Doutor Antonio Vicoso Moreira de Rezende | Instituto Adolfo Lutz, Interdiciplinary Procedures Center, Strategic Laboratory | Claudia Regina Gonçalves; Claudio Tavares Sacchi; Erica Valessa Ramos Gomes; Karoline Rodrigues Campos |
| EPI_ISL_476282, EPI_ISL_476288, EPI_ISL_476289, EPI_ISL_476297 | DB Diagnósticos do Brasil | Instituto de Medicina Tropical da Univesidade de São Paulo | Camila Alves Maia da Silva; Darlan da Silva Candido; Erika Regina Manuli; Ester Sabino; Flavia Cristina da Silva Sales; Giula Magalhaes Ferreira; Jaqueline Goes de Jesus; Julien Theze; Mariana Severo Ramundo; Nuno Faria; Samples: Nelson Gaburo Jr; Sequencing: Ingra Morales Claro; Thais de Moura Coletti |
| EPI_ISL_445215 | DNA Solution Ltd. | DNA Solution Ltd. | Abdul Khaleque; Abu Sufian; Hasan Ul Haider; Kazi Nadim Hasan; MSM Chowdhury; Mala Khan; Mamudul Hasan Razu; Md. Imran Khan; Mizanur Rahman; Mohammad Fazle Alam Rabbi; Mohammed Nafiz Imtiaz Polol |
| EPI_ISL_416542, EPI_ISL_421652 | Dasman Diabetes Institute | Dasman Diabetes Institute | Ebaa Al-Ozairi; Ebaa AlOzairi; Fahd Al-Mulla; Motasem Melhem; Qais Al-Duwairi; Rasheeba Iqbal; Sara Al-Qabandi; Sumi John |
| EPI_ISL_430796, EPI_ISL_430808 | Departamento de Biología y genética molecular, IACA Laboratorios. | Área de Secuenciación del Laboratorio de Virología del Hospital de Niños Dr. Ricardo Gutierrez on behalf of 'Proyecto Argentino Interinstitucional de genomica de SARS-CoV-2' (PAIS Consortium) | A; AS; E; Goya; LE; Lusso; MI; MS; Masciovecchio MV; Mistchenko; Nabaes Jodar; Natale; S; Streitenberger ER; Suárez; Tittarelli; Valinotto; Viegas, M. |
| EPI_ISL_603022, EPI_ISL_603034 | Departamento de Vigilância à Saúde | Instituto Adolfo Lutz, Interdiciplinary Procedures Center, Strategic Laboratory | Claudia Regina Gonçalves; Claudio Tavares Sacchi; Erica Valessa Ramos Gomes; Karoline Rodrigues Campos |
| EPI_ISL_516922, EPI_ISL_516924, EPI_ISL_516933 | Department for Molecular Diagnostics, Centre for Medical Microbiology, Institute of Public Health of Montenegro | Charité Universitätsmedizin Berlin, Institut für Virologie | Barbara Muehlemann; Christian Drosten; Julia Schneider; Jörn Beheim-Schwarzbach; Marija Govedarica and Danijela Vujošević; Talitha Veith; Terry Jones; Victor M Corman |
| EPI_ISL_420140, EPI_ISL_420144 | Department for Virology, Molecular Biology and Genome Research, R. G. Lugar Center for Public Health Research, National Center for Disease Control and Public Health (NCDC) of Georgia. | Department for Virology, Molecular Biology and Genome Research, R. G. Lugar Center for Public Health Research, National Center for Disease Control and Public Health (NCDC) of Georgia. | Adam Kotorashvili; Amiran Gamkrelidze; Amiran Gamkrelidze.; Ana Papkiauri; Ann Machablishvili; Anna Kasradze; Davit Tsaguria; Ekaterine Khmaladze; Ekaterine Zangaladze; Ekaterine Zhghenti; Giorgi Tomashvili; Gvantsa Brachveli; Gvantsa Chanturia; Irma Burjanadze; Ketevan Sidamonidze; Khatuna Zakhashvili; Lela Sabadze; Lela Urushadze; Magda Dgebuadze; Maia Alkhazashvili; Mari Gavashelidze; Mariam Zakalashvili; Marine Murtskhvaladze; Meri Pantsulaia; Nato Kotaria; Nino Berishvili; Paata Imnadze; Roena Sukhiashvili; Tamar Jashiashvili; Tata Imnadze; Tea Tevdoradze |
| EPI_ISL_447055, EPI_ISL_470876, EPI_ISL_470877, EPI_ISL_481483, EPI_ISL_754180, EPI_ISL_754181, EPI_ISL_763062 | | | |
| see above | Department for Virology, Molecular Biology and Genome Research, R. G. Lugar Center for Public Health Research, National Center for Disease Control and Public Health (NCDC) of Georgia. | Department for Virology, Molecular Biology and Genome Research, R. G. Lugar Center for Public Health Research, National Center for Disease Control and Public Health (NCDC) of Georgia. | Adam Kotorashvili; Amiran Gamkrelidze.; Ana Papkiauri; Ann Machablishvili; Anna Kasradze; Davit Tsaguria; Ekaterine Khmaladze; Ekaterine Zangaladze; Ekaterine Zhghenti; Giorgi Tomashvili; Gvantsa Brachveli; Gvantsa Chanturia; Irma Burjanadze; Ketevan Sidamonidze; Khatuna Zakhashvili; Lela Sabadze; Lela Urushadze; Magda Dgebuadze; Maia Alkhazashvili; Mari Gavashelidze; Mariam Zakalashvili; Marine Murtskhvaladze; Meri Pantsulaia; Nato Kotaria; Nino Berishvili; Paata Imnadze; Roena Sukhiashvili; Salome Javashvili; Tamar Jashiashvili; Tata Imnadze; Tea Tevdoradze |

| | | | |
|---|---|---|---|
| EPI_ISL_632934 | Department of Acute Infectious Diseases Control and Prevention,Yunnan Provincial Center for Disease Control and Prevention | Department of Acute Infectious Diseases Control and Prevention,Yunnan Provincial Center for Disease Control and Prevention | Jienan Zhou; Meiling Zhang; Senquan Jia; Xiaonan Zhao; Xiaoqing Fu |
| EPI_ISL_515084, EPI_ISL_515098 | Department of Biochemistry, Cell and Molecular Biology | WACCBIP, University of Ghana | A.K.; Adu, B.; Amenga-Etego; Ampofo, W.; Amuzu; Anang; Arjarquah, A.; Asante, I.; Awandare; Bediako, Y.; Boatemaa, L.; Bonney, E.; Bonney, K.; C.M.; D.S.; Eshun, M.; G.A.; G.B.; J.K.; J.M.; Kotey, E.; Kumordjie, S.; Kyei; L.N.; Magnussen, V.; Morang'a; Mutungi; Ngoi; Quashie, P.; Tei-Maya, F. |
| EPI_ISL_429239 | Department of Clinical Laboratory, the First People's Hospital of Yunnan Province | Department of Clinical Laboratory, the First People's Hospital of Yunnan Province | Guiqian Zhang; Xin Fan; Ya Xu; Yi Sun; Yu Zhang; Ziqin Dian |
| EPI_ISL_417020, EPI_ISL_418659, EPI_ISL_455968, EPI_ISL_498149, EPI_ISL_515078, EPI_ISL_540487, EPI_ISL_540551, EPI_ISL_540574, EPI_ISL_581613, EPI_ISL_581629, EPI_ISL_581658, EPI_ISL_644619, EPI_ISL_678399, EPI_ISL_707729, EPI_ISL_722937, EPI_ISL_722965, EPI_ISL_737372 | | | |
| see above | Department of Clinical Microbiology | GIGA Medical Genomics | Artesi Maria; Axelle Chaslain; Bontems Sébastien; Boreux Raphaël; Bouchra Boujemla; Bours Vincent.; Cecile Meex; Celine Fombellida-Lopez; Cécile Meex; Céline Fombellida-Lopez; Durkin Keith; Hayette Marie-Pierre; Keith Durkin; Maria Artesi; Marie-Pierre Hayette; Meex Cécile; Melin Pierrette; Pierrette Melin; Raphael Boreux; Raphaël Boreux; Sebastien Bontems; Sébastien Bontems; Vincent Bours; Vincent Bours. |
| EPI_ISL_417193, EPI_ISL_419245, EPI_ISL_419250 | Department of Clinical Pathology, Pamela Youde Nethersole Eastern Hospital | Department of Health Technology and Informatics, Faculty of Health and Social Science, The Hong Kong Polytechnic University | Alan Ka-Lun WU; Alex Yat-Man HO; Barry Kin-Chung WONG; David Ho-Keung SHUM; Eugene Yuk-Keung TSO; Gilman Kit-Hang SIU; Hiu-Yin LAO; Kam-Tong YIP; Kam-Tong Yip; Kenneth Siu-Sing LEUNG; Kingsley King-Gee TAM; Kit-Man SIN; Kitty Sau-Chun FUNG; Kwok-Cheung LUNG; Lam-Kwong LEE; Man-Chun CHAN; Ming-Pan CHOI; Miranda Chong-Yee YAU; Raymond Wai-To LIU; Sandy Ka-Yee CHAU; Shea Ping YIP; Tak-Lun QUE; Tak-Lun Que; Timothy Ting-Leung NG; Wai-Shing LEUNG; Wing Cheong YAM; Wing-Kin TO; Yuk-Yung NG |
| EPI_ISL_481260 | Department of Emerging Infectious Diseases, Institute of Tropical Medicine, Nagasaki University | Department of Emerging Infectious Diseases, Institute of Tropical Medicine, Nagasaki University | Haruka Abe; Jiro Yasuda; Rokusuke Yoshikawa; Yuichiro Furusato |
| EPI_ISL_507206 | Department of Experimental Modeling and Pathogenesis of Infectious Diseases | WHO National Influenza Centre Russian Federation | Andrey Komissarov; Anna Ivanova; Artem Fadeev; Daria Danilenko; Mariia Sergeeva |
| EPI_ISL_610169, EPI_ISL_610205, EPI_ISL_610208, EPI_ISL_610209, EPI_ISL_610217, EPI_ISL_610218, EPI_ISL_610225 | | | |
| see above | Department of Health Technology and Informatics, The Hong Kong Polytechnic University | Department of Health Technology and Informatics, The Hong Kong Polytechnic University | A.K.-L.; A.Y.-M.; B.K.-C.; C.T.-M.; Chan; Chau; D.H.-K.; Fung; G.K.-H.; H.-Y.; Ho; J.S.-L.; K.-T.; K.K.-G.; K.S.-C.; K.S.-S.; L.-K.; Lai; Lao; Lee; Leung; Luk, K.; M.C.-Y.; Ng; Que; S.K.-Y.; S.P.; Shum; Siu; T.-L.; T.T.-L.; Tam; To; W.-K.; W.C.; Wong; Wu; Y.W.-M.; Yam; Yau; Yip |
| EPI_ISL_417444 | Department of Healthcare Biotechnology, National University of Sciences and Technology (NUST) | Department of Healthcare Biotechnology, National University of Sciences and Technology (NUST) | Corman; Ghani, E.; H.A.; Janjua; Javed, A.; Niazi; S.K.; Saqib, M.; V.M. and Zohaib, A. |
| EPI_ISL_568574, EPI_ISL_637016, EPI_ISL_637018, EPI_ISL_637019, EPI_ISL_637020, EPI_ISL_637021 | Department of Infectious Diseases and Immunology, National Hospital Organization Nagoya Medical Center | Clinical Research Center, National Hospital Organization Nagoya Medical Center | Hirotaka Ode; Kazuhiro Matsuoka; Mai Kubota; Masakazu Matsuda; Mayumi Imahashi; Miho Nakasuji; Mikiko Mori; Nakasuji Miho; Yasumasa Iwatani; Yoshihiro Nakata; Yoshiyuki Yokomaku |
| EPI_ISL_412974 | Department of Infectious Diseases, Istituto Superiore di Sanità, Rome, Italy | Virology Laboratory, Scientific Department, Army Medical Center | Andrea Ciammaruconi; Antonella Fortunato; Antonella Marchi; Concetta Fabiani; Eleonora Benedetti; Filippo Molinari; Florigio Lista; Giancarlo Petralito; Giovanni Faggioni; Paola Stefanelli; Riccardo De Santis; Silvia Fillo; Stefano Fiore |
| EPI_ISL_644945, EPI_ISL_644948, EPI_ISL_644951, EPI_ISL_644954, EPI_ISL_644955, EPI_ISL_644961, EPI_ISL_644964, EPI_ISL_644970, EPI_ISL_644980, EPI_ISL_644993, EPI_ISL_644997, EPI_ISL_644999 | | | |
| see above | Department of Infectious Diseases, Keio University School of Medicine, Tokyo, Japan | Center for Medical Genetics, Keio University School of Medicine, Tokyo, Japan | Haruhiko Siomi; Hirotsugu Ishizu; Kenjiro Kosaki; Kodai Abe; Yuka Iwasaki |
| EPI_ISL_480100, EPI_ISL_480211 | Department of Infectious Diseases, Kobe Institute of Health | Pathogen Genomics Center, National Institute of Infectious Diseases | Hajime Kamiya; Kentaro Itokawa; Makoto Kuroda; Masanori Hashino; Motoi Suzuki; Rina Tanaka; Ryohei Nomoto; Tsuyoshi Sekizuka |
| EPI_ISL_411219 | Department of Infectious and Tropical Diseases, Bichat Claude Bernard Hospital, Paris | Laboratoire Virpath, CIRI U111, UCBL1, INSERM, CNRS, ENS Lyon | Alexandre Gaymard; Aurélien Traversier; Bruno Lina; Julien Fouret; Manuel Rosa-Calatrava; Olivier Terrier; Xavier Lescure; Yazdan Yazdanpanah |
| EPI_ISL_479490 | Department of Laboratory Medicine Tan Tock Seng Hospital | Department of Laboratory Medicine Tan Tock Seng Hospital | Barkham TMS; Chen YYC; Li C; Maurer-Stroh S; Nagarajan N; Sessions OM; Tang WY; Zair X |
| EPI_ISL_422422, EPI_ISL_447622, EPI_ISL_534336, EPI_ISL_693302, EPI_ISL_693305, EPI_ISL_738064 | Department of Laboratory Medicine, National Taiwan University Hospital | Microbial Genomics Core Lab, National Taiwan University Centers of Genomic and Precision Medicine | Chiao-Ling Li; Pei-Jer Chen; Shan-Chwen Chang; Shiou-Hwei Yeh; Sui-Yuan Chang; Ya-Yun Lai; You-Yu Lin |
| EPI_ISL_476819, EPI_ISL_538460, EPI_ISL_648094, EPI_ISL_648662, EPI_ISL_648717, EPI_ISL_648740, EPI_ISL_648807 | | | |
| see above | Department of Laboratory Medicine, Tan Tock Seng Hospital | Department of Laboratory Medicine, Tan Tock Seng Hospital | Barkham TMS; Chen YYC; Li C; Lim JX; Maurer-Stroh S; Nagarajan N; Sessions OM; Tang WY; Zair X |
| EPI_ISL_796681 | Department of Medical Microbiology - section Molde, Molde Hospital | Norwegian Institute of Public Health, Department of Virology | Atiya R Ali; Hilde Elshaug; Hilde Vollan; Kamilla Heddeland Instefjord; Karoline Bragstad; Kathrine Stene-Johansen; Marie Paulsen Madsen; Olav Hungnes; Rasmus Riis Kopperud |
| EPI_ISL_501178, EPI_ISL_501200, EPI_ISL_501206, EPI_ISL_501220, EPI_ISL_501221, EPI_ISL_501222 | Department of Medical Microbiology, University Malaya Medical Centre | Department of Medical Microbiology, Faculty of Medicine, University of Malaya | I-Ching SAM; Jennifer Chong; University Malaya Medical Centre COVID Team; Yoke Fun CHAN; Yoong Min CHONG |
| EPI_ISL_512844 | Department of Medical Research | DMR_Myanmar | Aung Kyaw Kyaw; Aung Zaw Latt; Hlaing Myat Thu; Hnin Ohnmar Soe; Htin Lin; Kay Thi Aye; Lai Lai San; Myat Htut Nyunt; Nan Aye Thida Oo; Ni Ni Zaw; Phyu Win Ei; Su Mon Win; Theingi Win Myat; Wah Wah Aung; Yi Yi Kyaw; Zaw Than Htun |
| EPI_ISL_528813 | Department of Medicine, Gandhi hospital, Hyderabad | CSIR-Centre for Cellular and Molecular Biology | Ajay Sarawagi; Archana Bharadwaj Siva; Dhiviya Vedagiri; Divya Gupta; Divya Tej Sowpati; Karthik Bharadwaj Tallapaka; Krishnan Harinivas Harshan; Lamuk Zaveri; M Soujanya Reddy; Namami Gaur; Nikhil Hajirnis; Onkar Kulkarni; Payel Mukherjee; Pratheusa Maccha; Priya Singh; Priyanka Pant; Purushotham Vodnala; Rajarao Mesipogu; Rajkanwar Nathawat; Rakesh K Mishra; Sakshi Shambhavi; Santosh Kumar Kuncha; Shagufta Khan; Sofia Banu; Thrilok Chander Bingi; Tulasi Nagabandi; Umesh Kumar; Unis Ahmad Bhat; Vinayasekhar Aedula; Vishal Sah |
| EPI_ISL_483874 | Department of Microbiology, Government Medical College, Surat | Gujarat Biotechnology Research Centre | A M Kadri; Afzal Ansari; Amit gamit; Apurvasinh Puvar; Chaitanya Joshi; Dinesh Kumar; Harsh Bakshi; Janvi Raval; Komal Patel; Labdhi Pandya; Madhvi Joshi; Maharshi Pandya; Monika Gandhi; Naresh Chauhan; Nidhi Patel; Nikha Trivedi; Nitin Savaliya; Pinal Trivedi; R D Dixit; Raghawendra Kumar; Summaiya Mullan; Zarna Patel; Zuber Saiyed |
| EPI_ISL_497770, EPI_ISL_497771, EPI_ISL_497778, EPI_ISL_497786, EPI_ISL_497791, EPI_ISL_497818, EPI_ISL_497865, EPI_ISL_498270, EPI_ISL_498271 | | | |
| see above | Department of Microbiology, The University of Hong Kong | Department of Microbiology, The University of Hong Kong | Kelvin K.W. To; Kwok-Yung Yuen |
| EPI_ISL_693299 | Department of Microbiology, Yokohama City University School of Medicine | Department of Microbiology, Yokohama City University School of Medicine | Akihide Ryo; Ichiro Takeuchi; Kazuya Sakai; Kei Miyakawa; Mototsugu Nishii; Reo Matsumura; Ryo Saji |
| EPI_ISL_463741, EPI_ISL_463743, EPI_ISL_463744, EPI_ISL_463745, EPI_ISL_463746, EPI_ISL_463747, EPI_ISL_463748 | | | |
| see above | Department of Molecular Virology, Cyprus Institute of Neurology and Genetics | Department of Molecular Virology, Cyprus Institute of Neurology and Genetics | Christina Christodoulou; Christina Tryfonos; Dana Koptides; George Krashias; Jan Richter; Stavros Bashiardes |
| EPI_ISL_582033, EPI_ISL_594186, EPI_ISL_594187, EPI_ISL_594188, EPI_ISL_596451 | Department of Pathology, School of Medicine, Imam Khomeini Hospital, Tehran University of Medical Sciences | Genetics Research Center, University of Social Welfare and Rehabilitation Sciences | Ali Jafarpour; Alireza Abdollahi; Azam Ghaziasadi; Azar Hadadi; Hossein Najmabadi; Khadijeh Jalalvand; Kimia Kahrizi; Marzieh Mohseni; Reza Najafipour; Saber Soltani; Seyed Mohammad Jazayeri; Seyedeh elham Mortazavi; Zohreh Fattahi |
| EPI_ISL_452944, EPI_ISL_456731 | Department of Pathology, University of Cambridge | COVID-19 Genomics UK (COG-UK) Consortium | Aminu S. Jahun; Anna Yakovleva; Charlotte J. Houldcroft; Fahad A Khokhar; Grant Hall; Ian Goodfellow; Laura G Caller; Luke W Meredith; M. Estée Török; Martin D. Curran; Myra Hosmillo; Sarah L. Caddy; Theresa Feltwell; William L. Hamilton |
| EPI_ISL_576147, EPI_ISL_576149 | Department of Respiratory & Other Viral Infections of L.V. Gromashevsky Institute of Epidemiology & Infectious Diseases NAMS of Ukraine | Department of Respiratory & Other Viral Infections of L.V. Gromashevsky Institute of Epidemiology & Infectious Diseases NAMS of Ukraine, JSC "Farmak" | Alla Mironenko; Andriy Goy; Ihor Kravchuk; Larysa Radchenko; Liudmyla Bolotova; Nataliia Teteriuk |
| EPI_ISL_654820 | Department of Respiratory and other Viral Infections of L.V.Gromashevsky Institute of Epidemiology & Infectious Diseases NAMS of Ukraine | Department of Respiratory and other Viral Infections of L.V.Gromashevsky Institute of Epidemiology & Infectious Diseases NAMS of Ukraine, JSC "Farmak" | Alla Mironenko; Andriy Goy; Ihor Kravchuk; Larysa Radchenko; Ludmyla Bolotova; Nataliia Teteriuk |
| EPI_ISL_732952 | Department of Tropical Parasitology | Laboratory of Recombinant Vaccines | Boguslaw Szewczyk; Kirsi Aaltonen; Lukasz Rabalski; Maciej Grzybek; Maciej Kosinski; Ravi Kant; Tarja Sironen; Teemu Smura |
| EPI_ISL_487275 | Department of Veterinary Pathology, University of Liege - FARAH | Department of Veterinary Pathology, University of Liege - FARAH | A.-S.; Clercx, C.; Desmecht, D.; Eloit, M.; Escriou, N.; Garigliany, M.; Giet, D.; Huon, C.; Vanlaere; van der Werf, S. |
| EPI_ISL_450198, EPI_ISL_450203, EPI_ISL_450208 | Department of Virology | Department of Virology | Ackermann, N.; Antwerpen, M.; Bengs, K.; Berger, A.; Boehm, S.; Boehmer; Boender; Buchholz, U.; Cai, W.; Corman; D.V.; Dangel, A.; Drosten, C.; Eberle, U.; Fingerle, V.; Grahl, A.; Haas, W.; Hamouda, O.; Hoch, M.; Hoermansdorfer, S.; Ippisch, S.; Jones; Katz, K.; Konrad, R.; Liebl, B.; M.M.; Marosevic; Muehlemann, B.; Muller, N.; Poertner, K.; Protzer, U.; Reich, A.; Rexroth, U.; Schneider, J.; Sing, A.; T.C.; T.S.; Treis, B.; V.M.; Veith, T.; Walter, M.; Wicklein, B.; Woelfel, R.; Woudenberg, T.; Zapf, A.; Zeitlmann, N.; an der Heiden, M. |
| EPI_ISL_413603, EPI_ISL_414641, EPI_ISL_481520, EPI_ISL_481557, EPI_ISL_481565, EPI_ISL_481570, EPI_ISL_481635, EPI_ISL_481662, EPI_ISL_481694, EPI_ISL_481709, EPI_ISL_481724, EPI_ISL_732582, EPI_ISL_732585, EPI_ISL_732649, EPI_ISL_737210, EPI_ISL_737216, EPI_ISL_755966, EPI_ISL_756012, EPI_ISL_756089, EPI_ISL_756099, EPI_ISL_756173, EPI_ISL_756247, EPI_ISL_757310, EPI_ISL_757331, EPI_ISL_757366, EPI_ISL_757368, EPI_ISL_757379, EPI_ISL_757384, EPI_ISL_757388, EPI_ISL_759833, EPI_ISL_759935 | | | |
| see above | Department of Virology and Immunology, University of Helsinki and Helsinki University Hospital, Huslab Finland | Department of Virology, Faculty of Medicine, University of Helsinki, Helsinki, Finland | Hannimari Kallio-Kokko; Harri Kangas; Hussein Alburkat; Jenni Virtanen; Maija Suvanto; Olli Vapalahti; Pekka Ellonen; Phuoc Truong; Ravi Kant; Sari Hannula; Teemu Smura |
| EPI_ISL_669290, EPI_ISL_670186, EPI_ISL_671247, EPI_ISL_682642, EPI_ISL_683218, EPI_ISL_683270, EPI_ISL_712349, EPI_ISL_714501, EPI_ISL_748296, EPI_ISL_758976, EPI_ISL_794396, EPI_ISL_795473 | | | |

| | | | |
|---|---|---|---|
| see above | Department of Virus and Microbiological Special Diagnostics, Statens Serum Institut, Copenhagen, Denmark | Albertsen Lab, Department of Chemistry and Bioscience, Aalborg University, Denmark | Danish Covid-19 Genome Consortium |
| EPI_ISL_641516 | Department of Virus and Microbiological Special Diagnostics, Statens Serum Institut, Copenhagen, Denmark | Albertsen lab, Department of Chemistry and Bioscience, Aalborg University, Denmark | Jannik Fonager; Morten Rasmussen; Thomas Bruun Rasmussen |
| EPI_ISL_429466, EPI_ISL_429582, EPI_ISL_444854, EPI_ISL_444956 | Department of Virus and Microbiological Special Diagnostics, Statens Serum Institut, Copenhagen, Denmark, Artillerivej 5, 2300 Copenahgen S | Albertsen lab, Department of Chemistry and Bioscience, Aalborg University, Denmark | Rasmus Kirkegaard |
| EPI_ISL_614743, EPI_ISL_614753, EPI_ISL_614771, EPI_ISL_614828, EPI_ISL_615386, EPI_ISL_615592, EPI_ISL_616349, EPI_ISL_616496, EPI_ISL_616613, EPI_ISL_616986, EPI_ISL_617737, EPI_ISL_618008, EPI_ISL_618032, EPI_ISL_618198, EPI_ISL_618242, EPI_ISL_618277, EPI_ISL_618343, EPI_ISL_618597, EPI_ISL_619344, EPI_ISL_619897, EPI_ISL_619978, EPI_ISL_620403, EPI_ISL_620904, EPI_ISL_621885, EPI_ISL_622413, EPI_ISL_622434, EPI_ISL_622600, EPI_ISL_622727 | | | |
| see above | Department of Virus and Microbiological Special Diagnostics, Statens Serum Institut, Denmark | Albertsen lab, Department of Chemistry and Bioscience, Aalborg University, Denmark | Danish Covid-19 Genome Consortia |
| EPI_ISL_416143 | Department of Virus and Microbiological Special diagnostics, Statens Serum Institut, Copenhagen, Denmark. | ViFU | Anders Fomsgaard; Maiken Worsoe Rosenstierne; Morten Rasmussen |
| EPI_ISL_449793, EPI_ISL_590960 | Dept. of Medical Microbiology, Stavanger University Hospital, Helse Stavanger HF | Norwegian Institute of Public Health, Department of Virology | Hilde Elshaug; Hilde Vollan; Iren Löhr; Kamilla Heddeland Instefjord; Karoline Bragstad; Kathrine Stene-Johansen; Olav Hungnes; Rasmus Riis Kopperud |
| EPI_ISL_665254, EPI_ISL_666593, EPI_ISL_666595, EPI_ISL_666602, EPI_ISL_666612 | Dept. of Microbiology and Infection Control, Akershus University Hospital HF | Dept. of Microbiology and Infection Control, Akershus University Hospital HF | Alexander Hesselberg Løvestad; Hege Vangstein Aamot; Nina Handal; Ole Herman Ambur; Silje Bakken Jørgensen |
| EPI_ISL_410532 | Dept. of Pathology, National Institute of Infectious Diseases | Pathogen Genomics Center, National Institute of Infectious Diseases | Harutaka Katano; Hideki Hasegawa; Kazuya Shirato; Makoto Kuroda; Makoto Takeda; Motoi Suzuki; Naganori Nao; Shutoku Matsuyama; Tadaki Suzuki; Takaji Wakita; Tsuyoshi Sekizuka |
| EPI_ISL_408667 | Dept. of Virology III, National Institute of Infectious Diseases | Pathogen Genomics Center, National Institute of Infectious Diseases | Kazuya Shirato; Makoto Kuroda; Makoto Takeda; Naganori Nao; Shutoku Matsuyama; Tsuyoshi Sekizuka |
| EPI_ISL_774935, EPI_ISL_774992, EPI_ISL_775013 | Designated Reference Institute for Chemical Measurements (DRICM) | DNA SOLUTION LTD. | Abdul Khaleque; Abu Sufian; Hasan Ul Haider; Jannatun Naima; Kazi Nadim Hasan; MSM Chowdhury; Mala Khan; Mamudul Hasan Razu; Md. Imran Khan; Mizanur Rahman; Mohammad Fazle Alam Rabbi |
| EPI_ISL_699506, EPI_ISL_699507, EPI_ISL_699508, EPI_ISL_699509 | Diagnostic Virology Laboratory, USDA National Veterinary Services Laboratories | Diagnostic Virology Laboratory, USDA National Veterinary Services Laboratories | Akpalu, Y.; Auckland; Barton Behravesh, C.; C.M.; Davila, E.; E.R.; Fischer; Franzen; G.L.; Ghai; Hamer; I.B.; Jenkins-Moore, M.; K.M.; Killian; L.D.; Love; M.L.; Pauvolid-Correa, A.; R.R.; R.S.; Roundy; S.A.; Spengler, J.; Tang, W.; Torchetti, M.; Zecca |
| EPI_ISL_653916 | Diagnostic- and Research Institute of Pathology, Medical University of Graz | Diagnostic- and Research Institute of Pathology, Medical University of Graz | Gregor Gorkiewicz; Karl Kashofer; Martin Zacharias; Peter Regitnig |
| EPI_ISL_475562 | Din Klinik | The Public Health Agency of Sweden | Anna Risberg; Anna-Malin Linde; Karin Tegmark-Wisell; Maria Lind Karlberg; Mattias Haukland; Mia Brytting; Olov Svartstrom; Oskar Karlsson Lindsjo; Reza Advani; Sandra Broddesson |
| EPI_ISL_722855, EPI_ISL_722897 | Dipartimento di Scienze Biomediche e Oncologia Umana - Azienda Ospedaliero Universitaria Consorziale Policlinico | Istituto Zooprofilattico Sperimentale della Puglia e della Basilicata | Bianco A.; Capozzi L.; Chironna M.; Del Sambro L.; Loconsole D.; Parisi A. |
| EPI_ISL_534312 | Distrito Sanitario Sul | Instituto Adolfo Lutz, Interdiciplinary Procedures Center, Strategic Laboratory | Claudia Regina Gonçalves; Claudio Tavares Sacchi; Erica Valessa Ramos Gomes |
| EPI_ISL_583499 | Distrito Sanitario Sul Campinas | Instituto Adolfo Lutz, Interdiciplinary Procedures Center, Strategic Laboratory | Claudia Regina Gonçalves; Claudio Tavares Sacchi; Erica Valessa Ramos Gomes; Karoline Rodrigues Campos |
| EPI_ISL_747251, EPI_ISL_747328, EPI_ISL_747337, EPI_ISL_747349, EPI_ISL_747375, EPI_ISL_747384, EPI_ISL_747444, EPI_ISL_759993, EPI_ISL_760004, EPI_ISL_760005, EPI_ISL_760026, EPI_ISL_760113, EPI_ISL_760124, EPI_ISL_760186, EPI_ISL_760195, EPI_ISL_760199, EPI_ISL_760220 | | | |
| see above | Division of Emerging Infectious Diseases, Bureau of Infectious Diseases Diagnosis Control, Korea Disease Control and Prevention Agency | Division of Emerging Infectious Diseases, Bureau of Infectious Diseases Diagnosis Control, Korea Disease Control and Prevention Agency | Ae Kyung Park; Chaeyoung Lee; Eun-Jin Kim; Heui Man Kim; Il-Hwan Kim; Jeong-Min Kim; Namjoo Lee; Sang Hee Woo |
| EPI_ISL_425118, EPI_ISL_426182, EPI_ISL_497955, EPI_ISL_497961, EPI_ISL_497998, EPI_ISL_498001, EPI_ISL_498019, EPI_ISL_498048, EPI_ISL_506993, EPI_ISL_506995, EPI_ISL_510562, EPI_ISL_510594, EPI_ISL_510608, EPI_ISL_510651, EPI_ISL_514788, EPI_ISL_514828, EPI_ISL_514914, EPI_ISL_514925, EPI_ISL_514960, EPI_ISL_515017, EPI_ISL_526713 | | | |
| see above | Division of Viral Diseases, Center for Laboratory Control of Infectious Diseases, Korea Centers for Diseases Control and Prevention | Division of Viral Diseases, Center for Laboratory Control of Infectious Diseases, Korea Centers for Diseases Control and Prevention | Daesang Lee; Dong Hyun Song; Heui Man Kim; Hye-Jun Jo; Jeong-Min Kim; Jun-Sub Kim; Junhyeong Jang; Mi-Seon Kim; Myung Guk Han; Namjoo Lee; Sang Hee Woo; Sehee Park; Seong Tae Jeong; Yoon-Seok Chung |
| EPI_ISL_722209 | Dom Zdravlja Sarajevo | Alea Genetic Center | Konjhodzic R.; Pecar D.; Salihefendic L. |
| EPI_ISL_700552 | Dr Abdurahman CDC wc DAC | NHLS/UCT | Arash Iranzadeh; Bruna Galvao; Carolyn Williamson; Deelan Doolabh; Diana Hardie; Innocent Mudau; Kruger Marais; Lynn Tyers; Marvin Hsiao; Stephen Korsman |
| EPI_ISL_583735, EPI_ISL_583755 | Dr. Gernot Walder GmbH | Bergthaler laboratory, CeMM Research Center for Molecular Medicine of the Austrian Academy of Sciences | Adi Steinrigl; Alexander Lercher; Alexandra Popa; Andreas Bergthaler; Benedikt Agerer; Christian Paar; Christoph Bock; Daniela Schmid; Dorothee von Laer; Elisabeth Puchhammer-Stoeckl; Franz Allerberger; Gernot Walder; Gregor Hörmann; Guenter Weiss; Gunther Vogl; Henrique Colaco; Jakob-Wendelin Genger; Jan Laine; Judith Aberle; Kinga Rigler-Hohenwarter; Lukas Endler; Manfred Nairz; Mark Smyth; Martin Senekowitsch; Michael Schuster; Peter Hufnagl; Peter Obrist; Rainer Gattringer; Sabine Sussitz-Rack; Stephan Aberle; Thomas Penz; Wegene Borena |
| EPI_ISL_415530, EPI_ISL_422678, EPI_ISL_455175, EPI_ISL_460953, EPI_ISL_461018, EPI_ISL_461074, EPI_ISL_461129, EPI_ISL_461189, EPI_ISL_522995, EPI_ISL_523023, EPI_ISL_523046, EPI_ISL_523051, EPI_ISL_523058, EPI_ISL_523084, EPI_ISL_523086, EPI_ISL_523093, EPI_ISL_523094, EPI_ISL_523101, EPI_ISL_523109, EPI_ISL_523147, EPI_ISL_523158, EPI_ISL_523273, EPI_ISL_523366, EPI_ISL_523413, EPI_ISL_523546, EPI_ISL_523598, EPI_ISL_523618, EPI_ISL_523686, EPI_ISL_523740, EPI_ISL_574789, EPI_ISL_577749, EPI_ISL_577770, EPI_ISL_577774, EPI_ISL_577783, EPI_ISL_577791, EPI_ISL_577810, EPI_ISL_577825, EPI_ISL_577939, EPI_ISL_577970, EPI_ISL_632436, EPI_ISL_632451, EPI_ISL_632453, EPI_ISL_632492, EPI_ISL_632503, EPI_ISL_632530, EPI_ISL_632595, EPI_ISL_632774, EPI_ISL_632777, EPI_ISL_722448, EPI_ISL_722534, EPI_ISL_722583, EPI_ISL_722734, EPI_ISL_722814, EPI_ISL_722842, EPI_ISL_763189, EPI_ISL_763244, EPI_ISL_801506, EPI_ISL_802533 | | | |
| see above | Dutch COVID-19 response team | Erasmus Medical Center | Anne van der Linden; Annemiek van der Eijk; Aura Timen; Bas Oude Munnink; Claudia Schapendonk; Corien Swaan; Corine GeurtsvanKessel; David Nieuwenhuijse; Emmanuelle Munger; Irina Chestakova; Jeroen van Kampen; Jolanda Voermans; Madelief Mollers; Manon Haverkate; Marion Koopmans; Marjan Boter; Mark Pronk; Mart Stein; OH consortium; Pascal Lexmond; Reina Sikkema; Richard Molenkamp; Sandra Kengne Kamga Mobou; Stefan van Nieuwkoop; Theo Bestebroer; on behalf of the Dutch national COVID-19 response team. |
| EPI_ISL_547445, EPI_ISL_547449, EPI_ISL_547450, EPI_ISL_547453, EPI_ISL_636500, EPI_ISL_636514, EPI_ISL_636517, EPI_ISL_636518, EPI_ISL_636519, EPI_ISL_636585, EPI_ISL_636603, EPI_ISL_723189, EPI_ISL_723396, EPI_ISL_723402, EPI_ISL_790986 | | | |
| see above | Dutch COVID-19 response team | National Institute for Public Health and the Environment (RIVM) | Adam Meijer; AnneMarie van den Brandt; Bas van der Veer; Chantal Reusken; Dennis Schmitz; Florian Zwagemaker; Harry Vennema; Jeroen Cremer; Sharon van den Brink; on behalf of the national COVID-19 response team |
| EPI_ISL_426287, EPI_ISL_450519, EPI_ISL_501894, EPI_ISL_512313, EPI_ISL_534199, EPI_ISL_639638, EPI_ISL_639684, EPI_ISL_639686, EPI_ISL_770053 | | | |
| see above | E. Gulbja Laboratorija | Latvian Biomedical Research and Study Centre | Dmitrijs Perminovs; Ivars Silamiķelis; Jānis Kloviņš; Jānis Pjalkovskis; Kaspars Megnis; Mikus Gavars; Monta Ustinova; Uga Dumpis; Vita Rovīte; Ņikita Zrelovs |
| EPI_ISL_486438 | E. Gulbja laboratorija | Latvian Biomedical Research and Study Centre | Dmitrijs Perminovs; Ivars Silamiķelis; Jānis Kloviņš; Kaspars Megnis; Mikus Gavars; Monta Ustinova; Uga Dumpis; Vita Rovīte; Ņikita Zrelovs |
| EPI_ISL_593498, EPI_ISL_593502 | Eastern Ontario Regional Laboratory Association | McMaster University | Ahmed Draia; Andrew G. McArthur; Emily Panousis; Hooman Derakhshani; Jalees Nasir; Leanne Mortimer; Robert Slinger |
| EPI_ISL_468055, EPI_ISL_479711, EPI_ISL_529144 | Egyptian National Cancer Institute (ENCI) | Egyptian National Cancer Institute (ENCI) | A.A.; Abdel Rahman N; Abdel Rahman N.; Abdelhamid, W.; Abouelhoda; Ahmed; Ali; Ali, M.; Amer; Bahnassy; Elkhateeb; Elsissy; Ezzelarab; Gad, A.; H.K.; Hafez; Hamdy; Hassan; Hassan, W.; K.E.; Khattab; M.A.; M.H.; M.M.; M.S.; Mahmoud; Mohamed; O.S.; Raouf, A.; S.M.; Samir, M.; Soliman; W.A.; Zekri |
| EPI_ISL_710611 | Ektorps Vardcentral | The Public Health Agency of Sweden | Department of Microbiology; The Public Health Agency of Sweden |
| EPI_ISL_450873, EPI_ISL_450874 | Evandro Chagas Institute | Evandro Chagas Institute | A.M.; Barbagelata; E.C.; E.M.A.; Ferreira; G.M.R; J.A.; Junior; L.C.; L.S.; M.C.; Martins; P.S.; Santos; Silva; Sousa; Sousa Junior; Viana; W.D.C.; da Silva |
| EPI_ISL_752605 | Faculty of Medicine, Al-Quds University | Faculty of Medicine, Al-Quds University | A. and Al-Jawabreh, A.; Ereqat, S.; Nasereddin |
| EPI_ISL_526940, EPI_ISL_614298, EPI_ISL_614304, EPI_ISL_614307, EPI_ISL_614308, EPI_ISL_614312 | Faroese National Reference Laboratory for Fish and Animal Diseases | Faroese National Reference Laboratory for Fish and Animal Diseases | Debes Hammershaimb Christiansen; Maria Marjunardóttir Dahl; Petra Elisabeth Petersen |
| EPI_ISL_549245 | Florida Bureau of Public Health Laboratories | Florida Bureau of Public Health Laboratories | Jason Blanton; Sarah Schmedes |
| EPI_ISL_476139 | Folkhalsomyndigheten | The Public Health Agency of Sweden | Anna Risberg; Anna-Malin Linde; Karin Tegmark-Wisell; Maria Lind Karlberg; Mattias Haukland; Olov Svartstrom; Oskar Karlsson Lindsjo; Petra Edquist; Reza Advani; Sandra Broddesson; Shamam Muradrasoli |
| EPI_ISL_581487, EPI_ISL_581490, EPI_ISL_581493 | Fondation Congolaise pour la recherche medicale (FCRM) | NGS Competence Center Tübingen, Institut für Medizinische Mikrobiologie und Hygiene, Universitätsklinikum Tübingen | Angel Angelov |
| EPI_ISL_411060, EPI_ISL_411066, EPI_ISL_431180, EPI_ISL_431782 | Fujian Center for Disease Control and Prevention | Fujian Center for Disease Control and Prevention | Chen Wei; He Wenxiang; Huang Zhimiao; Lin Qi; Weng Yuwei; Zhang Yanhua |
| EPI_ISL_707928 | Fujita Health University Hospital | Fujita Health University School of Medicine, Department of Microbiology | Aki Sakurai; Masahiro Suzuki; Yohei Doi |
| EPI_ISL_728154 | Fujita Health University Okazaki Medical Center | Department of Virology and Parasitology, Fujita Health University School of Medicine | Aki Sakurai; Masahiro suzuki; Satoshi Komoto; Takayuki Murata; Takuma Ishihara; Tomihiko Ide; Yohei Doi |
| EPI_ISL_794654 | Fundación Valle del Lili | Instituto Nacional de Salud - Dirección de Investigación en Salud Pública | Carlos Franco-Muñoz; Diego A. Álvarez-Díaz; Diego Andrés Prada; Gerardo Santamaría; Jonathan Reales; Julian Naizaque; Katherine Laiton-Donato; Magdalena Wiesner; Marcela Mercado-Reyes; Maria T. Herrera; Martha Lucia Ospina Martinez; Mauricio Pacheco-Montealegre; Paola Muñoz-Laiton; Sheryl Corchuelo |

| | | | |
|---|---|---|---|
| EPI_ISL_493367, EPI_ISL_500790, EPI_ISL_549074, EPI_ISL_590881, EPI_ISL_708129 | Furst Medical Laboratory | Norwegian Institute of Public Health, Department of Virology | Hilde Elshaug; Hilde Synnøve Vollan; Hilde Vollan; Kamilla Heddeland Instefjord; Karoline Bragstad; Kathrine Stene-Johansen; Marie Paulsen Madsen; Olav Hungnes; Rasmus Riis Kopperud |
| EPI_ISL_522549 | Félix Guyon Hospital | UMR PIMIT Université de La Réunion | Camille Lebarbenchon; David Wilkinson; Patrick Mavingui |
| EPI_ISL_524765 | GMERS Medical College and Hospital, Dharpur, Patan | Gujarat Biotechnology Research Centre | A M Kadri; A N Parmar; Afzal Ansari; Apurvasinh Puvar; Chaitanya Joshi; Dinesh Kumar; Harsh Bakshi; Janvi Raval; Komal Patel; Labdhi Pandya; Madhvi Joshi; Maharshi Pandya; Monika Gandhi; Nidhi Patel; Nikha Trivedi; Nitin Savaliya; Pinal Trivedi; R D Dixit; Raghawendra Kumar; Zarna Patel; Zuber Saiyed |
| EPI_ISL_467035 | GMERS Medical College and Hospital, Gandhinagar | Gujarat Biotechnology Research Centre | A M Kadri; Ankit Hinsu; Apurvasinh Puvar; Bharti Rajani; Bhavesh Modi; Chaitanya Joshi; Dinesh Kumar; Gaurishankar Shrimali; Harsh Bakshi; Janvi Raval; Komal Patel; Labdhi Pandya; Madhvi Joshi; Maharshi Pandya; Monika Gandhi; Neha Rajpara; Nidhi Patel; Nitin Savaliya; Pinal Trivedi; Pritesh Sabara; R D Dixit; Raghawendra Kumar; Seema Bhatt; Snehal Bagatharia; Tejas Shah; Zarna Patel; Zuber Saiyed |
| EPI_ISL_730567, EPI_ISL_730573, EPI_ISL_730575, EPI_ISL_730577 | Gazi University Faculty of Medicine, Medical Virology Laboratory | Gazi University Faculty of Medicine, Medical Virology Laboratory | Erdem Şahin; Gülendam Bozdayı; Hager Muftah; Işıl Fidan; Kayhan Çağlar; Murat Dizbay; Selin Yiğit; Shaknoza Sarzhanova; Özlem Güzel Tunçcan |
| EPI_ISL_677707, EPI_ISL_678252 | General Hospital - Kumanovo | Research Center for Genetic Engineering and Biotechnology "Georgi D. Efremov" , Macedonian Academy of Sciences and Arts | RCGEB - MASA |
| EPI_ISL_678256 | General Hospital - Ohrid | Research Center for Genetic Engineering and Biotechnology "Georgi D. Efremov" , Macedonian Academy of Sciences and Arts | RCGEB - MASA |
| EPI_ISL_514354, EPI_ISL_677674 | General Hospital - Prilep | Research Center for Genetic Engineering and Biotechnology "Georgi D. Efremov" , Macedonian Academy of Sciences and Arts | RCGEB - MASA |
| EPI_ISL_677719 | General Hospital - Struga | Research Center for Genetic Engineering and Biotechnology "Georgi D. Efremov" , Macedonian Academy of Sciences and Arts | RCGEB - MASA |
| EPI_ISL_677710 | General Hospital - Veles | Research Center for Genetic Engineering and Biotechnology "Georgi D. Efremov" , Macedonian Academy of Sciences and Arts | RCGEB - MASA |
| EPI_ISL_623097 | General practitioner | National Reference Center for Viruses of Respiratory Infections, Institut Pasteur, Paris | Angela Brisebarre; Camille Capel; Etienne Simon-Lorière; Marion Barbet; Maud Vanpeene; Méline Bizard; Sylvie Behillil; Sylvie van der Werf; Vincent Enouf |
| EPI_ISL_746486, EPI_ISL_746501, EPI_ISL_746531, EPI_ISL_746532, EPI_ISL_746537, EPI_ISL_746539, EPI_ISL_746550, EPI_ISL_746573, EPI_ISL_746595, EPI_ISL_746666, EPI_ISL_746692, EPI_ISL_746729, EPI_ISL_746734, EPI_ISL_746772, EPI_ISL_746785, EPI_ISL_746822 | | | |
| see above | Genetica Molecular and Subdepartamento de Virologia ISP Chile | Instituto de Salud Publica de Chile | Andres Castillo; Barbara Parra; Gisselle Barra; Jaime Lagos; Javier Tognarelli; Jorge Fernandez; Loredana Arata; Patricia Bustos; Rodrigo Fasce |
| EPI_ISL_605783 | Genome Center | Genome Center | A. S. M. Rubayet- Ul- Alam; Hassan M. Al-Emran; Md. Anwar Hossain; Md. Iqbal Kabir Jahid; Md. Shazid Hasan; Md. Tanvir Islam; Najmuj Sakib; Ovinu Kibria Islam; Pravas Chandra Roy; S. M. Tanjil Shah; Selina Akter; Shireen Nigar; Shovon Lal Sarkar |
| EPI_ISL_480230, EPI_ISL_480235, EPI_ISL_480269, EPI_ISL_735281, EPI_ISL_735288, EPI_ISL_735346, EPI_ISL_735360, EPI_ISL_735369 | | | |
| see above | Genomic Laboratory (GLAB) (Conjoint lab of Health Directorate of Istanbul and Istanbul Technical University) | Genomic Laboratory (GLAB), Istanbul Technical University | Arzu Irvem; Ayse Serra Ozel; Betsi Kose; Betsi Köse; Bugra Agaoglu; Elifnaz Çelik; Gizem Alkurt; Gizem Dinler Doganay; Ilker Karacan; Jale Yildiz; Levent Doganay; Mehtap Aydin; Mehtap Aydın; Nihat Bugra Agaoglu; Nilsun Altunal; Nisan Denizce Can; Ozlem Akgun Dogan; Payam Zolfagharian; Tugba Kizilboga Akgun; Yasemin Kendir Demirkol |
| EPI_ISL_803899 | Genomic Medicine Laboratory, IRCCS Santa Lucia Foundation | National Institute for Infectious Diseases, INMI, "L. Spallanzani" IRCCS | A Di Caro; B Bartolini; C.E.M Gruber; E Giardina; E Giombini; F Messina; M. Rueca; MR Capobianchi; O Butera |
| EPI_ISL_483688 | Genomic Research Lab, BCSIR | Genomic Research Lab, BCSIR | A. K. M. Shamsuzzaman; Abu Sayeed Mohammad Mahmud; Asish Kumar Ghosh; Barna Goswami; Eshrar Osman; Iffat Jahan; Mahmuda Yeasmin; Md. Ahasan Habib; Md. Maruf Ahmed Molla; Md. Murshed Hasan Sarkar; Md. Saddam Hossain; Md. Salim Khan; Mohammad Samir Uzzaman; Salek Ahmed Sajib; Shahina Akter; Sheikh Md. Selim Al Din; Tanjina Akhter Banu; Tasnim Nafisa; Utpal Chandra Ray |
| EPI_ISL_632908 | Genomic Sciences, Rehman Medical Institute | Genomic Sciences, Rehman Medical Institute | Afridi; Ali, J.; H. and Jehanzeb, V.; Haider; Jan; S.A.; Sabiha, B.; U.K. |
| EPI_ISL_730202, EPI_ISL_730209, EPI_ISL_730213, EPI_ISL_730219 | Genomica Lab Molecular, M√©xico | Andersen lab at Scripps Research | Jose Horacio Reyna Verdugo; Jose Roman Chavez Mendez; Luis Alberto Rangel Gonzalez; Martin Gonzalez Ibarra; SEARCH Alliance San Diego with Jonathan Gonzalez Garcia |
| EPI_ISL_774976 | Gonoshasthya-RNA Molecular Diagnostic and Research Center | Gonoshasthya-RNA Molecular Diagnostic and Research Center | Firoz Ahmed; Maha Jamiruddin; Mahfuza Marzan; Md. Ahsanul Haq; Mohd. Raeed Jamiruddin; Mohib Ullah Khondoker; Mousumi Chaity; Mumtarin Jannat Oishee; Nafisa Azmuda; Nihad Adnan; Nowshin Jahan; Salma Akter; Sayeda Moriam Liza; Shahad Saif Khandker; Shahana Sharmin; Tamanna Ali; Taslin Jahan Mou |
| EPI_ISL_775213, EPI_ISL_775216, EPI_ISL_775218 | Gonoshasthya-RNA Molecular Research Center | Gonoshasthya-RNA Molecular Research Center | Firoz Ahmed; Maha Jamiruddin; Mahfuza Marzan; Md. Ahsanul Haq; Mohd. Raeed Jamiruddin; Mohib Ullah Khondoker; Mousumi Chaity; Mumtarin Jannat Oishee; Nafisa Azmuda; Nihad Adnan; Nowshin Jahan; Salma Akter; Sayeda Moriam Liza; Shahad Saif Khandker; Shahana Sharmin; Tamanna Ali; Taslin Jahan Mou |
| EPI_ISL_415152 | Gorgas Memorial Institute for Health Studies | Gorgas Memorial Institute for Health Studies | Alexander A. Martinez.; Ambar Moreno; Claudia Gonzalez; Danilo Franco; Elimelec Valdespino; Juan M. Pascale; Leyda Abrego; Oris Chavarria; Sandra Lopez-Verges; Yamilka Diaz |
| EPI_ISL_640040 | Groote Schuur Hospital wc GSH | NHLS/UCT | Arash Iranzadeh; Bruna Galvao; Carolyn Williamson; Deelan Doolabh; Diana Hardie; Innocent Mudau; Kruger Marais; Lynn Tyers; Marvin Hsiao; Stephen Korsman |
| EPI_ISL_698223, EPI_ISL_698248, EPI_ISL_699038, EPI_ISL_699138 | Group 42 (G42) Healthcare, Abu Dhabi, United Arab Emirates; Department of Health, The United Arab Emirates | G42 Healthcare | Ashish Koshy; Budoor Alqarni; Denghui Liu; Fang Chen; Hanif Khalak; Huanming Yang; Javier Quilez; Jian Wang; Junhua Li; Ke Liang; Long Lin; Mohammed Saifuddin Fasihuddin; Nan Qiao; Nawal Ahmed Mohamed Al Kaabi; Pauline Ogrodzki; Pei Wu; Peng Xiao; Pengjuan Liu; Rong Liu; Sally Mahmoud; Siyang Liu; Stephen S. Francis; Tao Ma; Vinay Kusuma; Walid Abbas Zaher; Weibin Liu; Wenjun He; Xavier Anton; Xin Jin; Xin Meng; Xinyu Huang; Xun Xu; Zhaorong Yuan |
| EPI_ISL_403932, EPI_ISL_403936 | Guangdong Provincial Center for Diseases Control and Prevention; Guangdong Provincial Public Health | Department of Microbiology, Guangdong Provincial Center for Diseases Control and Prevention | Baisheng Li; Changwen Ke; Feng Ruan; Guanhao He; Haojie Zhong; Huihong Deng; Jianfeng He; Jianpeng Xiao; Jianxiang Geng; Jianxiong Hu; Jie Wu; Jing Lu; Lifeng Lin; Lijun Liang; Lirong Zou; Min Kang; Qi Zhu; Shujiang Mei; Songjian Xiao; Tao Liu; Tie Song; Weilin Zeng; Wenjun Ma; Xing Li; Xiujuan Tang; Xue Zhuang; Xuguang Chen; Ying Wang; Yingchao Song; Yingtao Zhang; Yuhuang Liao; Zhe Liu |
| EPI_ISL_406534 | Guangdong Provincial Center for Diseases Control and Prevention; Guangdong Provincial Public Health | Guangdong Provincial Center for Diseases Control and Prevention | Baisheng Li; Changwen Ke; Feng Ruan; Guanhao He; Haojie Zhong; Huihong Deng; Jianfeng He; Jianpeng Xiao; Jianxiang Geng; Jianxiong Hu; Jie Wu; Jing Lu; Lifeng Lin; Lijun Liang; Lirong Zou; Min Kang; Qi Zhu; Shujiang Mei; Songjian Xiao; Tao Liu; Tie Song; Weilin Zeng; Wenjun Ma; Xing Li; Xiujuan Tang; Xue Zhuang; Xuguang Chen; Ying Wang; Yingchao Song; Yingtao Zhang; Yuhuang Liao; Zhe Liu |
| EPI_ISL_428457 | Guangdong Provincial Center for Diseases Control and Prevention;Guangdong Provincial Institute of Public Health | School of Public Health, The University of Hong Kong | Bosheng Li; Hanri Zeng; Haogao Gu; Hui-Ling Yen; Jie Wu; Leo L.M. Poon; Lijun Liang; Tie Song; Yao Hu; Yingchao Song; Zhencui Li |
| EPI_ISL_413853, EPI_ISL_413855 | Guangdong Provincial Institution of Public Health, Guangdong Provinical Center for Disease Control and Prevention | Guangdong Provincial Institution of Public Health | Andrew Rambaut; Bo Peng; Changwen Ke; Chuming Liang; Huanying Zheng; Huifang Lin; Jing Lu; Jingju Peng; Jiufeng Sun; Josh Quick; Juan Su; Kang Min; Kuibiao Li; Lilian Zeng; Liu Zhe; Louis du Plessis; Minfeng Liang; Moritz Kraemer; Nick Loman; Nuno Faria; Oliver Pybus; Pingping Zhou; Qianlin Xiong; Ru bai; Ruilin Sun; Runyu Yuan; Sarah François; Shisong Fang; Song Tie; Tao Liu; Verity Hill; Wei Li; Wenjun Ma; Wenzhe Su; Xi Tang |
| EPI_ISL_444969 | Guangzhou Eighth People's Hospital (Jiahe Sector) | Institute of Human Virology, Zhongshan School of Medicine, Sun Yat-sen University | Bingfeng Liu; Fang Li; Fei Yu; Feng Huang; Fengyu Hu; Hui Zhang; Huimin Fan; Jun Liu; Junsong Zhang; Kai Deng; Mang Shi; Ruosu Ying; Ting Pan; Xu Zhang; Yiwen Zhang |
| EPI_ISL_509695, EPI_ISL_509701 | Guatemala Ministry of Public Health | Pathogen Discovery, Respiratory Viruses Branch, Division of Viral Diseases, Centers for Disease Control and Prevention | Anna Uehara; Clinton Paden; Haibin Wang; Jing Zhang; Krista Queen; Suxiang Tong; Yan Li; Ying Tao |
| EPI_ISL_700576 | Guguletu CHC wc GDH | NHLS/UCT | Arash Iranzadeh; Bruna Galvao; Carolyn Williamson; Deelan Doolabh; Diana Hardie; Innocent Mudau; Kruger Marais; Lynn Tyers; Marvin Hsiao; Stephen Korsman |
| EPI_ISL_471151, EPI_ISL_660809, EPI_ISL_661099 | Gundersen Molecular Diagnostics Laboratory | Kabara Cancer Research Institute | Craig S. Richmond; Paraic A. Kenny |
| EPI_ISL_745201 | Guntur General Hospital | CSIR-Centre for Cellular and Molecular Biology | Archana Bharadwaj Siva; B Himasri; Blessy B John; Divya Tej Sowpati; Dr.P.Shashikala Reddy; Dr.S.Pavani; Dr.Satyaprasad; Dr.V.Sudha Rani; Karthik Bharadwaj Tallapaka; Lamuk Zaveri; M Soujanya Reddy; Namami Gaur; Nikhil Hajirnis; Onkar Kulkarni; Payel Mukherjee; Pratheusa Maccha; Priya Singh; Purushotham Vodnala; Rakesh K Mishra; Sakshi Shambhavi; Shagufta Khan; Sofia Banu; Tulasi Nagabandi; Viswagithe S L |
| EPI_ISL_511361, EPI_ISL_693560, EPI_ISL_693600, EPI_ISL_693615 | H Beatriz Angelo | Instituto Nacional de Saude (INSA) | Borges et al |
| EPI_ISL_418022 | H Braga | Instituto Nacional de Saude (INSA) | Guiomar et al |
| EPI_ISL_454190 | H Dr Nelio Mendonca - Funchal | Instituto Nacional de Saude (INSA) | Borges et al |
| EPI_ISL_454293 | H Evora | Instituto Nacional de Saude (INSA) | Borges et al |
| EPI_ISL_732241 | H Evora | Instituto Nacional de Saude (INSA) and Instituto Gulbenkian de Ciencia (IGC) | Borges et al |
| EPI_ISL_453828, EPI_ISL_454201, EPI_ISL_454220, EPI_ISL_693634, EPI_ISL_693637 | H Vila Franca Xira | Instituto Nacional de Saude (INSA) | Borges et al |
| EPI_ISL_596255, EPI_ISL_596282 | HELIX LCC | WHO National Influenza Centre Russian Federation | Andrey Komissarov; Anna Ivanova; Artem Fadeev; Daria Danilenko; Dmitry Bazhenov; Kseniya Komissarova |
| EPI_ISL_602380, EPI_ISL_733062, | HELIX LLC | WHO National Influenza Centre Russian Federation | Andrey Komissarov; Anna Ivanova; Artem Fadeev; Daria Danilenko; Dmitry Bazhenov; Dmitry Lioznov; Elena Nabieva; Georgii Bazykin; Ksenia Safina; Kseniya Komissarova |

| | | | |
|---|---|---|---|
| EPI_ISL_733077, EPI_ISL_733089, EPI_ISL_733151, EPI_ISL_733436 | | | |
| EPI_ISL_445348 | HOSPITAL DR.HERNAN HENRIQUEZ ARAVENA | Instituto de Salud Publica de Chile | Alejandra Acevedo; Andrés E Castillo; Bárbara Parra; Carolina Tambley; Gabriel Leal; Jaime Lagos; Jorge Fernandez; Loredana Arata; Patricia Bustos; Paz Tapia; Rodrigo Fasce; Winston Andrade |
| EPI_ISL_682270 | HOSPITAL SAN JUAN DE DIOS | Inciensa, Instituto Costarricense de Investigación y Enseñanza en Nutrición y Salud | Adriana Godinez & Melany Calderon; Claudio Soto-Garita; Estela Cordero; Francisco Duarte; Hebleen Porras |
| EPI_ISL_682255 | HOSPITAL SAN VICENTE DE PAUL | Inciensa, Instituto Costarricense de Investigación y Enseñanza en Nutrición y Salud | Adriana Godinez & Melany Calderon; Claudio Soto-Garita; Estela Cordero; Francisco Duarte; Hebleen Porras |
| EPI_ISL_636977 | HP Pemba | KRISP, KZN Research Innovation and Sequencing Platform | Giandhari J; Ismael N; Nadia Sitoe; Nedio Mabunda; Paulo Arnaldo; Pillay S; Tegally H; Wilkinson E; de Oliveira T |
| EPI_ISL_731918 | HSE Ilha Terceira - Angra do Heroismo | Instituto Nacional de Saude (INSA) | Borges et al |
| EPI_ISL_475157, EPI_ISL_582805, EPI_ISL_615107 | Halmstad klinisk mikrobiologi | The Public Health Agency of Sweden | Anna Risberg; Anna-Malin Linde; Karin Tegmark-Wisell; Maria Lind Karlberg; Mattias Haukland; Mia Brytting; Olov Svartstrom; Oskar Karlsson Lindsjo; Petra Edquist; Reza Advani; Sandra Broddesson; Shamam Muradrasoli |
| EPI_ISL_660139 | Hamadi | National Health Laboratory Service (NHLS), Tygerberg | Bronwyn Kleinhans; Davis M-A; Draper C; Eduan Wilkindon; Gert van Zyl; Houriiyah Tegally; Hsiao M; Kayla Delaney; Siegfried N; Susan Engelbrecht; Tulio de Oliveira; Williamson C; Wolfgang Preiser |
| EPI_ISL_418514 | Hangzhou Center for Disease Control and Prevention | Inspection Center of Hangzhou Center for Disease Control and Prevention | Li jun; Pan jingcao; Wang haoqiu; Yu hua; Yu xinfeng |
| EPI_ISL_421236, EPI_ISL_482577, EPI_ISL_482578, EPI_ISL_482584, EPI_ISL_482585 | Hangzhou Center for Diseases Control and Prevention | Hangzhou Center for Diseases Control and Prevention | Haoqiu Wang; Hua Yu; Jun Li; Junfang Chen; Lingfeng Mao; Shuchang Chen; Xin Qian; Xinfen Yu; Xuchu Wang; Zhou Sun |
| EPI_ISL_700515 | Hanover Park CHC wc HPH | NHLS/UCT | Arash Iranzadeh; Bruna Galvao; Carolyn Williamson; Deelan Doolabh; Diana Hardie; Innocent Mudau; Kruger Marais; Lynn Tyers; Marvin Hsiao; Stephen Korsman |
| EPI_ISL_796015 | Hebei Provincial Center for Disease Control and Prevention, Shijiazhuang, Hebei Province; National Institute for Viral Disease Control and Prevention, China CDC | Hebei Provincial Center for Disease Control and Prevention, Shijiazhuang, Hebei Province; National Institute for Viral Disease Control and Prevention, China CDC | George F. Gao; Nankun Liu; Qi Li; Shunxiang Qi; Wenbo Xu; Xiang Zhao; Yang Song |
| EPI_ISL_700549 | Heideveld CDC wc HVP | NHLS/UCT | Arash Iranzadeh; Bruna Galvao; Carolyn Williamson; Deelan Doolabh; Diana Hardie; Innocent Mudau; Kruger Marais; Lynn Tyers; Marvin Hsiao; Stephen Korsman |
| EPI_ISL_700545 | Heideveld Emergency Centre | NHLS/UCT | Arash Iranzadeh; Bruna Galvao; Carolyn Williamson; Deelan Doolabh; Diana Hardie; Innocent Mudau; Kruger Marais; Lynn Tyers; Marvin Hsiao; Stephen Korsman |
| EPI_ISL_501233, EPI_ISL_501236, EPI_ISL_501248, EPI_ISL_501250 | Hellenic Pasteur Institute, National Influenza Reference laboratory of Southern Greece & Unit of Bioinformatics and Applied Genomics | Hellenic Pasteur Institute, National Influenza Reference laboratory of Southern Greece & Unit of Bioinformatics and Applied Genomics | Andreas Mentis; Androniki Voulgari-Kokota; Antonios Kalliaropoulos; Aspasia Kontou; Athanasios Kossyvakis; Evangelidou Maria; Horefti Elina; Timokratis Karamitros; Vasiliki Pogka |
| EPI_ISL_430469 | Hellenic Pasteur Institute, Public Health Laboratories | Hellenic Pasteur Institute, Public Health Laboratories, Unit of Bioinformatics and Applied Genomics | Andreas Mentis; Androniki Voulgari-Kokota; Antonios Kalliaropoulos; Aspasia Kontou; Athanasios Kossyvakis; Evangelidou Maria; Horefti Elina; Timokratis Karamitros; Vasiliki Pogka |
| EPI_ISL_512298 | Hematology Laboratory, Section of Molecular Diagnostics, University Clinical Centre, Medical University of Gdansk | Department of Virology, Faculty of Medicine, University of Helsinki, Helsinki, Finland | Aneta Szulc; Ewa Miłosz; Maciej Grzybek; Marlena Robakowska; Olli Vapalahti; Teemu Smura |
| EPI_ISL_699749, EPI_ISL_699792, EPI_ISL_699850, EPI_ISL_699874, EPI_ISL_700006, EPI_ISL_700070, EPI_ISL_700175, EPI_ISL_700216, EPI_ISL_700239, EPI_ISL_700270, EPI_ISL_700302, EPI_ISL_700322 | | | |
| see above | Hematopathology Laboratory, ACTREC, TMC | Hematopathology Laboratory, ACTREC, TMC | ACTREC; Hematopathology Laboratory |
| EPI_ISL_767873 | Histopath | NSW Health Pathology - Institute of Clinical Pathology and Medical Research; Westmead Hospital; University of Sydney | CIDM-PH et al. |
| EPI_ISL_770013 | Hle - Asilos De Ancianos | Inciensa, Instituto Costarricense de Investigación y Enseñanza en Nutrición y Salud | Adriana Godínez; Claudio Soto-Garita; Estela Cordero; Francisco Duarte; Hebleen Porras; Melany Calderón & Mariel López |
| EPI_ISL_770006 | Hle - Asociacion Hogar De Ancianos De Palmar Sur De Osa | Inciensa, Instituto Costarricense de Investigación y Enseñanza en Nutrición y Salud | Adriana Godínez; Claudio Soto-Garita; Estela Cordero; Francisco Duarte; Hebleen Porras; Melany Calderón & Mariel López |
| EPI_ISL_770031 | Hle-Asociacion De Atencion Integral Del Anciano San Cayetano | Inciensa, Instituto Costarricense de Investigación y Enseñanza en Nutrición y Salud | Adriana Godínez; Claudio Soto-Garita; Estela Cordero; Francisco Duarte; Hebleen Porras; Melany Calderón & Mariel López |
| EPI_ISL_414569, EPI_ISL_760036, EPI_ISL_760037, EPI_ISL_760041, EPI_ISL_760058 | Hong Kong Department of Health | School of Public Health, The University of Hong Kong | Carrie Wan; Daisy Ng; Daniel Chu; Daniel K.W. Chu; Dominic N.C. Tsang; Gigi Liu; Haogao Gu; Leo L.M. Poon; Leo Poon; Malik Peiris; Pavithra Krishnan |
| EPI_ISL_471562, EPI_ISL_471581, EPI_ISL_471582, EPI_ISL_515564, EPI_ISL_523977 | Hosp. Municipal Prof. Dr. Alípio Corrêa Netto | Instituto Adolfo Lutz, Interdiciplinary Procedures Center, Strategic Laboratory | Claudia Regina Gonçalves; Claudio Tavares Sacchi; Erica Valessa Ramos Gomes |
| EPI_ISL_560643 | Hospital | National Reference Center for Viruses of Respiratory Infections, Institut Pasteur, Paris | Etienne Simon-Lorière; Fabiana Gambaro; Maud Vanpeene; Sylvie Behillil; Sylvie van der Werf; Vincent Enouf |
| EPI_ISL_693229 | Hospital 8 de Maio | Instituto Adolfo Lutz, Interdiciplinary Procedures Center, Strategic Laboratory | Claudia Regina Gonçalves; Claudio Tavares Sacchi; Erica Valessa Ramos Gomes; Karoline Rodrigues Campos |
| EPI_ISL_471554 | Hospital Bosque da Saúde | Instituto Adolfo Lutz, Interdiciplinary Procedures Center, Strategic Laboratory | Claudia Regina Gonçalves; Claudio Tavares Sacchi; Erica Valessa Ramos Gomes |
| EPI_ISL_574580 | Hospital Cidade Tiradentes Carmen Prudente | Instituto Adolfo Lutz, Interdiciplinary Procedures Center, Strategic Laboratory | Claudia Regina Gonçalves; Claudio Tavares Sacchi; Erica Valessa Ramos Gomes; Karoline Rodrigues Campos |
| EPI_ISL_539536, EPI_ISL_539538 | Hospital Clínic | Instituto de Salud Carlos III | A. Monzón; F. Casas; I; I. Jiménez; Iglesias-Caballero; M. Camarero; M. Cuesta; M. González-Esguevillas; M. Molinero Calamita; M. Zaballos; M.A Marcos; P. Jiménez; S. Juliá; S. Pozo; S. Varona |
| EPI_ISL_691700 | Hospital Clínico San Carlos | Instituto de Salud Carlos III | A. Monzón; F. Casas; I. Jiménez; I. Rodríguez, I.; Iglesias-Caballero; M. Camarero; M. Cuesta; M. González-Esguevillas; M. Pozo; M. Zaballos; P. Jiménez; S. Juliá; S. Molinero Calamita; S. Varona |
| EPI_ISL_574591, EPI_ISL_574592, EPI_ISL_603030, EPI_ISL_693218 | Hospital Domingos Leonardo Ceravolo Presidente Prudente | Instituto Adolfo Lutz, Interdiciplinary Procedures Center, Strategic Laboratory | Claudia Regina Gonçalves; Claudio Tavares Sacchi; Erica Valessa Ramos Gomes; Karoline Rodrigues Campos |
| EPI_ISL_547578 | Hospital Doutor Domingos Leonardo Cerávolo | Instituto Adolfo Lutz, Interdiciplinary Procedures Center, Strategic Laboratory | Claudia Regina Gonçalves; Claudio Tavares Sacchi; Erica Valessa Ramos Gomes; Karoline Rodrigues Campos |
| EPI_ISL_527752 | Hospital Dr. Rafael A. Calderon Guardia | Inciensa, Instituto Costarricense de Investigación y Enseñanza en Nutrición y Salud | Adriana Godinez & Melany Calderon; Claudio Soto-Garita; Estela Cordero; Francisco Duarte; Hebleen Porras |
| EPI_ISL_512654 | Hospital Dr. Rafael A. Calderon Guardia [San Jose/San Jose] | Inciensa, Instituto Costarricense de Investigación y Enseñanza en Nutrición y Salud | Adriana Godinez & Melany Calderon; Claudio Soto-Garita; Estela Cordero; Francisco Duarte; Hebleen Porras |
| EPI_ISL_693213 | Hospital E Maternidade Municipal Governador Mario Covas | Instituto Adolfo Lutz, Interdiciplinary Procedures Center, Strategic Laboratory | Claudia Regina Gonçalves; Claudio Tavares Sacchi; Erica Valessa Ramos Gomes; Karoline Rodrigues Campos |
| EPI_ISL_574594 | Hospital Escola da Universidade de Taubate | Instituto Adolfo Lutz, Interdiciplinary Procedures Center, Strategic Laboratory | Claudia Regina Gonçalves; Claudio Tavares Sacchi; Erica Valessa Ramos Gomes; Karoline Rodrigues Campos |
| EPI_ISL_574588, EPI_ISL_583490 | Hospital Estadual Sumare | Instituto Adolfo Lutz, Interdiciplinary Procedures Center, Strategic Laboratory | Claudia Regina Gonçalves; Claudio Tavares Sacchi; Erica Valessa Ramos Gomes; Karoline Rodrigues Campos |
| EPI_ISL_583501 | Hospital Estadual de CampanhaCOVID 19 Barradas | Instituto Adolfo Lutz, Interdiciplinary Procedures Center, Strategic Laboratory | Claudia Regina Gonçalves; Claudio Tavares Sacchi; Erica Valessa Ramos Gomes; Karoline Rodrigues Campos |
| EPI_ISL_541894, EPI_ISL_541915, EPI_ISL_654116, EPI_ISL_654212, EPI_ISL_654317, EPI_ISL_654338, EPI_ISL_780002, EPI_ISL_780092 | | | |
| see above | Hospital General Universitario Gregorio Marañón | SeqCOVID-SPAIN consortium/IBV(CSIC) | Darío García de Viedma; Darío García de Viedma and SeqCOVID-SPAIN consortium; Jon Sicilia; Julia Suárez; Laura Pérez-Lago; Marta Herranz; Patricia Muñoz; Patricia Muñoz and SeqCOVID-SPAIN consortium; Pilar Catalán |
| EPI_ISL_471541, EPI_ISL_523971 | Hospital Geral Santa Marcelina | Instituto Adolfo Lutz, Interdiciplinary Procedures Center, Strategic Laboratory | Claudia Regina Gonçalves; Claudio Tavares Sacchi; Erica Valessa Ramos Gomes |
| EPI_ISL_534317 | Hospital Geral de Itapevi | Instituto Adolfo Lutz, Interdiciplinary Procedures Center, Strategic Laboratory | Claudia Regina Gonçalves; Claudio Tavares Sacchi; Erica Valessa Ramos Gomes |
| EPI_ISL_603037 | Hospital Geral de Pedreira | Instituto Adolfo Lutz, Interdiciplinary Procedures Center, Strategic Laboratory | Claudia Regina Gonçalves; Claudio Tavares Sacchi; Erica Valessa Ramos Gomes; Karoline Rodrigues Campos |
| EPI_ISL_515555 | Hospital Geral de Vila Nova Cachoeirinha | Instituto Adolfo Lutz, Interdiciplinary Procedures Center, Strategic Laboratory | Claudia Regina Gonçalves; Claudio Tavares Sacchi; Erica Valessa Ramos Gomes |
| EPI_ISL_574595 | Hospital Geral de Vila Penteado Dr. Jose Pamgella | Instituto Adolfo Lutz, Interdiciplinary Procedures Center, Strategic Laboratory | Claudia Regina Gonçalves; Claudio Tavares Sacchi; Erica Valessa Ramos Gomes; Karoline Rodrigues Campos |
| EPI_ISL_481248, EPI_ISL_574431 | Hospital IESS Babahoyo | Institute of Microbiology, Universidad San Francisco de Quito | Belén Prado-Vivar; Bernardo Gutiérrez; Carla Torres; Fernanda Zurita; Francisco Cordova; Gabriel Trueba; Juan José Guadalupe; Killen Briones-Claudette; Killen Briones-Zamora; Michelle Grunauer; Monica Becerra-Wong; Ninfa Henriquez; Patricio Rojas-Silva; Paúl Cárdenas; Sully Márquez; Verónica Barragán |
| EPI_ISL_412964 | Hospital Israelita Albert Einstein | Instituto Adolfo Lutz Interdisciplinary Procedures Center Strategic Laboratory | Andrew Rambaut; Claudia Regina Gonçalves; Claudio Tavares Sacchi; Daniela Bernardes Borges da Silva; Ester Cerdeira Sabino; Flávia Cristina da Silva Sales; Ingra Morales Claro; Jaqueline Goes de Jesus; Joshua Quick; Maria do Carmo; Nicholas James Loman; Nuno Rodrigues Faria; Sampaio Tavares Timenetsky |
| EPI_ISL_414014, EPI_ISL_416033, EPI_ISL_416034 | Hospital Israelita Albert Einstein | Instituto Adolfo Lutz, Interdiciplinary Procedures Center, Strategic Laboratory | Adriana Bugno; Adriano Abbud; Carlos Henrique Camargo; Claudia Regina Gonçalves; Claudio Tavares Sacchi; Daniela Bernardes Borges da Silva; Erica Valessa Ramos Gomes; Ester Cerdeira Sabino; Fabiana Cristina Pereira dos Santos; Katia Correia dos Santos; Maria do Carmo Sampaio Tavares Timenetsky; Maria do Carmo Sampaito Tavares Timenetsky; Simone Guadagnucci Morillo; Terezinha Maria de Paiva |
| EPI_ISL_413016 | Hospital Israelita Albert Einstein | Instituto Adolfo Lutz, Interdisciplinary Procedures Center, Strategic Laboratory | Andrew Rambaut; Claudia Regina Gonçalves; Claudio Tavares Sacchi; Ester Cerdeira Sabino; Fabiana Cristina Pereira dos Santos; Flávia Cristina da Silva Sales; Ingra Morales Claro; Jaqueline Goes de Jesus; Joshua Quick; Maria do Carmo Sampaio Tavares Timenetsky; Nicholas James Loman; Nuno Rodrigues Faria |
| EPI_ISL_523957 | Hospital Itamaraty | Instituto Adolfo Lutz, Interdiciplinary Procedures Center, Strategic Laboratory | Claudia Regina Gonçalves; Claudio Tavares Sacchi; Erica Valessa Ramos Gomes |

| | | | |
|---|---|---|---|
| EPI_ISL_769992 | Hospital Metropolitano | Inciensa, Instituto Costarricense de Investigación y Enseñanza en Nutrición y Salud | Adriana Godínez; Claudio Soto-Garita; Estela Cordero; Francisco Duarte; Hebleen Porras; Melany Calderón & Margarita Lee Lui |
| EPI_ISL_524462 | Hospital Metropolitano | Instituto Adolfo Lutz, Interdiciplinary Procedures Center, Strategic Laboratory | Claudia Regina Gonçalves; Claudio Tavares Sacchi; Erica Valessa Ramos Gomes |
| EPI_ISL_527739 | Hospital Mexico [San Jose/San Jose] | Inciensa, Instituto Costarricense de Investigación y Enseñanza en Nutrición y Salud | Adriana Godinez & Melany Calderon; Claudio Soto-Garita; Estela Cordero; Francisco Duarte; Hebleen Porras |
| EPI_ISL_515541, EPI_ISL_515561 | Hospital Montemagno | Instituto Adolfo Lutz, Interdiciplinary Procedures Center, Strategic Laboratory | Claudia Regina Gonçalves; Claudio Tavares Sacchi; Erica Valessa Ramos Gomes |
| EPI_ISL_534324 | Hospital Mun Ver Jose Storopolli | Instituto Adolfo Lutz, Interdiciplinary Procedures Center, Strategic Laboratory | Claudia Regina Gonçalves; Claudio Tavares Sacchi; Erica Valessa Ramos Gomes |
| EPI_ISL_534318, EPI_ISL_693208, EPI_ISL_693209 | Hospital Municipal Antonio Giglio | Instituto Adolfo Lutz, Interdiciplinary Procedures Center, Strategic Laboratory | Claudia Regina Gonçalves; Claudio Tavares Sacchi; Erica Valessa Ramos Gomes; Karoline Rodrigues Campos |
| EPI_ISL_547571 | Hospital Municipal Antônio Giglio | Instituto Adolfo Lutz, Interdiciplinary Procedures Center, Strategic Laboratory | Claudia Regina Gonçalves; Claudio Tavares Sacchi; Erica Valessa Ramos Gomes; Karoline Rodrigues Campos |
| EPI_ISL_471549, EPI_ISL_523991, EPI_ISL_523992, EPI_ISL_527869 | Hospital Municipal Carmen Prudente | Instituto Adolfo Lutz, Interdiciplinary Procedures Center, Strategic Laboratory | Claudia Regina Gonçalves; Claudio Tavares Sacchi; Erica Valessa Ramos Gomes |
| EPI_ISL_515562 | Hospital Municipal Doutor Alexandre Zaio | Instituto Adolfo Lutz, Interdiciplinary Procedures Center, Strategic Laboratory | Claudia Regina Gonçalves; Claudio Tavares Sacchi; Erica Valessa Ramos Gomes |
| EPI_ISL_693203 | Hospital Municipal Doutor Arthur Ribeiro de Saboya | Instituto Adolfo Lutz, Interdiciplinary Procedures Center, Strategic Laboratory | Claudia Regina Gonçalves; Claudio Tavares Sacchi; Erica Valessa Ramos Gomes; Karoline Rodrigues Campos |
| EPI_ISL_468311, EPI_ISL_468312 | Hospital Municipal Dr Ignacio Proenca de Gouvea | Instituto Adolfo Lutz, Interdiciplinary Procedures Center, Strategic Laboratory | Claudia Regina Gonçalves; Claudio Tavares Sacchi; Erica Valessa Ramos Gomes |
| EPI_ISL_515521 | Hospital Municipal Dr Waldemar Tebaldi | Instituto Adolfo Lutz, Interdiciplinary Procedures Center, Strategic Laboratory | Claudia Regina Gonçalves; Claudio Tavares Sacchi; Erica Valessa Ramos Gomes |
| EPI_ISL_523985 | Hospital Municipal Dr. Benedicto Montenegro | Instituto Adolfo Lutz, Interdiciplinary Procedures Center, Strategic Laboratory | Claudia Regina Gonçalves; Claudio Tavares Sacchi; Erica Valessa Ramos Gomes |
| EPI_ISL_515553, EPI_ISL_574577, EPI_ISL_574579 | Hospital Municipal Dr. Ignacio Proença de Gouvea | Instituto Adolfo Lutz, Interdiciplinary Procedures Center, Strategic Laboratory | Claudia Regina Gonçalves; Claudio Tavares Sacchi; Erica Valessa Ramos Gomes; Karoline Rodrigues Campos |
| EPI_ISL_515548, EPI_ISL_515563, EPI_ISL_574582, EPI_ISL_574589 | Hospital Municipal Dr. Jose Soares Hungria | Instituto Adolfo Lutz, Interdiciplinary Procedures Center, Strategic Laboratory | Claudia Regina Gonçalves; Claudio Tavares Sacchi; Erica Valessa Ramos Gomes; Karoline Rodrigues Campos |
| EPI_ISL_515557, EPI_ISL_524467 | Hospital Municipal Dr. Moysés Deutsch | Instituto Adolfo Lutz, Interdiciplinary Procedures Center, Strategic Laboratory | Claudia Regina Gonçalves; Claudio Tavares Sacchi; Erica Valessa Ramos Gomes |
| EPI_ISL_583498 | Hospital Municipal Dr. Waldemar Tebaldi | Instituto Adolfo Lutz, Interdiciplinary Procedures Center, Strategic Laboratory | Claudia Regina Gonçalves; Claudio Tavares Sacchi; Erica Valessa Ramos Gomes; Karoline Rodrigues Campos |
| EPI_ISL_693206 | Hospital Municipal Mario Gatti | Instituto Adolfo Lutz, Interdiciplinary Procedures Center, Strategic Laboratory | Claudia Regina Gonçalves; Claudio Tavares Sacchi; Erica Valessa Ramos Gomes; Karoline Rodrigues Campos |
| EPI_ISL_527870, EPI_ISL_574578, EPI_ISL_603029, EPI_ISL_603039 | Hospital Municipal Mário Gatti | Instituto Adolfo Lutz, Interdiciplinary Procedures Center, Strategic Laboratory | Claudia Regina Gonçalves; Claudio Tavares Sacchi; Erica Valessa Ramos Gomes; Karoline Rodrigues Campos |
| EPI_ISL_527856 | Hospital Municipal Prof. Waldomiro de Paula | Instituto Adolfo Lutz, Interdiciplinary Procedures Center, Strategic Laboratory | Claudia Regina Gonçalves; Claudio Tavares Sacchi; Erica Valessa Ramos Gomes |
| EPI_ISL_603028 | Hospital Municipal Santa Ana | Instituto Adolfo Lutz, Interdiciplinary Procedures Center, Strategic Laboratory | Claudia Regina Gonçalves; Claudio Tavares Sacchi; Erica Valessa Ramos Gomes; Karoline Rodrigues Campos |
| EPI_ISL_524468, EPI_ISL_527859 | Hospital Municipal Vereador Jose Storopolli | Instituto Adolfo Lutz, Interdiciplinary Procedures Center, Strategic Laboratory | Claudia Regina Gonçalves; Claudio Tavares Sacchi; Erica Valessa Ramos Gomes |
| EPI_ISL_471647 | Hospital Municipal de Barueri Dr. Francisco Moran | Instituto Adolfo Lutz, Interdiciplinary Procedures Center, Strategic Laboratory | Claudia Regina Gonçalves; Claudio Tavares Sacchi; Erica Valessa Ramos Gomes |
| EPI_ISL_527860 | Hospital Municipal de Parelheiros Josanias Castanha Braga | Instituto Adolfo Lutz, Interdiciplinary Procedures Center, Strategic Laboratory | Claudia Regina Gonçalves; Claudio Tavares Sacchi; Erica Valessa Ramos Gomes |
| EPI_ISL_527862 | Hospital Municipal de Urgência | Instituto Adolfo Lutz, Interdiciplinary Procedures Center, Strategic Laboratory | Claudia Regina Gonçalves; Claudio Tavares Sacchi; Erica Valessa Ramos Gomes |
| EPI_ISL_468308, EPI_ISL_468315, EPI_ISL_515520, EPI_ISL_515526, EPI_ISL_515546, EPI_ISL_515551, EPI_ISL_515552, EPI_ISL_523955, EPI_ISL_523974, EPI_ISL_523976, EPI_ISL_527863 | | | |
| see above | Hospital Municipal do Tatuape Carmino Caricchio | Instituto Adolfo Lutz, Interdiciplinary Procedures Center, Strategic Laboratory | Claudia Regina Gonçalves; Claudio Tavares Sacchi; Erica Valessa Ramos Gomes |
| EPI_ISL_735399 | Hospital Municiplal Dr Ignacio de gouvea | Instituto Adolfo Lutz, Interdiciplinary Procedures Center, Strategic Laboratory | Claudia Regina Gonçalves; Claudio Tavares Sacchi; Erica Valessa Ramos Gomes; Karoline Rodrigues Campos |
| EPI_ISL_491451 | Hospital México | Inciensa, Instituto Costarricense de Investigación y Enseñanza en Nutrición y Salud | Adriana Godinez & Melany Calderon; Claudio Soto-Garita; Estela Cordero; Francisco Duarte; Hebleen Brenes |
| EPI_ISL_735428, EPI_ISL_735429, EPI_ISL_735431, EPI_ISL_735432 | Hospital Nipo Brasileiro | Instituto Adolfo Lutz, Interdiciplinary Procedures Center, Strategic Laboratory | Claudia Regina Gonçalves; Claudio Tavares Sacchi; Erica Valessa Ramos Gomes; Karoline Rodrigues Campos |
| EPI_ISL_539496 | Hospital Nostra Senyora de Meritxell | Instituto de Salud Carlos III | A. Monzón; F. Casas; F. Fernández; I; I. Jiménez; Iglesias-Caballero; M. Camarero; M. Cuesta; M. González-Esguevillas; M. Molinero Calamita; M. Zaballos; P. Jiménez; S. Juliá; S. Pozo; S. Varona |
| EPI_ISL_471269, EPI_ISL_471270 | Hospital Oncológico Solca Núcleo de Quito | Institute of Microbiology, Universidad San Francisco de Quito | Belén Prado-Vivar; Bernardo Gutiérrez; Gabriel Trueba; Grace Salazar; Juan José Guadalupe; Marcos Di Stefano; Michelle Grunauer; Patricio Rojas-Silva; Paúl Cárdenas; Sully Márquez; Verónica Barragán |
| EPI_ISL_527857 | Hospital Regional Vale do Ribeira | Instituto Adolfo Lutz, Interdiciplinary Procedures Center, Strategic Laboratory | Claudia Regina Gonçalves; Claudio Tavares Sacchi; Erica Valessa Ramos Gomes |
| EPI_ISL_523956 | Hospital Regional de Assis | Instituto Adolfo Lutz, Interdiciplinary Procedures Center, Strategic Laboratory | Claudia Regina Gonçalves; Claudio Tavares Sacchi; Erica Valessa Ramos Gomes |
| EPI_ISL_524463 | Hospital Regional de Cotia | Instituto Adolfo Lutz, Interdiciplinary Procedures Center, Strategic Laboratory | Claudia Regina Gonçalves; Claudio Tavares Sacchi; Erica Valessa Ramos Gomes |
| EPI_ISL_735418 | Hospital Regional do Vale do Paraiba | Instituto Adolfo Lutz, Interdiciplinary Procedures Center, Strategic Laboratory | Claudia Regina Gonçalves; Claudio Tavares Sacchi; Erica Valessa Ramos Gomes; Karoline Rodrigues Campos |
| EPI_ISL_471552, EPI_ISL_523967 | Hospital Sancta Maggiore | Instituto Adolfo Lutz, Interdiciplinary Procedures Center, Strategic Laboratory | Claudia Regina Gonçalves; Claudio Tavares Sacchi; Erica Valessa Ramos Gomes |
| EPI_ISL_603036 | Hospital Santa Ana | Instituto Adolfo Lutz, Interdiciplinary Procedures Center, Strategic Laboratory | Claudia Regina Gonçalves; Claudio Tavares Sacchi; Erica Valessa Ramos Gomes; Karoline Rodrigues Campos |
| EPI_ISL_515527, EPI_ISL_693196 | Hospital Santa Clara | Instituto Adolfo Lutz, Interdiciplinary Procedures Center, Strategic Laboratory | Claudia Regina Gonçalves; Claudio Tavares Sacchi; Erica Valessa Ramos Gomes; Karoline Rodrigues Campos |
| EPI_ISL_693233 | Hospital Santa Cruz | Instituto Adolfo Lutz, Interdiciplinary Procedures Center, Strategic Laboratory | Claudia Regina Gonçalves; Claudio Tavares Sacchi; Erica Valessa Ramos Gomes; Karoline Rodrigues Campos |
| EPI_ISL_693236 | Hospital Santa Marcelina Sao Paulo | Instituto Adolfo Lutz, Interdiciplinary Procedures Center, Strategic Laboratory | Claudia Regina Gonçalves; Claudio Tavares Sacchi; Erica Valessa Ramos Gomes; Karoline Rodrigues Campos |
| EPI_ISL_468310 | Hospital Sao Paulo de Ensino da UNIFESP | Instituto Adolfo Lutz, Interdiciplinary Procedures Center, Strategic Laboratory | Claudia Regina Gonçalves; Claudio Tavares Sacchi; Erica Valessa Ramos Gomes |
| EPI_ISL_471545, EPI_ISL_471551, EPI_ISL_515528, EPI_ISL_515545, EPI_ISL_515559, EPI_ISL_515560, EPI_ISL_523969, EPI_ISL_523981, EPI_ISL_523988, EPI_ISL_693201 | | | |
| see above | Hospital Sao Paulo de Ensino da Unifesp | Instituto Adolfo Lutz, Interdiciplinary Procedures Center, Strategic Laboratory | Claudia Regina Gonçalves; Claudio Tavares Sacchi; Erica Valessa Ramos Gomes; Karoline Rodrigues Campos |
| EPI_ISL_414015, EPI_ISL_414016, EPI_ISL_414017 | Hospital São Joaquim Beneficencia Portuguesa | Instituto Adolfo Lutz, Interdiciplinary Procedures Center, Strategic Laboratory | Audrey Cilli; Carlos Henrique Camargo; Claudia Regina Gonçalves; Claudio Tavares Sacchi; Daniela Bernardes Borges da Silva; Ester Cerdeira Sabino; Fabiana Cristina Pereira dos Santos; Fabiana Cristina Pereira dos Santos Terezinha Maria de Paiva; Maria do Carmo Sampaito Tavares Timenetsky; SimoneGuadagnucci Morillo; Terezinha Maria de Paiva |
| EPI_ISL_452701 | Hospital Universitario Araba. Vitoria-Gasteiz, | SeqCOVID-SPAIN consortium/IBV(CSIC) | Amaia Aguirre Quiñonero; Andrés Canut Blasco and SeqCOVID-SPAIN consortium; Carmen Gómez González; Maria Concepción Lecaroz Agara; Maria Rosario Almela Ferrer; Marina Fernández Torres; Silvia Hernáez Crespo |
| EPI_ISL_530040, EPI_ISL_530042, EPI_ISL_530060, EPI_ISL_530083 | Hospital Universitario La Paz | Hospital Universitario La Paz | Elias Dahdouh; Esther Viedma; Fernando Lázaro; Jesús Mingorance; Juan Carlos Galán; Julio García; María Rodríguez; Mª Dolores Folgueira; Natalia Stella; Rafael Cantón; Rafael Delgado; Raúl Recio; Sara González |
| EPI_ISL_582106 | Hospital Universitario Marqués de Valdecilla - IDIVAL (Santander, Cantabria) | SeqCOVID-SPAIN consortium/IBV(CSIC) | Daniel Pablo Marcos; Jesús Rodríguez Rodríguez; Jose Manuel Méndez Legaza; María Eliecer Cano García; María Siller Ruiz and SeqCOVID-SPAIN consortium; Mónica Gozalo Margüello |
| EPI_ISL_691729 | Hospital Universitario Severo Ochoa | Instituto de Salud Carlos III | A. Monzón; F. Casas; I. García, M.; I. Jiménez; Iglesias-Caballero; M. Camarero; M. Cuesta; M. González-Esguevillas; M. Pozo; M. Zaballos; P. Jiménez; S. Juliá; S. Molinero Calamita; S. Varona |
| EPI_ISL_510447 | Hospital Universitario Virgen de las Nieves de Granada-SAS | SeqCOVID-SPAIN consortium/IBV(CSIC) | Irene Pedrosa Corral; José M. Navarro-Marí and SeqCOVID-SPAIN consortium; Mercedes Pérez Ruiz; Sara Sanbonmatsu Gámez |
| EPI_ISL_468318, EPI_ISL_468321, EPI_ISL_547574 | Hospital Universitario da USP | Instituto Adolfo Lutz, Interdiciplinary Procedures Center, Strategic Laboratory | Claudia Regina Gonçalves; Claudio Tavares Sacchi; Erica Valessa Ramos Gomes; Karoline Rodrigues Campos |
| EPI_ISL_471539 | Hospital Universitario da USP Sao Paulo | Instituto Adolfo Lutz, Interdiciplinary Procedures Center, Strategic Laboratory | Claudia Regina Gonçalves; Claudio Tavares Sacchi; Erica Valessa Ramos Gomes |
| EPI_ISL_534314 | Hospital Universitario da USP de SP | Instituto Adolfo Lutz, Interdiciplinary Procedures Center, Strategic Laboratory | Claudia Regina Gonçalves; Claudio Tavares Sacchi; Erica Valessa Ramos Gomes |
| EPI_ISL_476373, EPI_ISL_476435, EPI_ISL_476439, EPI_ISL_476445, EPI_ISL_476446, EPI_ISL_476469, EPI_ISL_476490 | | | |

| | | | |
|---|---|---|---|
| see above | Hospital da Clínicas da Faculdade de Medicina da Universidade de São Paulo | Instituto de Medicina Tropical da Univesidade de São Paulo | Camila Alves Maia da Silva; Carolina S. Lazar; Cecilia Salete Alencar; Darlan da Silva Candido; Erika Regina Manuli; Ester Sabino; Flavia Cristina da Silva Sales; Giula Magalhaes Ferreira; Jaqueline Goes de Jesus; Julien Theze; Mariana Severo Ramundo; Nuno Faria; Samples: Ingra Morales Claro; Sequencing: Ingra Morales Claro; Silvia F. Costa; Thais de Moura Coletti |
| EPI_ISL_534313 | Hospital da Sta Casa de Sto Amaro | Instituto Adolfo Lutz, Interdiciplinary Procedures Center, Strategic Laboratory | Claudia Regina Gonçalves; Claudio Tavares Sacchi; Erica Valessa Ramos Gomes |
| EPI_ISL_722129 | Hospital das Clínicas Universidade de São Paulo Medical School | Laboratório de Parasitologia Médica - Instituto de Medicina Tropical - Universidade de São Paulo | Brazil-UK Centre for Arbovirus Discovery Diagnosis Genomics and Epidemiology (CADDE) Genomic Network - Instituto de Medicina Tropical |
| EPI_ISL_693205 | Hospital de Campanha Covid-19 Assis | Instituto Adolfo Lutz, Interdiciplinary Procedures Center, Strategic Laboratory | Claudia Regina Gonçalves; Claudio Tavares Sacchi; Erica Valessa Ramos Gomes; Karoline Rodrigues Campos |
| EPI_ISL_735396 | Hospital de Camplanha COVID 19 SER | Instituto Adolfo Lutz, Interdiciplinary Procedures Center, Strategic Laboratory | Claudia Regina Gonçalves; Claudio Tavares Sacchi; Erica Valessa Ramos Gomes; Karoline Rodrigues Campos |
| EPI_ISL_534319, EPI_ISL_534320 | Hospital do Serv Pub ESTAFCO Morato de Oliveira | Instituto Adolfo Lutz, Interdiciplinary Procedures Center, Strategic Laboratory | Claudia Regina Gonçalves; Claudio Tavares Sacchi; Erica Valessa Ramos Gomes |
| EPI_ISL_693199 | Hospital do Servidor Publico Estadual Francisco Morato de Oliveira | Instituto Adolfo Lutz, Interdiciplinary Procedures Center, Strategic Laboratory | Claudia Regina Gonçalves; Claudio Tavares Sacchi; Erica Valessa Ramos Gomes; Karoline Rodrigues Campos |
| EPI_ISL_471548, EPI_ISL_515565, EPI_ISL_523965, EPI_ISL_523972, EPI_ISL_523978, EPI_ISL_523982, EPI_ISL_524470 | | | |
| see above | Hospital do Servidor Público Estadual Francisco Morato de Oliveira | Instituto Adolfo Lutz, Interdiciplinary Procedures Center, Strategic Laboratory | Claudia Regina Gonçalves; Claudio Tavares Sacchi; Erica Valessa Ramos Gomes |
| EPI_ISL_527861 | Hospital e Maternidade Celso Pierro | Instituto Adolfo Lutz, Interdiciplinary Procedures Center, Strategic Laboratory | 01246-1301; 355 - Brazil; Av. Dr. Arnaldo; Cerqueira Cesar; São Paulo - SP |
| EPI_ISL_693200 | Hospital e Maternidade Mairipora | Instituto Adolfo Lutz, Interdiciplinary Procedures Center, Strategic Laboratory | Claudia Regina Gonçalves; Claudio Tavares Sacchi; Erica Valessa Ramos Gomes; Karoline Rodrigues Campos |
| EPI_ISL_547577 | Hospital e Maternidade Nossa Senhora das Graças | Instituto Adolfo Lutz, Interdiciplinary Procedures Center, Strategic Laboratory | Claudia Regina Gonçalves; Claudio Tavares Sacchi; Erica Valessa Ramos Gomes; Karoline Rodrigues Campos |
| EPI_ISL_693241, EPI_ISL_735425 | Hospital e Maternidade Sao Lucas | Instituto Adolfo Lutz, Interdiciplinary Procedures Center, Strategic Laboratory | Claudia Regina Gonçalves; Claudio Tavares Sacchi; Erica Valessa Ramos Gomes; Karoline Rodrigues Campos |
| EPI_ISL_527865 | Hospital e Maternidade São Cristóvão | Instituto Adolfo Lutz, Interdiciplinary Procedures Center, Strategic Laboratory | Claudia Regina Gonçalves; Claudio Tavares Sacchi; Erica Valessa Ramos Gomes |
| EPI_ISL_527868 | Hospital e Maternidade do Braz | Instituto Adolfo Lutz, Interdiciplinary Procedures Center, Strategic Laboratory | Claudia Regina Gonçalves; Claudio Tavares Sacchi; Erica Valessa Ramos Gomes |
| EPI_ISL_534323 | Hospital e Pronto Socorro Comunitario Vila Yolanda | Instituto Adolfo Lutz, Interdiciplinary Procedures Center, Strategic Laboratory | Claudia Regina Gonçalves; Claudio Tavares Sacchi; Erica Valessa Ramos Gomes |
| EPI_ISL_527864 | Hospital e Pronto Socorro Comunitário Vila Iolanda | Instituto Adolfo Lutz, Interdiciplinary Procedures Center, Strategic Laboratory | Claudia Regina Gonçalves; Claudio Tavares Sacchi; Erica Valessa Ramos Gomes |
| EPI_ISL_693195, EPI_ISL_693230, EPI_ISL_693232, EPI_ISL_735412 | Hospital e Pronto Socorro Portinari | Instituto Adolfo Lutz, Interdiciplinary Procedures Center, Strategic Laboratory | Claudia Regina Gonçalves; Claudio Tavares Sacchi; Erica Valessa Ramos Gomes; Karoline Rodrigues Campos |
| EPI_ISL_707709 | Hospital for Tropical Diseases | COVID-19 Network Investigations (CONI) Alliance | Akanitt Jittmittraphap; Angkana Huang; Anthony R. Jones; Arporn Wangwiwatsin; Bhakbhoom Panthan; Chonticha Klungtong; Ekawat Pasomsub; Elizabeth Batty; Insee Sensorn; Janjira Thaipadungpanit; Khajohn Joonsalak; Kingkan Rakmanee; Krittikorn Kumpornsin; Namfon Kotanan; Nantarat Chantawat; Pornsawan Leaungwutiwong; Stefan Fernandez; Thanat Chookajorn; Theerarat Kochakarn; Treewat Watthanachockchai; Viravarn Luvira; Wasun Chantratita; Weena Janwitthayanan; Wudtichai Manasatienkij |
| EPI_ISL_593921 | Hospital, Fameck | National Reference Center for Viruses of Respiratory Infections, Institut Pasteur, Paris | Etienne Simon-Lorière; Fabiana Gambaro; Maud Vanpeene; Sylvie Behillil; Sylvie van der Werf; Vincent Enouf |
| EPI_ISL_784739, EPI_ISL_788322, EPI_ISL_788716, EPI_ISL_789289, EPI_ISL_789426 | Houston Methodist Hospital | Houston Methodist Hospital | David W. Bernard; Heather Hendrickson; James J. Davis; Layne Pruitt; Marcus Nguyen; Matthew Ojeda Saavedra; Maulik Shukla; Paul A. Christensen; Prasanti Yerramilli; Randall J. Olsen; S. Wesley Long; Sishir Subedi; and James M. Musser |
| EPI_ISL_469059 | Hovas Askim Familjelakare och BVC | The Public Health Agency of Sweden | Anna Risberg; Anna-Malin Linde; Karin Tegmark-Wisell; Maria Lind Karlberg; Mattias Haukland; Olov Svartstrom; Oskar Karlsson Lindsjo; Petra Edquist; Reza Advani; Sandra Broddesson; Shamam Muradrasoli |
| EPI_ISL_645000, EPI_ISL_645057, EPI_ISL_645072, EPI_ISL_645093 | Human Genome Variation Research Group, Malopolska Centre of Biotechnology | Human Genome Variation Research Group, Malopolska Centre of Biotechnology | Botwina, P.; Branicki, W.; Dabrowska, A.; Foremny, J.; Gromowski, T.; Klajmon, A.; Kopera, K.; Kowalski, M.; Labaj; Marszalek, K.; Owczarek, K.; P.P.; Pisarek, A.; Pospiech, E.; Pyrc, K.; Sanak, M.; Swadzba, J.; Szczepanski, A. |
| EPI_ISL_526222, EPI_ISL_526223, EPI_ISL_526224, EPI_ISL_526225, EPI_ISL_526236, EPI_ISL_526237, EPI_ISL_526238 | | | |
| see above | Hungarian Defence Forces Military Medical Centre | National Laboratory of Virology, Szentágothai Research Centre | Balázs Somogyi; Bálint Eszenyi; Endre Gábor Tóth; Ferenc Jakab; Gábor Kemenesi |
| EPI_ISL_710532, EPI_ISL_710540, EPI_ISL_710575 | Hôpital Fattouma-Bourguiba de Monastir | Laboratoire des Procédés de Criblage Moléculaire et Cellulaire-Centre de Biotechnologie de Sfax | A. and Masmoudi, S.; Abdelmoulah, F.; Abid, N.; Ajili, F.; Aouni, M.; Ben Ayed, I.; Bensaid, M.; Chtourou, A.; Elargoubi, A.; Fki-berrajah, L.; Gaaloul, I.; Gargouri, S.; Hammami, A.; Kamoun, S.; Karray Hakim, H.; Kharat, N.; Mastouri, M.; Mhalla, S.; Nabli, A.; Rebai; Smeti, I.; Souissi, A.; Stambouli, N.; Turki, M. |
| EPI_ISL_415650 | Hôpital Instruction des Armées - BEGIN | National Reference Center for Viruses of Respiratory Infections, Institut Pasteur, Paris | Angela Brisebarre; Christine Bigaillon; Flora Donati Vincent Enouf; Marion Barbet; Maud Vanpeene; Méline Bizard; Mélnie Albert; Sylvie Behillil; Sylvie van der Werf |
| EPI_ISL_535718 | Hôpital Pierre-Boucher | Laboratoire de santé publique du Québec | Guillaume Bourque; Ioannis Ragoussis; Jesse Shapiro; Mark Lathrop and Michel Roger; Sandrine Moreira |
| EPI_ISL_653819 | I.R.C.C.S. "S. De Bellis" - Ente Ospedaliero | Istituto Zooprofilattico Sperimentale della Puglia e della Basilicata | Bianco A.; Capozzi L.; Cipolletta D.; Del Sambro L.; Galante D.; Lippolis A.; Notarnicola M.; Parisi A. |
| EPI_ISL_404253, EPI_ISL_410045 | IL Department of Public Health Chicago Laboratory | Pathogen Discovery, Respiratory Viruses Branch, Division of Viral Diseases, Centers for Dieases Control and Prevention | Anna Uehara; Brett L. Whitaker; Brian Lynch; Clinton R. Paden; Janna' R. Murray; Jing Zhang; Krista Queen; Lijuan Wang; Senthil Kumar K. Sakthivel; Shifaq Kamili; Stephen Lindstrom; Susan I. Gerber; Suxiang Tong; Xiaoyan Lu; Yan Li; Ying Tao |
| EPI_ISL_648677 | INBIRS-UBA | Laboratorio Mixto de Biotecnología Acuática (LMBA) | Adriana Giri; Agustina Cerri; Ana Cavatorta; Ana Paletta; Diego Chouhy; Elisa Bolatti; Elizabeth Tapia; Federico Remes Lenicov; Flavio Spetale; Gastón Viarengo; Ignacio García Labari; Javier Murillo; Joaquín Ezpeleta; Julian Acosta; Laura Angelone; Leandro Ciappina; María Re; Pablo Casal; Pilar Bulacio; Silvana Spinelli; Silvia Arranz; Sofía Lavista Llanos; Vanina Villanova; Victoria Posner |
| EPI_ISL_590695, EPI_ISL_603143, EPI_ISL_603157, EPI_ISL_603169, EPI_ISL_603170, EPI_ISL_603182, EPI_ISL_609989, EPI_ISL_609997 | | | |
| see above | INMI Lazzaro Spallanzani IRCCS | INMI Lazzaro Spallanzani IRCCS | A Di Caro; Antonino Di Caro; B Bartolini; Barbara Bartolini; Beatrice Valli; C.E.M Gruber; Cesare E.M. Gruber; E Giombini; Eleonora Lalle; Emanuela Giombini; F Messina; Francesco Messina; Francesco Vairo; Fulvia Pimpinelli; M Rueca; MR Capobianchi; Maria R. Capobianchi; Martina Rueca; Simone Lanini |
| EPI_ISL_732115 | INSA | Instituto Nacional de Saude (INSA) | Borges et al |
| EPI_ISL_693576 | IPO Lisboa | Instituto Nacional de Saude (INSA) | Borges et al |
| EPI_ISL_751331, EPI_ISL_751338, EPI_ISL_751339 | IRCCS Sacro Cuore Don Calabria Hospital, Department of Infectious, Tropical Diseases & Microbiology | University of Verona, Department of Biotechnology | Antonio Mori; Chiara Degli Esposti; Chiara Piubelli; Cristina Beltrami; Elena Pomari; Emanuela Cosentino; Giulia Lopatriello; Luca Marcolungo; Massimo Delledonne; Michela Deiana |
| EPI_ISL_584071 | IZSM | IZSM | Giovanna Fusco; Lorena Cardillo; Maurizio Viscardi |
| EPI_ISL_738126 | IZSM-U.O.C. Virologia | Istituto Zooprofilattico Sperimentale del Mezzogiorno | Giovanna Fusco; Lorena Cardillo; Maurizio Viscardi |
| EPI_ISL_479941 | Ibaraki Prefectural Institute of Public Health | Pathogen Genomics Center, National Institute of Infectious Diseases | Hajime Kamiya; Keiko Goto; Kentaro Itokawa; Makoto Kuroda; Masanori Hashino; Motoi Suzuki; Rina Tanaka; Tsuyoshi Sekizuka |
| EPI_ISL_476702, EPI_ISL_476704 | Incubadora Venezolana de Ciencia, Venezuela | Incubadora Venezolana de Ciencia, Venezuela / Instituto Nacional de Salud, Bogotá, Colombia / Grupo de Investigaciones Microbiológicas-UR (GIMUR), Departamento de Biología, Facultad de Ciencias Naturales, Universidad del Rosario, Bogotá, Colombia / Icahn School of Medicine at Mount Sinai, New York, USA | Alberto Paniz-Mondolfi; Ana S. Gonzalez-Reiche; Angelica Rico; Aníbal A. Teherán; Carolina Florez; Carolina Hernández; Emilia Mia Sordillo; Esther C. Barros; Harm van Bakel; Jesús E. Jaimes; Juan David Ramírez; Lisseth Pardo; Lourdes Delgado; Luis Perez-Garcia; Marina Muñoz; Matthew M. Hernandez; Sergio Gomez; Viviana Simon |
| EPI_ISL_413522 | Indian Council of Medical Research - National Institute of Virology | National Influenza Center, Indian Council of Medical Research - National Institute of Virology | Choudhary ML; Potdar V; Shete-Aich A; Yadav PD |
| EPI_ISL_413523 | Indian Council of Medical Research-National Institute of Virology | National Influenza Center, Indian Council of Medical Research-National Institute of Virology | Choudhary ML; Potdar V; Shete-Aich A; Yadav PD |
| EPI_ISL_637100 | Indian Council of Medical Research-National Institute of Virology, Microbial Containment Complex | Indian Council of Medical Research-National Institute of Virology, Microbial Containment Complex | Anita Shete-Aich; Dimpal A. Nyayanit; Gururaj Rao Deshpande; Padinjaremattathil Thankappan Ullas; Pragya D. Yadav; Prasad Sarkale; Priya Abraham; Varsha Potdar |
| EPI_ISL_413521, EPI_ISL_514752 | Infectious Disease Control Center, Center for Disease Control and Prevention of PLA | Infectious Disease Control Center, Center for Disease Control and Prevention of PLA | Li; Li, J.; Li, L.; Li, P.; Li, Z.; P. and Li, P.; Qiu, S.; Song, H. |
| EPI_ISL_496374 | Infectolab | Andersen lab at Scripps Research | Carlos A. Cota Haros; Octavio Renteria Pacheco; SEARCH Alliance San Diego with Samuel Navarro Alvarez |
| EPI_ISL_708153 | Innlandet Hospital Trust, Division Lillehammer, Department for Medical Microbiology | Norwegian Institute of Public Health, Department of Virology | Hilde Elshaug; Hilde Vollan; Kamilla Heddeland Instefjord; Karoline Bragstad; Kathrine Stene-Johansen; Marie Paulsen Madsen; Olav Hungnes; Rasmus Riis Kopperud |
| EPI_ISL_418209, EPI_ISL_418210, EPI_ISL_418213, EPI_ISL_420078, EPI_ISL_476149, EPI_ISL_480556, EPI_ISL_482875, EPI_ISL_482876, EPI_ISL_486865, EPI_ISL_486866, EPI_ISL_486871, EPI_ISL_486872 | | | |
| see above | Institut Pasteur Dakar | Institut Pasteur de Dakar | Amadou Alpha Sall; Amadou Alpha Sall.; Mamadou Diop; Mamadou Malado Jallow; Mamadou malado Jallow; Marie Henriette Dior Ndione; Moussa Moise Diagne; Ndongo Dia; Ousmane Faye; Safietou Sanke; Safietou Sankhe |
| EPI_ISL_498238 | Institut Pasteur de Dakar | Institut Pasteur de Dakar | Amadou Alpha Sall.; Mamadou Diop; Mamadou Malado Jallow; Marie Henriette Dior Ndione; Moussa Moise Diagne; Ndongo Dia; Ousmane Faye; Safietou Sankhe Mbengue |
| EPI_ISL_613423, EPI_ISL_613429, EPI_ISL_613433, EPI_ISL_613445, EPI_ISL_613446, EPI_ISL_613448, EPI_ISL_613456 | | | |
| see above | Institut Pasteur de la Guadeloupe | Institut Pasteur de la Guadeloupe | Angela Brisebarre; Antoine Talarmin; Camille Capel; Cherina Fleming; Etienne Simon-Lorière; Marion Barbet; Maud Vanpeene; Méline Bizard; Radjin Steingrover; Stéphanie Guyomard; Sylvie Behillil; Sylvie van der Werf; Sébastien Breurec; Vincent Enouf |
| EPI_ISL_459967, EPI_ISL_459973, | Institut Pasteur du Maroc | Institut Pasteur du Maroc | Abdellah Faouzi; Anass Abbad; Anderrahmane Maaroufi; Angela Brisebarre; Camille Capel; Etienne Simon-Lorière; Jalal Nourlil; Latifa Anga; Marion Barbet; Maud Vanpeene; Mjid Eloualid; Méline Bizard; Sylvie Behillil; Sylvie van der Werf; Vincent Enouf |

| | | | |
|---|---|---|---|
| EPI_ISL_459984 | | | |
| EPI_ISL_418429, EPI_ISL_508891 | Institut des Agents Infectieux (IAI), Hospices Civils de Lyon | CNR Virus des Infections Respiratoires - France SUD | Alexandre Gaymard; Antonin Bal; Bruno Lina; Carine Moustaud; Florence Morfin-Sherpa; Gregory Destras; Gwendolyne Burfin; Laurence Josset; Martine Valette; Maude Bouscambert-Duchamp; Raphaelle Lamy; Solenne Brun |
| EPI_ISL_437932, EPI_ISL_475828, EPI_ISL_583616 | Institut für Virologie am Department für Hygiene, Mikrobiologie und Public Health | Bergthaler laboratory, CeMM Research Center for Molecular Medicine of the Austrian Academy of Sciences | Adi Steinrigl; Alexander Lercher; Alexandra Popa; Andreas Bergthaler; Benedikt Agerer; Christian Paar; Christoph Bock; Daniela Schmid; Dorothee von Laer; Elisabeth Puchhammer-Stoeckl; Franz Allerberger; Gernot Walder; Gregor Hörmann; Guenter Weiss; Gunther Vogl; Henrique Colaco; Jakob-Wendelin Genger; Jan Laine; Judith Aberle; Kinga Rigler-Hohenwarter; Lukas Endler; Manfred Nairz; Mark Smyth; Martin Senekowitsch; Michael Schuster; Peter Hufnagl; Peter Obrist; Rainer Gattringer; Sabine Sussitz-Rack; Stephan Aberle; Thomas Penz; Wegene Borena |
| EPI_ISL_574820 | Institute for Infectious Diseases, University of Bern | Institute for Infectious Diseases, University of Bern | Alban Ramette; Christian Baumann; Cora Sägesser; Franziska Suter-Riniker; Michel C Koch; Miguel A Terrazos Miani; Peter Keller; Stephen L Leib |
| EPI_ISL_583860 | Institute for Laboratory Diagnostics and Microbiology, Klinikum Klagenfurt am Worthersee | Bergthaler laboratory, CeMM Research Center for Molecular Medicine of the Austrian Academy of Sciences | Adi Steinrigl; Alexander Lercher; Alexandra Popa; Andreas Bergthaler; Benedikt Agerer; Christian Paar; Christoph Bock; Daniela Schmid; Dorothee von Laer; Elisabeth Puchhammer-Stoeckl; Franz Allerberger; Gernot Walder; Gregor Hörmann; Guenter Weiss; Gunther Vogl; Henrique Colaco; Jakob-Wendelin Genger; Jan Laine; Judith Aberle; Kinga Rigler-Hohenwarter; Lukas Endler; Manfred Nairz; Mark Smyth; Martin Senekowitsch; Michael Schuster; Peter Hufnagl; Peter Obrist; Rainer Gattringer; Sabine Sussitz-Rack; Stephan Aberle; Thomas Penz; Wegene Borena |
| EPI_ISL_455791 | Institute for Medical Research, Infectious Disease Research Centre, National Institutes of Health, Ministry of Health Malaysia | Malaysia Genome Institute | Azrin Ahmad; Enizza Kasim; Irni Suhayu Sapian; Jeyanthi Suppiah; Mohd Faizal Abu Bakar; Mohd Noor Mat Isa; Norazah Ahmad; Nurhezreen Md Iqbal; Ravindran Thayan; Shahrul Hisham Zainal Ariffin; Shamsidar Sopie; Siti Noraini Othman; Tahir Aris; Yusuf Muhammad Noor; Zarina Mohd Zawawi |
| EPI_ISL_430442, EPI_ISL_430444, EPI_ISL_489992, EPI_ISL_490089, EPI_ISL_490100, EPI_ISL_490103, EPI_ISL_718267, EPI_ISL_718268, EPI_ISL_718272, EPI_ISL_718280, EPI_ISL_718282, EPI_ISL_718285, EPI_ISL_718298, EPI_ISL_728161, EPI_ISL_728247, EPI_ISL_728251 | | | |
| see above | Institute for Medical Research, Infectious Disease Research Centre, National Institutes of Health, Ministry of Health Malaysia | Institute for Medical Research, Infectious Disease Research Centre, National Institutes of Health, Ministry of Health Malaysia | Abdul-Wahid.M-Z; Ahmad.N; Azizan.M-A; Hisham.H-A; Kalyanasundram J; Kalyanasundram.J; Kamel K; Mat-Sharani.S; Mohd-Zain.R; Mohd-Zawawi Z; Mohd-Zawawi.Z; Suppiah J; Suppiah.J; Tan.L-P; Tengku-Rogayah.TAR; Thayan R; Thayan.R |
| EPI_ISL_454606, EPI_ISL_468656 | Institute for Public Health | Laboratory for advanced genomics | Filip Rokić; Igor Jurak; Lovro Trgovec-Greif; Neven Sučić; Oliver Vugrek; Tomislav Rukavina |
| EPI_ISL_492076 | Institute for Public Health of the Republic of North Macedonia | Charite Universitatsmedizin Berlin, Institute of Virology | Barbara Mühlemann; Christian Drosten; Elizabeta Jancheska; Golubinka Bosevska; Joern Beheim-Schwarzbach; Julia Schneider; Maja Kuzmanovska; Talitha Veith; Terry Jones; Victor M Corman |
| EPI_ISL_707707 | Institute for Urban Disease Control and Prevention | COVID-19 Network Investigations (CONI) Alliance | Amornmas Kongklieng; Anek Mungaomklang; Angkana Huang; Anthony R. Jones; Arporn Wangwiwatsin; Bhakbhoom Panthan; Chonticha Klungtong; Ekawat Pasomsub; Elizabeth Batty; Insee Sensorn; Janjira Thaipadungpanit; Kamolthip Atsawawaranunt; Khajohn Joonsalak; Kingkan Rakmanee; Krittikorn Kumpornsin; Namfon Kotanan; Prayuth Kaewmalang; Pukkaporn Parnwijitkul; Stefan Fernandez; Thanat Chookajorn; Theerarat Kochakarn; Treewat Watthanachockchai; Wasun Chantratita; Wudtichai Manasatienkij |
| EPI_ISL_602472 | Institute for Virology, University Hospital Essen | Center of Medical Microbiology, Virology, and Hospital Hygiene, University of Duesseldorf | Alexander Dilthey; Andreas Walker; Daniel Strelow; Jessica Nicolai; Jörg Timm; Klaus Pfeffer; Lisanna Hülse; Malte Kohns Vasconcelos; Maximilian Damagnez; Nadine Lübke; Olympia E. Anastasiou; Tobias Wienemann; Torsten Houwaart; Ulf Dittmer |
| EPI_ISL_539333, EPI_ISL_539334, EPI_ISL_539338 | Institute of Disease Control and Prevention, People's Liberation Army | Institute of Disease Control and Prevention, People's Liberation Army | Li, P.; Qiu, S. |
| EPI_ISL_485399 | Institute of Human Genetics, Polish Academy of Sciences | Institute of Human Genetics, Polish Academy of Sciences | Adam Ustaszewski; Andrzej Pławski; Emilia Lis; Ewa Ziętkiewicz; Marta Kaczmarek-Ryś; Michał Witt; Szymon Hryhorowicz |
| EPI_ISL_450294 | Institute of Human Genetics, Polish Academy of Sciences Sanitary and Epidemiological Station in Poznań | Institute of Human Genetics, Polish Academy of Sciences | Adam Ustaszewski; Andrzej Pławski; Emilia Lis; Marta Kaczmarek-Ryś; Michał Witt; Szymon Hryhorowicz |
| EPI_ISL_463038 | Institute of Life Sciences, Bhubaneswar | Immunogenomics lab, Institute of Life Sciences, Bhubaneswar | Ajay Parida; Aliva Minz; Arup Ghosh; Atimukta Jha; DBT's PAN-INDIA 1000 SARS-CoV2 RNA genome sequencing consortium; Debyashrita Barik; Ghulam Hussain Syed; ILS COVID-19 TEAM; Manasi Priyadarshini; O. P. Shriwas; Orissa COVID-19 Study Group; Priyanka Mohapatra; Punit Prasad; Rajeeb Swain; Rupesh Dash; Satya Ranjan Sahu; Shanti Senapati; Shuchi Smita; Soma Chattopadhyay; Sunil Raghav; Swati Madhulika; Tushar K. Beuria; Viplov K. Biswas |
| EPI_ISL_723082 | Institute of Medical Genetics and Applied Genomics | Institute of Medical Genetics and Applied Genomics | Angel Angelov; Caspar Gross; Daniela Bezdan; Michael Bitzer; Michael Sonnabend; Michaela Pogoda; Nicolas Casadei; Siri Göpel; Stephan Ossowski; Thomas Iftner; Tina Ganzenmüller |
| EPI_ISL_635201, EPI_ISL_635228, EPI_ISL_635254, EPI_ISL_635270, EPI_ISL_635272, EPI_ISL_635294, EPI_ISL_635299 | | | |
| see above | Institute of Microbiology and Immunology, Faculty of Medicine, University of Ljubljana | Institute of Microbiology and Immunology, Faculty of Medicine, University of Ljubljana | Mario Poljak; Miša Korva; Samo Zakotnik; Tatjana Avšič - Županc; Tatjana Avšič – Županc; Tomaž Mark Zorec |
| EPI_ISL_548944 | Institute of Microbiology, University of Veterinary and Animal sciences | Institute of Microbiology, University of Veterinary and Animal sciences | Ali; Altaf, I.; Anwar, M.; Ashraf; Asif, A.; Attique; Awan; Aziz; Bhatti; Cheema; Fazal, S.; Hassan, S.; Khan; Khan, N.; M.A.; M.B.; M.M.; M.S.; M.T.; M.U.; M.W.; M.Z.; Mehmood, A.; Mukhtar, N.; N.A.; Nawaz, M.; Rafique, S.; Rana; Raza, S.; S.Q.; S.Z.; Sardar, N.; Sarwar, H.; Shabbir; Shah; Tahir, Z.; Yaqub, T.; Younis |
| EPI_ISL_477014, EPI_ISL_477015, EPI_ISL_486843, EPI_ISL_486846, EPI_ISL_516649, EPI_ISL_516650, EPI_ISL_516652, EPI_ISL_525432, EPI_ISL_539786, EPI_ISL_660533, EPI_ISL_660535, EPI_ISL_697793, EPI_ISL_697794 | | | |
| see above | Institute of Microbiology, Universidad San Francisco de Quito | Institute of Microbiology, Universidad San Francisco de Quito | Alejandra Ramones; Alexandra Tino; Andrea Macias; Belen Prado-Vivar; Belén Prado-Vivar; Bernardo Gutierrez; Bernardo Gutiérrez; Carla Torres; Carlos Mena; Edison Ligña; Edy Quizhpe; Fausto Maldonado; Francisco Mora; Franklin Espinoza; Freddy Iza; Gabriel Trueba; Geovanny Carzola; Hermelinda Paguay; Jonathan Araujo; Jorge Luis Velez; Jorge Montaño; Jorge Reyes; Juan Gaviria; Juan Jose Guadalupe; Juan José Guadalupe; Ligia Briceño; Michelle Grunauer; Monica Becerra-Wong; Nabih Dahik; Patricio Rojas-Silva; Paul Cardenas; Paúl Cárdenas; Prado-Vivar; Sully Marquez; Sully Márquez; Veronica Barragan; Verónica Barragán; Yomara Napa |
| EPI_ISL_508394, EPI_ISL_508397 | Institute of Post Graduate Medical Education & Research | National Institute of Biomedical Genomics | Arindam Maitra; Aritra Biswas; Jayeeta Haldar; Monimoy Banerjee; Raja Ray; Saumitra Das |
| EPI_ISL_672705, EPI_ISL_672711, EPI_ISL_672719, EPI_ISL_672720, EPI_ISL_672748 | Institute of Tropical Medicine at the University of São Paulo (IMT-USP) | Laboratório de Parasitologia Médica - Instituto de Medicina Tropical - Universidade de São Paulo | Brazil-UK Centre for Arbovirus Discovery Diagnosis Genomics and Epidemiology (CADDE) Genomic Network - Instituto de Medicina Tropical |
| EPI_ISL_408514 | Institute of Viral Disease Control and Prevention, China CDC | Institute of Viral Disease Control and Prevention, China CDC | Beiwei Ye; Dayan Wang; George F. Gao; Guizhen Wu; Juan Song; Jun Han; Mengjie Yang; Peihua Niu; Peipei Liu; Roujian Lu; Shumei Zou; Wei Zhen; Weifeng Shi; Weimin Zhou; Wenbo Xu; Wenjie Tan; Wenwen Lei; William J. Liu; Xiang Zhao; Yingze Zhao; Yuchao Wu; Zhixiao Chen; Ziqian Xu |
| EPI_ISL_577734, EPI_ISL_577736, EPI_ISL_577737, EPI_ISL_577738, EPI_ISL_577739, EPI_ISL_577741, EPI_ISL_577742, EPI_ISL_583482, EPI_ISL_583485, EPI_ISL_718253, EPI_ISL_718260, EPI_ISL_788985 | | | |
| see above | Institute of Virology, Biomedical Research Center of the Slovak Academy of Sciences, Bratislava | Faculty of Natural Sciences, Comenius University, Bratislava | Alena Koščálová; Boris Klempa; Broňa Brejová; Dominika Fričová; Edita Staroňová; Elena Tichá; Jozef Nosek; Juraj Kopáček; Kristína Boršová; Martina Ličková; Martina Neboháčová; Monika Sláviková; Peter Sabaka; Sabina Fumačová Havlíková; Tomáš Vinař; Viktória Hodorová; Viktória Čabanová; Ľubomíra Lukáčiková |
| EPI_ISL_417880 | Institute of Virology, Biomedical Research Center of the Slovak Academy of Sciences, Bratislava; Public Health Authority of the Slovak Republic, Bratislava | Institute of Virology, Biomedical Research Center of the Slovak Academy of Sciences, Bratislava; Comenius University Science Park, Bratislava | Boris Klempa; Diana Rusňáková; Edita Staroňová; Elena Tichá; Jaroslav Budiš; Juraj Kopáček; Juraj Koči; Martina Ličková; Miroslav Böhmer; Monika Slávikova; Sabina Fumačová Havlíková; Tomáš Szemeš; Werner Krampl |
| EPI_ISL_455680 | Institute of pathogenic microbiology, Jiangsu Provincial Center for Disease Control and Prevention | Institute of pathogenic microbiology, Jiangsu Provincial Center for Disease Control and Prevention | Cui, L. |
| EPI_ISL_755640, EPI_ISL_755642, EPI_ISL_755643, EPI_ISL_755651, EPI_ISL_755653, EPI_ISL_755654, EPI_ISL_776750, EPI_ISL_776751, EPI_ISL_776752, EPI_ISL_776753, EPI_ISL_776754, EPI_ISL_776755, EPI_ISL_776756, EPI_ISL_776759, EPI_ISL_776760, EPI_ISL_776761, EPI_ISL_776762, EPI_ISL_776763, EPI_ISL_792101, EPI_ISL_792102, EPI_ISL_792104, EPI_ISL_792105, EPI_ISL_792106, EPI_ISL_792107, EPI_ISL_792108, EPI_ISL_792109, EPI_ISL_792110, EPI_ISL_792111, EPI_ISL_792112, EPI_ISL_792113, EPI_ISL_792114 | | | |
| see above | Instituto Adolfo Lutz - Central | Instituto Adolfo Lutz, Interdiciplinary Procedures Center, Strategic Laboratory | Claudia Regina Gonçalves; Claudio Tavares Sacchi; Erica Valessa Ramos Gomes; Karoline Rodrigues Campos |
| EPI_ISL_776768 | Instituto Adolfo Lutz - Regional de Aracatuba | Instituto Adolfo Lutz, Interdiciplinary Procedures Center, Strategic Laboratory | Claudia Regina Gonçalves; Claudio Tavares Sacchi; Erica Valessa Ramos Gomes; Karoline Rodrigues Campos |
| EPI_ISL_755655 | Instituto Adolfo Lutz - Regional de Campinas | Instituto Adolfo Lutz, Interdiciplinary Procedures Center, Strategic Laboratory | Claudia Regina Gonçalves; Claudio Tavares Sacchi; Erica Valessa Ramos Gomes; Karoline Rodrigues Campos |
| EPI_ISL_776757, EPI_ISL_776758, EPI_ISL_776767 | Instituto Adolfo Lutz - Regional de Marilia | Instituto Adolfo Lutz, Interdiciplinary Procedures Center, Strategic Laboratory | Claudia Regina Gonçalves; Claudio Tavares Sacchi; Erica Valessa Ramos Gomes; Karoline Rodrigues Campos |
| EPI_ISL_735401, EPI_ISL_735402, EPI_ISL_735403, EPI_ISL_735404, EPI_ISL_735410 | Instituto Adolfo Lutz - Regional de Rio Claro | Instituto Adolfo Lutz, Interdiciplinary Procedures Center, Strategic Laboratory | Claudia Regina Gonçalves; Claudio Tavares Sacchi; Erica Valessa Ramos Gomes; Karoline Rodrigues Campos |
| EPI_ISL_755641, EPI_ISL_755646, EPI_ISL_755647, EPI_ISL_755649, EPI_ISL_776764, EPI_ISL_776765, EPI_ISL_776766, EPI_ISL_776769, EPI_ISL_792103 | | | |
| see above | Instituto Adolfo Lutz - Regional de Santo Andre | Instituto Adolfo Lutz, Interdiciplinary Procedures Center, Strategic Laboratory | Claudia Regina Gonçalves; Claudio Tavares Sacchi; Erica Valessa Ramos Gomes; Karoline Rodrigues Campos |
| EPI_ISL_735400, EPI_ISL_735427, EPI_ISL_735430 | Instituto Adolfo Lutz - Regional de Santos | Instituto Adolfo Lutz, Interdiciplinary Procedures Center, Strategic Laboratory | Claudia Regina Gonçalves; Claudio Tavares Sacchi; Erica Valessa Ramos Gomes; Karoline Rodrigues Campos |
| EPI_ISL_755648, EPI_ISL_755650, EPI_ISL_792115, EPI_ISL_792116 | Instituto Adolfo Lutz - Regional de Taubate | Instituto Adolfo Lutz, Interdiciplinary Procedures Center, Strategic Laboratory | Claudia Regina Gonçalves; Claudio Tavares Sacchi; Erica Valessa Ramos Gomes; Karoline Rodrigues Campos |
| EPI_ISL_491251, EPI_ISL_491263 | Instituto Gulbenkian de Ciência | Instituto Gulbenkian de Ciência | Cathy Paulino; Joao Sobral; João Costa; Ricardo Leite; Susana Ladeiro |
| EPI_ISL_792353 | Instituto Nacional de Epidemiología Dr. Jara | Área de Secuenciación del Laboratorio de Virología del Hospital de Niños Dr. Ricardo Gutierrez on behalf of 'Proyecto Argentino Interinstitucional de genomica de SARS-CoV-2' (PAIS Consortium) | CJ; Cimmino; Goya; I; LE; Lusso; MI; MS; Nabaes Jodar; Natale; O; Pagano; S; Uez; Valinotto; Viegas, M. |
| EPI_ISL_491945, EPI_ISL_491946, EPI_ISL_491949, EPI_ISL_491953 | Instituto Nacional de Investigación en Salud Pública - INSPI | INSPI - Charité | Alberto Orlando; Alexandra Usiña; Alfredo Bruno Caicedo; Andres Moreira-Soto; Anna-Lena Sander; Denisses Portugal; Domenica de Mora Coloma; Jan Felix Drexler; Juan Carlos Zeballos; Manuel Gonzalez; Maritza Olmedo; Nina Krause; Silvia Salgado |

| | | | |
|---|---|---|---|
| EPI_ISL_522980 | Instituto Nacional de Medicina Genómica | Instituto Nacional de Medicina Genómica | Cedro-Tanda A; Cisneros-Villanueva M; Herrera-Montalvo LA; Hidalgo-Miranda A; Hurtado-Cordova E; Mendoza-Vargas A; Peñaloza-Figueroa F; Reyes-Grajeda JP |
| EPI_ISL_536482, EPI_ISL_536486, EPI_ISL_536518, EPI_ISL_536521, EPI_ISL_536545, EPI_ISL_536557 | Instituto Nacional de Salud | Laboratorio de Infecciones Respiratorias Agudas | David Tarazona; Dennis Carhuaricra; Eduardo Juscamayta Lopez; Faviola Valdivia Guerrero; Lenin Maturrano Hernandez; Nancy Rojas Serrano; Ronnie Gavilan Chavez |
| EPI_ISL_791087, EPI_ISL_791093 | Instituto Nacional de Salud - Unidad de Secuenciación y Análisis Genómico | Instituto Nacional de Salud - Dirección de Investigación en Salud Pública | Carlos Franco-Muñoz; Diego A. Álvarez-Díaz; Diego Andrés Prada; Gerardo Santamaría; Jonathan Reales; Julian Naizaque; Katherine Laiton-Donato; Magdalena Wiesner; Marcela Mercado-Reyes; Maria T. Herrera; Martha Lucia Ospina Martinez; Mauricio Pacheco-Montealegre; Paola Muñoz-Laiton; Sheryl Corchuelo |
| EPI_ISL_456117, EPI_ISL_456126, EPI_ISL_456138 | Instituto Nacional de Salud - Unidad de Secuenciación y Análisis Genómico | Instituto Nacional de Salud, Universidad Cooperativa de Colombia, Instituto Alexander von Humboldt, Imperial College-London, London School of Hygiene & Tropical Medicine | Astrid C. Flórez; Carlos Franco-Muñoz; Christian Julian Villabona-Arenas; Diana Marcela Walteros-Acero; Diego A. Álvarez-Díaz; Erika Ospitia; Gloria Puerto; Jose A. Usme-Ciro; Juliana Barbosa; Katherine Laiton-Donato; Liz Villabona-Arenas; Luz Dary Rodriguez; Mailyn A.Gonzalez; Marcela Mercado-Reyes.; Martha Lucia Ospina Martinez; Nicolas D. Franco-Sierra; Sergio Gomez-Rangel; Sussy Echeverria; Zulma M. Cucunubá |
| EPI_ISL_498165, EPI_ISL_526949, EPI_ISL_526950, EPI_ISL_526955, EPI_ISL_526958, EPI_ISL_526971, EPI_ISL_653746, EPI_ISL_653754, EPI_ISL_653756, EPI_ISL_653757, EPI_ISL_739672, EPI_ISL_739673 | | | |
| see above | Instituto Nacional de Salud, Bogotá, Colombia | Instituto Nacional de Salud, Bogotá, Colombia | Astrid C. Flórez; Carlos Andrés Durán; Carlos Franco-Muñoz; Carolina Ferro; Christian Julian Villabona-Arenas; Diana Marcela Walteros-Acero; Diego A. Álvarez-Díaz; Diego Andrés Prada; Franklin Prieto; Jonathan Reales; Jose A. Usme-Ciro; Katherine Laiton-Donato; Liz Villabona-Arenas; Magdalena Weisner; Marcela Mercado-Reyes; Martha Lucia Ospina Martinez; Mauricio Pacheco-Montealegre; Nicolas D. Franco-Sierra; Sheryl Corchuelo; Sussy Echeverria; Zulma M. Cucunubá |
| EPI_ISL_511298 | Instituto Nacional de Saude (INSA) and Instituto Gulbenkian de Ciencia (IGC) | Instituto Nacional de Saude (INSA) and Instituto Gulbenkian de Ciencia (IGC) | Borges et al |
| EPI_ISL_426580 | Instituto Sabin | Laboratory of Virology | Bergmann M Ribeiro; Fernando L Melo; Gustavo Barra; Ikaro A Andrade; Pedro G Mesquita; Tatsuya Nagata; Ticiane H Santa-Rita |
| EPI_ISL_492032, EPI_ISL_492033, EPI_ISL_492034, EPI_ISL_492035, EPI_ISL_492036, EPI_ISL_492037, EPI_ISL_492038, EPI_ISL_492039, EPI_ISL_492040, EPI_ISL_492041, EPI_ISL_492042, EPI_ISL_492043, EPI_ISL_492044, EPI_ISL_492045, EPI_ISL_492046, EPI_ISL_492047, EPI_ISL_492048 | | | |
| see above | Instituto de Biologia do Exército | Laboratório Metabolismo Macromolecular FirminoTorres de Castro, Instituto de Biofísica Carlos Chagas Filho, Universidade Federal do Rio de Janeiro | Aline Rosa Vianna de Souza; Bianca Catarina Azevedo Cabral; Caleb GM Santos; Clarissa Damaso; Elizabeth Valentin; Marcio da Costa Cipitelli; Marcos Dornelas-Ribeiro; Nádia Vaez Gonçalves da Cruz; Rodrigo Soares de Moura Neto; Rosane Silva; Tatiana LS Nogueira; Virginia Sara Grancieri do Amaral |
| EPI_ISL_455438, EPI_ISL_493334, EPI_ISL_493335, EPI_ISL_493336, EPI_ISL_576276, EPI_ISL_658865, EPI_ISL_658901, EPI_ISL_658904 | | | |
| see above | Instituto de Diagnostico y Referencia Epidemiologicos (INDRE) | Instituto de Diagnostico y Referencia Epidemiologicos (INDRE) | Abril Rodriguez-Maldonado; Adnan Araiza-Rodriguez; Araiza-Rodriguez Adnan; Barrera-Badillo Gisela; Claudia Wong-Arambula; Dayanira Arellano-Suarez; Edgar Mendieta-Condado; Ernesto Ramirez-Gonzalez; Ernesto Ramirez-Gonzalez.; Fabiola Garces-Ayala; Garces-Ayala Fabiola. Ramirez-Gonzalez Ernesto; Gaudalupe Herrera-Ramirez; Gisela Barrera-Badillo; Gisela Barrera-Badillo.; Hernandez-Rivas Lucia; Irma Lopez-Martinez; Jazmin Galicia-Hernandez.; Linda Andarade-Sanchez; Lopez-Martinez Irma; Lucia Hernandez-Rivas; Marisol Galindo-Galindo; Mendieta-Condado Edgar; Natividad Cruz-Ortiz; Octavio Ruiz-Muñiz; Pamela Ramirez-Medina; Rodriguez-Maldonado Abril; Roman Canul-Aguilar; Taboada Ramírez Blanca.; Tatiana Nunez-Garcia; Wong-Arambula Claudia |
| EPI_ISL_452139 | Instituto de Diagnostico y Referencia Epidemiologicos (INDRE) | Instituto de diagnóstico y Referencia Epidemiologicos (INDRE) | Araiza-Rodriguez Adnan; Barrera-Badillo Gisela; Garces-Ayala Fabiola; Hernandez-Rivas Lucia; Lopez-Martinez Irma; Mendieta-Condado Edgar; Ramirez-Gonzalez Ernesto; Rodriguez-Maldonado Abril; Wong-Arambula Claudia |
| EPI_ISL_747615, EPI_ISL_748139 | Instituto de Investigaciones Biológicas Clemente Estable | Institut Pasteur de Montevideo | Ana Carolina Mendonça; Andrés Lizasoain; Camila Simoes; Cecilia Alonso; Cecilia Salazar; Daiana Mir; Fernando López-Tort; Fernando Motta; Gonzalo Bello; Ighor Arantes; Ignacio Ferrés; Jose Sotelo; Leticia Maya; Letícia Garay Martins; Luciana Appolinario; Lucía Spangenberg; Mailen Arleo; Mariana Brandes; Marilda Mendonça Siqueira; Marilda Tereza Mar da Rosa; María José Benitez-Galeano; Martín Graña; Matías Castells; Matías Victoria; Matías Salvo; Natalia Rego; Natalia Reyes; Pablo Smircich; Paola Cristina Resende; Rodney Colina; Tamara Fernandez-Calero; Tania Possi; Tatiana Schäffer Gregianini; Verónica Noya; Yasser Vega |
| EPI_ISL_792322, EPI_ISL_792330 | Instituto de Investigaciones Biomédicas en Retrovirus y SIDA | Área de Secuenciación del Laboratorio de Virología del Hospital de Niños Dr. Ricardo Gutierrez on behalf of 'Proyecto Argentino Interinstitucional de genomica de SARS-CoV-2' (PAIS Consortium) | F; Goya; H; LE; Lusso; MI; MS; Nabaes Jodar; Natale; Remes Lenicov; S; Salomón; Seery; V; Valinotto; Viegas, M. |
| EPI_ISL_729870, EPI_ISL_729903, EPI_ISL_729908, EPI_ISL_729910, EPI_ISL_729914 | Instituto de Medicina Tropical, Universidad Nacional Toribio Rodríguez de Mendoza de Amazonas | Laboratorio de Genómica Microbiana, Universidad Peruana Cayetano Heredia | Alejandra Dávila-Barclay; Brenda Ayzanoa; Carla Montenegro; Cecilia Pajuelo; Janet Huancachoque; Luis González; Pablo Tsukayama; Pedro E. Romero; Pool Marcos; Rafael Tapia; Stella Chenet |
| EPI_ISL_474978, EPI_ISL_475017, EPI_ISL_514292, EPI_ISL_516887, EPI_ISL_516893, EPI_ISL_516902, EPI_ISL_516915, EPI_ISL_516917, EPI_ISL_575333, EPI_ISL_649064, EPI_ISL_649104, EPI_ISL_745034, EPI_ISL_745041, EPI_ISL_745068, EPI_ISL_745083, EPI_ISL_776636 | | | |
| see above | Israel Central Virology laboratory | Israel Central Virology laboratory | Efrat Dahan Bucris; Ella Mendelson; Michal Mandelboim; Neta Zuckerman; Oran Erster; Orna Mor |
| EPI_ISL_594158, EPI_ISL_594159 | Israel Institute for Biological Research | Israel Institute for Biological Research | Adi Beth-Din and Anat Zvi; Dana Stein; Emanuelle Mamroud; Galia Zaide; Inbar Cohen-Gihon; Libby Weiss; Ofir Israeli; Orly Laskar; Shay Weiss; Yoav Gal |
| EPI_ISL_736800, EPI_ISL_776965, EPI_ISL_776997, EPI_ISL_778683, EPI_ISL_778707, EPI_ISL_778729, EPI_ISL_778817 | | | |
| see above | Istituto Zooprofilattico Sperimentale del Mezzogiorno | TIGEM | Andrea Ballabio; Anna Manfredi; Antonio Grimaldi; Antonio Limone; Biancamaria Pierri; Chiara Colantuono; Davide Cacchiarelli.; Denise Di Concilio; Francesco Panariello; Lucio Di Filippo; Marcello Salvi; Maria Concetta Cuomo; Patrizia Annunziata; Pellegrino Cerino; Valentina Bouche |
| EPI_ISL_763069 | Istituto Zooprofilattico Sperimentale dell' Umbria e delle Marche -Togo Rosati | Istituto Superiore di Sanità | Gabriele Vaccari; Giovanni Ianiro; Ilaria Di Bartolo; Luca De Sabato; Massimo Biagetti; Monica Giammarioli |
| EPI_ISL_649191, EPI_ISL_722893 | Istituto Zooprofilattico Sperimentale della Puglia e della Basilicata | Istituto Zooprofilattico Sperimentale della Puglia e della Basilicata | Bianco A.; Capozzi L.; Cipolletta D.; Del Sambro L.; Galante D.; Manzulli V; Pace L.; Parisi A.; Rondinone V. |
| EPI_ISL_450793, EPI_ISL_450795 | Jamaica Ministry of Health and Wellness | Pathogen Discovery, Respiratory Viruses Branch, Division of Viral Diseases, Centers for Disease Control and Prevention | Anna Montmayeur; Anna Uehara; Bettina Bankamp; Clinton R. Paden; Haibin Wang; Jing Zhang; Krista Queen; Rachel Marine; Suxiang Tong; Yan Li; Ying Tao; Zachary Weiner |
| EPI_ISL_779256, EPI_ISL_779257, EPI_ISL_779265, EPI_ISL_779280, EPI_ISL_779281, EPI_ISL_779284, EPI_ISL_779289 | | | |
| see above | Jamil-ur-Rahman Center for Genome Research, Dr. Panjwani Center for Molecular Medicine and Drug Research | Jamil-ur-Rahman Center for Genome Research, Dr. Panjwani Center for Molecular Medicine and Drug Research | Ansari, S.; Irfan, M.; Khan, I.; Nisa, Z.; Rashid, M.; Shakeel, M. |
| EPI_ISL_451958 | Jamil-ur-Rahman Center for Genome Research, Dr. Panjwani Center for Molecular Medicine and Drug Research, International Center for Chemical and Biological Sciences, University of Karachi | Jamil-ur-Rahman Center for Genome Research, Dr. Panjwani Center for Molecular Medicine and Drug Research, International Center for Chemical and Biological Sciences, University of Karachi | B.A.; I.A; Khan; Khan, S.; M.A.and Khan; Qureshi; Raza; S.A.; Shakeel, M.; Zahid, M. |
| EPI_ISL_421237, EPI_ISL_421242, EPI_ISL_421243, EPI_ISL_421244, EPI_ISL_421246, EPI_ISL_421248, EPI_ISL_421250, EPI_ISL_421252, EPI_ISL_421253, EPI_ISL_421258, EPI_ISL_421259, EPI_ISL_421260, EPI_ISL_421262, EPI_ISL_455461, EPI_ISL_455462, EPI_ISL_455463, EPI_ISL_455466, EPI_ISL_455467 | | | |
| see above | Jiangxi Province Center for Disease Control and Prevention | Jiangxi Province Center for Disease Control and Prevention | DaJin Xiao; Fang Xiao; Gang Xu; JianXiong Li; Jun Zhou; ShiWen Liu; Tian Gong; XiaoQing Liu; Xin Ran; YanNi Zhang; Ying Xiong; Yong Shi |
| EPI_ISL_417420 | Jiangxi province Center for Disease Control and Prevention | Jiangxi province Center for Disease Control and Prevention | Li jian Xiong |
| EPI_ISL_412459 | Jingzhou Center for Disease Control and Prevention | Hubei Provincial Center for Disease Control and Prevention | Bin Fang; Bo Yang; Bo Yu; Chunlin Mao; Faxian Zhan; Guojun Ye; Jie Hu; Jing Li; Junqiang Xu; Kun Cai; Linlin Liu; Maoyi Chen; Xiang Li; Xiao Yu; Xixiang Huo; Yongzhong Jiang. |
| EPI_ISL_457875, EPI_ISL_457897, EPI_ISL_457906, EPI_ISL_457913, EPI_ISL_457928 | KEMRI-CGMR-C | KEMRI-Wellcome Trust Research Programme/KEMRI-CGMR-C Kilifi | Githinji G. et al 2020 |
| EPI_ISL_568725, EPI_ISL_568764, EPI_ISL_568847, EPI_ISL_568872 | KEMRI-Wellcome Trust Research Programme/KEMRI-CGMR-C Kilifi | KEMRI-Wellcome Trust Research Programme/KEMRI-CGMR-C Kilifi | Githinji et al 2020 |
| EPI_ISL_660233 | KRISP, KZN Research Innovation and Sequencing Platform | KRISP, KZN Research Innovation and Sequencing Platform | Giandhari J; Khan S; Lessells R; Mdlalose K; Pillay S; Tegally H; Wilkinson E; York D; de Oliveira T |
| EPI_ISL_407976, EPI_ISL_417427, EPI_ISL_418981 | KU Leuven, Clinical and Epidemiological Virology | KU Leuven, Clinical and Epidemiological Virology | Annabel Rector; Bert Vanmechelen; Elke Wollants; Els Keyaerts; Joan Marti-Carerras; Joan Marti-Carreras; Lies Laenen; Marc Van Ranst; Piet Maes; Tony Wawina; and Piet Maes |
| EPI_ISL_462177 | KU Leuven, Rega Institute, Clinical and Epidemiological Virology | KU Leuven, Rega Institute, Clinical and Epidemiological Virology | Bert Vanmechelen; Joan Marti-Carerras; Piet Maes; Tony Wawina-Bokalanga |
| EPI_ISL_539308 | KWR Watercycle Research Institute | Erasmus Medical Center | Bas B. Oude Munnink; Claudia M. E. Schapendonk; David Nieuwenhuijse; Frank M. Aarestrup; Gertjan Medema; Goffe Elsinga; Leo Heijnen; Lu Lu; Marion P.G. Koopmans; Matthijs Kon; Miranda de Graaf; Ray Izquierdo-Lara; Samantha Lycett |
| EPI_ISL_495413, EPI_ISL_495433, EPI_ISL_495454 | Kafkas University, Faculty of Medicine, Department of Medical Microbiology | Kafkas University, Faculty of Medicine, Department of Medical Microbiology | Didem Ozgur; E. Ediz Tutuncu; Murat Karamese |
| EPI_ISL_454447, EPI_ISL_454477 | Karolinska Universitetslaboratoriet | The Public Health Agency of Sweden | Anna Risberg; Anna-Malin Linde; Karin Tegmark-Wisell; Maria Lind Karlberg; Mattias Haukland; Olov Svartstrom; Oskar Karlsson Lindsjo; Petra Edquist; Reza Advani; Shamam Muradrasoli |
| EPI_ISL_560984 | Karolinska universitetslaboratoriet SOLNA | The Public Health Agency of Sweden | Anna Risberg; Anna-Malin Linde; Karin Tegmark-Wisell; Maria Lind Karlberg; Mattias Haukland; Mia Brytting; Olov Svartstrom; Oskar Karlsson Lindsjo; Petra Edquist; Reza Advani; Sandra Broddesson |
| EPI_ISL_469288, EPI_ISL_469292, EPI_ISL_469295, EPI_ISL_469296 | Keio University Hospital | Keio University Hospital | Kenjiro Kosaki |

| | | | |
|---|---|---|---|
| EPI_ISL_512812, EPI_ISL_512815, EPI_ISL_512818, EPI_ISL_512821 | Kenema Government Hospital, Ministry of Health and Sanitation | Kenema Government Hospital, Ministry of Health and Sanitation | Andersen, K.; Garry, R.; Goba, A.; Grant, D.; Happi, C.; Jalloh, S.; Mehta, S.; Momoh, M.; Olawoye, I.; Oluniyi, P.; Park, D.; Sandi, J.; Siddle, K.; Tomkins-Tinch, C. |
| EPI_ISL_489997, EPI_ISL_489999, EPI_ISL_490006, EPI_ISL_490008, EPI_ISL_490010 | King Fahad Medical City | King Fahad Medical City | Alghoraibi, M.; Alosaimi, B.; Enani, M.; Naeem, A. |
| EPI_ISL_483543, EPI_ISL_483547, EPI_ISL_483549, EPI_ISL_483553, EPI_ISL_483562, EPI_ISL_483564 | Kingdom of Bahrein Ministry of Health | Erasmus Medical Center | Amjad Ghanem Mohamed; Anne van der Linden; Bas Oude Munnink; Claudia Schapendonk; David Nieuwenhuijse; Ebrahim Shehad; Fatema; Hashmeya Al Wasti; Irina Chestakova; Marion Koopmans; Mark Pronk; Pascal Lexmond; Reina Sikkema; Richard Molenkamp; Stefan van Nieuwkoop; Theo Bestebroer; on behalf of the Dutch national COVID-19 response team. |
| EPI_ISL_676501 | Klinisk Mikrobiologi | The Public Health Agency of Sweden | Department of Microbiology; The Public Health Agency of Sweden |
| EPI_ISL_654942, EPI_ISL_661289, EPI_ISL_661302, EPI_ISL_766730, EPI_ISL_789047 | Klinisk mikrobiologi | The Public Health Agency of Sweden | Anna Risberg; Anna-Malin Linde; Department of Microbiology; Karin Tegmark-Wisell; Maria Lind Karlberg; Mattias Haukland; Mia Brytting; Olov Svartstrom; Oskar Karlsson Lindsjo; Petra Edquist; Reza Advani; Sandra Broddesson; The Public Health Agency of Sweden |
| EPI_ISL_615097 | Klinisk mikrobiologi Lanssjukhuset Ryhov, Jonkoping | The Public Health Agency of Sweden | Anna Risberg; Anna-Malin Linde; Karin Tegmark-Wisell; Maria Lind Karlberg; Mattias Haukland; Mia Brytting; Olov Svartstrom; Oskar Karlsson Lindsjo; Petra Edquist; Reza Advani; Sandra Broddesson |
| EPI_ISL_510868 | Klinisk mikrobiologi NAL Trollhattan | The Public Health Agency of Sweden | Anna Risberg; Anna-Malin Linde; Karin Tegmark-Wisell; Maria Lind Karlberg; Mattias Haukland; Mia Brytting; Olov Svartstrom; Oskar Karlsson Lindsjo; Petra Edquist; Reza Advani; Sandra Broddesson |
| EPI_ISL_510853, EPI_ISL_582836, EPI_ISL_623090 | Klinisk mikrobiologi Vasternorrland | The Public Health Agency of Sweden | Anna Risberg; Anna-Malin Linde; Karin Tegmark-Wisell; Maria Lind Karlberg; Mattias Haukland; Mia Brytting; Olov Svartstrom; Oskar Karlsson Lindsjo; Petra Edquist; Reza Advani; Sandra Broddesson |
| EPI_ISL_775545 | Klinisk mikrobiologi, Viruslab | The Public Health Agency of Sweden | Department of Microbiology; The Public Health Agency of Sweden |
| EPI_ISL_594149 | Klinsisk mikrobiologi Linkoping | The Public Health Agency of Sweden | Anna Risberg; Anna-Malin Linde; Karin Tegmark-Wisell; Maria Lind Karlberg; Mattias Haukland; Mia Brytting; Olov Svartstrom; Oskar Karlsson Lindsjo; Petra Edquist; Reza Advani; Sandra Broddesson |
| EPI_ISL_407193 | Korea Centers for Disease Control & Prevention (KCDC) Center for Laboratory Control of Infectious Diseases Division of Viral Diseases | Korea Centers for Disease Control & Prevention (KCDC) Center for Laboratory Control of Infectious Diseases Division of Viral Diseases | Heui Man Kim; Hye-Joon Jo; Jeong-Min Kim; Mi-Seon Kim; Myung Guk Han; Namjoo Lee; SangHee Woo; Sehee Park; Yoon-Seok Chung |
| EPI_ISL_515181 | Kumasi Centre for Collaborative Research in Tropical Medicine, Kumasi. | Institute of Virology, Charité – Universitätsmedizin Berlin | Augustina Sylverken; Christian Drosten; Eric Adu; Jesse Addo Asamoah; Julia Schneider; Jörn Beheim-Schwarzbach; Michael Owusu; Philip El-Duah; Richard Phillips.; Richmond Gorman; Richmond Yeboah; Sherihane Aryeetey; Victor Max Corman |
| EPI_ISL_682277 | LABORATORIOS LABIN | Inciensa, Instituto Costarricense de Investigación y Enseñanza en Nutrición y Salud | Adriana Godinez; Claudio Soto-Garita; Estela Cordero; Francisco Duarte; Hebleen Porras; Melany Calderon & Pei Ling Chan |
| EPI_ISL_717809, EPI_ISL_717832, EPI_ISL_717833, EPI_ISL_717834, EPI_ISL_717835, EPI_ISL_717836, EPI_ISL_717841, EPI_ISL_717910, EPI_ISL_717911, EPI_ISL_717912, EPI_ISL_717913, EPI_ISL_717914, EPI_ISL_717915, EPI_ISL_717916, EPI_ISL_717917, EPI_ISL_717918, EPI_ISL_717919, EPI_ISL_717958, EPI_ISL_717963, EPI_ISL_717964 | | | |
| see above | LACEN Dr. Francisco Rimolo Neto | Bioinformatics Laboratory / LNCC | Alexandra L Gerber; Amilcar Tanuri; Ana Paula de C Guimarães; Ana Tereza R de Vasconcelos; Andréa Cony Cavalcanti; Carolina M Voloch; Claudia dos Santos Rodrigues; Cynthia C Cardoso; Diana Mariani; Luiz G P de Almeida; Otavio Bustrolini; Ronaldo da Silva F Jr; Terezinha M P P Castiñeira |
| EPI_ISL_717899, EPI_ISL_717900, EPI_ISL_717901, EPI_ISL_717902, EPI_ISL_717903, EPI_ISL_717904, EPI_ISL_717905, EPI_ISL_717906, EPI_ISL_717907, EPI_ISL_717908, EPI_ISL_717909, EPI_ISL_717962 | | | |
| see above | LACEN RJ - Noel Nutels | Bioinformatics Laboratory / LNCC | Alexandra L Gerber; Amilcar Tanuri; Ana Paula de C Guimarães; Ana Tereza R de Vasconcelos; Andréa Cony Cavalcanti; Carolina M Voloch; Claudia dos Santos Rodrigues; Cynthia C Cardoso; Diana Mariani; Luiz G P de Almeida; Otavio Bustrolini; Ronaldo da Silva F Jr; Terezinha M P P Castiñeira |
| EPI_ISL_502779, EPI_ISL_502875 | LACEN/PE | LABBE, Federal University of Pernambuco | ANTONIO CARLOS DE FREITAS; BRUNO SAMPAIO; HEIDI LACERDA ALVES DA CRUZ; MAIRA GALDINO DA ROCHA PITTA; MARCOS ANTONIO DE MORAIS JUNIOR; MARCOS DA SILVEIRA REGUEIRA NETO; MICHELLY CRISTINY PEREIRA; REGINALDO GONCALVES DE LIMA NETO; SERGIO DE SA LEITAO PAIVA JUNIOR; VALDIR DE QUEIROZ BALBINO.; WILSON JOSE DA SILVA JUNIOR; ZILDENE DE SOUSA SILVEIRA |
| EPI_ISL_469049 | LNR National Reference Laboratory, Mohammed VI University of Health Sciences | Medical Biotechnology Laboratory, Rabat Medical and Pharmacy School, Mohammed The Vth University in Rabat | Chakib NEJJARI; Houda BENRAHMA; Idrissa Diawara; Imane SMYEJ; Jalil El Atar; Jalila RAHOUI; Lahcen BELYAMANI and Azeddine IBRAHIMI; Laila SBABOU; Loubna ALLAM; M.W. CHEMAO-ELFIHRI; Meriem LAAMARTI; Mouna OUADGHIRI; Rachid EL JAOUDI; Rachid MENTAG; Rokaia LAAMRTI; Saaid AMZAZI; Souad KARTTI |
| EPI_ISL_578968, EPI_ISL_653625, EPI_ISL_653654, EPI_ISL_768407 | LSUHS Emerging Viral Threat Laboratory | Microbial Genome Sequencing Center | Andrew D. Yurochko; Camille F. Abshire; Christopher G. Kevil; Daniel J. Snyder; Jennifer L. Carroll; Jeremy P. Kamil; John A. Vanchiere; Katarzyna Zwolinska; Maarten Van Diest; Malgorzata Bienkowska-Haba; Martin J. Sapp; Rona S. Scott; Vaughn S. Cooper |
| EPI_ISL_528637, EPI_ISL_528638 | LVM/UFRJ | Bioinformatics Laboratory / LNCC | Amilcar Tanuri; Ana Teresa R. Vasconcelos; Bruno B. Bezerra; Diana Marianni; Elena Cobos; Fabio Limonte; Gustavo D. P. Silva; Isadora A. Correa; Luciana B. Arruda; Luciana J. Costa; Lucio A. Caldas; Luiz Almeida; Luiza Higga; M. Romário M. de Souza; Marcelo Bozza; Orlando Ferreira; Sharton V. A. Coelho; Terezinha M. Castineiras; Wanderley de Souza |
| EPI_ISL_528539 | LVM/UFRJ | LNCC | Amilcar Tanuri; Ana Teresa R. Vasconcelos; Bruno B. Bezerra; Diana Marianni; Elena Cobos; Fabio Limonte; Gustavo M. Romário M. de Souza; Isadora A. Correa; Luciana B. Arruda; Luciana J. Costa.; Lucio A. Caldas; Luiz Almeida; Luiza Higga; Marcelo Bozza; Orlando Ferreira; Sharton V. A. Coelho; Terezinha M. Castineiras; Wanderley de Souza |
| EPI_ISL_755644, EPI_ISL_755645, EPI_ISL_755652 | Lab LOC - Itapecerica da Serra | Instituto Adolfo Lutz, Interdiciplinary Procedures Center, Strategic Laboratory | Claudia Regina Gonçalves; Claudio Tavares Sacchi; Erica Valessa Ramos Gomes; Karoline Rodrigues Campos |
| EPI_ISL_634878, EPI_ISL_717613, EPI_ISL_718000, EPI_ISL_718020 | Lab voor klinische biologie | Onderzoeksgroep Virologie | Bruno Verhasselt; Hans Nauwynck; Laurens Lambrechts; Linos Vandekerckhove; Marthe Pauwels; Nick Vereecke; Sebastiaan Theuns |
| EPI_ISL_456176, EPI_ISL_456382, EPI_ISL_456394, EPI_ISL_548120, EPI_ISL_579111, EPI_ISL_579425, EPI_ISL_622803, EPI_ISL_732965, EPI_ISL_755630, EPI_ISL_755631 | | | |
| see above | LabPLUS | Institute of Environmental Science and Research (ESR) | Anja Werno; Antje van der Linden; Arlo Upton; Chris Mansell; David Hammer; Dragana Drinkovic; Erasmus Smit; Gary McAuliffe; Hana Sofia Andersson; Hermes Perez; James Ussher; Jill Sherwood; Jing Wang; Joep de Ligt; Josh Freeman; Julia Howard; Juliet Elvy; Lauren Jelly; Mary DeAlmeida; Matt Blakiston; Matt Storey; Matthew Rogers; Max Bloomfield; Michael Addidle; Michelle Balm; Muhammad Faisal; Nikki Freed; Olin Silander; Sally Roberts; Sarah Jefferies; Sharmini Muttaiyah; Susan Morpeth; Susan Taylor; Timothy Blackmore; Vani Sathyendran; Veronica Playle; Virginia Hope; Xiaoyun Ren |
| EPI_ISL_547994, EPI_ISL_579424 | LabTests | Institute of Environmental Science and Research (ESR) | Anja Werno; Antje van der Linden; Arlo Upton; Chris Mansell; David Hammer; Dragana Drinkovic; Erasmus Smit; Gary McAuliffe; Hana Sofia Andersson; Hermes Perez; James Ussher; Jill Sherwood; Jing Wang; Joep de Ligt; Josh Freeman; Julia Howard; Juliet Elvy; Lauren Jelly; Mary DeAlmeida; Matt Blakiston; Matt Storey; Matthew Rogers; Max Bloomfield; Michael Addidle; Michelle Balm; Muhammad Faisal; Nikki Freed; Olin Silander; Sally Roberts; Sarah Jefferies; Sharmini Muttaiyah; Susan Morpeth; Susan Taylor; Timothy Blackmore; Vani Sathyendran; Veronica Playle; Virginia Hope; Xiaoyun Ren |
| EPI_ISL_560638 | Labo Analyses Med | National Reference Center for Viruses of Respiratory Infections, Institut Pasteur, Paris | Etienne Simon-Lorière; Fabiana Gambaro; Maud Vanpeene; Sylvie Behillil; Sylvie van der Werf; Vincent Enouf |
| EPI_ISL_416029 | Laboiratório Fleury | Instituto Adolfo Lutz, Interdiciplinary Procedures Center, Strategic Laboratory | Adriana Bugno; Adriano Abbud; Carlos Henrique Camargo; Claudia Regina Gonçalves; Claudio Tavares Sacchi; Daniela Bernardes Borges da Silva; Fabiana Cristina Pereira dos Santos; Maria do Carmo Sampaio Tavares Timenetsky; Simone Guadagnucci Morillo; Terezinha Maria de Paiva |
| EPI_ISL_445220 | Laborarory for Respiratory Viruses, "Cantacuzino" National Military-Medical Institute for Resararch and Development | Cantacuzino Institute | A.Cretu; L.Ustea; M.Lazar |
| EPI_ISL_769863, EPI_ISL_775256 | Laboratoire Biolife | Laboratoire de Biotechnologie | Afaf Alaoui; Amina Benouda; Bouchra Belfquih; Hanae Dakka; Lahcen belyamani; Mohamed Chenaoui; Mohammed Walid Chemao Elfihri; Mouna Ouadghiri; Otmane Touzani; Saaid Amzazi and Azeddine Ibrahimi; Tarik Aanniz |
| EPI_ISL_428956, EPI_ISL_429733, EPI_ISL_429784, EPI_ISL_434507, EPI_ISL_445065, EPI_ISL_445066, EPI_ISL_459896, EPI_ISL_459905 | | | |
| see above | Laboratoire National de Sante, Microbiology, Virology | Laboratoire National de Sante, Microbiology, Epidemiology and Microbial Genomics | Anke Wienecke-Baldacchino; Ardashel Latsuzbaia; Catherine Ragimbeau; Guillaume Fournier; Jessica Tapp; Joel Mossong; Tamir Abdelrahman; Trung Nguyen Nguyen |
| EPI_ISL_413593 | Laboratoire National de Santé | Erasmus Medical Center | Anne van der Linden; Annemiek van der Eijk; Bas Oude Munnink; Claudia Schapendonk; Corine GeurtsvanKessel; David Nieuwenhuijse; G. Fournier; Irina Chestakova; J. Mossong; Jeroen van Kampen; Jolanda Voermans; Marion Koopmans; Mark Pronk; Pascal Lexmond; Reina Sikkema; Richard Molenkamp; T. Abdelrahman; T. Nguyen; on behalf of the Dutch national COVID-19 response team. |
| EPI_ISL_419566, EPI_ISL_419604 | Laboratoire National de Santé, Microbiology, Virology | Laboratoire National de Santé, Microbiology, Epidemiology and Microbial Genomics | Anke Wienecke-Baldacchino; Ardashel Latsuzbaia; Catherine Ragimbeau; Guillaume Fournier; Jessica Tapp; Joel Mossong; Tamir Abdelrahman; Trung Nguyen Nguyen |
| EPI_ISL_754147, EPI_ISL_754158 | Laboratoire de Microbiologie | National Reference Center for Viruses of Respiratory Infections, Institut Pasteur, Paris | Angela Brisebarre; Camille Capel; Etienne Simon-Lorière; Marie-Sarah Fangous; Marion Barbet; Maud Vanpeene; Méline Bizard; Sylvie Behillil; Sylvie van der Werf; Vincent Enouf |
| EPI_ISL_660446, EPI_ISL_660450, EPI_ISL_660452, EPI_ISL_660454, EPI_ISL_660457, EPI_ISL_660465, EPI_ISL_660472, EPI_ISL_660473, EPI_ISL_660475, EPI_ISL_660479, EPI_ISL_660480, EPI_ISL_660488, EPI_ISL_660489, EPI_ISL_660492, EPI_ISL_660494 | | | |
| see above | Laboratoire de Microbiologie CHU Souro Sanou | Centre Muraz | Abdoul-Salam Ouedraogo; Arsène Zongo; Essia Belarbi; Fabian Leendertz; Grit Schubert; Halidou Tinto; Lassana Sangaré; Soumeya Ouangraoua; Yacouba Sawadogo; Zekiba Tarnagda |
| EPI_ISL_712064, EPI_ISL_712568 | Laboratoire de Microbiologie- CHU Habib Bourguiba – Sfax adresse | Laboratoire des Procédés de Criblage Moléculaire et Cellulaire-Centre de Biotechnologie de Sfax | A. and Masmoudi, S.; Abdelmoulah, F.; Abid, N.; Ajili, F.; Aouni, M.; Ben Ayed, I.; Bensaid, M.; Chtourou, A.; Elargoubi, A.; Fki-berrajah, L.; Gaaloul, I.; Gargouri, S.; Hammami, A.; Kamoun, S.; Karray Hakim, H.; Kharat, N.; Mastouri, M.; Mhalla, S.; Nabli, A.; Rebai; Smeti, I.; Souissi, A.; Stambouli, N.; Turki, M. |

| | | | |
|---|---|---|---|
| EPI_ISL_414623 | Laboratoire de Virologie Institut de Virologie - INSERM U 1109 Hôpitaux Universitaires de Strasbourg | National Reference Center for Viruses of Respiratory Infections, Institut Pasteur, Paris | Angela Brisebarre; Flora Donati Vincent Enouf; Marion Barbet; Maud Vanpeene; Méline Bizard; Mélnie Albert; Samira Fafi-Kremer; Sylvie Behillil; Sylvie van der Werf |
| EPI_ISL_413996 | Laboratoire de Virologie, HUG | Swiss National Reference Centre for Influenza | LAUBSCHER Florian et al. |
| EPI_ISL_444500 | Laboratoire de microbiologie, Hopital de Verdun | Smith Laboratory, Centre de Recherche CHU Sainte-Justine | Ivan Pavlov; Marieke Rozendaal; Martin Smith |
| EPI_ISL_779823 | Laboratoire de virologie, CHU de Grenoble | CNR Virus des Infections Respiratoires - France SUD | Antonin Bal; Bruno Lina; Gregory Destras; Gwendolyne Burfin; Hadrien Règue; Laurence Josset; Martine Valette; Quentin Semanas; Sylvie Larrat |
| EPI_ISL_476822, EPI_ISL_476826, EPI_ISL_476828, EPI_ISL_476829, EPI_ISL_476831 | Laboratoire des Fièvres Hémorragiques Virales du Benin | Charité-Universitätsmedizin Berlin | Anges; Drexler; Jan Felix; Moreira-Soto Andres; Sander Anna-Lena; Yadouleton |
| EPI_ISL_744819 | Laboratoire national de santé, Microbiology, Virology | Laboratoire national de santé, Microbiology, Epidemiology and Microbial Genomics | Anke Wienecke-Baldacchino; Catherine Ragimbeau; Fatu Djabi; Jessica Tapp; Tamir Abdelrahman; Trung Nguyen Nguyen |
| EPI_ISL_739733, EPI_ISL_739925, EPI_ISL_739934, EPI_ISL_739954, EPI_ISL_740005, EPI_ISL_740063, EPI_ISL_740189, EPI_ISL_740289, EPI_ISL_740375, EPI_ISL_740438, EPI_ISL_740469, EPI_ISL_740489, EPI_ISL_744149, EPI_ISL_744246, EPI_ISL_744253, EPI_ISL_744260, EPI_ISL_744480, EPI_ISL_744575, EPI_ISL_744586, EPI_ISL_744633, EPI_ISL_744711, EPI_ISL_744766, EPI_ISL_744822, EPI_ISL_744931, EPI_ISL_744941, EPI_ISL_745013, EPI_ISL_745022, EPI_ISL_770925, EPI_ISL_771022, EPI_ISL_771113 | | | |
| see above | Laboratoire national de santé, Microbiology, Virology | Laboratoire national de santé, Microbiology, Microbial Genomics Platform | Anke Wienecke-Baldacchino; Catherine Ragimbeau; Fatu Djabi; Jessica Tapp; Lise Pignon; Raoul Salmon; Tamir Abdelrahman |
| EPI_ISL_792519, EPI_ISL_792521 | Laboratorio Central de la Ciudad de Santa Fe | Grupo de Genómica y Bioinformática del Instituto de Investigación de la Cadena Láctea CONICET-INTA on behalf of 'Proyecto Argentino Interinstitucional de genomica de SARS-CoV-2' (PAIS Consortium) | AF; Amadio; C; Eberhardt; G; Irazoqui; JM; MF; Mugna; Ojeda; Pastor; Rompato; V |
| EPI_ISL_792545 | Laboratorio Central, Ministerio de Salud Córdoba | Instituto de Patologia Vegetal (CIAP-INTA) on behalf of 'Proyecto Argentino Interinstitucional de genomica de SARS-CoV-2' (PAIS Consortium) | Barbas, G.; Castro, G.; Debat, HJ.; FD; Fernández; MB; Pisano; Re; V |
| EPI_ISL_424670 | Laboratorio Estatal de Salud Publica del Estado de Queretaro | Instituto de Diagnóstico y Referencia Epidemiologicos | Adnan Araiza Rodríguez; Alejandro Sánchez; Alfredo Ponce de León Garduño; Blanca Taboada; Carlos F. Arias; Carolina González Torres; Celia Boukadida; Cesar Raúl González Bonilla; Concepción Grajales Muñiz; Edgar Mendieta Condado; Eduardo Becerril Vargas; Fabiola Garcés Ayala; Fernando Ledesma Barrientos; Francisco Javier Gaytán Cervantes; Francisco Pulido; Gisela Barrera Badillo; Gloria Vázquez; Guillermo M. Ruiz-Palacios; Irma López Martínez; Joel Armando Vázquez Pérez; José Arturo Martínez Orozco; José Ernesto Ramírez González; José Esteban Muñoz Medina; Lucia Hernández Rivas; Luis Alberto García Andrade; Mario Mújica Sánchez; Pavel Isa; Pilar Ramos Cervantes; Ricardo Grande; Santiago Avila Ríos; Victor Hugo Borja Aburto; Violeta Ibarra Gonzalez |
| EPI_ISL_779189, EPI_ISL_779190, EPI_ISL_779191, EPI_ISL_779194, EPI_ISL_779195, EPI_ISL_779198 | Laboratorio Estatal de Salud Pública de Nuevo León | Laboratorio de Infectología Molecular, Departamento de Bioquímica y Medicina Molecular,Facultad de Medicina - Universidad Autónoma de Nuevo León | Ana M. Rivas-Estilla; Consuelo Treviño-Garza; Daniel Arellanos-Soto; Else del Carmen García-García; Gloria A. Jasso-de-la-Peña; Kame A. Galán-Huerta; Manuel E. de-la-O-Cavazos; María F. Herrera-Saldivar; Natalia Martínez-Acuña; Roberto Montes-de-Oca; Samuel Buentello-Wong; Sonia A. Lozano-Sepúlveda |
| EPI_ISL_735398 | Laboratorio Fleury | Instituto Adolfo Lutz, Interdiciplinary Procedures Center, Strategic Laboratory | Claudia Regina Gonçalves; Claudio Tavares Sacchi; Erica Valessa Ramos Gomes; Karoline Rodrigues Campos |
| EPI_ISL_693246 | Laboratorio Municipal de Rio Grande da Serra | Instituto Adolfo Lutz, Interdiciplinary Procedures Center, Strategic Laboratory | Claudia Regina Gonçalves; Claudio Tavares Sacchi; Erica Valessa Ramos Gomes; Karoline Rodrigues Campos |
| EPI_ISL_457948, EPI_ISL_457953, EPI_ISL_457957, EPI_ISL_457965, EPI_ISL_457972 | Laboratorio de Biología Molecular Asociación Española Primera en Salud | Departments of Pathology and Medicine, New York University School of Medicine | Adriana Heguy; Christian Marier; Gael Westby; Gonzalo Manrique; Maria Noel Zubillaga; Maria Victoria Elizondo; Matthew T Maurano; Paul Zappile |
| EPI_ISL_626564 | Laboratorio de Biología Molecular, Facultad de Medicina, Universidad de Atacama, Copiapo, Chile/ FONDAP CRG, Universidad Andrés Bello, Santiago, Chile | Center for Mathematical Modeling and Center for Genome Regulation. Santiago, Chile | Allende ML; Bastias M; Castro E; Echeverría C; González M; M; Maass A; Manríquez R; Meneses C.; Montecino; Orellana A; Sanhueza D; Travisany D |
| EPI_ISL_792355 | Laboratorio de Biología Molecular. Hospital Dr. Héctor Cura | Área de Secuenciación del Laboratorio de Virología del Hospital de Niños Dr. Ricardo Gutierrez on behalf of 'Proyecto Argentino Interinstitucional de genomica de SARS-CoV-2' (PAIS Consortium) | Ghiano; Goya; J; LE; Lusso; MB; MI; MS; N; Nabaes Jodar; Natale; R; S; Spina; Turrina; Valinotto; Viegas, M.; Zaffanella |
| EPI_ISL_417034, EPI_ISL_792560, EPI_ISL_801386, EPI_ISL_801387, EPI_ISL_801388, EPI_ISL_801389, EPI_ISL_801390, EPI_ISL_801391, EPI_ISL_801392, EPI_ISL_801393, EPI_ISL_801394, EPI_ISL_801395, EPI_ISL_801396 | | | |
| see above | Laboratorio de Ecologia de Doencas Transmissiveis na Amazonia, Instituto Leonidas e Maria Deane - Fiocruz Amazonia | Laboratorio de Ecologia de Doencas Transmissiveis na Amazonia, Instituto Leonidas e Maria Deane - Fiocruz Amazonia | André Corado; Debora Duarte; Felipe Naveca on behalf of the Fiocruz COVID-19 Genomic Surveillance Network; Fernanda Nascimento; George Silva; Karina Pessoa; Luciana Gonçalves; Maria Júlia Brandão; Michele Jesus; Sérgio Luz; Valdinete Nascimento; Victor Souza; Ágatha Costa |
| EPI_ISL_779170, EPI_ISL_779172 | Laboratorio de Infectología, Servicio de Infectología, Hospital Universitario Dr. José Eleuterio González - Universidad Autónoma de Nuevo León | Laboratorio de Infectología Molecular, Departamento de Bioquímica y Medicina Molecular,Facultad de Medicina - Universidad Autónoma de Nuevo León | Adrian Camacho-Ortiz; Ana M. Rivas-Estilla; Daniel Arellanos-Soto; Eduardo Perez-Alba; Elvira Garza-González; Kame A. Galán-Huerta; Laura Nuzzolo-Shihadeh; María F. Herrera-Saldivar; Natalia Martínez-Acuña; Paola Bocanegra-Ibarias; Samantha M. Flores-Treviño; Sonia A. Lozano-Sepúlveda |
| EPI_ISL_792442 | Laboratorio de Inmunología del Hospital Perrando e Instituto de Medicina Regional de la UNNE | Instituto de Biotecnología, IABIMO (CONICET), Instituto de Virología, IVIT(CONICET), Instituto de Patobiología, IPVET(CONICET), CICVyA, INTA on behalf of 'Proyecto Argentino Interinstitucional de genomica de SARS-CoV-2' (PAIS Consortium) | A; AF; AJ; AV; Asurmendi; Ayala; Bengoa Luoni; Cacciabué; Cayré; D; Deluca; Distéfano; Farber; Fass; Foussal; G; GA; Giusiano; Gómez; H; König; L; LC; Lescano; Lozano Calderón; Lucero; M; MD; MG; MPD; MV; Marin; Muñoz Hidalgo; NA; NB; PA; Paniego; Pedroarias; Peralta; Puebla; Rivarola; S; VC; Vera; Viegas, M.; Zavallo |
| EPI_ISL_648313, EPI_ISL_648315, EPI_ISL_648324, EPI_ISL_648325, EPI_ISL_648330, EPI_ISL_648339, EPI_ISL_648340, EPI_ISL_648347, EPI_ISL_648353, EPI_ISL_648355, EPI_ISL_648367, EPI_ISL_648379, EPI_ISL_649158, EPI_ISL_649164, EPI_ISL_649166, EPI_ISL_649170, EPI_ISL_649172 | | | |
| see above | Laboratorio de Investigaciones de Baney | University Hospital Basel, Clinical Bacteriology | Adrian Egli; Alfredo Mari; Bonifacio Manguire Nlavo; Carlos Cortes; Claudia Daubenberger; Diosdado Odjama Nseng Ada; Elizabeth Nyakarungu; Guillermo Garcia; Helena Seth-Smith; Madlen Stange; Maximilian Mpina; Mitoha Ondo O Ayekaba; Philip Wonder Phiri; Salome Hosch; Tim Roloff; Tobias Schindler |
| EPI_ISL_516722, EPI_ISL_517713, EPI_ISL_527787 | Laboratorio de Referencia Nacional de Virus Respiratorio. Centro Nacional de Salud Publica. Instituto Nacional de Salud Peru. | Laboratorio de Referencia Nacional de Biotecnología y Biología Molecular. Centro Nacional de Salud Publica. Instituto Nacional de Salud Peru. | Carlos Padilla Rojas; Henri Bailon Calderon; Johanna Balbuena Torres; Karolyn Vega Chozo; Marco Galarza Perez; Maribel Huaringa Nuñez; Nancy Rojas Serrano.; Omar Caceres Rey; Priscila Lope Pari |
| EPI_ISL_489987, EPI_ISL_491429, EPI_ISL_491433 | Laboratorio de Referencia Nacional de Virus Respiratorio. Instituto Nacional de Salud Perú | Laboratorio de Referencia Nacional de Biotecnología y Biología Molecular. Instituto Nacional de Salud Perú | Carlos Padilla Rojas; Henri Bailon Calderon; Johanna Balbuena Torres; Johanna Balbuena Torrez; Karolyn Chozo Vega; Karolyn Vega Chozo; Marco Galarza Perez; Maribel Huaringa Nuñez; Nancy Rojas Serrano; Nancy Rojas Serrano.; Omar Caceres Rey; Priscila Lope Pari |
| EPI_ISL_529067, EPI_ISL_540927, EPI_ISL_540945, EPI_ISL_568549, EPI_ISL_729884, EPI_ISL_729886 | Laboratorio de Referencia Nacional de Virus Respiratorios, Instituto Nacional de Salud Peru | Laboratorio de Genómica Microbiana, Universidad Peruana Cayetano Heredia | Alejandra Dávila-Barclay; Brenda Ayzanoa; Camila Castillo-Vilcahuaman; Guillermo Salvatierra; Janet Huancachoque; Luis González; Marco Galarza; Maribel Huaringa; Nancy Rojas; Pablo Tsukayama; Pedro E. Romero; Pool Marcos; Priscila Lope |
| EPI_ISL_792365 | Laboratorio de Virologia - HIEAyC San Juan de Dios | Área de Secuenciación del Laboratorio de Virología del Hospital de Niños Dr. Ricardo Gutierrez on behalf of 'Proyecto Argentino Interinstitucional de genomica de SARS-CoV-2' (PAIS Consortium) | A; Colmeiro; Ercole; Ferioli; Gatelli; Goya; LE; Lusso; M; MI; MS; Nabaes Jodar; Natale; R; S; Valinotto; Viegas, M. |
| EPI_ISL_623108, EPI_ISL_623110, EPI_ISL_623112, EPI_ISL_623114, EPI_ISL_623116, EPI_ISL_623118, EPI_ISL_623119, EPI_ISL_623120, EPI_ISL_623124, EPI_ISL_623126, EPI_ISL_623129, EPI_ISL_623130, EPI_ISL_623132, EPI_ISL_623134, EPI_ISL_623136, EPI_ISL_623138, EPI_ISL_623140, EPI_ISL_623142, EPI_ISL_623143, EPI_ISL_623144, EPI_ISL_623145, EPI_ISL_623146, EPI_ISL_623147, EPI_ISL_623148, EPI_ISL_623149, EPI_ISL_623152, EPI_ISL_623154, EPI_ISL_623157, EPI_ISL_623158, EPI_ISL_623160, EPI_ISL_623161, EPI_ISL_623162, EPI_ISL_623164, EPI_ISL_623165, EPI_ISL_623166, EPI_ISL_623168, EPI_ISL_717806, EPI_ISL_717807, EPI_ISL_717808, EPI_ISL_717810, EPI_ISL_717811, EPI_ISL_717812, EPI_ISL_717813, EPI_ISL_717814, EPI_ISL_717815, EPI_ISL_717816, EPI_ISL_717817, EPI_ISL_717818, EPI_ISL_717819, EPI_ISL_717820, EPI_ISL_717821, EPI_ISL_717822, EPI_ISL_717823, EPI_ISL_717824, EPI_ISL_717825, EPI_ISL_717826, EPI_ISL_717827, EPI_ISL_717828, EPI_ISL_717829, EPI_ISL_717830, EPI_ISL_717831, EPI_ISL_717837, EPI_ISL_717838, EPI_ISL_717839, EPI_ISL_717840, EPI_ISL_717842, EPI_ISL_717843, EPI_ISL_717844, EPI_ISL_717845, EPI_ISL_717846, EPI_ISL_717847, EPI_ISL_717848, EPI_ISL_717849, EPI_ISL_717850, EPI_ISL_717851, EPI_ISL_717852, EPI_ISL_717853, EPI_ISL_717854, EPI_ISL_717855, EPI_ISL_717857, EPI_ISL_717858, EPI_ISL_717859, EPI_ISL_717860, EPI_ISL_717861, EPI_ISL_717862, EPI_ISL_717863, EPI_ISL_717864, EPI_ISL_717865, EPI_ISL_717866, EPI_ISL_717867, EPI_ISL_717868, EPI_ISL_717869, EPI_ISL_717870, EPI_ISL_717871, EPI_ISL_717872, EPI_ISL_717873, EPI_ISL_717874, EPI_ISL_717875, EPI_ISL_717876, EPI_ISL_717877, EPI_ISL_717878, EPI_ISL_717879, EPI_ISL_717880, EPI_ISL_717881, EPI_ISL_717882, EPI_ISL_717883, EPI_ISL_717884, EPI_ISL_717885, EPI_ISL_717886, EPI_ISL_717887, EPI_ISL_717888, EPI_ISL_717889, EPI_ISL_717890, EPI_ISL_717891, EPI_ISL_717892, EPI_ISL_717893, EPI_ISL_717894, EPI_ISL_717895, EPI_ISL_717896, EPI_ISL_717897, EPI_ISL_717898, EPI_ISL_717920, EPI_ISL_717921, EPI_ISL_717922, EPI_ISL_717923, EPI_ISL_717924, EPI_ISL_717925, EPI_ISL_717926, EPI_ISL_717927, EPI_ISL_717928, EPI_ISL_717929, EPI_ISL_717930, EPI_ISL_717931, EPI_ISL_717932, EPI_ISL_717933, EPI_ISL_717934, EPI_ISL_717935, EPI_ISL_717936, EPI_ISL_717937, EPI_ISL_717938, EPI_ISL_717939, EPI_ISL_717940, EPI_ISL_717941, EPI_ISL_717942, EPI_ISL_717943, EPI_ISL_717944, EPI_ISL_717945, EPI_ISL_717946, EPI_ISL_717947, EPI_ISL_717948, EPI_ISL_717949, EPI_ISL_717950, EPI_ISL_717951, EPI_ISL_717952, EPI_ISL_717953, EPI_ISL_717954, EPI_ISL_717955, EPI_ISL_717956, EPI_ISL_717957, EPI_ISL_717959, EPI_ISL_717960, EPI_ISL_717961 | | | |
| see above | Laboratorio de Virologia Molecular / UFRJ | Bioinformatics Laboratory / LNCC | Alexandra L Gerber; Amilcar Tanuri; Ana Paula de C Guimarães; Ana Tereza R de Vasconcelos; Andréa Cony Cavalcanti; Carolina M Voloch; Claudia dos Santos Rodrigues; Covid19-UFRJ Workgroup; Cynthia C Cardoso; Diana Mariani; Luiz G P de Almeida; Luís Cristóvão Pôrto; Orlando C. Ferreira; Otavio Bustrolini; Otavio J. Brustolini; Renato S Aguiar; Ronaldo S Francisco Jr; Ronaldo da Silva F Jr; Terezinha M P P Castiñeira; Terezinha M P P Castiñeiras |
| EPI_ISL_792522 | Laboratorio de Virología del Hospital de Niños Dr. Ricardo Gutierrez | Grupo de Genómica y Bioinformática del Instituto de Investigación de la Cadena Láctea CONICET-INTA on behalf of 'Proyecto Argentino Interinstitucional de genomica de SARS-CoV-2' (PAIS Consortium) | AF; AS; Acevedo; Alexay; Alvarez Lopez; Amadio; Aulicino; C; Eberhardt; G; Goya; Irazoqui; Jacques; König; M; ME; MF; MS; Mistchenko; Nabaes Jodar; O; P; S; Torres; Viegas, M. |
| EPI_ISL_792172, EPI_ISL_792216, EPI_ISL_792278 | Laboratorio de Virología del Hospital de Niños Dr. Ricardo Gutierrez | Área de Secuenciación del Laboratorio de Virología del Hospital de Niños Dr. Ricardo Gutierrez on behalf of 'Proyecto Argentino Interinstitucional de genomica de SARS-CoV-2' (PAIS Consortium) | AS; Acevedo; Alexay; Alvarez Lopez; C; E; Goya; Gravis; Jacques; LE; Lusso; ME; MI; MS; Mistchenko; Nabaes Jodar; Natale; O; S; Valinotto; Viegas, M. |
| EPI_ISL_671974 | Laboratorio de Virología y Microbiología Molecular, Depto. de Microbiología, Facultad de Medicina, Universidad de El Salvador/INS-laboratorio de Ref. Ministerio de Salud | Laboratorio de Virología y Microbiología Molecular, Depto. de Microbiología, Facultad de Medicina, Universidad de El Salvador/INS-laboratorio de Ref. Ministerio de Salud | Carlos Hernández Ávila; Ortega-Pérez C A; Rivera NR; Xochitl Sandoval López |
| EPI_ISL_792525 | Laboratorio del Hospital Interzonal General de Agudos Evita | Grupo de Genómica y Bioinformática del Instituto de Investigación de la Cadena Láctea CONICET-INTA on behalf of 'Proyecto Argentino Interinstitucional de genomica de SARS-CoV-2' (PAIS Consortium) | AF; Alexay; Amadio; Aulicino; C; Desimone; E; Eberhardt; G; Goya; Grossi; I; Irazoqui; König; L; Luczac; M; MF; MS; Musto; Nabaes Jodar; O; P; S; Serrano; Torres; Viegas, M. |
| EPI_ISL_792444 | Laboratorio del Hospital Regional Ushuaia Gdor. Ernesto Campos | Hospital Regional Ushuaia - Centro Austral De Investigaciones Científicas - Universidad Nacional De Tierra Del Fuego on behalf of 'Proyecto Argentino Interinstitucional de genomica de SARS-CoV-2' (PAIS Consortium) | Boutureira, MF.; CA; CB; CF; Castro; Ceballos; Cáceres; De Roccis; F; G; Gallego; Gramundi; ID; Nardi; SB; SG; Yulan |
| EPI_ISL_511879 | Laboratorium Kesehatan Provinsi Jawa Barat | Molecular Genetics Laboratory-Faculty of Medicine-Universitas Padjadjaran; School of Life Sciences and Technology & School of Pharmacy-Institut Teknologi Bandung; Laboratorium Kesehatan Provinsi Jawa Barat | Agung Eru Wibowo; Azzania Fibriani; Catur Riani; Cut Nur Cinthia Alamanda; Ema Rahmawati; Hammam Riza; Hesti Lina Wiraswati; Husna Nugrahapraja; Lia Faridah; Marselina Irasonia Tan; Rifky Waluyajati Rachman; Ryan Bayusantika Ristandi; Savira Ekawardhani; Soni Solistia Wirawan; Tarwadi; Yunia Sribudiani |
| EPI_ISL_514427, EPI_ISL_514428, EPI_ISL_541651 | Laboratory Diagnostic, Veterinary Specialized Institute Kraljevo | Laboratory Diagnostic, Veterinary Specialized Institute Kraljevo | Afonso, C.; Banovic Djeri, B.; Jankovic, M.; Jovanovic, T.; Knezevic, A.; Petrovic, T.; Sekler, M.; Tesovic, B.; Vidanovic, D.; Volkening, J. |

| | | | |
|---|---|---|---|
| EPI_ISL_482744 | Laboratory Diagnostic, Veterinary Specialized Institute Kraljevo | Laboratory Diagnostic, Veterinary Specialized Institute Kraljevo | Afonso, C.; Banovic Djeri, B.; Knezevic, A.; Tesovic, B.; Vidanovic, D. |
| EPI_ISL_754902, EPI_ISL_754904, EPI_ISL_754907, EPI_ISL_754908, EPI_ISL_754909, EPI_ISL_754913 | Laboratory Diagnostics and Clinical Immunology of Developmental Age, Medical University of Warsaw | genXone SA, Research & Development Laboratory; The Faculty of Mathematics, Informatics and Mechanics of the University of Warsaw | Anna Gambin; Grzegorz Nowicki; Jakub Grabowski; Maciej Sykulski; Michał Kaszuba; Monika Mańkowska-Woźniak; Natalia Drwęska-Matelska; Urszula Demkow; Łukasz Krych |
| EPI_ISL_476221 | Laboratory Fleury | Instituto de Medicina Tropical da Univesidade de São Paulo | Camila Alves Maia da Silva; Darlan da Silva Candido; Erika Regina Manuli; Ester Sabino; Flavia Cristina da Silva Sales; Giula Magalhaes Ferreira; Jaqueline Goes de Jesus; Julien Theze; Mariana Severo Ramundo; Nuno Faria; Samples: Celso Granato; Sequencing: Ingra Morales Claro; Thais de Moura Coletti |
| EPI_ISL_415742, EPI_ISL_417521, EPI_ISL_424970, EPI_ISL_452178, EPI_ISL_464093, EPI_ISL_464094, EPI_ISL_660543, EPI_ISL_660544, EPI_ISL_660545, EPI_ISL_667809 | | | |
| see above | Laboratory Medicine | Department of Laboratory Medicine, Lin-Kou Chang Gung Memorial Hospital, Taoyuan, Taiwan | Cheng-Hsun Chiu; Cheng-Ta Yang; Chi-Hsien Huang; Chung-Guei Huang; Guang-Wu Chen; Kuang-Tso Le; Kuo-Chien Tsao; Kuo-Ming Lee; Mei-Jen Hsiao; Peng-Nien Huang; Po-Wei Huang; Shin-Ru Shih; Shu-Li Yang; Shu-Min Lin; Yi-Chun Liu; Yu-Nong Gong |
| EPI_ISL_411915 | Laboratory Medicine | Department of Laboratory Medicine, Lin-Kou Chang Gung Memorial Hospital, Taoyuan, Taiwan. | Chung-Guei Huang; Kuo-Chien Tsao; Shin-Ru Shih; Shu-Li Yang; Yhu-Chering Huang; Yi-Chun Li; Yu-Nong Gong |
| EPI_ISL_455472 | Laboratory for Respiratory Viruses, Cantacuzino National Military-Medical Institute for Research and Development | Cantacuzino Institute | A.Cretu; L.Ustea; M.Lazar; Tim Durfee |
| EPI_ISL_754238 | Laboratory for Respiratory Viruses, Cantacuzino National Military-Medical Institute for Research and Development | Cantacuzino Institute Virology | Luiza Ustea; Mihaela Lazar; Mühlemann Barbara |
| EPI_ISL_435045 | Laboratory of Applied Genetics | RSE "National Center for Biotechnology" | Alexandr Shevtsov; Asylulan Amirgazin; Ilyas Akhmetollayev; Ruslan Kalendar; Viktoriya Lutsay; Yerlan Ramanculov |
| EPI_ISL_477160 | Laboratory of Dr. John Lednicky | University of Florida | Chang-Yu Wu; John A. Lednicky; and John Glenn Morris, Jr. |
| EPI_ISL_437625 | Laboratory of Genomics & Bioinformatics, Institute of Immunology and Experimental Therapy, Polish Academy of Sciences Oddzial Mikrobiologii Wojewodzkiej Stacji Sanitarno-Epidemiologicznej. | Laboratory of Genomics & Bioinformatics, Institute of Immunology and Experimental Therapy, Polish Academy of Sciences | Aleksandra Herud; Dariusz Martynowski; Dorota Kujawa; Grazyna Zalewska; Joanna Sikorska; Krzysztof Jakub Pawlik; Oskar Karpinski and Lukasz Laczmanski; Paulina Zebrowska |
| EPI_ISL_536399 | Laboratory of Immunovirology. Universidad de Antioquia | Instituto Nacional de Salud - Unidad de Secuenciación y Genómica | Carlos Franco-Muñoz; Diego Álvarez-Diaz and Marcela Mercado-Reyes; Francisco J. Díaz; Katherine Laiton-Donato; Lizdany Flórez; Wbeimar Aguilar-Jimenez |
| EPI_ISL_452328, EPI_ISL_452332, EPI_ISL_452349, EPI_ISL_452352 | Laboratory of Infectious Diseases Center of Beijing Ditan Hospital | Laboratory of Infectious Diseases Center of Beijing Ditan Hospital | Chengjie Jie; Fengting Yu; Linghang Wang; Liting Yan; Siyuan Yang; Yunxia Tang |
| EPI_ISL_717979 | Laboratory of Microbiology and Infectious Diseases, Faculty of Veterinary Medicine, Aristotle University of Thessaloniki, University Campus, 541 24, Thessaloniki, Greece. | Laboratory of Biology, Department of Medicine, Democritus University of Thrace, Alexandroupolis, Greece | Bampali M.; Chaintoutis S.; Dimitriou M.; Dovas C.; Dovrolis N.; Karakasiliotis I. |
| EPI_ISL_418265 | Laboratory of Microbiology, Department of Medicine, National and Kapodistrian University of Athens, Greece | Laboratory of Biology, Department of Medicine, Democritus University of Thrace, Greece | Elisavet Gatzidou; Ioannis Karakasiliotis; Maria Bampali; Nikolaos Dovrolis; Nikolaos Spanakis; Stavroula Veletza |
| EPI_ISL_434466, EPI_ISL_434473, EPI_ISL_437905, EPI_ISL_437911 | Laboratory of Microbiology, Medical School, National and Kapodistrian University of Athens | Laboratory of Biology, Department of Medicine, Democritus University of Thrace | Bampali, M.; Dovrolis, N.; Froukala, E.; Gatzidou, E.; Kassela K.; N. and Karakasiliotis, I.; Spanakis; Stavropoulou, A.; Tsakris, A.; Veletza, S. |
| EPI_ISL_654018, EPI_ISL_654020 | Laboratory of Microbiology, National Reference Lab, Charles Nicolle Hospital; 2-University of Tunis ElManar, Faculty of Medicine of Tunis, LR99ES09, Tunis, Tunisia | 1-Clinical and Experimental Pharmacology Lab, LR16SP02, National Center of Pharmacovigilance, University of Tunis El Manar, Tunis, Tunisia. 2-Neurodegenerative diseases and psychiatric troubles, LR18SP03, Razi Hospital, University of Tunis El Manar, Tunis, Tunisia. 3- Ministry of Health, National Observatory of New and Emerging Diseases, 1006, Tunis, Tunisia | Alia Ben Kahla; Gaies Emna; Ilhem Boutiba-Ben Boubaker; Imen Kacem; Imen Mkada; Jalila Ben Khelil; Maher Kharrat; Mouna Ben Sassi; Mouna Safer; Nissaf Ben Alaya; Riadh Daghfous; Riadh Gouider.; Salma Abid; Sameh Trabelsi; Sana Ferjani; Soumaya Rammeh |
| EPI_ISL_796782, EPI_ISL_803119, EPI_ISL_803817 | Laboratory of Microbiology, National Reference Lab, Charles Nicolle Hospital; 2-University of Tunis ElManar, Faculty of Medicine of Tunis, LR99ES09, Tunis, Tunisia | Clinical and Experimental Pharmacology Lab, LR16SP02, National Center of Pharmacovigilance, University of Tunis El Manar, Tunis, Tunisia. 2-Neurodegenerative diseases and psychiatric troubles, LR18SP03, Razi Hospital, University of Tunis El Manar, Tunis, Tunisia. 3- Ministry of Health, National Observatory of New and Emerging Diseases, 1006, Tunis, Tunisia | Ahmed Fakhfakh; Alia BenKahla; Gaies Emna; Guedi Ali Barreh; Habiba Ben Romdhane; Hanen El Jebari; Ilhem Boutiba-Ben Boubaker; Jalila Ben Khelil; Maher Kharrat; Mouna Ben Sassi; Mouna Safer; Nissaf Ben Alaya; Riadh Daghfous; Riadh Gouider.; Salma Abid; Sameh Trabelsi; Sana Ferjani; Sarra Chamman; Souissi Amira; Zaineb Hamzaoui |
| EPI_ISL_613563, EPI_ISL_613564, EPI_ISL_613707, EPI_ISL_613708 | Laboratory of Molecular Biology, Blood Center of Ribeirão Preto | Laboratory of Molecular Biology, Blood Center of Ribeirão Preto, Faculty of Medicine of Ribeirão Preto, University of São Paulo | Aparecida Y Yamamoto; Diego Villa Clé; Dimas T Covas; Elaine V Santos; Evandra S Rodrigues; Glauco de Carvalho Pereira; Joilson Xavier; Luiz CJ Alcantara; Marta Giovanetti; Rodrigo T Calado; Simone Kashima; Svetoslav N Slavov; Talita Adelino; Vagner Fonseca |
| EPI_ISL_613709, EPI_ISL_613951 | Laboratory of Molecular Biology, Blood Center of Ribeirão Preto, Faculty of Medicine of Ribeirão Preto, University of São Paulo | Laboratory of Molecular Biology, Blood Center of Ribeirão Preto, Faculty of Medicine of Ribeirão Preto, University of São Paulo | Aparecida Y Yamamoto; Diego Villa Clé; Dimas T Covas; Elaine V Santos; Evandra S Rodrigues; Glauco de Carvalho Pereira; Joilson Xavier; Luiz CJ Alcantara; Marta Giovanetti; Rodrigo T Calado; Simone Kashima; Svetoslav N Slavov; Talita Adelino; Vagner Fonseca |
| EPI_ISL_801608, EPI_ISL_801697, EPI_ISL_801703, EPI_ISL_801737, EPI_ISL_801821, EPI_ISL_801840, EPI_ISL_801853 | | | |
| see above | Laboratory of Molecular Virology, Pontificia Universidad Católica de Chile | MSHS Pathogen Surveillance Program | Adolfo Garcia-Sastre; Adriana van De Guchte; Ajay Obla; Aldo Gaggero; Ana Maria Contreras; Ana S. Gonzalez-Reiche; Bremy Alburquerque; Carlos Palma; Constanza Maldonado; Edward C. Holmes; Eileen Serrano; Erick Salinas; Fernando Valiente; Hala Alshammary; Harm van Bakel; Jayeeta Dutta; Jorge Levican; Juan Soto; Leonardo I. Almonacid; M. Belen Leyton; Manuel Ampuero; Marcela Ferres; Matthew M. Hernandez; Melissa Smith; Rafael A. Medina.; Robert Sebra; Shwetha Hara Sridhar; Tamara García-Salum; Viviana Simon; Ying-Chih Wang; Zenab Khan |
| EPI_ISL_427294, EPI_ISL_427295, EPI_ISL_427296, EPI_ISL_427297, EPI_ISL_427298, EPI_ISL_427299, EPI_ISL_427300, EPI_ISL_427301, EPI_ISL_427302, EPI_ISL_427303, EPI_ISL_427304, EPI_ISL_456071, EPI_ISL_456072, EPI_ISL_456073, EPI_ISL_456074, EPI_ISL_456075, EPI_ISL_456079, EPI_ISL_456080, EPI_ISL_456081, EPI_ISL_456084, EPI_ISL_456085, EPI_ISL_456086, EPI_ISL_456087, EPI_ISL_456089, EPI_ISL_456090, EPI_ISL_456091, EPI_ISL_456092, EPI_ISL_456093, EPI_ISL_456094, EPI_ISL_456095, EPI_ISL_456096, EPI_ISL_456097, EPI_ISL_456098, EPI_ISL_456099, EPI_ISL_456100, EPI_ISL_456101, EPI_ISL_456102, EPI_ISL_456103, EPI_ISL_456104, EPI_ISL_456105, EPI_ISL_456106, EPI_ISL_467344, EPI_ISL_467345, EPI_ISL_467346, EPI_ISL_467347, EPI_ISL_467348, EPI_ISL_467349, EPI_ISL_467350, EPI_ISL_467351, EPI_ISL_467352, EPI_ISL_467353, EPI_ISL_467354, EPI_ISL_467355, EPI_ISL_467356, EPI_ISL_467357, EPI_ISL_467358, EPI_ISL_467359, EPI_ISL_467360, EPI_ISL_467361, EPI_ISL_467362, EPI_ISL_467363, EPI_ISL_467364, EPI_ISL_467365, EPI_ISL_467366, EPI_ISL_467367, EPI_ISL_467368, EPI_ISL_467369, EPI_ISL_467370, EPI_ISL_467371, EPI_ISL_541347, EPI_ISL_541348, EPI_ISL_541349, EPI_ISL_541350, EPI_ISL_541351, EPI_ISL_541352, EPI_ISL_541353, EPI_ISL_541354, EPI_ISL_541355, EPI_ISL_541356, EPI_ISL_541357, EPI_ISL_541358, EPI_ISL_541359, EPI_ISL_541360, EPI_ISL_541361, EPI_ISL_541362, EPI_ISL_541363, EPI_ISL_541364, EPI_ISL_541365, EPI_ISL_541366, EPI_ISL_541367, EPI_ISL_541368, EPI_ISL_541369 | | | |
| see above | Laboratory of Respiratory Viruses and Measles, Oswaldo Cruz Institute, FIOCRUZ | Laboratory of Respiratory Viruses and Measles, Oswaldo Cruz Institute, FIOCRUZ | Aline Mattos; Ana Carolina Mendonca; Ana Carolina Mendonça; Anna Carolina Paixao; Anna Carolina Paixão; Braulia Caetano; Cinthia Avila; Cristiana Garcia; Fernando Motta; Jonathan Lopes; Luciana Appolinario; Maria Ogrzewalska; Marilda Siqueira on behalf of the Fiocruz COVID-19 Genomic Surveillance Network; Milene Miranda; Paola Resende; Priscila Born; Roxana Loayza; Sunando Roy |
| EPI_ISL_451298 | Laboratory of Virology, INMI Lazzaro Spallanzani IRCCS | Laboratory of Virology, INMI Lazzaro Spallanzani IRCCS | Antonino Di Caro; Barbara Bartolini; Cesare E.M. Gruber; Francesco Messina; Giuseppe Ippolito; Maria R. Capobianchi; Martina Rueca |
| EPI_ISL_454575 | Laboratory of virology, National Center of Expertise | Laboratory of molecular-genetic research, National Center for Expertise, Kazakhstan National Center for Biotechnology, Kazakhstan | ; Abdaliyev Askar; Akhmetollayev Ilyas; Amirgazin Asylulan; Aushakhmetova Zabira; Kalendar Ruslan; Lutsay Viktoriya; Rakhmetova Akbota; Ramankulov Yerlan; Shevtsov Alexandr |
| EPI_ISL_454585 | Laboratory of virology, National Center of Expertise | Laboratory of molecular-genetic research, National Center for Expertise, Kazakhstan National Center for Biotechnology, Kazakhstan | ; Abdaliyev Askar; Akhmetollayev Ilyas; Amirgazin Asylulan; Aushakhmetova Zabira; Kalendar Ruslan; Lutsay Viktoriya; Rakhmetova Akbota; Ramankulov Yerlan; Shevtsov Alexandr |
| EPI_ISL_454576, EPI_ISL_454579 | Laboratory of virology, National Center of Expertise | Laboratory of molecular-genetic research, National Center of Expertise, Kazakhstan National Center for Biotechnology, Kazakhstan | ; Abdaliyev Askar; Akhmetollayev Ilyas; Amirgazin Asylulan; Aushakhmetova Zabira; Kalendar Ruslan; Lutsay Viktoriya; Rakhmetova Akbota; Ramankulov Yerlan; Shevtsov Alexandr |
| EPI_ISL_576117, EPI_ISL_576120 | Laboratory, The Bio Arte Limited | Laboratory, The Bio Arte Limited | Biazzo, M.; Madeddu, S.; Pinzauti, D.; Santoro, F. |
| EPI_ISL_414045, EPI_ISL_456076, EPI_ISL_456077, EPI_ISL_456082, EPI_ISL_456083, EPI_ISL_456088 | Laboratório Central de Saúde Pública Noel Nutels (LACEN-RJ) | Laboratory of Respiratory Viruses and Measles, Oswaldo Cruz Institute, FIOCRUZ | Aline Mattos; Alisson Fabri; Braulia Caetano; Cristiana Garcia; Fernando Motta; Joilson Xavier; Jonathan Lopes; Luciana Appolinario; Maria Nóbrega; Maria Ogrzewalska; Marilda Siqueira on behalf of the Fiocruz COVID-19 Genomic Surveillance Network; Marilda SiqueiraFiocruz COVID-19 Genomic Surveillance Network; Milene Miranda; Paola Resende; Sunando Roy |
| EPI_ISL_415105, EPI_ISL_427293 | Laboratório Central de Saúde Pública Professor Gonçalo Moniz (LACEN-BA) | Laboratory of Respiratory Viruses and Measles, Oswaldo Cruz Institute, FIOCRUZ | Aline Mattos; Allison Fabri; Braulia Caetano; Cristiana Garcia; Fernando Motta; Joilson Xavier; Jonathan Lopes; Luciana Appolinario; Maria Nóbrega; Maria Ogrzewalska; Marilda Siqueira Fiocruz COVID-19 Genomic Surveillance Network; Marilda Siqueira on behalf of the Fiocruz COVID-19 Genomic Surveillance Network; Milene Miranda; Paola Resende; Priscila Born; Sunando Roy |
| EPI_ISL_792561, EPI_ISL_792562, EPI_ISL_792563, EPI_ISL_792564, EPI_ISL_792565, EPI_ISL_792566, EPI_ISL_792567, EPI_ISL_792568, EPI_ISL_792569, EPI_ISL_792570, EPI_ISL_792571, EPI_ISL_792572, EPI_ISL_792573, EPI_ISL_792574, EPI_ISL_792575, EPI_ISL_792576, EPI_ISL_792577, EPI_ISL_792578, EPI_ISL_792579, EPI_ISL_792580, EPI_ISL_792581, EPI_ISL_792582, EPI_ISL_792583, EPI_ISL_792584, EPI_ISL_792585, EPI_ISL_792586, EPI_ISL_792587, EPI_ISL_792588, EPI_ISL_792589, EPI_ISL_792590, EPI_ISL_792591, EPI_ISL_792592, EPI_ISL_792593, EPI_ISL_792594, EPI_ISL_792595, EPI_ISL_792596, EPI_ISL_792597, EPI_ISL_792598, EPI_ISL_792599, EPI_ISL_792600, EPI_ISL_792601, EPI_ISL_792602, EPI_ISL_792603, EPI_ISL_792604, EPI_ISL_792605, EPI_ISL_792606, EPI_ISL_792607, EPI_ISL_792608, EPI_ISL_792609, EPI_ISL_792610, EPI_ISL_792611, EPI_ISL_792612, EPI_ISL_792613, EPI_ISL_792614, EPI_ISL_792615, EPI_ISL_792616, EPI_ISL_792617, EPI_ISL_792618, EPI_ISL_792619, EPI_ISL_792620, EPI_ISL_792621, EPI_ISL_792622, EPI_ISL_792623, EPI_ISL_792624, EPI_ISL_792625, EPI_ISL_792626, EPI_ISL_792627, EPI_ISL_792628, EPI_ISL_792629, EPI_ISL_792630, EPI_ISL_792631, EPI_ISL_792632, EPI_ISL_792633, EPI_ISL_792634, EPI_ISL_792635, EPI_ISL_792636, EPI_ISL_792637, EPI_ISL_792638 | | | |
| see above | Laboratório Central de Saúde Pública do Estado da Paraíba (LACEN-PB) | Laboratory of Respiratory Viruses and Measles, Oswaldo Cruz Institute, FIOCRUZ | Ana Carolina Mendonca; Anna Carolina Paixao; Dalane Loudal Florentino Teixeira; Fernando Motta; João Felipe Bezerra; Luciana Appolinario; Marilda Siqueira on behalf of the Fiocruz COVID-19 Genomic Surveillance Network; Paola Resende; Romero Henrique Teixeira de Vasconcelos; Thiago Franco de Oliveira Carneiro |
| EPI_ISL_427292, EPI_ISL_792639, EPI_ISL_792640, EPI_ISL_792641, EPI_ISL_792643, EPI_ISL_792644 | Laboratório Central de Saúde Pública do Estado de Alagoas (LACEN-AL) | Laboratory of Respiratory Viruses and Measles, Oswaldo Cruz Institute, FIOCRUZ | Aline Mattos; Ana Carolina Mendonca; Anderson Brandao Leite; Anna Carolina Paixao; Braulia Caetano; Cristiana Garcia; Fernando Motta; Jonathan Lopes; Luciana Appolinario; Maria Ogrzewalska; Marilda Siqueira on behalf of the Fiocruz COVID-19 Genomic Surveillance Network; Milene Miranda; Paola Resende; Priscila Born; Sunando Roy |
| EPI_ISL_500460, EPI_ISL_500461, EPI_ISL_500462, EPI_ISL_500463, EPI_ISL_500471, EPI_ISL_500473, EPI_ISL_500475, EPI_ISL_500477, EPI_ISL_500480, EPI_ISL_500481, EPI_ISL_500482, EPI_ISL_500483, EPI_ISL_500484, EPI_ISL_500485, EPI_ISL_500486, EPI_ISL_500872, EPI_ISL_572334, EPI_ISL_572335, EPI_ISL_572351, EPI_ISL_572353, EPI_ISL_572366, EPI_ISL_572371, EPI_ISL_572386 | | | |

| | | | |
|---|---|---|---|
| see above | Laboratório Central de Saúde Pública do Estado de Pernambuco (LACEN-PE) | WallauLab, Aggeu Magalhaes Institute | Alexandre Freitas da Silva; Antonio Mauro Rezende; Armando de Menezes Neto; Bruna Santos Lima Figueiredo de Sá; Caroline Targino Alves da Silva; Claudio Eduardo Cavalcanti; Constância Flávia Junqueira Ayres; Cássia Docena; Derciliano Lopes da Cruz; Duschinka Ribeiro Duarte Guedes; Elisama Helvecio; Filipe Zimmer Dezordi; Gabriel Luz Wallau on behalf of the Fiocruz COVID-19 Genomic Surveillance Network; Gonzalo Bello; Kamila Gaudêncio da Silva Sales; Larissa Krokovsky; Laís Ceschini Machado; Luciane Caroline Albuquerque Bezerra; Luydson Richardson Silva Vasconcelos; Marcelo Henrique Santos Paiva; Maria Almerice Lopes da Silva; Matheus Filgueira Bezerra; Michelle da Silva Barros; Paola Cristina Resende; Renata Pessôa Germano Mendes; Rodrigo Moraes Loyo Arcoverde; Severino Jefferson Ribeiro da Silva; Sinval Pinto Brandão Filho; Tiago Gräf; Wheverton Ricardo Correia do Nascimento |
| EPI_ISL_427305, EPI_ISL_427306, EPI_ISL_541370 | Laboratório Central de Saúde Pública do Estado de Santa Catarina (LACEN-SC) | Laboratory of Respiratory Viruses and Measles, Oswaldo Cruz Institute, FIOCRUZ | Aline Mattos; Ana Carolina Mendonça; Anna Carolina Paixão; Braulia Caetano; Cristiana Garcia; Fernando Motta; Jonathan Lopes; Luciana Appolinario; Maria Ogrzewalska; Marilda Siqueira on behalf of the Fiocruz COVID-19 Genomic Surveillance Network; Milene Miranda; Paola Resende; Priscila Born; Sandra Bianchini; Sunando Roy |
| EPI_ISL_541372, EPI_ISL_541373, EPI_ISL_541374, EPI_ISL_541375, EPI_ISL_541376, EPI_ISL_541377, EPI_ISL_541378, EPI_ISL_541379, EPI_ISL_541380, EPI_ISL_541381, EPI_ISL_541382, EPI_ISL_541383, EPI_ISL_541384, EPI_ISL_541385, EPI_ISL_541386, EPI_ISL_541387, EPI_ISL_541388, EPI_ISL_541389, EPI_ISL_541390, EPI_ISL_541391, EPI_ISL_541393, EPI_ISL_541394, EPI_ISL_541395, EPI_ISL_541396 | | | |
| see above | Laboratório Central de Saúde Pública do Estado de Sergipe (LACEN-SE) | Laboratory of Respiratory Viruses and Measles, Oswaldo Cruz Institute, FIOCRUZ | Ana Carolina Mendonça; Anna Carolina Paixão; Clioma Santos; Fernando Motta; Jonathan Lopes; Luciana Appolinario; Marilda Siqueira on behalf of the Fiocruz COVID-19 Genomic Surveillance Network; Paola Resende |
| EPI_ISL_801397, EPI_ISL_801398, EPI_ISL_801399, EPI_ISL_801400, EPI_ISL_801401, EPI_ISL_801402, EPI_ISL_801403 | | | |
| see above | Laboratório Central de Saúde Pública do Estado do Amazonas (LACEN-AM) | Laboratorio de Ecologia de Doencas Transmissiveis na Amazonia, Instituto Leonidas e Maria Deane - Fiocruz Amazonia | André Corado; Debora Duarte; Felipe Naveca on behalf of the Fiocruz COVID-19 Genomic Surveillance Network; Fernanda Nascimento; George Silva; Luciana Gonçalves; Maria Júlia Brandão; Michele Jesus; Valdinete Nascimento; Victor Souza; Ágatha Costa |
| EPI_ISL_415128 | Laboratório Central de Saúde Pública do Estado do Espírito Santo (LACEN-ES) | Laboratory of Respiratory Viruses and Measles, Oswaldo Cruz Institute, FIOCRUZ | Aline Mattos; Allison Fabri; Braulia Caetano; Cristiana Garcia; Fernando Motta; Joilson Xavier; Jonathan Lopes; Luciana Appolinario; Maria Nóbrega; Maria Ogrzewalska; Marilda Siqueira on behalf of the Fiocruz COVID-19 Genomic Surveillance Network; Milene Miranda; Paola Resende; Sunando Roy |
| EPI_ISL_541340, EPI_ISL_541341, EPI_ISL_541342, EPI_ISL_541343, EPI_ISL_541344, EPI_ISL_541345, EPI_ISL_541346, EPI_ISL_792645, EPI_ISL_792646, EPI_ISL_792647, EPI_ISL_792648, EPI_ISL_792649, EPI_ISL_792650, EPI_ISL_792651, EPI_ISL_792652, EPI_ISL_792653, EPI_ISL_792654 | | | |
| see above | Laboratório Central de Saúde Pública do Estado do Paraná (LACEN-PR) | Laboratory of Respiratory Viruses and Measles, Oswaldo Cruz Institute, FIOCRUZ | Ana Carolina Mendonca; Ana Carolina Mendonça; Anna Carolina Paixao; Anna Carolina Paixão; Fernando Motta; Irina Nastassja Riediger; Irina Riediger; Jonathan Lopes; Luciana Appolinario; Maria do Carmo Debur; Marilda Siqueira on behalf of the Fiocruz COVID-19 Genomic Surveillance Network; Paola Resende |
| EPI_ISL_729794, EPI_ISL_729800, EPI_ISL_729801, EPI_ISL_729802, EPI_ISL_729803, EPI_ISL_729804, EPI_ISL_729805, EPI_ISL_729806, EPI_ISL_729807, EPI_ISL_729808, EPI_ISL_729809, EPI_ISL_729810, EPI_ISL_729811, EPI_ISL_729812, EPI_ISL_729813, EPI_ISL_729814, EPI_ISL_729815, EPI_ISL_729816, EPI_ISL_729817, EPI_ISL_729818, EPI_ISL_729819, EPI_ISL_729820, EPI_ISL_729821, EPI_ISL_729822, EPI_ISL_729823, EPI_ISL_729824, EPI_ISL_729825, EPI_ISL_729826, EPI_ISL_729827, EPI_ISL_729828, EPI_ISL_729829, EPI_ISL_729830, EPI_ISL_729831, EPI_ISL_729832, EPI_ISL_729833, EPI_ISL_729834, EPI_ISL_729835, EPI_ISL_729836, EPI_ISL_729837, EPI_ISL_729838, EPI_ISL_729839, EPI_ISL_729840, EPI_ISL_729841, EPI_ISL_729842, EPI_ISL_729843, EPI_ISL_729844, EPI_ISL_729845, EPI_ISL_729846, EPI_ISL_729847, EPI_ISL_729848, EPI_ISL_729849, EPI_ISL_729850, EPI_ISL_729851, EPI_ISL_729852, EPI_ISL_729853, EPI_ISL_729854, EPI_ISL_729855, EPI_ISL_729856, EPI_ISL_729857, EPI_ISL_729858, EPI_ISL_729859, EPI_ISL_729860, EPI_ISL_729861 | | | |
| see above | Laboratório Central de Saúde Pública do Estado do Rio Grande do Sul (LACEN-RS) | Laboratory of Respiratory Viruses and Measles, Oswaldo Cruz Institute, FIOCRUZ | Ana Carolina Mendonça; Anna Carolina Paixão; Fernando Motta; Luciana Appolinario; Marilda Siqueira on behalf of the Fiocruz COVID-19 Genomic Surveillance Network; Marilda Tereza Mar da Rosa; Paola Resende; Tatiana Schaffer Gregianini |
| EPI_ISL_693220, EPI_ISL_693223, EPI_ISL_693224, EPI_ISL_693243 | Laboratório Municipal de Piracicaba | Instituto Adolfo Lutz, Interdiciplinary Procedures Center, Strategic Laboratory | Claudia Regina Gonçalves; Claudio Tavares Sacchi; Erica Valessa Ramos Gomes; Karoline Rodrigues Campos |
| EPI_ISL_636737, EPI_ISL_636834, EPI_ISL_636835, EPI_ISL_636837 | Laboratório de Imunofarmacologia - Instituto Oswaldo Cruz | Laboratório de Imunofarmacologia - Instituto Oswaldo Cruz | A.D.; C.Q.; De Paula; F.B.; Ferreira; Fintelman-Rodrigues, N.; M.A. and Sacramento; Saraiva; Souza; T.M. |
| EPI_ISL_770551, EPI_ISL_770552, EPI_ISL_770553, EPI_ISL_770554, EPI_ISL_770555, EPI_ISL_770556, EPI_ISL_770557, EPI_ISL_770558, EPI_ISL_770559, EPI_ISL_770560, EPI_ISL_770561, EPI_ISL_770562, EPI_ISL_770563, EPI_ISL_770564, EPI_ISL_770565, EPI_ISL_770566, EPI_ISL_770567, EPI_ISL_770568, EPI_ISL_770569, EPI_ISL_770570, EPI_ISL_770571, EPI_ISL_770572, EPI_ISL_770573, EPI_ISL_770574, EPI_ISL_770575, EPI_ISL_770576, EPI_ISL_770577, EPI_ISL_770578, EPI_ISL_770579, EPI_ISL_770580, EPI_ISL_770581, EPI_ISL_770582, EPI_ISL_770583, EPI_ISL_770584, EPI_ISL_770585, EPI_ISL_770586, EPI_ISL_770587, EPI_ISL_770588, EPI_ISL_770589, EPI_ISL_770590, EPI_ISL_770591, EPI_ISL_770592, EPI_ISL_770593, EPI_ISL_770594, EPI_ISL_770595, EPI_ISL_770596, EPI_ISL_770597, EPI_ISL_770598, EPI_ISL_770599, EPI_ISL_770600, EPI_ISL_770601, EPI_ISL_770602, EPI_ISL_770603, EPI_ISL_770604, EPI_ISL_770605, EPI_ISL_770606, EPI_ISL_770607, EPI_ISL_770608, EPI_ISL_770609, EPI_ISL_770610, EPI_ISL_770611, EPI_ISL_770612, EPI_ISL_770613, EPI_ISL_770614, EPI_ISL_770615, EPI_ISL_770616, EPI_ISL_770617, EPI_ISL_770618, EPI_ISL_770619, EPI_ISL_770620, EPI_ISL_770621, EPI_ISL_770622, EPI_ISL_770623, EPI_ISL_770624, EPI_ISL_770625, EPI_ISL_770626, EPI_ISL_770627, EPI_ISL_770628, EPI_ISL_770629, EPI_ISL_770630, EPI_ISL_779155, EPI_ISL_779156, EPI_ISL_779157, EPI_ISL_779158, EPI_ISL_779159, EPI_ISL_779160, EPI_ISL_779161, EPI_ISL_779162, EPI_ISL_779163, EPI_ISL_779164, EPI_ISL_779165, EPI_ISL_779166, EPI_ISL_779167, EPI_ISL_779168 | | | |
| see above | Laboratório de Microbiologia Molecular - Universidade FEEVALE | Bioinformatics Laboratory / LNCC | Alana Witt Hansen; Alessandra Pavan Lamarca da Silva; Alexandra L Gerber; Ana Karolina Eisen Antunes; Ana Luiza Ziulkoski; Ana Paula de C Guimarães; Ana Tereza R de Vasconcelos; Bruna Hermann; Fagner Henrique Heldt; Felipe Benites; Fernando Rosado Spilki; Juliana Schons; Juliane Deise Fleck; Karoline Schallenberger; Larissa Mallmann; Luiz G P de Almeida; Matheus Nunes Weber; Meriane Demoliner; Paula Rodrigues de Almeida; Ronaldo da Silva F Jr; Vyctoria Goes |
| EPI_ISL_476341, EPI_ISL_476395, EPI_ISL_476398 | Laboratório de Patologia Clínica - UNICAMP | Laboratório de Estudos de Vírus Emergentes - UNICAMP | Angelica Schreiber; Camila Simeoni; Darlan da Silva Candido; Jaqueline Goes Jesus e William Marciel de Souza; José Luiz Proença-Modena; Julia Forato; Julien Theze; Luiz Gonzaga; Magnun Nueldo Nunes dos Santos; Marcilio Jorge Fumagalli; Mariene Ribeiro Amorim; Nuno Rodrigues Faria |
| EPI_ISL_738338 | Landstuhl Regional Medical Center | United States Air Force School of Aerospace Medicine | Amanda Javorina; Anthony Fries; Clarise Starr; Cole Anderson; Elizabeth Macias; Fritz Castillo; Jennifer Meyer; Sarah Purves; William Gruner |
| EPI_ISL_407079 | Lapland Central Hospital | Department of Virology, University of Helsinki and Helsinki University Hospital, Helsinki, Finland | Hannimari Kallio-Kokko; Olli Vapalahti; Suvi Kuivanen; Teemu Smura |
| EPI_ISL_486434, EPI_ISL_486435, EPI_ISL_512652, EPI_ISL_515185, EPI_ISL_770047 | Latvijas Infektoloģijas centrs | Latvian Biomedical Research and Study Centre | Ivars Silamiķelis; Jeļena Storoženko; Jānis Kloviņš; Jānis Pjalkovskis; Kaspars Megnis; Monta Ustinova; Oksana Savicka; Tatjana Kolupajeva; Uga Dumpis; Vita Rovīte; Ņikita Zrelovs |
| EPI_ISL_498554 | Lebanese American University | Lebanese American University | Abdallah, J.; Abi Habib, W.; El Shesheny, R.; Goldstein; J. and Kayali, G.; Mokhbat, J.; R.J.; Webby |
| EPI_ISL_434677 | Lednicky Laboratory at Emerging Pathogens Institute | Lednicky Laboratory at Emerging Pathogens Institute | C.-Y.; Clugston; Elbadry; J.A.; J.G. Jr. and Lednicky; J.R.; M.A.; Morris; S.N.; Shankar; Wu |
| EPI_ISL_649293, EPI_ISL_656608, EPI_ISL_796865, EPI_ISL_798401 | Lighthouse Lab in Alderley Park | Wellcome Sanger Institute for the COVID-19 Genomics UK (COG-UK) Consortium | Cordelia Langford; David K. Jackson; Dominic Kwiatkowski; Ewan Harrison; Ian Johnston; Jacquelyn Wynn; John Sillitoe on behalf of the Wellcome Sanger Institute COVID-19 Surveillance Team; John Sillitoe on behalf of the Wellcome Sanger Institute COVID-19 Surveillance Team (http://www.sanger.ac.uk/covid-team); Mairead Hyland; Roberto Amato; Sonia Goncalves; The Lighthouse Lab in Alderley Park and Alex Alderton |
| EPI_ISL_551760, EPI_ISL_553964, EPI_ISL_554730, EPI_ISL_555672, EPI_ISL_557072, EPI_ISL_557673, EPI_ISL_558873, EPI_ISL_607149 | | | |
| see above | Lighthouse Lab in Alderley Park | Wellcome Sanger Institute for the COVID-19 Genomics UK (COG-UK) consortium | Cordelia Langford; David K. Jackson; Dominic Kwiatkowski; Ewan Harrison; Ian Johnston; Jacquelyn Wynn; John Sillitoe on behalf of the Wellcome Sanger Institute COVID-19 Surveillance Team; John Sillitoe on behalf of the Wellcome Sanger Institute COVID-19 Surveillance Team (http://www.sanger.ac.uk/covid-team); Mairead Hyland; Roberto Amato; Sonia Goncalves; The Lighthouse Lab in Alderley Park and Alex Alderton |
| EPI_ISL_702043, EPI_ISL_718937 | Lighthouse Lab in Cambridge | Wellcome Sanger Institute for the COVID-19 Genomics UK (COG-UK) Consortium | Cordelia Langford; David K. Jackson; Dominic Kwiatkowski; Ewan Harrison; Ian Johnston; John Sillitoe on behalf of the Wellcome Sanger Institute COVID-19 Surveillance Team; Rob Howes; Roberto Amato; Sonia Goncalves; The Lighthouse Lab in Cambridge and Alex Alderton |
| EPI_ISL_551875, EPI_ISL_556549, EPI_ISL_609159 | Lighthouse Lab in Cambridge | Wellcome Sanger Institute for the COVID-19 Genomics UK (COG-UK) consortium | Cordelia Langford; David K. Jackson; Dominic Kwiatkowski; Ewan Harrison; Ian Johnston; John Sillitoe on behalf of the Wellcome Sanger Institute COVID-19 Surveillance Team; Rob Howes; Roberto Amato; Sonia Goncalves; The Lighthouse Lab in Cambridge and Alex Alderton |
| EPI_ISL_662943, EPI_ISL_676391, EPI_ISL_757152, EPI_ISL_797432, EPI_ISL_797840 | Lighthouse Lab in Glasgow | Wellcome Sanger Institute for the COVID-19 Genomics UK (COG-UK) Consortium | Anna Dominiczak and Alex Alderton; Carol Clugston; Cordelia Langford; David Gray; David K. Jackson; Dominic Kwiatkowski; Ewan Harrison; Harper VanSteenhouse; Ian Johnston; John Sillitoe on behalf of the Wellcome Sanger Institute COVID-19 Surveillance Team; Roberto Amato; Sonia Goncalves; Yumi Kasai |
| EPI_ISL_530611, EPI_ISL_531127, EPI_ISL_531375, EPI_ISL_531848, EPI_ISL_532578, EPI_ISL_532842, EPI_ISL_533067, EPI_ISL_533244, EPI_ISL_533266, EPI_ISL_533305, EPI_ISL_537083, EPI_ISL_537093, EPI_ISL_537136, EPI_ISL_540031, EPI_ISL_540302, EPI_ISL_549687, EPI_ISL_568324, EPI_ISL_579830, EPI_ISL_590489, EPI_ISL_601205, EPI_ISL_602112 | | | |
| see above | Lighthouse Lab in Glasgow | Wellcome Sanger Institute for the COVID-19 Genomics UK (COG-UK) consortium | Anna Dominiczak and Alex Alderton; Carol Clugston; Cordelia Langford; David Gray; David K. Jackson; Dominic Kwiatkowski; Ewan Harrison; Harper VanSteenhouse; Ian Johnston; John Sillitoe; John Sillitoe on behalf of the Wellcome Sanger Institute COVID-19 Surveillance Team; John Sillitoe on behalf of the Wellcome Sanger Institute COVID-19 Surveillance Team (http://www.sanger.ac.uk/covid-team); Roberto Amato; Sonia Goncalves; Yumi Kasai |
| EPI_ISL_720448, EPI_ISL_798412 | Lighthouse Lab in Milton Keynes | Wellcome Sanger Institute for the COVID-19 Genomics UK (COG-UK) Consortium | Cordelia Langford; David K. Jackson; Dominic Kwiatkowski; Ewan Harrison; Ian Johnston; John Sillitoe on behalf of the Wellcome Sanger Institute COVID-19 Surveillance Team; Roberto Amato; Sonia Goncalves; The Lighthouse Lab in Milton Keynes and Alex Alderton |
| EPI_ISL_549940, EPI_ISL_550448, EPI_ISL_552310, EPI_ISL_553102, EPI_ISL_566729, EPI_ISL_568476, EPI_ISL_575987, EPI_ISL_580649, EPI_ISL_600005, EPI_ISL_601762, EPI_ISL_608755, EPI_ISL_634484 | | | |
| see above | Lighthouse Lab in Milton Keynes | Wellcome Sanger Institute for the COVID-19 Genomics UK (COG-UK) consortium | Cordelia Langford; David K. Jackson; Dominic Kwiatkowski; Ewan Harrison; Ian Johnston; John Sillitoe on behalf of the Wellcome Sanger Institute COVID-19 Surveillance Team; John Sillitoe on behalf of the Wellcome Sanger Institute COVID-19 Surveillance Team (http://www.sanger.ac.uk/covid-team); Roberto Amato; Sonia Goncalves; The Lighthouse Lab in Alderley Park and Alex Alderton; The Lighthouse Lab in Milton Keynes and Alex Alderton |
| EPI_ISL_541859, EPI_ISL_541873 | Lithuanian University of Health Sciences Hospital, Department of Laboratory Medicine | Lithuanian University of Health Sciences, Laboratory of Molecular Cardiology | Arnoldas Pautienius; Astra Vitkauskiene; Dovydas Gecys; Kamile Tamusauskaite; Laura Pareckaite; Lukas Zemaitis; Vaiva Lesauskaite |
| EPI_ISL_603090, EPI_ISL_603116, EPI_ISL_636840, EPI_ISL_636842, EPI_ISL_636844, EPI_ISL_636856, EPI_ISL_636864, EPI_ISL_636872, EPI_ISL_636881, EPI_ISL_636886, EPI_ISL_636892, EPI_ISL_770511 | | | |
| see above | Lithuanian University of Health Sciences Hospital, Department of Laboratory Medicine | Lithuanian University of Health Sciences, Molecular cardiology lab. | Arnoldas Pautienius; Astra Vitkauskiene; Dovydas Gecys; Ingrida Olendraitė; Kamile Tamusauskaite; Laura Pareckaite; Lukas Zemaitis; Vaiva Lesauskaite |
| EPI_ISL_526384 | Liverpool Clinical Laboratories | COVID-19 Genomics UK (COG-UK) Consortium | A Alrezaihi; Alessandro Gerada; Alistair Darby; Angela Cowell; Anita Lucaci; Anu Chawla; Cassie Olateju; Catherine Hartley; Charlotte Nelson; Ecaterina Vamos; Elaine O'Toole; Eleanor G Bentley; Ghada T Shawli; Isabel García-Dorival; James Johnson; James P Stewart; Jenifer Manson; Joanne Watts; Jones Benjamin; Jordan J Clark; Julian Hiscox; L Luu; Lucille Rainbow; M Almsaud; Margaret Hughes; Mark Whitehead; Matthew Gemmell; Miren Iturriza-Gomara; Muhannad Alruwaili; N.P Randle; Neil Swainston; PKF Gilmore; Parul Sharma; Rebekah Penrice-Randal; Richard Eccles; Richard Gregory; Sam Haldenby; Steve Paterson; Stuart D Armstrong; Trevor Ian Robinson; Ximeng Han |
| EPI_ISL_534706 | MD PHL | MD PHL | Maryland Department of Health Laboratories Administration |
| EPI_ISL_568954, EPI_ISL_568973, EPI_ISL_568993, EPI_ISL_568998, EPI_ISL_569023, EPI_ISL_569084, EPI_ISL_569116, EPI_ISL_569208, EPI_ISL_644396, EPI_ISL_644516, EPI_ISL_644548 | | | |
| see above | MEPHI, Aix Marseille University | MEPHI, Aix Marseille University | Anthony LEVASSEUR |
| EPI_ISL_413574 | MHC West-Brabant | Erasmus Medical Center | Anne van der Linden; Annemiek van der Eijk; Aura Timen; Bas Oude Munnink; Claudia Schapendonk; Corien Swaan; Corine GeurtsvanKessel; David Nieuwenhuijse; Irina Chestakova; Jeroen van Kampen; Jolanda Voermans; Madelief Mollers; Manon Haverkate; Marion Koopmans; Mark Pronk; Mart Stein; Pascal Lexmond; Reina Sikkema; Richard Molenkamp; Sandra Kengne Kamga Mobou; |

| | | | |
|---|---|---|---|
| | | | on behalf of the Dutch national COVID-19 response team. |
| EPI_ISL_428856 | MRCG at LSHTM Genomics Lab | MRCG at LSHTM Genomics lab | Sesay et al |
| EPI_ISL_428857, EPI_ISL_471158, EPI_ISL_471164, EPI_ISL_471171, EPI_ISL_560995, EPI_ISL_561021, EPI_ISL_561034, EPI_ISL_561038, EPI_ISL_561052, EPI_ISL_561130, EPI_ISL_561218, EPI_ISL_561229, EPI_ISL_561283, EPI_ISL_561285, EPI_ISL_561292 | | | |
| see above | MRCG at LSHTM Genomics lab | MRCG at LSHTM Genomics lab | Abdoulie Kanteh; Abdul Karim sesay; Bakary Sanyang; Jarra Manneh; Mariama Kujabi; Sesay et al |
| EPI_ISL_428855 | MRCG at LSHTM Geomics lab | MRCG at LSHTM Genomics lab | Sesay et al |
| EPI_ISL_450008 | MSHS Clinical Microbiology Laboratories | MSHS Pathogen Surveillance Program | Adolfo Garcia-Sastre; Ajay Obla; Alberto Paniz-mondolfi; Ana S. Gonzalez-Reiche; Bremy Alburquerque; Emilia Sordillo; Florian Krammer; Gopi Patel; Harm van Bakel; Jayeeta Dutta; Jose Polanco; Juan Soto; Judith Aberg; Lisa Miorin; Matthew Hernandez; Melissa Gitman; Melissa Smith; Mitchell Sullivan; Randy Albrecht; Robert Sebra; Shelcie Fabre; Shwetha Sridhar Hara; Viviana Simon; Wen-chun Liu; Ying-Chih Wang; Zenab Khan |
| EPI_ISL_640259 | MVZ Laborärzte Singen | MVZ Laborärzte Singen | Folker Wenzel; Frithjof Blessing; Jonas Schmidt; Sandro Berghaus |
| EPI_ISL_528744 | Malaysia Genome Institute | Malaysia Genome Institute | Azrin Ahmad; Enizza Kasim; Irni Suhayu Sapian; Mohd Faizal Abu Bakar; Mohd Noor Mat Isa; Nor Azfa Johari; Nurhezreen Md Iqbal; Shahrul Hisham Zainal Ariffin; Shamsidar Sopie; Siti Noraini Othman; Yusuf Muhammad Noor |
| EPI_ISL_693021, EPI_ISL_693078 | Massachusetts State Public Health Laboratory | Massachusetts State Public Health Laboratory | Andrew Lang; Glen Gallagher; Sandra Smole; Timelia Fink |
| EPI_ISL_466888, EPI_ISL_466895, EPI_ISL_466902 | Max von Pettenkofer Institute, Virology, National Reference Center for Retroviruses, LMU München | Laboratory for Functional Genome Analysis, Dept. Genomics, Gene Center of the LMU Munich | Alexander Graf; Helmut Blum; Max Muenchhoff; Oliver Keppler; Stefan Krebs |
| EPI_ISL_569620 | McCrossan Boys Ranch | South Dakota Public Health Laboratory | Jacob Garfin; Matt Plumb; Xiong Wang; and Chris Carlson |
| EPI_ISL_482760, EPI_ISL_483036, EPI_ISL_483038 | Medical Ain Shams Research Institute (MASRI), Ain Shams University | Medical Ain Shams Research Institute (MASRI), Ain Shams University | Ahmad Moustafa; Ashraf Omar; Aya Mohamed; Ghada Ismael; Hala Hafez; Hesham Elghazaly; Mahmoud Elmeitini; Osama Mansour; Reham Mamdouh; Sara Elnakeep; Sara Hassan Agwa; Shaimaa Moustafa |
| EPI_ISL_495542 | Medical Disagnostics Services (MDS) | KRISP, KZN Research Innovation and Sequencing Platform | Chimukangara B; Giandhari J; Khan S; Lessells R; Mdlalose K; Pillay S; Tegally H; Wilkinson E; York D; de Oliveira T |
| EPI_ISL_635169, EPI_ISL_759977 | Medical Microbiology Unit, Department for Laboratory Medicine, Drammen Hospital, Vestre Viken Health Trust, | Norwegian Institute of Public Health, Department of Virology | Hilde Elshaug; Hilde Vollan; Kamilla Heddeland Instefjord; Karoline Bragstad; Kathrine Stene-Johansen; Marie Paulsen Madsen; Olav Hungnes; Rasmus Riis Kopperud |
| EPI_ISL_707692, EPI_ISL_707696 | Medical Research Center, Faculty of Medicine, Syarif Hidayatullah State Islamic University Jakarta | Medical Research Center, Faculty of Medicine, Syarif Hidayatullah State Islamic University Jakarta | Chris Adhiyanto; Dennis Nurjadi; Erike Suwarsono; Ferania Mela; Laifa Hendarmin; Rini Puspitaningrum; Zeti Harriyati |
| EPI_ISL_591527 | Medicina Norte U Chile - Servicio Medico Legal | Center for Mathematical Modeling and Center for Genome Regulation. Santiago, Chile | Allende ML; Ferres M.; Gaete A; Gaggero A; González M; Maass A; Palma R; Travisany D; Urra C; Valiente F; Varas M |
| EPI_ISL_536411 | Medtimes Molecular Laboratory | Medtimes Molecular Laboratory | Eric Chan; Isaac Chow; Jacqueline Tam; Winsome Wong |
| EPI_ISL_471885, EPI_ISL_577582, EPI_ISL_666950 | Michigan Department of Health and Human Services, Bureau of Laboratories | Michigan Department of Health and Human Services, Bureau of Laboratories | Blankenship HM; Riner D; Soehnlen MK |
| EPI_ISL_518776, EPI_ISL_519754, EPI_ISL_520546, EPI_ISL_521778, EPI_ISL_562821, EPI_ISL_562952, EPI_ISL_562994, EPI_ISL_563576, EPI_ISL_563957, EPI_ISL_591999, EPI_ISL_592326, EPI_ISL_593218, EPI_ISL_593475, EPI_ISL_663721 | | | |
| see above | Microbiological Diagnostic Unit - Public Health Laboratory (MDU-PHL) | MDU-PHL | M. B.; M.L.; N.L.; Sait; Sait, M.; Schultz; Schultz M.; Schultz M. B.; Schultz M.B.; Seemann T.; Seemann, T.; Sherry; Sherry, N. |
| EPI_ISL_513925 | Microbiology & Bioinformatics and Biostatistics, Kohat University of Science and Technology (Pakistan) & Shanghai Jiao Tong University (China) | Microbiology & Bioinformatics and Biostatistics, Kohat University of Science and Technology (Pakistan) & Shanghai Jiao Tong University (China) | A.S.; Ali, S.; D.Q.; Khan; M.T.; Muhammad; N. and Wei; T.A. |
| EPI_ISL_418269 | Microbiology and Immunology department, Pasteur institute in Ho Chi Minh city | Microbiology and Immunology department, Pasteur institute in Ho Chi Minh city | A.H.; C.Q.; Cao; D.T.; Dao; H.Q.; H.T.; H.T.T.; Hoang; Huynh; K.C.; L.T.; L.T. and Nguyen; L.T.K.; Le; Luong; M.H.; N.P.H.; N.T.; Nguyen; Pham; Phan; Phung; Q.C.; Q.D.; S.N.; T.M.; T.N.A.; T.T.; T.T.N.; T.V.; Tran; Tran, T.; Vo; Vu |
| EPI_ISL_447267 | Microbiology laboratory, Assuta Ashdod University-Affiliated Hospital | Stern Lab | Stern Lab |
| EPI_ISL_547433, EPI_ISL_547436, EPI_ISL_547442, EPI_ISL_547444, EPI_ISL_613664, EPI_ISL_615024 | Microbiology, Department of Pathology, St. Bernard's Hospital, Gibraltar Health Authority | Respiratory Virus Unit, Microbiology Services Colindale, Public Health England | Charlotte Gillborn-Jones (Gibraltar); Dr Nicholas Cortes (Gibraltar); PHE Covid Sequencing Team |
| EPI_ISL_548010, EPI_ISL_548132, EPI_ISL_579100, EPI_ISL_622817, EPI_ISL_637086, EPI_ISL_649122, EPI_ISL_682281, EPI_ISL_682288, EPI_ISL_707806, EPI_ISL_794626 | | | |
| see above | Middlemore Hospital | Institute of Environmental Science and Research (ESR) | Anja Werno; Antje van der Linden; Arlo Upton; Chris Mansell; David Hammer; Dragana Drinkovic; Erasmus Smit; Gary McAuliffe; Hana Sofia Andersson; Hermes Perez; James Ussher; Jill Sherwood; Jing Wang; Joep de Ligt; Josh Freeman; Julia Howard; Juliet Elvy; Lauren Jelly; Mary DeAlmeida; Matt Blakiston; Matt Storey; Matthew Rogers; Max Bloomfield; Michael Addidle; Michelle Balm; Muhammad Faisal; Nikki Freed; Olin Silander; Sally Roberts; Sarah Jefferies; Sharmini Muttaiyah; Susan Morpeth; Susan Taylor; Timothy Blackmore; Vani Sathyendran; Veronica Playle; Virginia Hope; Xiaoyun Ren |
| EPI_ISL_678288 | Mikrobiologie, RARI | Mikrobiologie, RARI | A.V.; Badanin; D.V.; E.A.; Fedorov; Guseva; Krasnov; Kutyrev; N.A.; N.P.; Naryshkina; Portenko; S.A.; Sharapova; Shcherbakova; Sosedova; V.V.; Y.M. |
| EPI_ISL_735413 | Militello Centro de Diagnosticos e Biopesquisa Clinica | Instituto Adolfo Lutz, Interdiciplinary Procedures Center, Strategic Laboratory | Claudia Regina Gonçalves; Claudio Tavares Sacchi; Erica Valessa Ramos Gomes; Karoline Rodrigues Campos |
| EPI_ISL_718139, EPI_ISL_718140, EPI_ISL_718143, EPI_ISL_718146, EPI_ISL_718156, EPI_ISL_718158, EPI_ISL_718168, EPI_ISL_718184, EPI_ISL_718210 | | | |
| see above | Ministry of Health Hospitals | Institute of Health and Community Medicine | Chan Chia Jui; Chua Hock Hin; David Perera; Ooi Mong How; Tonnii Sia Loong Loong; Wong Jyn Shan; Wong Kiing Aik |
| EPI_ISL_428712, EPI_ISL_437315, EPI_ISL_437329 | Ministry of Health Turkey | Ministry of Health Turkey | Ayşe Başak Altaş; Fatma Bayrakdar; Gülay Korukluoğlu; Selçuk Kılıç; Süleyman Yalçın; Tülin Demir; Yasemin Coşgun |
| EPI_ISL_427408 | Ministry of Public Health (MoPH) | Biomedical Research Center (BRC) | Abdullatif Al-Khal; Ajaeb D. M. H. Al-Nabet; Asmaa A. Al-Thani.; Einas A. E. Al-Kuwari; Fatiha M. Benslimane; Hadi M. Yassine; Hamad E. Al-Romaihi; Heba A. Al-Khatib; Mohammed Al-Thani; Muna A. S. Al-Maslamani; Nourah B. M. Younes; Peter V. Coyle; Salih Al-Marri; Sonia Boughattas |
| EPI_ISL_435119 | Mohammed Bin Rashid University of Medicine and Health Sciences | Al Jalila Children's Hospital | Abdulmajeed Alkhaja; Abiola Catherine Senok; Ahmad Abou Tayoun; Alawi Alsheikh-Ali; Divinlal Harilal; Hamda Khansaheb; Hanan Al Suwaidi; Mohammed Uddin; Norbert Nowotny; Qutayba Hamid; Rabih Halwani; Rifat Hamoudi; Rupa Murthy Varghese; Sathishkumar Ramaswamy; Tom Loney; Zulfa Omar Deesi |
| EPI_ISL_435129, EPI_ISL_435134, EPI_ISL_469280, EPI_ISL_520671, EPI_ISL_520706, EPI_ISL_520733, EPI_ISL_520734, EPI_ISL_520743 | | | |
| see above | Mohammed Bin Rashid University of Medicine and Health Sciences | Al Jalila Genomics Center | Abdulmajeed Alkhaja; Abiola Catherine Senok; Ahmad Abou Tayoun; Alawi Alsheikh-Ali; Divinlal Harilal; Hamda Khansaheb; Hanan Al Suwaidi; Mohammed Uddin; Norbert Nowotny; Qutayba Hamid; Rabih Halwani; Rifat Hamoudi; Rupa Murthy Varghese; Sathishkumar Ramaswamy; Tom Loney; Zulfa Omar Deesi |
| EPI_ISL_467506, EPI_ISL_482719 | Molecular Diagnostics Services (MDS) | KRISP, KZN Research Innovation and Sequencing Platform | Chimukangara B; Giandhari J; Khan S; Lessells R; Mdlalose K; Pillay S; Tegally H; Wilkinson E; York D; de Oliveira T |
| EPI_ISL_681692, EPI_ISL_681696 | Molecular Medicine Laboratory, University of Magallanes | Centro Asistencial Docente y de Investigacion, Universidad de Magallanes | Diego Alvarez; Hermy Alvarez; Jacqueline Aldridge; Jorge González; Marcelo Navarrete; Marco Montes de Oca; Roberto Uribe-Paredes |
| EPI_ISL_802551, EPI_ISL_802564 | Molecular Microbiology and Food Research Laboratory (MMFRLAB) - Universidad San Sebastián | Facultad de Ciencias de la Vida, UNAB | Claudio Meneses; César Echeverría; Dayán Sanhueza; Eduardo Castro; Jorge Olivares; Macarena Bastías; Sebastián Wolter; Waldo Díaz |
| EPI_ISL_460083 | Molecular Virology Unit, Fondazione IRCCS Policlinico San Matteo , Pavia | Laboratory of Virology, INMI Lazzaro Spallanzani IRCCS | Antonino Di Caro; Antonio Piralla; Barbara Bartolini; Cesare E.M. Gruber; Fausto Baldanti; Maria R. Capobianchi; Martina Rueca |
| EPI_ISL_510535 | Molecular Virology, Instituto Carlos Chagas / Fiocruz Paraná | Universidade Federal do Parana (UFPR) | Duarte dos Santos, C.; Raboni, S.; Suzukawa, A.; Tscha, M.; Zanluca, C. |
| EPI_ISL_653922, EPI_ISL_660436, EPI_ISL_732664 | Molecular diagnostic laboratory of Federal Budget Institution of Science "Central Research Institute of Epidemiology" of The Federal Service on Customers' Rights Protection and Human Well-being Surveillance | Group of Genomics and Postgenomic Technologies of Central Research Institute of Epidemilology | Akimkin VG; Bulanenko VP; Dudorova A.V.; Dudorova AV; Kaptelova VV; Korneenko EV; Saenko SS; Samoilov AE; Shipulina OY; Speranskaya AS; Tivanova EV; Valdokhina AV |
| EPI_ISL_614347, EPI_ISL_614348, EPI_ISL_614351, EPI_ISL_614357, EPI_ISL_614372, EPI_ISL_614376, EPI_ISL_614379, EPI_ISL_614381, EPI_ISL_614386, EPI_ISL_614391, EPI_ISL_614393, EPI_ISL_681834, EPI_ISL_681840 | | | |
| see above | Molecular diagnostic unit for viral haemorrhagic fevers and emerging viruses, Bouaké CHU Laboratory | Project group Epidemology of Highly Pathogenic Microorganisms, Robert Koch-Institute | Adjaratou Traoré; Bamba Fatoumata Touré; Chantal Akoua-Koffi; Coulibaly Mbegnan; Diané Bamourou; Essia Belarbi; Etilé Anoh; Fabian Leendertz; Grit Schubert; Kra Ouffoué; Monemo Pacome; Safiatou Karidioula; Soundélé Maïté |
| EPI_ISL_426889, EPI_ISL_426892 | Motol University Hospital | Institute of Applied Biotechnologies a.s. | Adam Novotný; Jan Geryk; Kateřina Kvapilová; Martin Kašný; Milan Macek; Pavel Dřevínek; Petr Brož; Petr Klempt; Petr Kvapil |
| EPI_ISL_751585 | NE Public Health Laboratory | Genomics and Discovery, Respiratory Viruses Branch, Division of Viral Diseases, Centers for Disease Control and Prevention | Anna Montmayeur; Anna Uehara; Clinton R. Paden; Haibin Wang; Jing Zhang; Justin Lee; Krista Queen; Mili Sheth; Peter W. Cook; Rachel Marine; Suxiang Tong; Yan Li; Ying Tao |
| EPI_ISL_605894 | NGS Lab, DNA SOLUTION LTD. | NGS Lab, DNA SOLUTION LTD. | Chowdhury, M.; H.U.; Haider; Hasan; K.N.; Khaleque, A.; Khan; Khan, M.; M.F.A.; M.H.; M.I.; M.N.I.; Polol; Rabbi; Rahman, M.; Razu; Sufian, A. |
| EPI_ISL_411952 | NHC Key laboratory of Enteric Pathogenic Microbiology, Institute of Pathogenic Microbiology | Jiangsu Provincial Center for Disease Control & Prevention | Baoli Zhu; Bin Wu; Fengcai Zhu; Kangchen Zhao; Lunbiao Cui; Ming Wu; Tao Wu; Xiaojuan Zhu; Yin Chen; Yiyue Ge |
| EPI_ISL_467441, EPI_ISL_482710, EPI_ISL_487313, EPI_ISL_487314, EPI_ISL_509294, EPI_ISL_509357, EPI_ISL_515711, EPI_ISL_529739, EPI_ISL_535396, EPI_ISL_535551, EPI_ISL_602660, EPI_ISL_602752, EPI_ISL_602803, EPI_ISL_602911, EPI_ISL_602930, EPI_ISL_660220, EPI_ISL_660237, EPI_ISL_660650, EPI_ISL_678610, EPI_ISL_736993 | | | |

| | | | |
|---|---|---|---|
| see above | NHLS-IALCH | KRISP, KZN Research Innovation and Sequencing Platform | Chimukangara B; ChimukangaraB; Giandhari J; Khan S; Lessells R; Mdlalose K; Pillay S; Tegally H; Wilkinson E; York D; de Oliveira T |
| EPI_ISL_459578, EPI_ISL_501635, EPI_ISL_532767, EPI_ISL_533256, EPI_ISL_534435, EPI_ISL_534599, EPI_ISL_534646 | | | |
| see above | NHSGGC West of Scotland Specialist Virology Centre / MRC-University of Glasgow Centre for Virus Research | Wellcome Sanger Institute for the COVID-19 Genomics UK (COG-UK) consortium | Alasdair MacLean; Alice Broos; Ana da Silva Filipe; Antonia Ho; Cordelia Langford; Daniel Mair; David K. Jackson; David L Robertson; Dominic Kwiatkowski; Elihu Aranday-Cortes; Emma Thomson and Alex Alderton; Ewan Harrison; Ian Johnston; James Shepherd; Jenna Nichols; John Sillitoe; John Sillitoe on behalf of the Wellcome Sanger Institute COVID-19 Surveillance Team (http://www.sanger.ac.uk/covid-team); Joseph Hughes; Kathy Li; Kathy Smollett; Kirstyn Brunker; Kyriaki Nomikou; Lily Tong; Marc Niebel; Natasha Jesudason; Natasha Johnson; Patawee Asamaphan; Rajiv Shah; Richard Orton; Roberto Amato; Rory Gunson; Sarah McDonald; Sonia Goncalves; Sreenu Vattipally; Stephen Carmichael; Yasmin Parr |
| EPI_ISL_418242, EPI_ISL_420037, EPI_ISL_766861, EPI_ISL_766863, EPI_ISL_766864, EPI_ISL_766865, EPI_ISL_766869 | | | |
| see above | NIC Viral Respiratory Unit - Institut Pasteur of Algeria | National Reference Center for Viruses of Respiratory Infections, Institut Pasteur, Paris | Angela Brisebarre; Etienne Simon-Lorière; Fawzi Derrar; Flora Donati; Marion Barbet; Maud Vanpeene; Mélanie Albert; Méline Bizard; Sylvie Behillil; Sylvie van der Werf; Vincent Enouf |
| EPI_ISL_452203, EPI_ISL_479493 | NIV Influenza | NIV Influenza | Potdar V |
| EPI_ISL_416742 | NRL for Influenza, Centrum Epidemiology and Microbiology of National Institute of Public Health, Czech Republic | Charite Universitaetsmedizin Berlin, Institute of Virology | Akexander Nagy; Barbara Muehlemann; Christian Drosten; Dusan Trnka; Helena Jirincova; Jaromira Vecerova; Jörn Beheim-Schwarzbach; Julia Schneider; Ludmila Novakova; Talitha Veith; Terry Jones; Victor M Corman |
| EPI_ISL_488162, EPI_ISL_488829 | NU-OMICS DNA Sequencing research facility, Northumbria University | Wellcome Sanger Institute for the COVID-19 Genomics UK (COG-UK) consortium | Andrew Nelson; Brendan Payne; Chris Duncan; Clive Graham; Cordelia Langford; Darren Smith and Alex Alderton; David K. Jackson; Debra Padgett; Dominic Kwiatkowski; Edward Barton; Emma Swindells; Ewan Harrison; Garren Scott; Gary Black; Gary Eltringham; Greg Young; Ian Johnston; Jane Greenaway; Jennifer Collins; John Allan; John Sillitoe on behalf of the Wellcome Sanger Institute COVID-19 Surveillance Team (http://www.sanger.ac.uk/covid-team); Joshua Loh; Lynn Dover; Matthew Bashton; Paul Baker; Roberto Amato; Sarah Essex; Sheia Waugh; Shirelle Burton-Fanning; Sonia Goncalves; Steve Ligget; Wen Yew; Yusri Taha |
| EPI_ISL_456063 | NYU Langone Health | Departments of Pathology and Medicine, New York University School of Medicine | Adriana Heguy; Alireza Khodadadi-Jamayran; Amy Rapkiewicz; Andre M. Ribeiro-dos-Santos; Andrew Lytle; Antonio Serrano; Brendan Belovarac; Christian Marier; Dacia Dimartino; Emily Guzman; Emily Huang; Gael Westby; George Jour; Guomiao Shen; Iman Osman; Jared Pinnell; John Cadley; John Chen; Lawrence H. Lin; Ludovic Boytard; Margaret Black; Maria Aguero-Rosenfeld; Marie Samanovic-Golden; Mark J. Mulligan; Matija Snuderl; Matthew T. Maurano; Megan Hogan; Nick Vulpescu; Paolo Cotzia; Paul Zappile; Peter Meyn; Raquel Ordonez Ciriza; Raven Luther; Sitharam Ramaswami; Tatyana Gindin; Theodore Vougiouklakis; Vanessa Raabe; Xiaojun Feng; Yutong Zhang |
| EPI_ISL_480051, EPI_ISL_480062 | Nagoya City Public Health Research Institute | Pathogen Genomics Center, National Institute of Infectious Diseases | Hajime Kamiya; Kentaro Itokawa; Makoto Kuroda; Masanori Hashino; Motoi Suzuki; Rina Tanaka; Shinichiro Shibata; Takuya Miki; Tsuyoshi Sekizuka |
| EPI_ISL_523962 | National Agency for Public Health, Republic of Moldova | Charite Universitatsmedizin Berlin, Institute of Virology | Barbara Mühlemann; Christian Drosten; Elizabeta Jancheska; Golubinka Bosevska; Joern Beheim-Schwarzbach; Julia Schneider; Maja Kuzmanovska; Talitha Veith; Terry Jones; Victor M Corman |
| EPI_ISL_454571 | National Center of Expertise | National Center for Expertise, National Center for Biotechnology, Kazakhstan | ; Abdaliyev Askar; Akhmetollayev Ilyas; Amirgazin Asylulan; Aushakhmetova Zabira; Kalendar Ruslan; Lutsay Viktoriya; Rakhmetova Akbota; Ramankulov Yerlan; Shevtsov Alexandr |
| EPI_ISL_636757 | National Centre for Disease control (NCDC) | NCDC/CSIR-IGIB | Ajit Shewale; Anurag Agrawal#; Aparna Swaminathan; Asangla Kamai; Bharathram Uppili2*; Ishtaq Ahmed; Mahesh S. Dhar1*; Manish Kumar; Mohammed Faruq#; Nishu Tyagi; Partha Rakshit#; Pooja Sharma2*; RadhaKrishnan VS; Robin Marwal1*; Sandhya Kabra; Saruchi Wadhwa; Shaista Khan; Simmi Tiwari; Sujeet Singh; Tushar Nale; Vivekanand A |
| EPI_ISL_435084, EPI_ISL_435102 | National Centre for Disease control (NCDC), CSIR-Institute of Genomics and Integrative Biology (CSIR-IGIB) | NCDC/CSIR-IGIB | Aarti Tewari; Anurag Agrawal; Bharathram Uppili; Bibhash Nandi; Debasis Dash; Dhirendra Kumar; Hema Gogia; Hemlata Lall; Himanshu Vashisht; Mahesh Dhar; Manju Bala; Meena Datta; Mitali Mukerji; Mohammed Faruq; Nidhi Saini; Nishu Tyagi; Partha Rakshit; Pooja Sharma; Poonam Gupta; Pramod Kumar; Prateek Singh; Preeti Madan; Priyanka Singh; Rajesh Pandey; Sandhya Kabra; Saruchi Wadhwa; Satyabrata Bag; Simrita Singh; Sujeet Singh; Uma Sharma; Varun Jaiswal; Vivekanand A |
| EPI_ISL_420099 | National Centre for Infectious Diseases | Programme in Emerging Infectious Diseases, Duke-NUS Medical School | Barnaby E Young; Danielle E Anderson; David CB Lye; Gavin JD Smith; Jayanthi Jayakumar; Martin Linster; Yan Zhuang; Yee Sin Leo; Yvonne CF Su |
| EPI_ISL_560386 | National Health Laboratory | Botswana Institute for Technology Research and innovation | Dineo Emang Tshiamo. Gape Nyepetsi; Kefentse Arnold Tumedi; Madisa Mine; Maitshwarelo Ignatius Matsheka; Thongbotho Mphoyakgosi |
| EPI_ISL_464112, EPI_ISL_464138 | National Health Laboratory Service (NHLS), Tygerberg | Division of Medical Virology, Stellenbosch University and National Health Laboratory Service (NHLS) | Bronwyn Kleinhans; Eduan Wilkindon; Gert van Zyl; Houriiyah Tegally; Kayla Delaney; Susan Engelbrecht; Tulio de Oliveira; Wolfgang Preiser |
| EPI_ISL_435305, EPI_ISL_455707, EPI_ISL_455709 | National Hospital of Tropical Diseases | Oxford University Clinical Research Unit, Hanoi, Vietnam | H. Rogier van Doorn; Le Nguyen Minh Hoa; Nguyen Thi Ngoc Diep; Nguyen Thi Tam; Nguyen Thu Trang; Pham Ngoc Thach; Van Dinh Trang; on behalf of the OUCRU COVID-19 research group |
| EPI_ISL_416028, EPI_ISL_416031, EPI_ISL_416032, EPI_ISL_416035, EPI_ISL_416036, EPI_ISL_515525 | National Influenza Center - Instituto Adolfo Lutz | Instituto Adolfo Lutz, Interdiciplinary Procedures Center, Strategic Laboratory | Adriana Bugno; Adriano Abbud; Carlos Henrique Camargo; Claudia Regina Gonçalves; Claudio Tavares Sacchi; Daniela Bernardes Borges da Silva; Erica Valessa Ramos Gomes; Fabiana Cristina Pereira dos Santos; Maria do Carmo Sampaio Tavares Timenetsky; Simone Guadagnucci Morillo; Terezinha Maria de Paiva |
| EPI_ISL_420551, EPI_ISL_421668 | National Influenza Center, Indian Council of Medical Research - National Institute of Virology | Indian Council of Medical Research-National Institute of Virology, Microbial Containment Complex | Amita Jain; Anita M. Shete; Anita Shete-Aich; Atanu Basu; Bharati Malhotra; Bharti Malhotra; Deepika Chaudhary; Dimpal A. Nyayanit; Gajanan Sapkal; Gururaj Deshpande; Hitesh Dighe; Lalit Dar; M Choudhary; Manohar. L. Chaudhary; Padinjaremattathil Thankappan Ullas; Pragya D. Yadav; Pragya D. Yadav. Savita Patil; Pranita Gawande; Prasad Sarkale; Priya Abraham; Sarah Cherian; Savita Patil; Sreelekshmy Mohandas; Triparna Majumdar; Varsha Potdar |
| EPI_ISL_416429 | National Influenza Center, National Institute of Hygiene and Epidemiology (NIHE) | National Influenza Center, National Institute of Hygiene and Epidemiology (NIHE) | Dang Duc Anh; Futoshi Hasebe; Hoang Vu Mai Phuong; Kouichi Morita; Le Quynh Mai; Le Thi Thanh; Meng Ling Moi; Nguyen Le Khanh Hang; Nguyen Phuong Anh; Nguyen Vu Son; Pham Hong Quynh Anh; Pham Thi Hien; Taichiro Takemura; Takeshi Nabeshima; Tran Thu Huong; Ung Thi Hong Trang; Vuong Duc Cuong |
| EPI_ISL_467778, EPI_ISL_467780 | National Influenza Centre Romania | Charite Universitatsmedizin Berlin, Institute of Virology | Barbara Muehlemann; Christian Drosten; Jorn Beheim-Schwarzbach; Julia Schneider; L. Ustea; M. Lazar; N. Paraschiv; Talitha Veith; Terry Jones; Victor M Corman |
| EPI_ISL_410301 | National Influenza Centre, National Public Health Laboratory, Kathmandu, Nepal | The University of Hong Kong | Alfonso J. Rodriguez-Morales; Anup Bastola; Basu Dev Pandey; Bibek Kumar Lal; Daniel Chu; Haogao Gu; Leo Poon; Malik Peiris; Ranjit Sah; Runa Jha |
| EPI_ISL_402125 | National Institute for Communicable Disease Control and Prevention (ICDC) Chinese Center for Disease Control and Prevention (China CDC) | National Institute for Communicable Disease Control and Prevention (ICDC) Chinese Center for Disease Control and Prevention (China CDC) | Chen; Dai; F.-H.; Hu, Y.; J.-H.; J.-J.; J.-L. and Zhu; Liu, Y.; Pei; Q.-M.; She; Song; T.-Y.; Tao; Tian; Wang; Wang, W.; Wu, F.; Xu, L.; Y.-L.; Y.-M.; Y.-Y.; Y.-Z.; Yu, B.; Z.-G.; Z.-W.; Zhang; Zhao, S.; Zheng |
| EPI_ISL_417186 | National Institute for Communicable Diseases of the National Health Laboratory Service | National Institute for Communicable Diseases of the National Health Laboratory Service | A; Allam M; Bhiman JN; Ismail A; Khumalo Z; Kwenda S; Mohale T; Subramoney K; van Heusden P; von Gottberg |
| EPI_ISL_721625, EPI_ISL_728280, EPI_ISL_728285, EPI_ISL_746835 | National Institute for Infectious Diseases, INMI, "L. Spallanzani" IRCCS | National Institute for Infectious Diseases, INMI, "L. Spallanzani" IRCCS | A Di Caro; B Bartolini; C.E.M Gruber; E Giombini; F Messina; M Rueca; MR Capobianchi |
| EPI_ISL_469254 | National Institute for Viral Disease Control and Prevention, China CDC | Institute of Viral Disease Control and Prevention, China CDC | Chun Huang; Dayan Wang; George Fu Gao; Guizhen Wu; Li Zhao; Lijuan Chen; Peihua Niu , Baoying Huang; Roujian Lu; Wenbo Xu; Wenjie Tan; Wenling Wang; Yubai Bi |
| EPI_ISL_408480, EPI_ISL_408482, EPI_ISL_408484, EPI_ISL_408485, EPI_ISL_408486, EPI_ISL_408488 | National Institute for Viral Disease Control and Prevention, China CDC | National Institute for Viral Disease Control & Prevention, CCDC | Baoying Huang; Fei Ye; George F. Gao; Guizhen Wu; Huiping Yang; Jianan Xu; Jianxiong Li; Kangchen Zhao; Li Zhao; Peihua Niu; Quanyi Wang; Roujian Lu; Shenjiao Wang; Ti Liu; Wenbo Xu; Wenjie Tan; Wenjie Tan , Xiaoqing Fu , Xiang Zhao , Wenling Wang , Peihua Niu , Roujian Lu; Wenling Wang; Xiang Zhao; Yang Pan; Yanhong Sun , Baoying Huang , Li Zhao , Fei Ye , Wenbo Xu , George F. Gao , Guizhen Wu; Yong Shi; Zhaoguo Wang |
| EPI_ISL_402120, EPI_ISL_469256, EPI_ISL_498693, EPI_ISL_575330, EPI_ISL_591272, EPI_ISL_591274, EPI_ISL_591280 | | | |
| see above | National Institute for Viral Disease Control and Prevention, China CDC | National Institute for Viral Disease Control and Prevention, China CDC | Cao Chen; Chun Huang; Dayan Wang; Dong Xia; Geogia Fu Gao; George F. Gao; George F.Gao; Guizhen Wu; Haijun Du; Hong Wang; Huilai Ma; Ji Wang; Jingdong Song; Juan Song; Jun Han; Kang Xiao; LingLing Mao; Qinqin Song; Rongbao Gao; Ruqin Gao; Shiwen Wang; Shuai Pang; Weifeng Shi; WenQing Yao , Wenbo Xu; Wenbo Xu; Wenjie Tan , Xiang Zhao , Wenling Wang , Xuejun Ma , Yongzhong Jiang , Roujian Lu , Ji Wang , Weimin Zhou , Peihua Niu , Peipei Liu , Faxian Zhan , Weifeng Shi , Baoying Huang , Jun Liu , Li Zhao , Yao Meng , Xiaozhou He , Fei Ye , Na Zhu , Yang Li , Jing Chen , Wenbo Xu , George F. Gao , Guizhen Wu; William J. Liu; Xiang Zhao; Xiang Zhao , Lijuan Chen; Xuancheng Lu; Yang Song; Yanhai Wang; Yao Meng; Yao Meng , Zhixiao Chen , Yuchao Wu; Yong Zhang; Yong Zhang , Bo Zhijian , Jianqun Zhang; Yuan He; Yuchao Wu; Zhaoguo Wang; ZhiQiang Xia; Zhixiao Chen |
| EPI_ISL_708839 | National Institute of Blood Diseases (NIBD), Molecular Biology Lab | Genomics Lab NIBD | Aneeta Shahni; Arshi Naz; Gul Sufaida; Samina Naz Mukry; Sayed Ali Raza; Shariq Ahmed; Tahir Sultan Shamsi |
| EPI_ISL_538504, EPI_ISL_538508, EPI_ISL_574433, EPI_ISL_576386, EPI_ISL_747234 | National Institute of Health Research and Development | National Institute of Health Research and Development | A; AA; Adam; Agustiningsih; D; F; Febriyani; Febriyanti; HA; HD; Hariastuti; Herman; Herna; IL; Ikawati; Indalau; Indrasari; J; K; KD; KNA; Kipuw; Kurniawati; M; Monika; Muna; Mursinah; N; ND; NI; NK; NL; Nikmah; Nugraha; Nurhadi; Paisal; Pangesti; Pawestri; Puspa; Puspandari; Puspandary; R; Ramadhany; Rizki; Setiawaty; Setiawaty, V.; Soekarso; Subangkit; Susanti; Susilarini; T; UA; V; Vivi; Wibowo; Wulandari |
| EPI_ISL_462093, EPI_ISL_466649, EPI_ISL_475167, EPI_ISL_480449, EPI_ISL_514248, EPI_ISL_603222, EPI_ISL_603247 | | | |
| see above | National Institute of Laboratory Medicine and Referral Center | Genomic Research Lab, BCSIR | A. K. M. Shamsuzzaman; Abu Sayeed Mohammad Mahmud; Arifa Akram; Asish Kumar Ghosh; Barna Goswami; Bayzid Bin Monir; Eshrar Osman; Iffat Jahan; Mahmuda Yeasmin; Md. Ahasan Habib; Md. Ahashan Habib; Md. Maruf Ahmed Molla; Md. Masum Hossain Arif; Md. Murshed Hasan Sarkar; Md. Murshed Hasan Sarker; Md. Saddam Hossain; Md. Salim Khan; Mohammad Samir Uzzaman; Monira Parveen; Salek Ahmed Sajib; Shahina Akter; Shahjahan Siddike; Sheikh Md. Selim Al Din; Tanjina Akhter Banu; Tanjina Akhter Banu; Tasnim Nafisa; Utpal Chandra Ray |
| EPI_ISL_539779, EPI_ISL_539780 | National Institute of Public Health (Czech Republic) | State Veterinary Institute Prague | A; D; H; Jirincova; L; Nagy; Novakova; Trnka; Vecerova, J. |
| EPI_ISL_434555 | National Institutes of Health, University of the Philippines Manila | Philippine Genome Center | Alessandra C. Sanchez; Benedict A. Maralit; Bernard Demot; Carlo M. Lapid; Christina Tan; El King D. Morado; Eva Maria Cutiongco-de la Paz; Francis A. Tablizo; Haifa L.Gaza; Jan Michael C. Yap; Jarvin E. Nipales; Jo-Hannah S. Llames; John Mark Velasco; Joshua Gregor A. Dizon; Joy Ann Petronio-Santos; Julius Aaron Mejia; Kris P. Punayan; Kristianne Arielle D. Gabriel; Maribell Dollete; Marissa M. Alejandria; Raul V. Destura; Shana F. Genavia; Shebna Rose D. Fabilloren; Shiela Mae M. Araiza; Sonia Salamat; and Cynthia P. Saloma |
| EPI_ISL_449799 | National Laboratory for Health, Environment and Food | National Laboratory for Health, Environment and Food | Blazun Vosner H.; Duh D.; Hedzet S.; Janezic S.; Mahnic A.; Rupnik M.; Zavrsnik J. |
| EPI_ISL_512609, EPI_ISL_512617, EPI_ISL_512628, EPI_ISL_512629, EPI_ISL_512636, EPI_ISL_512640, EPI_ISL_512810 | | | |
| see above | National Laboratory for Influenza/Virology reference | Respiratory Virus Unit, Microbiology Services Colindale, Public Health England | Dr. Iryna Demchyshyna; PHE Covid Sequencing Team |

| | | | |
|---|---|---|---|
| | laboratory, Public Health Center of the Ministry of Health of Ukraine | | |
| EPI_ISL_671417, EPI_ISL_671418 | National Laboratory of Virology, Szentágothai Research Centre | National Laboratory of Virology, Szentágothai Research Centre | Balázs Somogyi; Brigitta; Endre Gábor Tóth; Ferenc Jakab; Gábor Kemenesi |
| EPI_ISL_644789, EPI_ISL_644801, EPI_ISL_644821, EPI_ISL_647978 | National Microbiology Reference Laboratory | Quadram Institute Bioscience | Agnes Juru; Alexander Goredema; Ana-Victoria Gutierrez; Andrew J. Page; Andrew Tarupiwa; Barbra Murwira; Beuty Makamure; Charles Nyagupe; David Baker; Gaetan Thilliez; Gemma Kay; Gibson Mhlanga; Hlanai Gumbo; Isaac Phiri; Justin O'Grady; Leonardo de Oliveira Martins; Muchaneta Mugabe; Portia Manangazira; Robert Kingsley; Sekesai Zinyowera; Tapfumanei Mashe; Tatenda Takawira; Thanh Le Viet |
| EPI_ISL_457833, EPI_ISL_457843 | National Public Health Laboratory | KEMRI-Wellcome Trust Research Programme/KEMRI-CGMR-C Kilifi | Githinji G. et al 2020 |
| EPI_ISL_416829, EPI_ISL_416885 | National Public Health Laboratory | Malaysia Genome Institute | Azrin Ahmad; Enizza Kasim; Hani Mat Hussin; Irni Suhayu Sapian; Mohd Faizal Abu Bakar; Mohd Noor Mat Isa; Noorliza Mohamad Noordin; Nor Azfa Johari; Norazimah Tajudin; Nurhezreen Md Iqbal; Rehan Shuhada Abu Bakar; Selvanesan Sengol; Shahrul Hisham Zainal Ariffin; Shamsidar Sopie; Siti Noraini Othman; W Afiza W Mohd Arifin; Yu Kie Chem; Yusuf Muhammad Noor |
| EPI_ISL_410719 | National Public Health Laboratory | National Public Health Laboratory | Cui L; Lin RTP; Mak TM; Octavia S |
| EPI_ISL_418996, EPI_ISL_419000, EPI_ISL_428828, EPI_ISL_428836, EPI_ISL_462287, EPI_ISL_462433, EPI_ISL_479578, EPI_ISL_490049, EPI_ISL_490068, EPI_ISL_498595, EPI_ISL_498604, EPI_ISL_512837, EPI_ISL_512841, EPI_ISL_516818, EPI_ISL_516824, EPI_ISL_527364, EPI_ISL_527379, EPI_ISL_536420, EPI_ISL_536446, EPI_ISL_548973, EPI_ISL_574519, EPI_ISL_596462, EPI_ISL_596474, EPI_ISL_596492, EPI_ISL_626631, EPI_ISL_626633, EPI_ISL_645115, EPI_ISL_645124, EPI_ISL_654816, EPI_ISL_693313, EPI_ISL_693325, EPI_ISL_728181, EPI_ISL_754092, EPI_ISL_754096, EPI_ISL_768625 | | | |
| see above | National Public Health Laboratory, National Centre for Infectious Diseases | National Public Health Laboratory, National Centre for Infectious Diseases | Chavatte JM; Cui L; Lin Cui; Lin RTP; Mak TM; Octavia S; Raymond Tzer Pin Lin; Sophie Octavia; Tze Minn Mak; Zhenyang Zhou; Zhou Z |
| EPI_ISL_480224, EPI_ISL_480297, EPI_ISL_480300, EPI_ISL_480302, EPI_ISL_480304, EPI_ISL_480305, EPI_ISL_480307, EPI_ISL_480309, EPI_ISL_480310 | | | |
| see above | National Reference Laboratory "Influenza and acute respiratory diseases" | NRL-HIV | Ivailo Alexiev; Ivan Ivanov; Ivva Philipova |
| EPI_ISL_435677, EPI_ISL_443187 | National Virology Reference Laboratory | National Public Health Laboratory, National Centre for Infectious Diseases | Chavatte Jean-Marc; Cui Lin; Lin Raymond Tzer Pin; Mak Tze Minn; Octavia Sophie; Taib Surita; Zaini Zainun |
| EPI_ISL_605076 | National Virus Reference Laboratory | Irish Coronavirus Sequencing Consortium - Helixworks | Conor Crosbie; Nimesh Pinnamaneni; Sachin Chalapati |
| EPI_ISL_671339, EPI_ISL_671350, EPI_ISL_671397, EPI_ISL_681906, EPI_ISL_682057, EPI_ISL_767724, EPI_ISL_767792, EPI_ISL_767793, EPI_ISL_767814, EPI_ISL_791297, EPI_ISL_791314 | | | |
| see above | National Virus Reference Laboratory | Irish Coronavirus Sequencing Consortium - Teagasc Moorepark | Alejandro Abner Garcia Leon; Calm Walsh; Calum Walsh; Fiona Crispie; Genuity Ireland; John Kenny; Matthew McCabe; Paddy Mallon; Paul Cotter |
| EPI_ISL_500577, EPI_ISL_500595, EPI_ISL_512087, EPI_ISL_525352, EPI_ISL_525376, EPI_ISL_528445, EPI_ISL_578258, EPI_ISL_578294, EPI_ISL_578325, EPI_ISL_596903, EPI_ISL_639863, EPI_ISL_639865, EPI_ISL_671864, EPI_ISL_671913, EPI_ISL_732496, EPI_ISL_732505, EPI_ISL_732522, EPI_ISL_752514, EPI_ISL_752536 | | | |
| see above | National Virus Reference Laboratory | National Virus Reference Laboratory | Aditi Chaturvedi; Cillian F De Gascun; Daniel Hare; Gabriel Gonzalez; Jonathan Dean; Michael Carr; Suzie Coughlan |
| EPI_ISL_444998, EPI_ISL_444999 | Naval Health Research Center | Naval Medical Research Center Biological Defense Research Directorate | Adrian Paskey; Chris Myers; Dessiree Pena-Gomez; Ewell Hollis; Kimberly Bishop-Lilly; Kyle Long; Logan Voegtly; Melinda Balansay-Ames; Nathaniel Christy; Regina Cer; Roger Pan |
| EPI_ISL_515398 | Nevada State Public Health Laboratory | Nevada State Public Health Laboratory | Andrew Gorzalski; Chris Laverdure; Cyprian Rossetto; David Jackson; Heather Kerwin; Joel R. Sevinsky; Natalie Crawford; Paul Hartley; Richard Tillett; Stephanie Van Hooser; Subhash C. Verma; and Mark Pandori |
| EPI_ISL_516934 | Nicolae Testemitanu State University of Medicine and Pharmacy | International Centre for Genetic Engineering and Biotechnology (ICGEB) and ARGO Open Lab Platform for Genome Sequencing | Dal Monego S; Licastro D; Marcello A; Rajasekharan S; Ulinici M |
| EPI_ISL_455362, EPI_ISL_487091, EPI_ISL_487103, EPI_ISL_487106, EPI_ISL_527873, EPI_ISL_527874, EPI_ISL_527880, EPI_ISL_527889, EPI_ISL_527891, EPI_ISL_527892, EPI_ISL_729924, EPI_ISL_729929, EPI_ISL_729938, EPI_ISL_729946, EPI_ISL_729962, EPI_ISL_729964, EPI_ISL_729973, EPI_ISL_729976, EPI_ISL_729977, EPI_ISL_729978, EPI_ISL_729980, EPI_ISL_729982, EPI_ISL_729985, EPI_ISL_729993, EPI_ISL_729994, EPI_ISL_730034, EPI_ISL_730042 | | | |
| see above | Nigeria Centre for Disease Control (NCDC) | African Centre of Excellence for Genomics of Infectious Diseases (ACEGID), Redeemer's University, Ede, Osun State, Nigeria | Ajogbasile F.V.; Folarin O.A.; Happi C.T.; Ihekweazu C.; Kayode A.; Oguzie J.; Olawoye I.; Olumade T.; Oluniyi P.E.; Oluniyi P.E. et al; Uwanibe J. |
| EPI_ISL_479923 | Niigata City Public Health Research Institute | Pathogen Genomics Center, National Institute of Infectious Diseases | Hajime Kamiya; Kentaro Itokawa; Makoto Kuroda; Masanori Hashino; Motoi Suzuki; Rina Tanaka; Tsuyoshi Sekizuka; Yurie Takahashi |
| EPI_ISL_668431, EPI_ISL_796694 | Nordland Hospital - Bodo, Laboratory Department, Molecular Biology Unit | Norwegian Institute of Public Health, Department of Virology | Atiya R Ali; Hilde Elshaug; Hilde Vollan; Kamilla Heddeland Instefjord; Karoline Bragstad; Kathrine Stene-Johansen; Marie Paulsen Madsen; Olav Hungnes; Rasmus Riis Kopperud |
| EPI_ISL_548048, EPI_ISL_579406 | North Shore Hospital | Institute of Environmental Science and Research (ESR) | Anja Werno; Antje van der Linden; Arlo Upton; Chris Mansell; David Hammer; Dragana Drinkovic; Erasmus Smit; Gary McAuliffe; Hana Sofia Andersson; Hermes Perez; James Ussher; Jill Sherwood; Jing Wang; Joep de Ligt; Josh Freeman; Julia Howard; Juliet Elvy; Lauren Jelly; Mary DeAlmeida; Matt Blakiston; Matt Storey; Matthew Rogers; Max Bloomfield; Michael Addidle; Michelle Balm; Muhammad Faisal; Nikki Freed; Olin Silander; Sally Roberts; Sarah Jefferies; Sharmini Muttaiyah; Susan Morpeth; Susan Taylor; Timothy Blackmore; Vani Sathyendran; Veronica Playle; Virginia Hope; Xiaoyun Ren |
| EPI_ISL_493496 | Northumbria University / South Tees Hospitals NHS Foundation Trust / North Cumbria Integrated Care NHS Foundation Trust / North Tees and Hartlepool NHS Foundation Trust / Newcastle Hospitals NHS Foundation Trust | COVID-19 Genomics UK (COG-UK) Consortium | Andrew Nelson; Brendan Payne; Clive Graham; Darren L Smith; Debra Padgett; Edward Barton; Emma Swindells; Garren Scott; Gary Black; Gary Eltringham; Giles S Holt; Greg R Young; Jane Greenaway; Jennifer Collins; John Allan; Joshua Loh; Lynn Dover; Matthew Bashton; Mohammad A Tariq; Paul Baker; Sarah Essex; Steve Liggett; Wen C Yew; Yusri Taha |
| EPI_ISL_775356, EPI_ISL_796675 | Norwegian Institute of Public Health, Department of Virology | Norwegian Institute of Public Health, Department of Virology | Atiya R Ali; Hilde Elshaug; Hilde Vollan; Kamilla Heddeland Instefjord; Karoline Bragstad; Kathrine Stene-Johansen; Marie Paulsen Madsen; Olav Hungnes; Rasmus Riis Kopperud |
| EPI_ISL_534326 | Notre Dame Intermedica Saude AS | Instituto Adolfo Lutz, Interdiciplinary Procedures Center, Strategic Laboratory | Claudia Regina Gonçalves; Claudio Tavares Sacchi; Erica Valessa Ramos Gomes |
| EPI_ISL_735438 | Nuclei Acid Testing - Rwanda National Reference Laboratory | GIGA Medical Genomics | Bouchra Boujemla; Esperence Umumararungu; Jacob Souopgui; Keith Durkin; Léon Mutesa; Maria Artesi; Marie-Pierre Hayette; Patrick Tuyisenge; Robert Rutayisire; Sabin Nsanzimana; Swaibu Gatare; Sébastien Bontems; Vincent Bours; Yvan Butera |
| EPI_ISL_735448 | Nucleic Acid Testing - Rwanda National Reference Laboratory | GIGA Medical Genomics | Bouchra Boujemla; Esperence Umumararungu; Jacob Souopgui; Keith Durkin; Léon Mutesa; Maria Artesi; Marie-Pierre Hayette; Patrick Tuyisenge; Robert Rutayisire; Sabin Nsanzimana; Swaibu Gatare; Sébastien Bontems; Vincent Bours; Yvan Butera |
| EPI_ISL_700486 | Nyanga CDC wc NGC | NHLS/UCT | Arash Iranzadeh; Bruna Galvao; Carolyn Williamson; Deelan Doolabh; Diana Hardie; Innocent Mudau; Kruger Marais; Lynn Tyers; Marvin Hsiao; Stephen Korsman |
| EPI_ISL_512846 | O.I.J. MORGUE JUDICIAL | Inciensa, Instituto Costarricense de Investigación y Enseñanza en Nutrición y Salud | Adriana Godinez & Melany Calderon; Claudio Soto-Garita; Estela Cordero; Francisco Duarte; Hebleen Porras |
| EPI_ISL_534316 | OS Mun Santana Lauro Ribas Braga | Instituto Adolfo Lutz, Interdiciplinary Procedures Center, Strategic Laboratory | Claudia Regina Gonçalves; Claudio Tavares Sacchi; Erica Valessa Ramos Gomes |
| EPI_ISL_498192 | OUCRU | OUCRU | Dinh Nguyen Huy Man; Guy Thwaites; Lam Anh Nguyet; Lam Minh Yen; Le Manh Hung; Le Nguyen Truc Nhu; Nghiem My Ngoc; Ngo Ngoc Quang Minh; Nguyen Tan Binh; Nguyen Thanh Dung; Nguyen Thanh Phong; Nguyen Thanh Truong; Nguyen Thi Han Ny; Nguyen Thi Thu Hong; Nguyen Tri Dung; Nguyen Van Vinh Chau; Tang Chi Thuong; Tran Chanh Xuan; Tran Nguyen Hoang Tu; Tran Tan Thanh; Tran Tinh Hien; Vo Thanh Lam; and Le Van Tan; for OUCRU COVID-19 research group* |
| EPI_ISL_450743 | OUCRU/HTD | OUCRU/HTD | Dinh Nguyen Huy Man; Guy Thwaites; Lam Anh Nguyet; Lam Minh Yen; Le Manh Hung; Le Nguyen Truc Nhu; Nghiem My Ngoc; Ngo Ngoc Quang Minh; Nguyen Tan Binh; Nguyen Thanh Dung; Nguyen Thanh Phong; Nguyen Thanh Truong; Nguyen Thi Han Ny; Nguyen Thi Thu Hong; Nguyen Tri Dung; Nguyen Van Vinh Chau; Tang Chi Thuong; Tran Chanh Xuan; Tran Nguyen Hoang Tu; Tran Tan Thanh; Tran Tinh Hien; Vo Thanh Lam; and Le Van Tan; for OUCRU COVID-19 research group* |
| EPI_ISL_458117, EPI_ISL_458118 | Oman National Influenza Centre | Department of Microbiology and Immunology-SQUH | Abdulla Balkhair; Ahlam Al-Amri; Aisha Al-Amri; Aisha Al-Busaidi; Amina Al Jardani; Fahad Zadjali; Fatma BaAlawi; Hamida AL Barwani; Hanan Al-kindi; Intisar Al-Shukri; Khulood Al-Mammary; Mohammed Al-Tobi; Samiha Al Kharusi; Samira Al-Maruqi; Zeyana AL-Dahmani |
| EPI_ISL_457981, EPI_ISL_491978, EPI_ISL_491999, EPI_ISL_492007, EPI_ISL_492023, EPI_ISL_492026 | Oman-NIC | Department of Microbiology and Immunology-SQUH | Abdulla Balkhair; Ahlam Al-Amri; Aisha Al-Amri; Aisha Al-Busaidi; Amina Al Jardani; Fahad Zadjali; Fatma BaAlawi; Hamida AL Barwani; Hanan Al-kindi; Intisar Al-Shukri; Khulood Al-Mammary; Mohammed Al-Tobi; Samiha Al Kharusi; Samira Al-Maruqi; Zeyana AL-Dahmani |
| EPI_ISL_457702 | Oman-NIC | Microbiology laboratory- Sultan Qaboos University Hospital | Abdulla Balkhair; Ahlam Al-Amri; Aisha Al-Amri; Aisha Al-Busaidi; Amina Al Jardani; Fahad Zadjali; Fatma BaAlawi; Hamida AL Barwani; Hanan Al-kindi; Intisar Al-Shukri; Khulood Al-Mammary; Mohammed Al-Tobi; Samiha Al Kharusi; Samira Al-Maruqi; Zeyana AL-Dahmani |
| EPI_ISL_457701, EPI_ISL_457706, EPI_ISL_457939, EPI_ISL_457993, EPI_ISL_457997 | Oman-NIC | Oman-NIC | Abdulla Balkhair; Ahlam Al-Amri; Aisha Al-Amri; Aisha Al-Busaidi; Amina Al Jardani; Fahad Zadjali; Fatma BaAlawi; Hamida AL Barwani; Hanan Al-kindi; Intisar Al-Shukri; Khulood Al-Mammary; Mohammed Al-Tobi; Samiha Al Kharusi; Samira Al-Maruqi; Zeyana AL-Dahmani |
| EPI_ISL_491126, EPI_ISL_491137 | Oman-National Influenza Center | Biotechnology & OMICs Laboratory | Abdul Latif Khan; Adil Al-Wahaibi; Adil Khan; Ahlam Al-Amri; Ahmed Al-Harrasi; Ahmed Al-Rawahi; Aisha Al-Amri; Aisha Al-Busaidi; Amina Al-Jardani; Hanan Al-Kindi; Intisar Al-Shukri; Sajjad Asaf; Samiha Al-Kharusi; Samira Al-Mahruqi; Seif Al-Abri |
| EPI_ISL_492065 | Oman-National Influenza Center | Department of Microbiology and Immunology-SQUH Department of Microbiology and Immunology, Sultan Qaboos University Hopsital, P.O 35, Postal code 123 | Abdulla Balkhair; Amina Al Jardani; Azza Al-Rashdi; Fahad Zadjali; Fatma BaAlawi; Hamida AL Barwani; Hanan Al-kindi; Intisar Al-Shukri; Khulood Al-Mammary; Samiha Al Kharusi; Samira Al-Maruqi; Zeyana AL-Dahmani |
| EPI_ISL_569760, EPI_ISL_569763, EPI_ISL_569778, EPI_ISL_569791, EPI_ISL_569798, EPI_ISL_569836, EPI_ISL_639945, EPI_ISL_639947, EPI_ISL_733484, EPI_ISL_733490 | | | |
| see above | Omsk Research Institute of Natural Focal Infections | WHO National Influenza Centre Russian Federation | Aleksei Vasilenko; Andrey Komissarov; Anna Ivanova; Artem Fadeev; Daria Danilenko; Daria Nashatyreva; Dmitry Bazhenov; Dmitry Lioznov; Ekaterina Gradoboeva; Ekaterina Savkina; Elena Nabieva; Elena Poleshchuk; Georgii Bazykin; Ksenia Safina; Kseniya Komissarova; Valery Yakimenko |
| EPI_ISL_475134, EPI_ISL_582841, EPI_ISL_676513 | Orebro klinisk mikrobiologi | The Public Health Agency of Sweden | Anna Risberg; Anna-Malin Linde; Department of Microbiology; Karin Tegmark-Wisell; Maria Lind Karlberg; Mattias Haukland; Mia Brytting; Olov Svartstrom; Oskar Karlsson Lindsjo; Petra Edquist; Reza Advani; Sandra Broddesson; Shamam Muradrasoli; The Public Health Agency of Sweden |
| EPI_ISL_710586 | Orsa VC | The Public Health Agency of Sweden | Department of Microbiology; The Public Health Agency of Sweden |
| EPI_ISL_417484, EPI_ISL_471177, EPI_ISL_493355, EPI_ISL_549034, EPI_ISL_590908, EPI_ISL_590914, EPI_ISL_775416 | | | |

| | | | |
|---|---|---|---|
| see above | Oslo University Hospital, Department of Medical Microbiology | Norwegian Institute of Public Health, Department of Virology | Atiya R Ali; Hilde Elshaug; Hilde Synnøve Vollan; Hilde Vollan; Kamilla Heddeland Instefjord; Karoline Bragstad; Kathrine Stene-Johansen; Marie Paulsen Madsen; Olav Hungnes; Rasmus Riis Kopperud |
| EPI_ISL_788951 | Ospedale "Di Venere" | Beaconlab (Bioinformatics, Evolution and Comparative Genomics lab), Dept of Biosciences, University on Milan | Chiara M; Iacobellis M; Manzari C; Parisi A; Pesole G; Piluscio R; d'Avenia M |
| EPI_ISL_529020 | Ospedale Civile S. Liberatore-Atri | Istituto Zooprofilattico Sperimentale dell'Abruzzo e Molise "G.Caporale" | Ancora M; Cammà C; Curini V; Di Domenico M; Di Pasquale A; Lorusso A; Mangone I; Marcacci M; Puglia I; Rinaldi A; Savini G. |
| EPI_ISL_548248 | Ostersund klinisk mikrobiologi | The Public Health Agency of Sweden | Anna Risberg; Anna-Malin Linde; Karin Tegmark-Wisell; Maria Lind Karlberg; Mattias Haukland; Mia Brytting; Olov Svartstrom; Oskar Karlsson Lindsjo; Petra Edquist; Reza Advani; Sandra Broddesson |
| EPI_ISL_549093, EPI_ISL_591017, EPI_ISL_635116 | Ostfold Hospital Trust - Kalnes, Centre for Laboratory Medicine, Section for gene technology and infection serology | Norwegian Institute of Public Health, Department of Virology | Hilde Elshaug; Hilde Synnøve Vollan; Hilde Vollan; Kamilla Heddeland Instefjord; Karoline Bragstad; Kathrine Stene-Johansen; Marie Paulsen Madsen; Olav Hungnes; Rasmus Riis Kopperud |
| EPI_ISL_448504, EPI_ISL_559780, EPI_ISL_576757, EPI_ISL_595652, EPI_ISL_628442, EPI_ISL_628485, EPI_ISL_742248, EPI_ISL_764532, EPI_ISL_764539, EPI_ISL_764566 | | | |
| see above | Oxford Viromics, NDM, University of Oxford; Oxford University Hospitals; Basingstoke and North Hampshire Hospital | COVID-19 Genomics UK (COG-UK) Consortium | Alex Mobbs; Amy Trebes; Anita Justice; Catrin Moore; Christophe Fraser; David Bonsall; David Buck; Emma Wise; George Macintyre; Jessica Lynch; John Todd; Mariateresa de Cesare; Matilde Mori; Monique Andersson; Nathan Moore; Nick Cortes; Robert Shaw; Stephen Kidd; Tanya Golubchik; Timothy Peto |
| EPI_ISL_424885 | PA Department of Health, Bureau of Laboratories | Pathogen Discovery, Respiratory Viruses Branch, Division of Viral Diseases, Centers for Disease Control and Prevention | Alison S. Laufer Halpin; Anna Uehara; Christopher A. Elkins; Clinton R. Paden; Haibin Wang; Jing Zhang; Krista Queen; Mary S. Keckler; Rachel Marine; Suxiang Tong; Yan Li; Ying Tao |
| EPI_ISL_767897 | PALMS | NSW Health Pathology - Institute of Clinical Pathology and Medical Research; Westmead Hospital; University of Sydney | CIDM-PH et al. |
| EPI_ISL_443989, EPI_ISL_469531 | PHE South West Regional Laboratory, National Infection Service | Wellcome Sanger Institute for the COVID-19 Genomics UK (COG-UK) consortium | Barry Vipond; Cordelia Langford; David K. Jackson; Dominic Kwiatkowski; Dr Peter Muir; Ewan Harrison; Hannah Pymont; Ian Johnston; John Sillitoe on behalf of the Wellcome Sanger Institute COVID-19 Surveillance Team (http://www.sanger.ac.uk/covid-team); Rich Hopes; Roberto Amato; Sonia Goncalves; Stephanie Hutchings; and Alex Alderton |
| EPI_ISL_678257 | PHI Institute of Nephrology - Struga | Research Center for Genetic Engineering and Biotechnology "Georgi D. Efremov" , Macedonian Academy of Sciences and Arts | RCGEB - MASA |
| EPI_ISL_733570 | POK OI HOSPITAL | Hong Kong Department of Health | Alan K.L. Tsang; Dominic N.C. Tsang; Edman T.K. Lam; Peter C.W. Yip; Rickjason C.W. Chan |
| EPI_ISL_534322 | PS Mun Julio Tupy | Instituto Adolfo Lutz, Interdiciplinary Procedures Center, Strategic Laboratory | Claudia Regina Gonçalves; Claudio Tavares Sacchi; Erica Valessa Ramos Gomes |
| EPI_ISL_523973 | PS Municipal Dona Maria Antonieta Ferreira de Barros | Instituto Adolfo Lutz, Interdiciplinary Procedures Center, Strategic Laboratory | Claudia Regina Gonçalves; Claudio Tavares Sacchi; Erica Valessa Ramos Gomes |
| EPI_ISL_547570 | PS Municipal Dr Augusto Gomes de Mattos | Instituto Adolfo Lutz, Interdiciplinary Procedures Center, Strategic Laboratory | Claudia Regina Gonçalves; Claudio Tavares Sacchi; Erica Valessa Ramos Gomes; Karoline Rodrigues Campos |
| EPI_ISL_515523, EPI_ISL_515524, EPI_ISL_515566, EPI_ISL_524466, EPI_ISL_527866 | PS Municipal Dr Lauro Ribas Braga | Instituto Adolfo Lutz, Interdiciplinary Procedures Center, Strategic Laboratory | 01246-1301; 355 - Brazil; Av. Dr. Arnaldo; Cerqueira Cesar; Claudia Regina Gonçalves; Claudio Tavares Sacchi; Erica Valessa Ramos Gomes; São Paulo - SP |
| EPI_ISL_524465 | PS Municipal Dr. Caetano Virgílio Neto | Instituto Adolfo Lutz, Interdiciplinary Procedures Center, Strategic Laboratory | Claudia Regina Gonçalves; Claudio Tavares Sacchi; Erica Valessa Ramos Gomes |
| EPI_ISL_534321 | PS e Maternidade Nair Fonseca Leitao Arantes | Instituto Adolfo Lutz, Interdiciplinary Procedures Center, Strategic Laboratory | Claudia Regina Gonçalves; Claudio Tavares Sacchi; Erica Valessa Ramos Gomes |
| EPI_ISL_596503, EPI_ISL_596506, EPI_ISL_596509, EPI_ISL_596518, EPI_ISL_596521, EPI_ISL_596532, EPI_ISL_596535, EPI_ISL_596545, EPI_ISL_596555, EPI_ISL_596557, EPI_ISL_596561, EPI_ISL_596565 | | | |
| see above | Palestinian Ministry of Health | Molecular Genetics Lab | Damien Richard; Dana Najjar; Francois Balloux; Hisham Darwish; Husam Sallam; Issa Shtayeh; Lucy van Dorp; Mahmoud Ruzayqat; Nouar Qutob; Osama Najjar; Zaidoun Salah |
| EPI_ISL_707936, EPI_ISL_707939, EPI_ISL_708185, EPI_ISL_708186, EPI_ISL_708194 | Pamukkale University Hospital | Pamukkale University Department of Medical Genetics | Onur TOKGUN et al. |
| EPI_ISL_450790 | Pandit Deendayal Upadhyay Government Medical College, Rajkot | Gujarat Biotechnology Research Centre | A M Kadri; Amit Kanani; Ankit Hinsu; Apurvasinh Puvar; Bhavesh Modi; Chaitanya Joshi; Dinesh Kumar; Gaurishankar Shrimali; Janvi Raval; Komal Patel; Labdhi Pandya; Madhvi Joshi; Maharshi Pandya; Manish Pattani; Monika Gandhi; Nidhi Patel; Nitin Savaliya; Pinal Trivedi; Prakash Modi; Pritesh Sabara; R D Dixit; Raghawendra Kumar; Ramesh Pandit; Sejul Antala; Sharmistha Majumdar; Snehal Bagatharia; Tejas Shah; Zarna Patel; Zuber Saiyed |
| EPI_ISL_654866, EPI_ISL_654870, EPI_ISL_654883 | Pasteur Institute in Ho Chi Minh city | Department of Microbiology and Immunology - Pasteur Institute in Ho Chi Minh city | Cao Minh Thắng; Hoàng Kim Loan; Hoàng Minh; Hoàng Như Đào; Hoàng Quốc Cường; Huỳnh Phương Thảo; Huỳnh Thị Kim Loan; Lê Hoàng Chương; Lương Chấn Quang; Nguyễn Hoàng Anh; Nguyễn Hoàng Quân; Nguyễn Thanh Long; Nguyễn Thu Ngọc; Nguyễn Thị Ngọc Thảo; Nguyễn Thị Thanh Thương; Nguyễn Trung Hiếu; Nguyễn Trọng Toàn; Nguyễn Vũ Thượng; Phan Trọng Lân; Phạm Duy Quang; Phạm Thị Nhung; Phạm Thị Thu Hằng; Trần Thị Hồng Kim; Vũ Phạm Hồng Nhung; Đào Huy Mạnh; Đặng Thanh Giang |
| EPI_ISL_456221, EPI_ISL_622805, EPI_ISL_637088 | PathLab Bay of Plenty | Institute of Environmental Science and Research (ESR) | Anja Werno; Antje van der Linden; Arlo Upton; Chris Mansell; David Hammer; Dragana Drinkovic; Erasmus Smit; Gary McAuliffe; Hana Sofia Andersson; Hermes Perez; James Ussher; Jill Sherwood; Jing Wang; Joep de Ligt; Josh Freeman; Julia Howard; Juliet Elvy; Lauren Jelly; Mary DeAlmeida; Matt Blakiston; Matt Storey; Matthew Rogers; Max Bloomfield; Michael Addidle; Michelle Balm; Muhammad Faisal; Nikki Freed; Olin Silander; Sally Roberts; Sarah Jefferies; Sharmini Muttaiyah; Susan Morpeth; Susan Taylor; Timothy Blackmore; Vani Sathyendran; Veronica Playle; Virginia Hope; Xiaoyun Ren |
| EPI_ISL_420456 | PathWest Laboratory Medicine WA | PathWest Laboratory Medicine WA | Avram Levy; Chisha Sikazwe; David Smith and David Speers; Jurissa Lang |
| EPI_ISL_582023 | Pathlab Lakes | Institute of Environmental Science and Research (ESR) | Anja Werno; Antje van der Linden; Arlo Upton; Chris Mansell; David Hammer; Dragana Drinkovic; Erasmus Smit; Gary McAuliffe; Hana Sofia Andersson; Hermes Perez; James Ussher; Jill Sherwood; Jing Wang; Joep de Ligt; Josh Freeman; Julia Howard; Juliet Elvy; Lauren Jelly; Mary DeAlmeida; Matt Blakiston; Matt Storey; Matthew Rogers; Max Bloomfield; Michael Addidle; Michelle Balm; Muhammad Faisal; Nikki Freed; Olin Silander; Sally Roberts; Sarah Jefferies; Sharmini Muttaiyah; Susan Morpeth; Susan Taylor; Timothy Blackmore; Vani Sathyendran; Veronica Playle; Virginia Hope; Xiaoyun Ren |
| EPI_ISL_636972 | Pathogen Genomics Lab King Abdullah University of Science and Technology (KAUST) | Pathogen Genomics Lab King Abdullah University of Science and Technology (KAUST) | Afrah Alsomali; Amanda Ooi; Amit Kumar Subudhi; Anwar Hashem; Arnab Pain; Asim Khogeer; Fadwa Alofi; Fathia Ben Rached; Kahled Algithami; Luke Esau; Naif Almontashiri; Raeece Naeem; Rahul P Salunke; Sara Mfarrej; Sharif Hala |
| EPI_ISL_437739, EPI_ISL_512880, EPI_ISL_512927, EPI_ISL_512931, EPI_ISL_512992, EPI_ISL_513028, EPI_ISL_513074, EPI_ISL_513115, EPI_ISL_513141, EPI_ISL_513175, EPI_ISL_513188, EPI_ISL_513192, EPI_ISL_513206, EPI_ISL_513246, EPI_ISL_677974, EPI_ISL_677990, EPI_ISL_678034, EPI_ISL_678089, EPI_ISL_678095, EPI_ISL_678142, EPI_ISL_678143, EPI_ISL_678196, EPI_ISL_678215, EPI_ISL_678216, EPI_ISL_678224, EPI_ISL_678230, EPI_ISL_678246, EPI_ISL_751225, EPI_ISL_751237 | | | |
| see above | Pathogen Genomics Lab King Abdullah University of Science and Technology(KAUST) | Pathogen Genomics Lab King Abdullah University of Science and Technology(KAUST) | Abbas Al Mutair; Abdulaziz Alahmadi; Afrah Alsomali; Ahmad Bakur Mahmoud; Amanda; Amanda Ooi; Amit Kumar Subudhi; Anwar Hashem; Arnab Pain; Asim Khogeer; Awad Al-Omari; Fadwa Alofi; Fathia Ben Rached; Fathia Ben-Rached; Jumana Taha; Kahled Algithami; Khaled Algithami; Luke; Luke Esau; Muhammad Shuaib; Naif Almontashiri; Nashwa Al-khotani; Olga Douvropoulou; Raeece Naeem; Rahul P Salunke; Rahul Salunke; Raushan Nugmanova; Samer Salih; Sara Mfarrej; Sharif Hala |
| EPI_ISL_566077, EPI_ISL_596322, EPI_ISL_733168 | Pathogenic Microorganisms Variability Laboratory | WHO National Influenza Centre Russian Federation | Alexander Gintsburg; Alexey Shchetinin; Andrey Komissarov; Andrey Pochtovyy; Anna Ignatieva; Anna Ivanova; Artem Fadeev; Artem Tkachuk; Daria Danilenko; Dmitry Bazhenov; Dmitry Lioznov; Ekaterina Milashenko; Elena Burtseva; Elena Nabieva; Elena Shidlovskaya; Elena Vokalova; Elizaveta Divisenko; Evgeniya Mukasheva; Evgeny Usachev; Evgeny Usachev; Georgii Bazykin; Kirill Krasnoslobotsev; Ksenia Safina; Kseniya Komissarova; Ludmila Kolobukhina; Maria Nikiforova; Maxim Rubalsky; Nadezhda Kuznetsova; Oleg Rubalsky; Olga Burgasova; Sergey Alkhovsky; Svetlana Smetanina; Svetlana Trushakova; Tatyana Vishnevskaya; Valeria Bacalin; Vladimir Gushchin |
| EPI_ISL_410717 | Pathology Queensland | Public Health Virology Laboratory | Alyssa Pyke; Amanda De Jong; Andrew Van Den Hurk; Ben Huang; Carmel Taylor; David Warrilow; Doris Genge; Elisabeth Gamez; Glen Hewitson; Ian Maxwell Mackay; Inga Sultana; Jamie McMahon; Jean Barcelon; Judy Northill; Mitchell Finger; Natalie Simpson; Neelima Nair; Peter Burtonclay; Peter Moore; Sarah Wheatley; Sean Moody; Sonja Hall-Mendelin; Timothy Gardam; and Frederick Moore. |
| EPI_ISL_522547 | Platforme CYROI | UMR PIMIT Université de La Réunion | Camille Lebarbenchon; David Wilkinson; Patrick Mavingui |
| EPI_ISL_735433 | Posto de Atendimento Saude Cidade Pasc Cajati | Instituto Adolfo Lutz, Interdiciplinary Procedures Center, Strategic Laboratory | Claudia Regina Gonçalves; Claudio Tavares Sacchi; Erica Valessa Ramos Gomes; Karoline Rodrigues Campos |
| EPI_ISL_529013 | Presidio Ospedaliero "S.Filippo e Nicola"-Avezzano | Istituto Zooprofilattico Sperimentale dell'Abruzzo e Molise "G.Caporale" | Ancora M; Cammà C; Curini V; Di Domenico M; Di Pasquale A; Lorusso A; Mangone I; Marcacci M; Puglia I; Rinaldi A; Savini G. |
| EPI_ISL_417064 | Prince of Wales Hospital | Hong Kong Department of Health | Alan K.L. Tsang; Dominic N.C. Tsang; Edman T.K. Lam; Peter C.W. Yip; Rickjason C.W. Chan |
| EPI_ISL_539807 | Princess Margaret Hospital | Hong Kong Department of Health | Alan K.L. Tsang; Dominic N.C. Tsang; Edman T.K. Lam; Peter C.W. Yip; Rickjason C.W. Chan |
| EPI_ISL_513513, EPI_ISL_513514, EPI_ISL_513515, EPI_ISL_513518, EPI_ISL_513519, EPI_ISL_513520, EPI_ISL_513521, EPI_ISL_513522, EPI_ISL_513524, EPI_ISL_513525, EPI_ISL_513526, EPI_ISL_513527, EPI_ISL_513529, EPI_ISL_513530, EPI_ISL_513531, EPI_ISL_513532, EPI_ISL_513533, EPI_ISL_513534, EPI_ISL_513535, EPI_ISL_513536, EPI_ISL_513537, EPI_ISL_513538, EPI_ISL_513539, EPI_ISL_513540, EPI_ISL_513541, EPI_ISL_513542, EPI_ISL_513543, EPI_ISL_513544, EPI_ISL_513545, EPI_ISL_513546, EPI_ISL_513547, EPI_ISL_513548, EPI_ISL_513549, EPI_ISL_513550, EPI_ISL_513551, EPI_ISL_513553, EPI_ISL_513556, EPI_ISL_513557, EPI_ISL_513558, EPI_ISL_513559, EPI_ISL_513560, EPI_ISL_513561, EPI_ISL_513562, EPI_ISL_513563, EPI_ISL_513564, EPI_ISL_513567, EPI_ISL_513568, EPI_ISL_513571, EPI_ISL_513572, EPI_ISL_513573, EPI_ISL_513574, EPI_ISL_513575, EPI_ISL_513576, EPI_ISL_513577, EPI_ISL_513578, EPI_ISL_513579, EPI_ISL_513580, EPI_ISL_513581, EPI_ISL_513582, EPI_ISL_513583 | | | |
| see above | Programa de Oncovirologia, Instituto Nacional de Câncer | Programa de Oncovirologia, Instituto Nacional de Câncer | Andreia C. de Melo; Brunna M. Alves; Claudia Cicala; James Arthos; João P.B. Viola; Juliana D. Siqueira; Livia R. Goes; Marcelo A. Soares |
| EPI_ISL_527858 | Pronto Atendimento Sancta Maggiore Jardim Paulista | Instituto Adolfo Lutz, Interdiciplinary Procedures Center, Strategic Laboratory | Claudia Regina Gonçalves; Claudio Tavares Sacchi; Erica Valessa Ramos Gomes |
| EPI_ISL_603021, EPI_ISL_693204 | Pronto Socorro Dr. Conrado Cesarino Nuvolini | Instituto Adolfo Lutz, Interdiciplinary Procedures Center, Strategic Laboratory | Claudia Regina Gonçalves; Claudio Tavares Sacchi; Erica Valessa Ramos Gomes; Karoline Rodrigues Campos |
| EPI_ISL_471556 | Pronto Socorro Jose Ibrahin | Instituto Adolfo Lutz, Interdiciplinary Procedures Center, Strategic Laboratory | Claudia Regina Gonçalves; Claudio Tavares Sacchi; Erica Valessa Ramos Gomes |
| EPI_ISL_527867 | Pronto Socorro Municipal - Balneario São José | Instituto Adolfo Lutz, Interdiciplinary Procedures Center, Strategic Laboratory | Claudia Regina Gonçalves; Claudio Tavares Sacchi; Erica Valessa Ramos Gomes |
| EPI_ISL_523961 | Pronto Socorro Municipal 21 de Junho | Instituto Adolfo Lutz, Interdiciplinary Procedures Center, Strategic Laboratory | Claudia Regina Gonçalves; Claudio Tavares Sacchi; Erica Valessa Ramos Gomes |
| EPI_ISL_515529 | Pronto Socorro Municipal Julio Tupy | Instituto Adolfo Lutz, Interdiciplinary Procedures Center, Strategic Laboratory | Claudia Regina Gonçalves; Claudio Tavares Sacchi; Erica Valessa Ramos Gomes |
| EPI_ISL_693202 | Pronto Socorro Municipal Prof. Joao Catarin Mezomo | Instituto Adolfo Lutz, Interdiciplinary Procedures Center, Strategic Laboratory | Claudia Regina Gonçalves; Claudio Tavares Sacchi; Erica Valessa Ramos Gomes; Karoline Rodrigues Campos |
| EPI_ISL_515554, EPI_ISL_523958, EPI_ISL_523959 | Pronto Socorro Municipal de Perus | Instituto Adolfo Lutz, Interdiciplinary Procedures Center, Strategic Laboratory | Claudia Regina Gonçalves; Claudio Tavares Sacchi; Erica Valessa Ramos Gomes |
| EPI_ISL_693231 | Pronto Socorro Municipal de Santa Branca | Instituto Adolfo Lutz, Interdiciplinary Procedures Center, Strategic Laboratory | Claudia Regina Gonçalves; Claudio Tavares Sacchi; Erica Valessa Ramos Gomes; Karoline Rodrigues Campos |
| EPI_ISL_693210 | Pronto-Socorro Dr. Osmar Mesquita | Instituto Adolfo Lutz, Interdiciplinary Procedures Center, Strategic Laboratory | Claudia Regina Gonçalves; Claudio Tavares Sacchi; Erica Valessa Ramos Gomes; Karoline Rodrigues Campos |
| EPI_ISL_636973 | Public Health Lab | Public Health Lab | Alwasti; H |

| | | | |
|---|---|---|---|
| EPI_ISL_413015 | Public Health Ontario Laboratory | National Microbiology Laboratory | Adrian Zetner; Anna Majer; Darryl Falzarano; Erika Landry; Gerdts Volker; Grace Seo; Guillaume Poliquin; Jocelyne Lew; Jonathan Gubbay; Matthew Gilmour; Morag Graham; Natalie Knox; Nathalie Bastien; Philip Mabon; Rob Kozak; Samira Mubareka; Shari Tyson; Stephanie Booth; Timothy Booth; Tom Graefenhan; Yan Li |
| EPI_ISL_513310 | Public Health, United States Air Force School of Aerospace Medicine | Public Health, United States Air Force School of Aerospace Medicine | A.C.; A.K.; A.W.; B.C.; C.R.; Chapleau; Connors; E.A.; Fries; J.R.; Javorina; Lambert; Macias; Meyer; Purves; R.R. and Starr; S.M. |
| EPI_ISL_513312 | Public Health, United States Air Force School of Aerospace Medicine | Public Health, United States Air Force School of Aerospace Medicine | A.C.; A.K.; A.W.; B.C.; C.R.; Chapleau; Connors; E.A.; Fries; J.R.; Javorina; Lambert; Macias; Meyer; Purves; R.R. and Starr; S.M. |
| EPI_ISL_434547 | Puerto Rico Department of Health | Centers for Disease Control and Prevention, Dengue Branch | Betzabel Flores; Chaney Kalinich; Fabiola Cruz; Gilberto A. Santiago; Glenda Gonzalez; Jessica I. Falcon; Jorge L. Munoz-Jordan; Joseph Fauver; Keyla Charriez; Nathan Grubaugh |
| EPI_ISL_632936 | Puskesmas Mlati 1 Sleman | Genetics Working Group (Pokja Genetik) Faculty of Medicine, Public Health and Nursing Universitas Gadjah Mada (FK-KMK UGM); Disease Investigation Center Wates Ministry of Agriculture Indonesia; Department of Microbiology FK-KMK UGM; Laboratorium Diagnostik Yayasan Tahija World Mosquito Program (WMP) Yogyakarta Center for Tropical Medicine FK-KMK UGM; Integrated Research Center FK-KMK UGM; Department of Computer Science and Electronics FMIPA UGM; RSUP Dr. Sardjito | Afiahayati; Bambang Sigit Riyanto; Dwi AA Nugrahaningsih; Edwin W. Daniwijaya; Eggi Arguni; Eko Budiono; Endah Supriyati; Ernawati; Gunadi; Hendra Wibawa; Heni Retnowulan; Ika Trisnawati; Ira Puspitawati; Kristy Iskandar; Ludhang P. Rizki; Marcellus; Mohamad S. Hakim; Munawar Gani; Nungki Anggorowati; Nur Imma Fatimah Harahap; Nur Rahmi Ananda; Osman Sianipar; Riat El Khair; Satria Maulana; Siswanto; Sumardi; Titik Nuryastuti; Tri Wibawa; Umi Solekhah Intansari; Untung Wirawan; William Widitjiarso; Yunika Puspadewi; Elizabeth Henny Herningtiyas |
| EPI_ISL_579679 | QEII Health Sciences Centre | National Microbiology Laboratory (NML) | Anna Majer; Anneliese Landgraff; CanCOGeN's metadata curation team; Darian Hole; Elsie Grudeski; Gary Van Domselaar; Grace Seo; Jason LeBlanc; Jennifer Tanner; Morag Graham; Natalie Knox; Nathalie Bastien; Philip Mabon; Public Health Agency of Canada's CanCOGeN team; Rhiannon Huzarewich; Russell Mandes; Shari Tyson; Timothy Booth; Todd Hatchette; Yan Li |
| EPI_ISL_725189, EPI_ISL_725416 | Quadram Institute Bioscience | COVID-19 Genomics UK (COG-UK) Consortium | Alexander J Trotter; Alison E. Mather; Alp Aydin; Ana P. Tedim; Anastasia Kolyva; Andrew Bell; Andrew J. Page; Claire Stuart; Dave J. Baker; Gemma L. Kay; John Wain; Justin O'Grady; Leonardo de Oliveira Martins; Lizzie Meadows; Maria Diaz; Mark Webber; Muhammed Yasir; Nabil-Fareed Alikhan; Ngozi Elumogo; Nicholas M. Thomson; Rachael Stanley; Rachel Gilroy; Reenesh Prakash; Samir Dervisevic; Samuel Bloomfield; Steven Rudder; Thanh Le-Viet |
| EPI_ISL_487434, EPI_ISL_649154 | Queen Astrid Military Hospital | Institute of Tropical Medicine | Colin Anthony; Philippe Selhorst |
| EPI_ISL_733571 | Queen Mary Hospital | Hong Kong Department of Health | Alan K.L. Tsang; Dominic N.C. Tsang; Edman T.K. Lam; Peter C.W. Yip; Rickjason C.W. Chan |
| EPI_ISL_526395, EPI_ISL_741650 | Queens Medical Centre, Clinical Microbiology Department / DeepSeq Nottingham | COVID-19 Genomics UK (COG-UK) Consortium | Christopher Moore; Fei Sang; Gemma Clark; Hannah Howson-Wells; Johnny Debebe; Jonathan Ball; Joseph Chappell; Manjinder Khakh; Matthew Carlisle; Matthew Loose; Michelle M Lister; Nadine Holmes; Patrick McClure; Theocharis Tsoleridis; Vicki M Fleming; Victoria Wright; Wendy Smith |
| EPI_ISL_639790 | Queensland Health Forensic and Scientific Services | Queensland Health Forensic and Scientific Services | Son Nguyen et al |
| EPI_ISL_604647 | Quest Diagnostics | Quest Diagnostics | Anderson, B.; D.F.; Gerasimova, A.; Grover, D.; Hua, M.; K.E.; Kagan; Lacbawan, F.; Liu Y.; Livingston; Owen, R.; R.M.; Rosenthal; S.H.; Shalhout |
| EPI_ISL_415641, EPI_ISL_415642, EPI_ISL_416480 | R. G. Lugar Center for Public Health Research, National Center for Disease Control and Public Health (NCDC) of Georgia. | R. G. Lugar Center for Public Health Research, National Center for Disease Control and Public Health (NCDC) of Georgia. | Adam Kotorashvili; Amiran Gamkrelidze.; Ana Papkiauri; Ann Machablishvili; Anna Kasradze; Davit Tsaguria; Ekaterine Khmaladze; Ekaterine Zangaladze; Ekaterine Zhghenti; Giorgi Tomashvili; Gvantsa Brachveli; Gvantsa Chanturia; Irma Burjanadze; Ketevan Sidamonidze; Khatuna Zakhashvili; Lela Sabadze; Lela Urushadze; Magda Dgebuadze; Maia Alkhazashvili; Mari Gavashelidze; Mariam Zakalashvili; Marine Murtskhvaladze; Meri Pantsulaia; Nato Kotaria; Nino Berishvili; Paata Imnadze; Roena Sukhiashvili; Tamar Jashiashvili; Tata Imnadze; Tea Tevdoradze |
| EPI_ISL_766045 | RS Pelni | Eijkman Institute for Molecular Biology, Ministry of Research and Technology/National Agency for Research and Innovation | Amin Soebandrio; David H Muljono; Edison Johar; Frilasita A Yudhaputri; Hidayat Trimarsanto; Khin Saw Myint; Safarina G Malik |
| EPI_ISL_437190 | RS Pondok Indah Hospital – Pondok Indah | Eijkman Institute for Molecular Biology, Ministry of Research and Technology/National Agency for Research and Innovation | Amin Soebandrio; David H Muljono; Edison Johar; Frilasita A Yudhaputri; Hidayat Trimarsanto; Khin Saw Myint; Safarina G Malik |
| EPI_ISL_791985 | RS Santa Maria Pekanbaru | National Institute of Health Research and Development | AA; Fridayenti; HA; HD; Ikawati; KD; KNA; N; Nugraha; Pangesti; Pawestri; Puspa; Puspandari; Setiawaty; Soekarso; Subangkit; T; V |
| EPI_ISL_766042 | RS Sari Asih Ciledug | Eijkman Institute for Molecular Biology, Ministry of Research and Technology/National Agency for Research and Innovation | Amin Soebandrio; David H Muljono; Edison Johar; Frilasita A Yudhaputri; Hidayat Trimarsanto; Khin Saw Myint; Safarina G Malik |
| EPI_ISL_574606 | RS Siloam Purwakarta | Eijkman Institute for Molecular Biology, Ministry of Research and Technology/National Agency for Research and Innovation | Amin Soebandrio; David H Muljono; Edison Johar; Frilasita A Yudhaputri; Herawati Sudoyo; Hidayat Trimarsanto; Iskandar A Adnan; Khin Saw Myint; Safarina G Malik; Willy Agustine |
| EPI_ISL_454497, EPI_ISL_454515 | RSE "National Center for Biotechnology" | RSE "National Center for Biotechnology" | Akbota Rakhmetova; Alexandr Shevtsov; Askar Abdaliyev; Asylulan Amirgazin; Ilyas Akhmetollayev; Ruslan Kalendar; Viktoriya Lutsay; Yerlan Ramankulov; Zabira Aushakhmetova |
| EPI_ISL_766038 | RSJPD Harapan Kita | Eijkman Institute for Molecular Biology, Ministry of Research and Technology/National Agency for Research and Innovation | Amin Soebandrio; David H Muljono; Edison Johar; Frilasita A Yudhaputri; Hidayat Trimarsanto; Khin Saw Myint; Safarina G Malik |
| EPI_ISL_791980 | RSUD Blambangan Banyuwangi | National Institute of Health Research and Development | AA; HA; HD; Ikawati; KD; KNA; N; Noor; Nugraha; Pangesti; Pawestri; Puspa; Puspandari; RI; Setiawaty; Soekarso; Subangkit; T; V |
| EPI_ISL_610162 | RSUD Dr. Tjitrowardojo | Genetics Working Group (Pokja Genetik) Faculty of Medicine, Public Health and Nursing Universitas Gadjah Mada (FK-KMK UGM); Disease Investigation Center Wates Ministry of Agriculture Indonesia; Department of Microbiology FK-KMK UGM; Laboratorium Diagnostik Yayasan Tahija World Mosquito Program (WMP) Yogyakarta Center for Tropical Medicine FK-KMK UGM; Integrated Research Center FK-KMK UGM; Department of Computer Science and Electronics FMIPA UGM; Balai Besar Teknik Kesehatan Lingkungan dan Pengendalian Penyakit (BBTKLPP) Yogyakarta | Afiahayati; Alvin Santoso Kalim; Dwi AA Nugrahaningsih; Edwin W. Daniwijaya; Eggi Arguni; Endah Supriyati; Gunadi; Havid Setyawan; Hendra Wibawa; Indaryati; Irene; Kristy Iskandar; Ludhang P. Rizki; Marcellus; Mohamad S. Hakim; Nungki Anggorowati; Siswanto; Susan Simanjaya; Titik Nuryastuti; Tri Wibawa; Wuryanto |
| EPI_ISL_791986 | RSUD Morotai Maluku Utara | National Institute of Health Research and Development | AA; Denggo; HA; HD; Ikawati; KD; KNA; N; Nugraha; Pangesti; Pawestri; Puspa; Puspandari; SD; Setiawaty; Soekarso; Subangkit; T; V |
| EPI_ISL_632937 | RSUD Saptosari Gunung Kidul | Genetics Working Group (Pokja Genetik) Faculty of Medicine, Public Health and Nursing Universitas Gadjah Mada (FK-KMK UGM); Disease Investigation Center Wates Ministry of Agriculture Indonesia; Department of Microbiology FK-KMK UGM; Laboratorium Diagnostik Yayasan Tahija World Mosquito Program (WMP) Yogyakarta Center for Tropical Medicine FK-KMK UGM; Integrated Research Center FK-KMK UGM; Department of Computer Science and Electronics FMIPA UGM; Balai Besar Teknik Kesehatan Lingkungan dan Pengendalian Penyakit (BBTKLPP) Yogyakarta | Afiahayati; Audric Kenny Tedja; Dwi AA Nugrahaningsih; Edwin W. Daniwijaya; Eggi Arguni; Eko Darmawan; Endah Supriyati; Gunadi; Havid Setyawan; Hendra Wibawa; Indaryati; Irene; Kristy Iskandar; Ludhang P. Rizki; Marcellus; Maria Patricia Inggriani; Mohamad S. Hakim; Nungki Anggorowati; Siswanto; Titik Nuryastuti; Tri Wibawa |
| EPI_ISL_538510 | RSUD Ulin Banjarmasin South Kalimantan | National Institute of Health Research and Development | AA; HA; HD; Ikawati; KD; KNA; M; Nugraha; Paisal; Pangesti; Pasaribu; Pawestri; Puspa; Setiawaty, V.; Soekarso; Subangkit; T |
| EPI_ISL_766032 | RSUP Dr. Sardjito | Eijkman Institute for Molecular Biology, Ministry of Research and Technology/National Agency for Research and Innovation | Amin Soebandrio; David H Muljono; Edison Johar; Frilasita A Yudhaputri; Hidayat Trimarsanto; Khin Saw Myint; Safarina G Malik |
| EPI_ISL_576113 | RSUP Dr. Sardjito | Genetics Working Group (Pokja Genetik) Faculty of Medicine, Public Health and Nursing Universitas Gadjah Mada (FK-KMK UGM); Disease Investigation Center Wates Ministry of Agriculture Indonesia; Department of Microbiology FK-KMK UGM; Laboratorium Diagnostik Yayasan Tahija World Mosquito Program (WMP) Yogyakarta Center for Tropical Medicine FK-KMK UGM; Integrated Research Center FK-KMK UGM; Department of Computer Science and Electronics FMIPA UGM | . Marcellus; Afiahayati; Dwi AA Nugrahaningsih; Edwin W. Daniwijaya; Eggi Arguni; Endah Supriyati; Fadil Fahri; Gunadi; Hendra Wibawa; Ika Trisnawati; Kristy Iskandar; Ludhang P. Rizki; Mohamad S. Hakim; Nungki Anggorowati; Riat El Khair; Siswanto; Titik Nuryastuti; Tri Wibawa; William Widitjiarso |
| EPI_ISL_568691, EPI_ISL_766046 | RSUP Fatmawati | Eijkman Institute for Molecular Biology, Ministry of Research and Technology/National Agency for Research and Innovation | Amin Soebandrio; David H Muljono; Edison Johar; Frilasita A Yudhaputri; Herawati Sudoyo; Hidayat Trimarsanto; Iskandar A Adnan; Khin Saw Myint; Safarina G Malik; Willy Agustine |
| EPI_ISL_574604 | RSUP Prof. Dr. R. Kandou Manado | Eijkman Institute for Molecular Biology, Ministry of Research and Technology/National Agency for Research and Innovation | Amin Soebandrio; David H Muljono; Edison Johar; Frilasita A Yudhaputri; Herawati Sudoyo; Hidayat Trimarsanto; Iskandar A Adnan; Khin Saw Myint; Safarina G Malik; Willy Agustine |
| EPI_ISL_525492 | RSUP dr. SOERADJI TIRTONEGORO | Genetics Working Group (Pokja Genetik) Faculty of Medicine, Public Health and Nursing Universitas Gadjah Mada (FK-KMK UGM), Disease Investigation Center Wates Ministry of Agriculture Indonesia, Department of Microbiology FK-KMK UGM, Laboratorium Diagnostik Yayasan Tahija World Mosquito Program (WMP) Yogyakarta Center for Tropical Medicine FK-KMK UGM, Integrated Research Center FK-KMK UGM, Department of Computer Science and Electronics FMIPA UGM | . Marcellus; . Siswanto; Afiahayati; Alvin S. Kalim; Dwi AA Nugrahaningsih; Dwiki Afandy; Edwin W. Daniwijaya; Eggi Arguni; Endah Supriyati; Gunadi; Hendra Wibawa; Indah Juliana; Kurniyanto; Ludhang P. Rizki; Mohamad S. Hakim; Titik Nuryastuti; Tri Wibawa |
| EPI_ISL_450511, EPI_ISL_450512, EPI_ISL_450515 | Rafik Hariri University Hospital | Rafik Hariri University Hospital | Rita Feghali |
| EPI_ISL_429177, EPI_ISL_447019, EPI_ISL_455909, EPI_ISL_512863, EPI_ISL_693377 | Ramathibodi Hospital | COVID-19 Network Investigations (CONI) Alliance | Angkana Huang; Anthony R. Jones; Arporn Wangwiwatsin; Bhakbhoom Panthan; Chonticha Klungtong; Ekawat Pasomsub; Elizabeth Batty; Insee Sensorn; Janjira Thaipadungpanit; Khajohn Joonsalak; Kingkan Rakmanee; Krittikorn Kumpornsin; Namfon Kotanan; Poramate Jiaranai; Stefan Fernandez; Thanat Chookajorn; Theerarat Kochakarn; Treewat Watthanachockchai; Wasun Chantratita; Wudtichai Manasatienkij |
| EPI_ISL_735490 | Rangamati General Hospital RT-PCR lab, | Central Biological Research Laboratory and Department of Biochemistry and Molecular Biology | H. M. Abdullah Al Masud; Imam Hossen; Md. Arif Hossain; Md. Imranul Hoq; Md. Khondakar Raziur Rahman; Md. Omer Faruq; Mohammad Omar Faruque; Robiul Hasan Bhuiyan; Sajib Rudra; Shanta Paul |
| EPI_ISL_768534 | Regional Medical Sciences Center 5 Samut Songkhram | National Institute of Health, Department of Medical Sciences, Ministry of Public Health, Thailand | ; Natchaya Khiadsang; Pakorn Piromtong; Pilailuk Okada; Ratana Tacharoenmuang; Siripaporn Phuygun; Sittiporn Parnmen; Sunthareeya Waicharoen; Thanutsapa Thanadachakul; Warawan Wongboot; sirikanda wimol |
| EPI_ISL_441381, EPI_ISL_441410, EPI_ISL_441431, EPI_ISL_448951, EPI_ISL_585200, EPI_ISL_627543, EPI_ISL_627566, EPI_ISL_680198, EPI_ISL_680235, EPI_ISL_742130, EPI_ISL_742148, EPI_ISL_764387, EPI_ISL_764435 | | | |
| see above | Regional Virus Laboratory, Belfast Health and Social Care Trust | COVID-19 Genomics UK (COG-UK) Consortium | Alison Watt; Ciara Cox; Conall McCaughey; David Simpson; Derek Fairley; James McKenna; Mairead Connor; Susan Feeney; Tanya Curran; Zoltan Molnar |
| EPI_ISL_441698, EPI_ISL_470024, EPI_ISL_489187, EPI_ISL_489224, EPI_ISL_489257, EPI_ISL_489285, EPI_ISL_500968, EPI_ISL_501035, EPI_ISL_501040 | | | |
| see above | Regional Virus Laboratory, Belfast Health and Social Care Trust | Wellcome Sanger Institute for the COVID-19 Genomics UK (COG-UK) consortium | Alex Alderton; Alison Watt; Ciara Cox; Conall McCaughey; Cordelia Langford; David K. Jackson; David Simpson; Derek Fairley; Dominic Kwiatkowski; Ewan Harrison; Ian Johnston; James McKenna; John Sillitoe on behalf of the Wellcome Sanger Institute COVID-19 Surveillance Team (http://www.sanger.ac.uk/covid-team); Mairead Connor; Roberto Amato; Sonia Goncalves; Susan Feeney; Tanya Curran; Zoltan Molnar; and Alex Alderton |
| EPI_ISL_708811 | Regional medical sciences center 2 Phitsanulok | National Institute of Health, Department of Medical Sciences, Ministry of Public Health, Thailand | Malinee Chittaganpitch; Pakorn Piromtong; Pilailuk Okada; Siripaporn Phuygun; Sittiporn Parnmen; Sunthareeya Waicharoen; Thanutsapa Thanadachakul; Warawan Wongboot |
| EPI_ISL_708800, EPI_ISL_708801, EPI_ISL_708807 | Regional medical sciences center 6 chonburi | National Institute of Health, Department of Medical Sciences, Ministry of Public Health, Thailand | Malinee Chittaganpitch; Pakorn Piromtong; Pilailuk Okada; Siripaporn Phuygun; Sittiporn Parnmen; Sunthareeya Waicharoen; Thanutsapa Thanadachakul; Warawan Wongboot |
| EPI_ISL_451934 | Research Unit, University Hospital for Infectious Diseases "Dr. Fran Mihaljević" | Cicin Sain lab, Helmholtz Centre for Infection Research | Ivan-Christian Kurolt; Kathrin Eschke; Zeeshan Chaudhry; Željka Mačak Šafranko |

| | | | |
|---|---|---|---|
| EPI_ISL_454418 | Research and Experiment Center, Meizhou People Hospital | Research and Experiment Center, Meizhou People Hospital | Guo, X.; L. and Yu, Z.; Zeng |
| EPI_ISL_661271 | Research platform for Transfusion-transmitted Disease, Institute of Blood Transfusion, Chinese Academy of Medical Sciences | Research platform for Transfusion-transmitted Disease, Institute of Blood Transfusion, Chinese Academy of Medical Sciences | He; M. and Fan, Z. |
| EPI_ISL_407071, EPI_ISL_412116, EPI_ISL_417215, EPI_ISL_421769, EPI_ISL_464178, EPI_ISL_464193, EPI_ISL_464799, EPI_ISL_465485 | | | |
| see above | Respiratory Virus Unit, Microbiology Services Colindale, Public Health England | Respiratory Virus Unit, Microbiology Services Colindale, Public Health England | Angie Lackenby; Joanna Ellis; Jonathan Hubb; Kirstin Edwards; Leena Bhaw; Maria Zambon; Monica Galiano; Omolola Akinbami; PHE Covid Sequencing Team; Richard Myers; Shahjahan Miah; Steven Platt; Tiina Talts |
| EPI_ISL_682298 | Respiratory virus Laboratory, Chinese Academy of Medical Science | Respiratory virus Laboratory, Chinese Academy of Medical Science | Chen; Li, J.; Y. and Liu, L.; Zhen, H. |
| EPI_ISL_640035 | Riversdale Clinic wc RAV | NHLS/UCT | Arash Iranzadeh; Bruna Galvao; Carolyn Williamson; Deelan Doolabh; Diana Hardie; Innocent Mudau; Kruger Marais; Lynn Tyers; Marvin Hsiao; Stephen Korsman |
| EPI_ISL_514131, EPI_ISL_514132, EPI_ISL_514133, EPI_ISL_514134, EPI_ISL_514135, EPI_ISL_514136, EPI_ISL_514137, EPI_ISL_514138 | | | |
| see above | Rondônia Central Public Health Laboratory (LACEN/RO), vinculated to State Health Secretariat of Rondônia (SESAU/RO) | Molecular Virology Laboratory of Oswaldo Cruz Foundation of Rondônia | Adriana Cristina Salvador Maia; Alcione de Oliveira dos Santos; Alice Paula Di Sabatino Guimarães; Aline Linhares Ferreira de Melo Mendonça; Caio Henrique Nemeth Santos; Camila Flávia Gomes Azzi; Celina Aparecida Bertoni Lugtenburg; Cicileia Correia da Silva; Felipe Gomes Naveca; Felipe Souza Nogueira-Lima; Fernando Rodrigues Máximo; Jansen Fernandes de Medeiros; Juan Miguel Vilallobos-Salcedo and Deusilene Souza Vieira; Juan Miguel Vilallobos-Salcedo and Deusilene Souza Vieira.; Juan Miguel Villalobos-Salcedo and Deusilene Souza Vieira1; Juliana Loca Furtado; Luan Felipo Botelho-Souza; Suelen Cavalcante; Tárcio Peixoto Roca; Rita de Cássia Pontello Rampazzo |
| EPI_ISL_522679 | Royal Hobart Hospital Microbiology Department | MDU-PHL | Cooley L.; M.B.; Sait M.; Schultz; Seemann T.; Sherry N.; van Haeften R. |
| EPI_ISL_516800 | Rumah Sakit Akademik Universitas Gadjah Mada | Genetics Working Group (Pokja Genetik) Faculty of Medicine, Public Health and Nursing Universitas Gadjah Mada (FK-KMK UGM); Disease Investigation Center Wates Ministry of Agriculture Indonesia; Department of Microbiology FK-KMK UGM; Laboratorium Diagnostik Yayasan Tahija World Mosquito Program (WMP) Yogyakarta Center for Tropical Medicine FK-KMK UGM; Integrated Research center FK-KMK UGM; Department of Computer Science and Electronics FMIPA UGM | . Marcellus; Afiahayati; Alvin S. Kalim; Desyifa Mursalin; Dwi AA Nugrahaningsih; Edwin W. Daniwijaya; Eggi Arguni; Endah Supriyati; Gunadi; Hendra Wibawa; Ludhang P. Rizki; Mohamad S. Hakim; Siswanto; Titik Nuryastuti; Tri Wibawa |
| EPI_ISL_451132, EPI_ISL_622762, EPI_ISL_755570 | SA Pathology | SA Pathology | Chuan Kok Lim; Geoff Higgins; Ivan Bastian; Julien Soubrier; Karin Kassahn; Lex Leong; Mark Turra; Song Gao |
| EPI_ISL_455603 | SA Pathology | VPRL | Beard, MR.; C.K.; Coldbeck-Shackley, R.; Kirby, E.; L.E.X.; Leong; Lim; Llamas, B.; Merrett, J.; Shue, B.; Van Der Hoek, K. |
| EPI_ISL_648217 | SILAB | Laboratorio Mixto de Biotecnología Acuática (LMBA) | Adriana Giri; Agustina Cerri; Ana Cavatorta; Ana Paletta; Diego Chouhy; Elisa Bolatti; Elizabeth Tapia; Federico Remes Lenicov; Flavio Spetale; Gastón Viarengo; Ignacio García Labari; Javier Murillo; Joaquín Ezpeleta; Julian Acosta; Laura Angelone; Leandro Ciappina; María Re; Pablo Casal; Pilar Bulacio; Silvana Spinelli; Silvia Arranz; Sofía Lavista Llanos; Vanina Villanova; Victoria Posner |
| EPI_ISL_547575 | SVO Jundiaí | Instituto Adolfo Lutz, Interdiciplinary Procedures Center, Strategic Laboratory | Claudia Regina Gonçalves; Claudio Tavares Sacchi; Erica Valessa Ramos Gomes; Karoline Rodrigues Campos |
| EPI_ISL_796781 | SYNLAB | Instituto Nacional de Saude (INSA) | Borges et al |
| EPI_ISL_635333, EPI_ISL_635853 | San Diego County Public Health Laboratory | Andersen lab at Scripps Research | Brett Austin; Jovan Shephard; SEARCH Alliance San Diego with Tracy Basler |
| EPI_ISL_749155, EPI_ISL_750169, EPI_ISL_750178, EPI_ISL_750430 | Sanatorio Americano | Institut Pasteur de Montevideo | Ana Carolina Mendonça; Andrés Lizasoain; Camila Simoes; Cecilia Alonso; Cecilia Salazar; Daiana Mir; Fernando López-Tort; Fernando Motta; Gonzalo Bello; Ighor Arantes; Ignacio Ferrés; Jose Sotelo; Leticia Maya; Letícia Garay Martins; Luciana Appolinario; Lucía Spangenberg; Mailen Arleo; Mariana Brandes; Marilda Mendonça Siqueira; Marilda Tereza Mar da Rosa; María José Benitez-Galeano; Martín Graña; Matías Castells; Matías Victoria; Matías Salvo; Natalia Rego; Natalia Reyes; Pablo Smircich; Paola Cristina Resende; Rodney Colina; Tamara Fernandez-Calero; Tania Possi; Tatiana Schäffer Gregianini; Verónica Noya; Yasser Vega |
| EPI_ISL_583492 | Santa Casa Anna Cintra | Instituto Adolfo Lutz, Interdiciplinary Procedures Center, Strategic Laboratory | Claudia Regina Gonçalves; Claudio Tavares Sacchi; Erica Valessa Ramos Gomes; Karoline Rodrigues Campos |
| EPI_ISL_547580, EPI_ISL_603026, EPI_ISL_603032, EPI_ISL_693219 | Santa Casa da Misericórdia de Presidente Prudente | Instituto Adolfo Lutz, Interdiciplinary Procedures Center, Strategic Laboratory | Claudia Regina Gonçalves; Claudio Tavares Sacchi; Erica Valessa Ramos Gomes; Karoline Rodrigues Campos |
| EPI_ISL_735407 | Santa Casa de Marilia | Instituto Adolfo Lutz, Interdiciplinary Procedures Center, Strategic Laboratory | Claudia Regina Gonçalves; Claudio Tavares Sacchi; Erica Valessa Ramos Gomes; Karoline Rodrigues Campos |
| EPI_ISL_603024, EPI_ISL_603027, EPI_ISL_603038 | Santa Casa de Misericordia de Araçatuba | Instituto Adolfo Lutz, Interdiciplinary Procedures Center, Strategic Laboratory | Claudia Regina Gonçalves; Claudio Tavares Sacchi; Erica Valessa Ramos Gomes; Karoline Rodrigues Campos |
| EPI_ISL_693212 | Santa Casa de Misericordia de Braganca Paulista | Instituto Adolfo Lutz, Interdiciplinary Procedures Center, Strategic Laboratory | Claudia Regina Gonçalves; Claudio Tavares Sacchi; Erica Valessa Ramos Gomes; Karoline Rodrigues Campos |
| EPI_ISL_524469 | Santa Casa de Misericordia de Sao Paulo | Instituto Adolfo Lutz, Interdiciplinary Procedures Center, Strategic Laboratory | Claudia Regina Gonçalves; Claudio Tavares Sacchi; Erica Valessa Ramos Gomes |
| EPI_ISL_693198 | Santa Casa de Misericordia de Sao Paulo - Hospital Central | Instituto Adolfo Lutz, Interdiciplinary Procedures Center, Strategic Laboratory | Claudia Regina Gonçalves; Claudio Tavares Sacchi; Erica Valessa Ramos Gomes; Karoline Rodrigues Campos |
| EPI_ISL_693211 | Santa Casa de Misericordia e Maternidade | Instituto Adolfo Lutz, Interdiciplinary Procedures Center, Strategic Laboratory | Claudia Regina Gonçalves; Claudio Tavares Sacchi; Erica Valessa Ramos Gomes; Karoline Rodrigues Campos |
| EPI_ISL_547579 | Santa Casa de Misericórdia de Araçatuba | Instituto Adolfo Lutz, Interdiciplinary Procedures Center, Strategic Laboratory | Claudia Regina Gonçalves; Claudio Tavares Sacchi; Erica Valessa Ramos Gomes; Karoline Rodrigues Campos |
| EPI_ISL_603031 | Santa Casa de Presidente Epitácio | Instituto Adolfo Lutz, Interdiciplinary Procedures Center, Strategic Laboratory | Claudia Regina Gonçalves; Claudio Tavares Sacchi; Erica Valessa Ramos Gomes; Karoline Rodrigues Campos |
| EPI_ISL_524464 | Santa Casa de Santa Isabel | Instituto Adolfo Lutz, Interdiciplinary Procedures Center, Strategic Laboratory | Claudia Regina Gonçalves; Claudio Tavares Sacchi; Erica Valessa Ramos Gomes |
| EPI_ISL_479799 | Sapporo City Institute of Public Health | Pathogen Genomics Center, National Institute of Infectious Diseases | Asami Ohnishi; Hajime Kamiya; Kentaro Itokawa; Makoto Kuroda; Masanori Hashino; Motoi Suzuki; Rina Tanaka; Tsuyoshi Sekizuka |
| EPI_ISL_450403, EPI_ISL_759858 | School of Public Health, The University of Hong Kong | School of Public Health, The University of Hong Kong | Barrs; Brackman; C.J.; Chu; D.K.W.; D.N.C.; E.M.W.; K.W.S.; L.D.; L.L.M. and Peiris, M.; Law; P.Y.L.; Peiris, M.; Perera; Poon; R.A.P.M.; R.A.P.M. and Sit; Sims; Sit; T.H.C.; T.H.S.; Tam; To; Tsang; V.R.; V.Y.T.; Yu |
| EPI_ISL_510529 | School of Veterinary Medicine, Disease Control | School of Veterinary Medicine, Disease Control | A.L.; A.N.; Bates, M.; C. and Zumla, A.; Chambaro, H.; Chanda, D.; Changula, K.; Chilufya; Chipimo; Chitanga, S.; Fwoloshi, S.; K.S.; Kapata; Kapata, N.; Kapaya, F.; Kapin'a, M.; Kayeyi, N.; Liwewe; M.M.; Malama, K.; Masahiro, K.; Monze, M.; Morales; Mubemba, B.; Mukonka, V.; Mulenga, L.; Muleya, W.; Mupeta, F.; Musonda, K.; Nalubamba; Ngosa, W.; P.C.; P.J.; Saasa, N.; Sawa, H.; Shibemba; Simulundu, E.; Sinyange, N.; Takada, A.; Tembo, J.; Zulu, P. |
| EPI_ISL_661180, EPI_ISL_661189, EPI_ISL_661191, EPI_ISL_661198, EPI_ISL_676576, EPI_ISL_676585, EPI_ISL_676589, EPI_ISL_676590 | | | |
| see above | Scientific Veterinary Institute Novi Sad | Veterinary Specialized Institute "Kraljevo", Serbia | Afonso, C.; Banovic Djeri, B.; Jankovic, M.; Jovanovic, T.; Knezevic, A.; Petrovic, T.; Sekler, M.; Tesovic, B.; Vidanovic, D.; Volkening, J. |
| EPI_ISL_693238, EPI_ISL_693239, EPI_ISL_693247 | Secao Centro de Diagnostico Secedi | Instituto Adolfo Lutz, Interdiciplinary Procedures Center, Strategic Laboratory | Claudia Regina Gonçalves; Claudio Tavares Sacchi; Erica Valessa Ramos Gomes; Karoline Rodrigues Campos |
| EPI_ISL_412026 | Second Hospital of Anhui Medical University | Second Hospital of Anhui Medical University | Changtai Wang; Mengji Lu; Mengyuan Xua; Tengfei He; Xin Huang; Zhenhua Zhang; Zhongping Liua; Zixiang Chen |
| EPI_ISL_469253 | Second Military Medical University, Department of Microbiology | Second Military Medical University, Department of Microbiology | Jiang, L.; Peng, H.; Qi; Tang, H.; Z. and Zhao, P. |
| EPI_ISL_735405 | Secretaria Minucipal de Saude de Birigui | Instituto Adolfo Lutz, Interdiciplinary Procedures Center, Strategic Laboratory | Claudia Regina Gonçalves; Claudio Tavares Sacchi; Erica Valessa Ramos Gomes; Karoline Rodrigues Campos |
| EPI_ISL_708530 | Secretaria Municipal de Saude de Fernandópolis | Instituto Adolfo Lutz, Interdiciplinary Procedures Center, Strategic Laboratory | Carlos Henrique Camargo; Claudia Regina Gonçalves; Claudio Tavares Sacchi; Erica Valessa Ramos Gomes; Fernanda Modesto Tolentino Binhardi; Janaina Other Martins Montanha; Karoline Rodrigues Campos; Marcia Maria Costa Nunes Soares; Maricelia Navarro Pinheiro Flores |
| EPI_ISL_468320 | Secretaria Municipal de Saude de Hortolandia | Instituto Adolfo Lutz, Interdiciplinary Procedures Center, Strategic Laboratory | Claudia Regina Gonçalves; Claudio Tavares Sacchi; Erica Valessa Ramos Gomes |
| EPI_ISL_574583 | Secretaria Municipal de Saude de Jandira | Instituto Adolfo Lutz, Interdiciplinary Procedures Center, Strategic Laboratory | Claudia Regina Gonçalves; Claudio Tavares Sacchi; Erica Valessa Ramos Gomes; Karoline Rodrigues Campos |
| EPI_ISL_574597 | Secretaria Municipal de Saude de Jarinu | Instituto Adolfo Lutz, Interdiciplinary Procedures Center, Strategic Laboratory | Claudia Regina Gonçalves; Claudio Tavares Sacchi; Erica Valessa Ramos Gomes; Karoline Rodrigues Campos |
| EPI_ISL_547576, EPI_ISL_603035 | Secretaria Municipal de Saúde | Instituto Adolfo Lutz, Interdiciplinary Procedures Center, Strategic Laboratory | Claudia Regina Gonçalves; Claudio Tavares Sacchi; Erica Valessa Ramos Gomes; Karoline Rodrigues Campos |
| EPI_ISL_693221, EPI_ISL_693222 | Secretaria Municipal de Saúde de Birigui | Instituto Adolfo Lutz, Interdiciplinary Procedures Center, Strategic Laboratory | Claudia Regina Gonçalves; Claudio Tavares Sacchi; Erica Valessa Ramos Gomes; Karoline Rodrigues Campos |
| EPI_ISL_693215 | Secretaria Municipal de Saúde de Iracemapolis | Instituto Adolfo Lutz, Interdiciplinary Procedures Center, Strategic Laboratory | Claudia Regina Gonçalves; Claudio Tavares Sacchi; Erica Valessa Ramos Gomes; Karoline Rodrigues Campos |
| EPI_ISL_693228 | Secretaria Municipal de Sorocaba | Instituto Adolfo Lutz, Interdiciplinary Procedures Center, Strategic Laboratory | Claudia Regina Gonçalves; Claudio Tavares Sacchi; Erica Valessa Ramos Gomes; Karoline Rodrigues Campos |
| EPI_ISL_471542 | Secretaria de Saude de Mogi das Cruzes | Instituto Adolfo Lutz, Interdiciplinary Procedures Center, Strategic Laboratory | Claudia Regina Gonçalves; Claudio Tavares Sacchi; Erica Valessa Ramos Gomes |
| EPI_ISL_708529 | Secretária Municipal de Saude de Fernandópolis | Instituto Adolfo Lutz, Interdiciplinary Procedures Center, Strategic Laboratory | Carlos Henrique Camargo; Claudia Regina Gonçalves; Claudio Tavares Sacchi; Erica Valessa Ramos Gomes; Fernanda Modesto Tolentino Binhardi; Janaina Other Martins Montanha; Karoline Rodrigues Campos; Marcia Maria Costa Nunes Soares; Maricelia Navarro Pinheiro Flores |
| EPI_ISL_593902 | Sentinelles, Limay | National Reference Center for Viruses of Respiratory Infections, Institut Pasteur, Paris | Etienne Simon-Lorière; Fabiana Gambaro; Maud Vanpeene; Sylvie Behillil; Sylvie van der Werf; Vincent Enouf |
| EPI_ISL_583502 | Serv de Vig Sanitaria Epidemio e CTRL de Zoonoses Guaruja | Instituto Adolfo Lutz, Interdiciplinary Procedures Center, Strategic Laboratory | Claudia Regina Gonçalves; Claudio Tavares Sacchi; Erica Valessa Ramos Gomes; Karoline Rodrigues Campos |
| EPI_ISL_420600, EPI_ISL_778843 | Servicio Virosis Respiratorias-Departamento Virología-INEI | Instituto Nacional Enfermedades Infecciosas C.G.Malbran | Avaro M.; Baumeister E.; Benedetti E.; Campos J.; Cisterna D.; Dattero ME; Lorenzo F.; Molina V.; Perandones C.; Poklepovich T.; Pontoriero A.; Russo M.; Tuduri E. |
| EPI_ISL_436316, EPI_ISL_436337 | Servicio de Microbiologia. Hospital Clinico Universitario de Valencia | Sequencing and Bioinformatics Service and Molecular Epidemiology Research Group. FISABIO-Public Health | Beatriz Beamud; David Navarro; Fernando Gonzalez-Candelas; Giuseppe D'Auria; Griselda De Marco; Inma Galán Vendrell; Ivan Ansari; Lidia Ruiz Roldan; Llúcia Martínez-Priego; Loreto Ferrús Abad; Maria Alma Bracho; Mariana Reyes-Prieto; Marta Pla Diaz; Neris Garcia-Gonzalez; Paula Ruiz-Hueso; Sandra Carbo; Vicente Soriano Chirona |
| EPI_ISL_510377, EPI_ISL_654462 | Servicio de Microbiología, Hospital Miguel Servet, Zaragoza | SeqCOVID-SPAIN consortium/IBV(CSIC) | Alexander Tristancho Baró; Ana Milagro; Antonio Rezusta López; Nieves Martínez Cameo and SeqCOVID-SPAIN consortium; Yolanda Gracia Grataloup |

| | | | |
|---|---|---|---|
| EPI_ISL_654630 | Servicio de Microbiología, Hospital Universitario Central de Asturias | SeqCOVID-SPAIN consortium/IBV(CSIC) | Cristián Castelló Abietar; Jose A. Boga; Marta Elena Álvarez-Argüelles; Santiago Melón and SeqCOVID-SPAIN consortium; Susana Rojo-Alba |
| EPI_ISL_691663, EPI_ISL_779882, EPI_ISL_779888, EPI_ISL_779899, EPI_ISL_779902, EPI_ISL_779921 | Servicio de Microbiología, Hospital Universitario Son Espases | SeqCOVID-SPAIN consortium/IBV(CSIC) | Antonio Oliver and SeqCOVID-SPAIN consortium; Carla López-Causapé; Jordi Reina |
| EPI_ISL_654537, EPI_ISL_660303, EPI_ISL_671836, EPI_ISL_683607, EPI_ISL_683646 | Servicio de Microbiología, Laboratori Clínic Metropolitana Nord. Hospital Universitari Germans Trias i Pujol. Institut d'Investigació en Ciències de la Salut Germans Trias i Pujol (IGTP) | SeqCOVID-SPAIN consortium/IBV(CSIC) | Adrián Antuori; Anabel Fernández; Anna Not; Antoni E. Bordoy; Cristina Casañ and SeqCOVID-SPAIN consortium; Elisa Martró; Nona Romaní; Nona Romaní and SeqCOVID-SPAIN consortium; Verónica Saludes |
| EPI_ISL_419681 | Servicio de Microbiología. Consorcio Hospital General Universitario de Valencia | Sequencing and Bioinformatics Service and Molecular Epidemiology Research Group. FISABIO-Public Health | Concepcion Gimeno; Fernando Gonzalez-Candelas; Giuseppe D'Auria; Griselda De Marco; Maria Alma Bracho; Maria Dolores Ocete; Neris Garcia-Gonzalez |
| EPI_ISL_509641, EPI_ISL_509651, EPI_ISL_582070 | Servicio de Microbiología. Hospital Universitario Donostia. OSI Donostialdea. Área de Enfermedades Infecciosas, Grupo de Infección Respiratoria y Resistencia Antimicrobiana. Instituto de Investigación Sanitaria Biodonostia | SeqCOVID-SPAIN consortium/IBV(CSIC) | Gustavo Cilla; Jose Maria Marimón and SeqCOVID-SPAIN consortium; Luis Piñeiro; Milagrosa Montes |
| EPI_ISL_468868 | Servicio de Microbiología. Hospital Universitario Donostia. OSI Donostialdea. Área de Enfermedades Infecciosas, Grupo de Infección Respiratoria y Resistencia Antimicrobiana. Instituto de Investigación Sanitaria Biodonostia. | SeqCOVID-SPAIN consortium/IBV(CSIC) | Gustavo Cilla; Jose Maria Marimón and SeqCOVID-SPAIN consortium; Luis Piñeiro; Milagrosa Montes |
| EPI_ISL_574598 | Servico de Verificacao de Obito SVO | Instituto Adolfo Lutz, Interdiciplinary Procedures Center, Strategic Laboratory | Claudia Regina Gonçalves; Claudio Tavares Sacchi; Erica Valessa Ramos Gomes; Karoline Rodrigues Campos |
| EPI_ISL_529003 | Servizio di igiene epidemiologia e sanità pubblica (SIESP)-Chieti | Istitituto Zooprofilattico Sperimentale dell'Abruzzo e Molise "G.Caporale" | Ancora M; Cammà C; Curini V; Di Domenico M; Di Pasquale A; Lorusso A; Mangone I; Marcacci M; Puglia I; Rinaldi A; Savini G. |
| EPI_ISL_534315, EPI_ISL_583495 | Serviço de Verificação de Óbitos SVO Guaruulhos | Instituto Adolfo Lutz, Interdiciplinary Procedures Center, Strategic Laboratory | Claudia Regina Gonçalves; Claudio Tavares Sacchi; Erica Valessa Ramos Gomes; Karoline Rodrigues Campos |
| EPI_ISL_515543 | Serviço de Vigilância Sanitária e Epidemiológica | Instituto Adolfo Lutz, Interdiciplinary Procedures Center, Strategic Laboratory | Claudia Regina Gonçalves; Claudio Tavares Sacchi; Erica Valessa Ramos Gomes |
| EPI_ISL_414936, EPI_ISL_414941 | Shandong Provincial Center for Disease Control and Prevention | Beijing Institute of Microbiology and Epidemiology | Bo Pang; Can-Fang Lin; Cun-Bao Li; Dian-Ming Kang; Feng Gao; Guo-Lin Wang; Jie Lei; Li-Jun Duan; Lin Yao; Mai-Juan Ma; Ming-Xiao Yao; Sheng-Xiang Ji; Wen-Kui Sun; Xiang-Na Zhao; Xiao Wei; Xiao-Li Zhang; Xiao-Lin Jiang; Yang Hang; Zeng-Qiang Kou |
| EPI_ISL_416319, EPI_ISL_416334, EPI_ISL_416389, EPI_ISL_416394, EPI_ISL_416403 | Shanghai Public Health Clinical Center, Shanghai Medical College, Fudan University | National Research Center for Translational Medicine (Shanghai), Ruijin Hospital affiliated to Shanghai Jiao Tong University School of Medicine & Shanghai Public Health Clinical Center | Gang Lu; Hongzhou Lu; Saijuan Chen; Shengyue Wang; Xiaonan Zhang; Yun Ling; Yun Tan |
| EPI_ISL_497950 | Shaoxing CDC | Zhejiang Provincial Center for Disease Control and Prevention | Haiyan Mao; Hao Yan; Junhang Pan; Wen Shi; Xiuyu Lou; Yanjun Zhang; Yi Sun; Yin Chen; Zhen Li |
| EPI_ISL_463889, EPI_ISL_463896, EPI_ISL_463901 | Shaoxing Center for Disease Control and Prevention | Department of Pathology and Laboratory Medicine, University of California Los Angeles | Evann E. Hilt; Fan Li; Huan Wu; JiLing Wang; Jialiang Tang; Jinkun Chen; QinChao Zhang; Shangxin Yang; Yifang Wang; Zhuojing Jiang; Ziqin Li |
| EPI_ISL_636117, EPI_ISL_636118 | Sharp HealthCare Laboratory | Andersen lab at Scripps Research | Cathy Woerle; Jacquelyn Berumen; Liam McGinnis; SEARCH Alliance San Diego with Aaron Harding |
| EPI_ISL_582126 | Sheikh Khalifa Medical City | Molecular Surveillance lab Sheikh Khalifa Medical City | Amirtharaj Francis; Hala Imambaccus; Hiba Saud; Sahar Almarzooqi; Sajeed Abdul; Stefan Weber |
| EPI_ISL_582608, EPI_ISL_582620, EPI_ISL_582625, EPI_ISL_582642, EPI_ISL_582643, EPI_ISL_582659, EPI_ISL_582662, EPI_ISL_582674, EPI_ISL_582688 | | | |
| see above | Sheikh Khalifa Medical City | Molecular/Surveillance lab Sheikh Khalifa Medical City | Amirtharaj Francis; Hala Imambaccus; Hiba Saud; Sahar Almarzooqi; Sajeed Abdul; Stefan Weber |
| EPI_ISL_482695, EPI_ISL_482696, EPI_ISL_509394, EPI_ISL_768324 | Singapore General Hospital | Department of Microbiology | Chenhao Li; Chenhao Li; Karrie Ko; Karrie Ko; Kenneth Xin Long Chan; Kern Rei Chng; Kern Rei Chng; Kian Sing Chan; Kian Sing Chan; Kun Lee Lim; Kun Lee Lim; Lynette Oon; Lynette Oon; Niranjan Nagarajan; Niranjan Nagarajan; Nurdyana Abdul Rahman; Nurdyana Abdul Rahman; Sui Sin Goh |
| EPI_ISL_410536 | Singapore General Hospital, Molecular Laboratory, Division of Pathology | Programme in Emerging Infectious Diseases, Duke-NUS Medical School | Danielle E Anderson; Gavin JD Smith; Jayanthi Jayakumar; Jenny GH Low; Kian Sing Chan; Lynette LE Oon; Martin Linster; Shirin Kalimuddin; Yan Zhuang; Yvonne CF Su |
| EPI_ISL_756375 | Siriraj Hospital, Mahidol University | National Institute of Health, Department of Medical Sciences, Ministry of Public Health, Thailand | Archiraya Pattama; Chutikarn Chaimayo; Natchaya Khiadsang; Navin Horthongkham; Niracha Athipanyasilp; Pakorn Piromtong; Pilailuk Okada; Ratana Tacharoenmuang; Ruengpung Sutthent; Sirikanda wimol; Siripaporn Phuygun; Sittiporn Parnmen; Sunthareeya Waicharoen; Thanutsapa Thanadachakul; Wannee Kantakamalakul.; Warawan Wongboot |
| EPI_ISL_534227 | Skanes universitetssjukhus Lund | The Public Health Agency of Sweden | Anna Risberg; Anna-Malin Linde; Karin Tegmark-Wisell; Maria Lind Karlberg; Mattias Haukland; Mia Brytting; Olov Svartstrom; Oskar Karlsson Lindsjo; Petra Edquist; Reza Advani; Sandra Broddesson |
| EPI_ISL_475108, EPI_ISL_654956 | Skovde/Unilabs | The Public Health Agency of Sweden | Anna Risberg; Anna-Malin Linde; Karin Tegmark-Wisell; Maria Lind Karlberg; Mattias Haukland; Mia Brytting; Olov Svartstrom; Oskar Karlsson Lindsjo; Petra Edquist; Reza Advani; Sandra Broddesson; Shamam Muradrasoli |
| EPI_ISL_408431 | Sorbonne Université, Inserm et Assistance Publique-Hôpitaux de Paris (Pitié Salpétrière) | National Reference Center for Viruses of Respiratory Infections, Institut Pasteur, Paris | Angela Brisebarre; Anne-Geneviève Marcelin; David Boutolleau; Elise Klément; Eric Caumes.; Flora Donati; Marion Barbet; Maud Vanpeene; Mélanie Albert; Méline Bizard; Sonia Burrel; Sylvie Behillil; Sylvie van der Werf; Valérie Pourcher; Vincent Calvez; Vincent Enouf |
| EPI_ISL_710126 | South Eastern Area Laboratory Services (SEALS) | CIDM-PH et al. | CIDM-PH et al. |
| EPI_ISL_490031, EPI_ISL_490041, EPI_ISL_667797, EPI_ISL_667801, EPI_ISL_767886 | South Eastern Area Laboratory Services (SEALS) | NSW Health Pathology - Institute of Clinical Pathology and Medical Research; Westmead Hospital; University of Sydney | CIDM-PH et al. |
| EPI_ISL_456347, EPI_ISL_579175 | Southern Community Labs Dunedin | Institute of Environmental Science and Research (ESR) | Anja Werno; Antje van der Linden; Arlo Upton; Chris Mansell; David Hammer; Dragana Drinkovic; Erasmus Smit; Gary McAuliffe; Hana Sofia Andersson; Hermes Perez; James Ussher; Jill Sherwood; Jing Wang; Joep de Ligt; Josh Freeman; Julia Howard; Juliet Elvy; Lauren Jelly; Mary DeAlmeida; Matt Blakiston; Matt Storey; Matthew Rogers; Max Bloomfield; Michael Addidle; Michelle Balm; Muhammad Faisal; Nikki Freed; Olin Silander; Sally Roberts; Sarah Jefferies; Sharmini Muttaiyah; Susan Morpeth; Susan Taylor; Timothy Blackmore; Vani Sathyendran; Veronica Playle; Virginia Hope; Xiaoyun Ren |
| EPI_ISL_469252 | Special Infectious Agents Unit | Special Infectious Agents Unit | A.M.; Al-Sobahy; Azhar; E.I.; El-Kafrawy; Farraj; Hassan; N.A.; S.A.; T.L.; Tolah; Uthman |
| EPI_ISL_450352 | St.Olavs hospital/NTNU | Institute of Genomics Core Facility, University of Tartu | Aleksandr Ianevski; Denis Kainov; Janne-Fossum Malmring; Svein Arne Nordbø; Tuuli Reisberg |
| EPI_ISL_596652 | St.Vincent's University Hospital | St.Vincent's University Hospital | Aisling Purcell; Gabriel Gonzalez; Guerrino Macori; Kirsten Schaffer; Lynda Fenelon; Mary Lucey; Niamh Mullane; Suzie Coughlan; Séamus Fanning; Una Sutton-Fitzpatrick |
| EPI_ISL_418991 | State Key Laboratory for Diagnosis and Treatment of Infectious Diseases, National Clinical Research Center for Infectious Diseases, First Affiliated Hospital, Zhejiang University School of Medicine, Hangzhou, China 310003 | State Key Laboratory for Diagnosis and Treatment of Infectious Diseases, National Clinical Research Center for Infectious Diseases, First Affiliated Hospital, Zhejiang University School of Medicine, Hangzhou, China 310003 | Changzhong Jin; Chao Jiang; Fumin Liu; Haibo Wu; Hangping Yao; Lanjuan Li; Linfang Cheng; Min Zheng; Nanping Wu; Xiangyun Lu; Zhigang Wu |
| EPI_ISL_414687 | State Key Laboratory of Respiratory Disease, National Clinical Research Center for Respiratory Disease, Guangzhou Institute of Respiratory Health, the First Affiliated Hospital of Guangzhou Medical University | the First Affiliated Hospital of Guangzhou Medical University & BGI-Shenzhen | Zhao et al |
| EPI_ISL_752938 | State Laboratories Division, Hawaii State Department of Health | State Laboratories Division, Hawaii State Department of Health | Drew Kuwazaki; Edward Desmond; Pamela O'Brien; Razvan Sultana; Sabrina Diemert |
| EPI_ISL_428869, EPI_ISL_428887, EPI_ISL_428891, EPI_ISL_428901 | State Research Center of Virology and Biotechnology VECTOR, Department of Collection of Microorganisms | State Research Center of Virology and Biotechnology VECTOR, Department of Collection of Microorganisms | Alexander N. Shvalov; Elena V. Gavrilova; Oleg V. Pyankov; Rinat A. Maksyutov; Sergey A. Bodnev; Tatyana V. Tregubchak |
| EPI_ISL_451658 | State Sanitary Inspectorate | Laboratory of Recombinant Vaccines | Boguslaw Szewczyk; Jaroslaw Pinkas; Krystyna Bienkowska-Szewczyk; Lukasz Rabalski |
| EPI_ISL_626341, EPI_ISL_626346, EPI_ISL_626347 | Statens Serum Institute | Statens Serum Institute | A.S.; Afaro-Nunez, A.; Belsham; C.S.; Fonager, J.; Foomsgaard, A.; G.J. and Botner, A.; Halasa, T.; Hammer; Jorgensen; Lohse, L.; M.L.; M.W.; Mundbjerg, K.; Quaade; Rasmussen; Rasmussen, M.; Rosenstierne; Strandbygaard, B.; T.B. |
| EPI_ISL_491036, EPI_ISL_491037, EPI_ISL_491039, EPI_ISL_491048, EPI_ISL_491053, EPI_ISL_491055, EPI_ISL_491070, EPI_ISL_491075, EPI_ISL_491085, EPI_ISL_491088 | | | |
| see above | Suceava County Emergency Hospital | "Stefan cel Mare" University Metagenomics Lab | Antoniadis Panagiotis et al.; Lobiuc Andrei; Lobiuc Andrei et al. |
| EPI_ISL_747235 | Sukabumi Health Laboratory | West Java Health Laboratory; School of Life Sciences and Technology, Institut Teknologi Bandung | Azzania Fibriani; Cut Nur Cinthia Alamanda; Ema Rahmawati; Isak Solihin; Karimatu Khoirunnisa; Miftahul Faridl; Rifky Waluyajati Rachman; Rini Robiani; Ryan Bayusantika Ristandi |

| | | | |
|---|---|---|---|
| EPI_ISL_560986 | Sundsvalls sjukhus | The Public Health Agency of Sweden | Anna Risberg; Anna-Malin Linde; Karin Tegmark-Wisell; Maria Lind Karlberg; Mattias Haukland; Mia Brytting; Olov Svartstrom; Oskar Karlsson Lindsjo; Petra Edquist; Reza Advani; Sandra Broddesson |
| EPI_ISL_767940 | Sydney South West Pathology Service (SSWPS) - Liverpool Hospital - NSW Health Pathology | NSW Health Pathology - Institute of Clinical Pathology and Medical Research; Westmead Hospital; University of Sydney | CIDM-PH et al. |
| EPI_ISL_767854, EPI_ISL_767860, EPI_ISL_767863, EPI_ISL_803112 | Sydney South West Pathology Service (SSWPS) - Royal Prince Alfred Hospital - NSW Health Pathology | NSW Health Pathology - Institute of Clinical Pathology and Medical Research; Westmead Hospital; University of Sydney | CIDM-PH et al. |
| EPI_ISL_768522 | Synphaet Hospital | National Institute of Health, Department of Medical Sciences, Ministry of Public Health, Thailand | ; Natchaya Khiadsang; Pakorn Piromtong; Pilailuk Okada; Ratana Tacharoenmuang; Siripaporn Phuygun; Sittiporn Parnmen; Sunthareeya Waicharoen; Thanutsapa Thanadachakul; Warawan Wongboot; sirikanda wimol |
| EPI_ISL_682261 | TAMIZAJE COMUNITARIO- PASO CANOAS | Inciensa, Instituto Costarricense de Investigación y Enseñanza en Nutrición y Salud | Adriana Godinez & Melany Calderon; Claudio Soto-Garita; Estela Cordero; Francisco Duarte; Hebleen Porras |
| EPI_ISL_695536 | TGen North | TGen North | Ashlyn Pfeiffer; Chris French; Darrin Lemmer; Dave Engelthaler; Hayley Yaglom; Jolene Bowers; Megan Folkerts; The Arizona COVID Genomics Union (ACGU) |
| EPI_ISL_447252, EPI_ISL_457726 | TSGH-CP molecular lab | TSGH-CP molecular lab | Cherng-Lih Perng; Chien-Wen Chen; Chih-Kai Chang; Feng-Yee Chang; Hsing-Yi Chung; Hung-Sheng Shang; Jung-Chung Lin; Kuo-Ming Yeh; Kuo-Sheng Hung; Ming-Jr JIAN; Sheng-Kang Chiu; Shih-Hung Tsai; Tien-Yao Chang |
| EPI_ISL_412968 | Takayuki Hishiki Kanagawa Prefectural Institute of Public Health | Takayuki Hishiki Kanagawa Prefectural Institute of Public Health | An, T.; Hayashizaki; Hishiki, T.; Kawai, J.; Kogo, Y.; Matsuki, Y.; Sakuragi, J.; Suzuki, R.; Tanaka, Y.; Usui, K.; Y. and Takasaki, T. |
| EPI_ISL_733572, EPI_ISL_733574, EPI_ISL_733575, EPI_ISL_733576, EPI_ISL_759892, EPI_ISL_759893 | Temporary Specimen Collection Centre | Hong Kong Department of Health | Alan K.L. Tsang; Dominic N.C. Tsang; Edman T.K. Lam; Peter C.W. Yip; Rickjason C.W. Chan |
| EPI_ISL_450442 | The Department of Infectious Disease Prevention and Control, Henan Provincial Center for Disease Control and Prevention | The Department of Infectious Disease Prevention and Control, Henan Provincial Center for Disease Control and Prevention | Hu, X.; Huang; Li, D.; Li, X.; Lu, S.; Wu, B.; X. and Guo, W. |
| EPI_ISL_429086, EPI_ISL_429092, EPI_ISL_429102 | The First Affiliated Hospital of Guangzhou Medical University | BGI-shenzhen & The First Affiliated Hospital of Guangzhou Medical University | ; Daxi Wang; Lu Zhang; Wanying Sun; Yanqun Wang; Zhaoyong Zhang et al. |
| EPI_ISL_457692, EPI_ISL_457698 | The First Affiliated Hospital of Guangzhou Medical University, Guangzhou, China | BGI-shenzhen & The First Affiliated Hospital of Guangzhou Medical University | Daxi Wang; Lu Zhang; Wanying Sun; Yanqun Wang; Zhaoyong Zhang et al. |
| EPI_ISL_541333, EPI_ISL_545957, EPI_ISL_547967, EPI_ISL_577623, EPI_ISL_577633, EPI_ISL_577636, EPI_ISL_577637, EPI_ISL_584073, EPI_ISL_584074, EPI_ISL_584078, EPI_ISL_584080, EPI_ISL_584082, EPI_ISL_626582, EPI_ISL_626585, EPI_ISL_626599, EPI_ISL_626605, EPI_ISL_660554, EPI_ISL_660584, EPI_ISL_660588, EPI_ISL_737015, EPI_ISL_792685, EPI_ISL_792695, EPI_ISL_792699, EPI_ISL_792706, EPI_ISL_792707 | | | |
| see above | The National Institute of Public Health | State Veterinary Institute Prague | A; D; H; J; Jirincova; L; Nagy; Novakova; Trnka; Vecerova |
| EPI_ISL_471528, EPI_ISL_471550 | The National Institute of Public Health | State Veterinary Institute Prague and The National Institute of Public Health | A; D; H; J; Jirincova; L; Nagy; Novakova; Trnka; Vecerova |
| EPI_ISL_541335 | The National Institute of Public Health | Sídlištní 136/24 165 03, Prague Czech Republic | A; D; H; J; Jirincova; L; Nagy; Novakova; Trnka; Vecerova |
| EPI_ISL_437519 | The National Institute of Public Health Center for Epidemiology and Microbiology | The National Institute of Public Health Center for Epidemiology and Microbiology | Alexander Nagy; Dusan Trnka; Helena Jirincova; Jaromira Vecerova; Ludmila Novakova |
| EPI_ISL_417701, EPI_ISL_417765, EPI_ISL_417788, EPI_ISL_424458, EPI_ISL_424556 | The National University Hospital of Iceland | deCODE genetics | Agnar Helgason; Alma Moller; Arna B Agustsdottir; Arnaldur Gylfason; Asgeir Sigurdsson; Aslaug Jonasdottir; Berglind Eiriksdottir; Bjarni Thorbjornsson; Brynjar O Jensson; Daniel F Gudbjartsson; Droplaug N Magnusdottir; Elisabet E Gardarsdottir; Emil A Thorarensen; Gardar Sveinbjornsson; Gisli Masson; Gudmundur Georgsson; Gudmundur L Norddahl; Gudrun Sigmundsdottir; Hakon Jonsson; Hilma Holm; Ingileif Jonsdottir; Jona Saemundsdottir; Kamilla S Josefsdottir; Kari Stefansson; Karl G Kristinsson; Kjartan R Gudmundsson; Kristin E Sveinsdottir; Louise le Roux; Maney Sveinsdottir; Olafia S Gretarsdottir; Olafur T Magnusson; Pall Melsted; Patrick Sulem; Run Fridriksdottir; Thora R Gunnarsdottir; Thordur Kristjansson; Thorolfur Gudnason; Unnur Thorsteinsdottir |
| EPI_ISL_648182, EPI_ISL_648184, EPI_ISL_648190, EPI_ISL_648194, EPI_ISL_648196, EPI_ISL_654500, EPI_ISL_710581 | | | |
| see above | The Public Health Agency of Sweden | The Public Health Agency of Sweden | Anna Risberg; Anna-Malin Linde; Department of Microbiology; Karin Tegmark-Wisell; Maria Lind Karlberg; Mattias Haukland; Mia Brytting; Olov Svartstrom; Oskar Karlsson Lindsjo; Petra Edquist; Reza Advani; Sandra Broddesson; The Public Health Agency of Sweden |
| EPI_ISL_754233, EPI_ISL_754235 | The Republican Research and Practical Center for Epidemiology and Microbiology (RRPCEM) | WHO National Influenza Centre Russian Federation | Anatoly Krasko; Andrey Komissarov; Anna Ivanova; Artem Fadeev; Daria Danilenko; Dmitry Bazhenov; Dmitry Lioznov; Elena Gasich; Elena Nabieva; Georgii Bazykin; Kirill Bulda; Ksenia Safina; Kseniya Komissarova |
| EPI_ISL_779711 | The University Hospital Brno | Institute of Applied Biotechnologies a.s. | Martin Kašný; Ondřej Brzoň; Petr Klempt; Petr Kvapil |
| EPI_ISL_406030 | The University of Hong Kong - Shenzhen Hospital | Li Ka Shing Faculty of Medicine, The University of Hong Kong | C.C.-Y.; C.K.-M. and Yuen; Cai; Chan; Chen, H.; Cheng; Chu, H.; H.W.; Hui; Ip; J.D.; J.F.-W.; J.P.; K.H.; K.K.-W.; K.Y.; Kok; Liu, J.; Lo; Poon; R.W.-S.; S.K.-F.; To; Tsai; V.C.-C.; V.K.-M.; W.M.; Xing, F.; Yang, J.; Yip; Yuan, S. |
| EPI_ISL_412983 | Tianmen Center for Disease Control and Prevention | Hubei Provincial Center for Disease Control and Prevention | Bin Fang; Bo Yang; Bo Yu; Faxian Zhan; Guojun Ye; Jing Li; Junqiang Xu; Kun Cai; Linlin Liu; Xiang Li; Xiao Yu; Xierong Li; Xixiang Huo; Yangyang Tao; YiFa Zhu; Yongzhong Jiang. |
| EPI_ISL_463991, EPI_ISL_586319, EPI_ISL_591199, EPI_ISL_755672, EPI_ISL_755745, EPI_ISL_755748, EPI_ISL_755758, EPI_ISL_755768, EPI_ISL_755802, EPI_ISL_755806, EPI_ISL_755811, EPI_ISL_755813, EPI_ISL_755884, EPI_ISL_755938, EPI_ISL_792087, EPI_ISL_792088, EPI_ISL_792090 | | | |
| see above | Toronto Invasive Bacterial Diseases Network | McMaster University | Ahmed Draia; Allison McGeer; Andrew G. McArthur; Angel Li; Emily Panousis; Hooman Derakhshani; Jalees Nasir; Kuganya Nirmalarajah; Michael Surette; Patryk Aftanas; Samira Mubareka |
| EPI_ISL_480201 | Toyama Institute of Health | Pathogen Genomics Center, National Institute of Infectious Diseases | Hajime Kamiya; Kazunori Oishi; Kentaro Itokawa; Makoto Kuroda; Masae Itamochi; Masanori Hashino; Motoi Suzuki; Rina Tanaka; Tsuyoshi Sekizuka |
| EPI_ISL_735419 | UBS Alvarenga | Instituto Adolfo Lutz, Interdiciplinary Procedures Center, Strategic Laboratory | Claudia Regina Gonçalves; Claudio Tavares Sacchi; Erica Valessa Ramos Gomes; Karoline Rodrigues Campos |
| EPI_ISL_735422 | UBS Dematchi | Instituto Adolfo Lutz, Interdiciplinary Procedures Center, Strategic Laboratory | Claudia Regina Gonçalves; Claudio Tavares Sacchi; Erica Valessa Ramos Gomes; Karoline Rodrigues Campos |
| EPI_ISL_735420 | UBS Riacho Grande | Instituto Adolfo Lutz, Interdiciplinary Procedures Center, Strategic Laboratory | Claudia Regina Gonçalves; Claudio Tavares Sacchi; Erica Valessa Ramos Gomes; Karoline Rodrigues Campos |
| EPI_ISL_735421 | UBS Sta Terezinha | Instituto Adolfo Lutz, Interdiciplinary Procedures Center, Strategic Laboratory | Claudia Regina Gonçalves; Claudio Tavares Sacchi; Erica Valessa Ramos Gomes; Karoline Rodrigues Campos |
| EPI_ISL_693227 | UBS Vila Marchi | Instituto Adolfo Lutz, Interdiciplinary Procedures Center, Strategic Laboratory | Claudia Regina Gonçalves; Claudio Tavares Sacchi; Erica Valessa Ramos Gomes; Karoline Rodrigues Campos |
| EPI_ISL_523963 | UBS Vila Silvia | Instituto Adolfo Lutz, Interdiciplinary Procedures Center, Strategic Laboratory | Claudia Regina Gonçalves; Claudio Tavares Sacchi; Erica Valessa Ramos Gomes |
| EPI_ISL_471648 | UBS e Pronto Socorro Jd. Jacira | Instituto Adolfo Lutz, Interdiciplinary Procedures Center, Strategic Laboratory | Claudia Regina Gonçalves; Claudio Tavares Sacchi; Erica Valessa Ramos Gomes |
| EPI_ISL_483520 | UC San Diego Center for Advanced Laboratory Medicine | Andersen lab at Scripps Research | Ji H Shin; SEARCH Alliance San Diego with David Pride |
| EPI_ISL_414487, EPI_ISL_414586 | UCD National Virus Reference Laboratory | UCD National Virus Reference Laboratory | Alison Murphy; Brendan Loftus; Cillian F De Gascun; Gabriel Gonzalez; Jeff Connell; Jonathan Dean; Ken Wolfe; Kevin Byrne; Michael Carr; Suzie Coughlan |
| EPI_ISL_486345 | UCSF Clinical Microbiology Laboratory | Chan-Zuckerberg Biohub | CZB Cliahub Consortium |
| EPI_ISL_732195 | ULS Litoral Alentejano | Instituto Nacional de Saude (INSA) and Instituto Gulbenkian de Ciencia (IGC) | Borges et al |
| EPI_ISL_747487 | ULSS 5 Polesana | Istituto Zooprofilattico Sperimentale delle Venezie | Adelaide Milani; Alessia Schivo; Alice Fusaro; Ambra Pastori; Annalisa Salviato; Antonia Ricci; Bianca Zecchin; Calogero Terregino; Erika Giorgia Quaranta; Isabella Monne |
| EPI_ISL_452185 | ULSS9 Distretto di Bussolengo | Istituto Zooprofilattico Sperimentale delle Venezie | Adelaide Milani; Alessia Schivo; Alice Fusaro; Ambra Pastori; Annalisa Salviato; Antonia Ricci; Bianca Zecchin; Calogero Terregino; Erika Giorgia Quaranta; Isabella Monne |
| EPI_ISL_733238 | UMMC-Health | WHO National Influenza Centre Russian Federation | Andrey Komissarov; Anna Ivanova; Artem Fadeev; Daria Danilenko; Dmitry Bazhenov; Dmitry Lioznov; Elena Nabieva; Georgii Bazykin; Ksenia Safina; Kseniya Komissarova; Tatiana Platonova |
| EPI_ISL_445247 | UNIVERSIDAD DE LOS ANDES | Instituto de Salud Publica de Chile | Alejandra Acevedo; Andrés E Castillo; Bárbara Parra; Carolina Tambley; Gabriel Leal; Jaime Lagos; Jorge Fernandez; Loredana Arata; Patricia Bustos; Paz Tapia; Rodrigo Fasce; Winston Andrade |
| EPI_ISL_515522 | UPA 24HS de Itatiba | Instituto Adolfo Lutz, Interdiciplinary Procedures Center, Strategic Laboratory | Claudia Regina Gonçalves; Claudio Tavares Sacchi; Erica Valessa Ramos Gomes |
| EPI_ISL_523983, EPI_ISL_523993 | UPA Campo Limpo | Instituto Adolfo Lutz, Interdiciplinary Procedures Center, Strategic Laboratory | Claudia Regina Gonçalves; Claudio Tavares Sacchi; Erica Valessa Ramos Gomes |
| EPI_ISL_603025 | UPA Central de Caraguatatuba | Instituto Adolfo Lutz, Interdiciplinary Procedures Center, Strategic Laboratory | Claudia Regina Gonçalves; Claudio Tavares Sacchi; Erica Valessa Ramos Gomes; Karoline Rodrigues Campos |
| EPI_ISL_534311 | UPA III 26 de Agosto | Instituto Adolfo Lutz, Interdiciplinary Procedures Center, Strategic Laboratory | Claudia Regina Gonçalves; Claudio Tavares Sacchi; Erica Valessa Ramos Gomes |
| EPI_ISL_583496 | UPA Jandira | Instituto Adolfo Lutz, Interdiciplinary Procedures Center, Strategic Laboratory | Claudia Regina Gonçalves; Claudio Tavares Sacchi; Erica Valessa Ramos Gomes; Karoline Rodrigues Campos |
| EPI_ISL_693237, EPI_ISL_693245 | UPA Santa Isabel | Instituto Adolfo Lutz, Interdiciplinary Procedures Center, Strategic Laboratory | Claudia Regina Gonçalves; Claudio Tavares Sacchi; Erica Valessa Ramos Gomes; Karoline Rodrigues Campos |
| EPI_ISL_523975, EPI_ISL_523980 | UPA Tito Lopes | Instituto Adolfo Lutz, Interdiciplinary Procedures Center, Strategic Laboratory | Claudia Regina Gonçalves; Claudio Tavares Sacchi; Erica Valessa Ramos Gomes |
| EPI_ISL_468316 | UPA Vila Assis | Instituto Adolfo Lutz, Interdiciplinary Procedures Center, Strategic Laboratory | Claudia Regina Gonçalves; Claudio Tavares Sacchi; Erica Valessa Ramos Gomes |
| EPI_ISL_515550 | UPA Vila Santa Catarina | Instituto Adolfo Lutz, Interdiciplinary Procedures Center, Strategic Laboratory | Claudia Regina Gonçalves; Claudio Tavares Sacchi; Erica Valessa Ramos Gomes |
| EPI_ISL_735461, EPI_ISL_756237 | UW Virology Lab | UW Virology Lab | Alexander Greninger; Hong Xie; Keith R Jerome; Lasata Shrestha; Meei-Li Huang; Pavitra Roychoudhury |
| EPI_ISL_734494, EPI_ISL_734539, EPI_ISL_734580, EPI_ISL_734612, EPI_ISL_734661, EPI_ISL_734687, EPI_ISL_734728, EPI_ISL_734747, EPI_ISL_734749, EPI_ISL_734895, EPI_ISL_734920, EPI_ISL_734991, EPI_ISL_735050, EPI_ISL_735078, EPI_ISL_735136, EPI_ISL_738220, EPI_ISL_738452 | | | |

| | | | |
|---|---|---|---|
| see above | UZ Leuven, National Reference Laboratory for Coronaviruses, Laboratory Medicine, Leuven, Belgium | KU Leuven, Rega Institute, Clinical and Epidemiological Virology | Bert Vanmechelen; Joan Marti-Carerras; Piet Maes; Tony Wawina-Bokalanga |
| EPI_ISL_693225 | Ubs Vila Rosa - Olímpia Gomes De Almeida | Instituto Adolfo Lutz, Interdiciplinary Procedures Center, Strategic Laboratory | Claudia Regina Gonçalves; Claudio Tavares Sacchi; Erica Valessa Ramos Gomes; Karoline Rodrigues Campos |
| EPI_ISL_737935, EPI_ISL_737948, EPI_ISL_737969, EPI_ISL_737973, EPI_ISL_737974, EPI_ISL_737990, EPI_ISL_738000, EPI_ISL_738001, EPI_ISL_738011, EPI_ISL_738017, EPI_ISL_738035, EPI_ISL_738040 | | | |
| see above | Uganda Central Public Health Lab and Uganda Virus Research Institute | MRC/UVRI & LSHTM Uganda Research Unit | Dan Lule Bugembe; Matthew Cotten; My V.T. Phan; Pontiano Kaleebu et al. |
| EPI_ISL_451190, EPI_ISL_451195, EPI_ISL_451198, EPI_ISL_451199 | Uganda Virus Research Institute | MRC/UVRI & LSHTM Uganda Research Unit | Beatrice Dhaala; Dan Lule Bugembe; Deogratius Ssemwanga; Henry Kyobe; Henry Mwebesa; Jane Aceng; John Kayiwa; Jonas Lexow; Julius Lutwama; Matthew Cotten; My V.T Phan; Phionah Tushabe; Pontiano Kaleebu; Stephen Balinandi |
| EPI_ISL_445240 | Ulltuna Vardcentral | The Public Health Agency of Sweden | Anna Risberg; Anna-Malin Linde; Heidi Lindback; Karin Tegmark-Wisell; Maria Lind Karlberg; Mia Brytting; Olov Svartstrom; Oskar Karlsson Lindsjo; Theresa Enkirch |
| EPI_ISL_792304 | Unidad de Virología, Centro de Educación Médica en Investigaciones Clínicas CEMIC | Área de Secuenciación del Laboratorio de Virología del Hospital de Niños Dr. Ricardo Gutierrez on behalf of 'Proyecto Argentino Interinstitucional de genomica de SARS-CoV-2' (PAIS Consortium) | AS; Echavarría; Goya; LE; Lusso; M; MI; MS; Mistchenko; Nabaes Jodar; Natale; S; Valinotto, Viegas, M. |
| EPI_ISL_735397 | Unidade Respiratória Nova Hortolandia | Instituto Adolfo Lutz, Interdiciplinary Procedures Center, Strategic Laboratory | Claudia Regina Gonçalves; Claudio Tavares Sacchi; Erica Valessa Ramos Gomes; Karoline Rodrigues Campos |
| EPI_ISL_574590 | Unidade de Printo Atendimento UPA I Santa Isabel | Instituto Adolfo Lutz, Interdiciplinary Procedures Center, Strategic Laboratory | Claudia Regina Gonçalves; Claudio Tavares Sacchi; Erica Valessa Ramos Gomes; Karoline Rodrigues Campos |
| EPI_ISL_735409 | Unidade de Pronto Atendimento Carlos Lourenco | Instituto Adolfo Lutz, Interdiciplinary Procedures Center, Strategic Laboratory | Claudia Regina Gonçalves; Claudio Tavares Sacchi; Erica Valessa Ramos Gomes; Karoline Rodrigues Campos |
| EPI_ISL_693214 | Unidade de Pronto Atendimento Central de Caraguatatuba | Instituto Adolfo Lutz, Interdiciplinary Procedures Center, Strategic Laboratory | Claudia Regina Gonçalves; Claudio Tavares Sacchi; Erica Valessa Ramos Gomes; Karoline Rodrigues Campos |
| EPI_ISL_693226 | Unidade de Pronto Atendimento Sao José | Instituto Adolfo Lutz, Interdiciplinary Procedures Center, Strategic Laboratory | Claudia Regina Gonçalves; Claudio Tavares Sacchi; Erica Valessa Ramos Gomes; Karoline Rodrigues Campos |
| EPI_ISL_735406 | Unidade de Pronto Atendimento UPA I Sta Isabel | Instituto Adolfo Lutz, Interdiciplinary Procedures Center, Strategic Laboratory | Claudia Regina Gonçalves; Claudio Tavares Sacchi; Erica Valessa Ramos Gomes; Karoline Rodrigues Campos |
| EPI_ISL_735414, EPI_ISL_735415, EPI_ISL_735417 | Unidade de Pronto Atendimento de Agenor de Campos | Instituto Adolfo Lutz, Interdiciplinary Procedures Center, Strategic Laboratory | Claudia Regina Gonçalves; Claudio Tavares Sacchi; Erica Valessa Ramos Gomes; Karoline Rodrigues Campos |
| EPI_ISL_534325 | Unidade de Vigilancia em Saude de Guarulhos | Instituto Adolfo Lutz, Interdiciplinary Procedures Center, Strategic Laboratory | Claudia Regina Gonçalves; Claudio Tavares Sacchi; Erica Valessa Ramos Gomes |
| EPI_ISL_693216, EPI_ISL_693217 | Unidade de Vigilância Epiudemiológica de Araras | Instituto Adolfo Lutz, Interdiciplinary Procedures Center, Strategic Laboratory | Claudia Regina Gonçalves; Claudio Tavares Sacchi; Erica Valessa Ramos Gomes; Karoline Rodrigues Campos |
| EPI_ISL_635153 | Unilabs Laboratory Medicine | Norwegian Institute of Public Health, Department of Virology | Hilde Elshaug; Hilde Vollan; Kamilla Heddeland Instefjord; Karoline Bragstad; Kathrine Stene-Johansen; Marie Paulsen Madsen; Olav Hungnes; Rasmus Riis Kopperud |
| EPI_ISL_412980 | Union Hospital of Tongji Medical College, Huazhong University of Science and Technology | Hubei Provincial Center for Disease Control and Prevention | Bin Fang; Bo Yang; Bo Yu; Faxian Zhan; Guojun Ye; Jing Li; Junqiang Xu; Kun Cai; Linlin Liu; Xiang Li; Xiao Yu; Xixiang Huo; Yongzhong Jiang. |
| EPI_ISL_763006, EPI_ISL_763044 | Unit 17: Influenza & Other Respiratory Viruses, German National Influenza Center | Project group Epidemiology of Highly Pathogenic Microorganisms, Robert Koch-Institute | Andreas Sachse; Ariane Düx; Djin-Ye Oh; Fabian Leendertz; Grit Schubert; Marianne Wedde; Ralf Dürrwald; Sébastien Calvignac-Spencer; Thorsten Wolff |
| EPI_ISL_411951 | Unit for Laboratory Development and Technology Transfer, Public Health Agency of Sweden | Unit for Laboratory Development and Technology Transfer, Public Health Agency of Sweden | Appelberg, S.; Bengner, M.; Brave, A.; Lind Karlberg, M.; Lindsjo, O.; Monteil, V.; Muradrasoli; Palmerus, M.; S. and Tegmark-Wisell, K. |
| EPI_ISL_464008, EPI_ISL_569954, EPI_ISL_569996, EPI_ISL_569998, EPI_ISL_570008, EPI_ISL_570019, EPI_ISL_570021, EPI_ISL_570025, EPI_ISL_609822, EPI_ISL_671653 | | | |
| see above | Unity Health Toronto | Ontario Institute for Cancer Research | Annette Gower; Bernard Lam; Felicia Vincelli; Ilinca Lungu; Jared Simpson; Jared T. Simpson; Jeremy Johns; Karel Boissinot; Larissa M. Matukas; Le Luu; Mark Downing; Paul Krzyzanowski; Philip Zuzarte; Ramzi Fattouh; Richard de Borja; Samira Mubareka; TIBDN; Trina Otterman; Wai Sum Siu; Yan Chen; Zhi Cui |
| EPI_ISL_645213 | Unité des Virus Émergents | CNR Virus des Infections Respiratoires - France SUD | Antoine Nougairède; Antonin Bal; Bruno Lina; Gregory Destras; Gwendolyne Burfin; Hadrien Règue; Laetitia Ninove; Laurence Josset; Léa Luciani; Martine Valette; Quentin Semanas |
| EPI_ISL_525467, EPI_ISL_525468 | Universidad Iberoamericana | International Centre for Genetic Engineering and Biotechnology (ICGEB) and ARGO Open Lab Platform | Alejandro Vallejo Degaudenzi; Danilo Licastro; Eileen Riego; Leandro Tapia; Patricia Leon; Robert Paulino-Ramirez; Simeone Dal Monego; Sreejith Rajasekharan and Alessandro Marcello.; Victor Virgilio Calderon |
| EPI_ISL_523811 | Universidad Iberoamericana, Instituto de Medicina Tropical & Salud Global | International Centre for Genetic Engineering and Biotechnology (ICGEB) and ARGO Open Lab Platform | Alejandro Vallejo Degaudenzi; Danilo Licastro; Eileen Riego; Leandro Tapia; Robert Paulino-Ramirez; Simeone Dal Monego; Sreejith Rajasekharan and Alessandro Marcello.; Victor Virgilio Calderon |
| EPI_ISL_539783, EPI_ISL_539784 | Universidad Regional Amazonica IKIAM | Institute of Microbiology, Universidad San Francisco de Quito | Andrea Carrera; Belén Prado-Vivar; Bernardo Gutiérrez; Carolina Proaño-Bolaños; Fabian Aguilar; Gabriel Trueba; Giovanna Moran; Juan José Guadalupe; Katherine Apunte; Marcelo Ortiz; Michelle Grunauer; Monica Becerra-Wong; Nina Espinoza de los Monteros; Patricio Rojas-Silva; Paúl Cárdenas; Sonia Sislema; Sully Márquez; Verónica Barragán; Yeimy Rojas |
| EPI_ISL_445219 | Universidad del Valle, Laboratorio de Microbiologia, VIREM | Universidad del Valle, Universidad Nacional de Colombia-Sede Palmira, International Center for Tropical Agriculture | Beatriz Parra; Diana López-Alvarez; Wilmer J. Cuellar |
| EPI_ISL_529963, EPI_ISL_759958 | Universitas Airlangga Hospital | Institute of Tropical Disease, Universitas Airlangga | Aldise M Nastri; Gatot Soegiarto; Jezzy R Dewantari; Kazufumi Shimizu; Krisnoadi Rahardjo; Laksmi Wulandari; Maria I Lusida; Mitsuhiro Nishimura; Nasronudin; Resti Yudhawati; Retno A Setyoningrum; Rima R Prasetya; Soetjipto; Yasuko Mori; Yohko K Shimizu |
| EPI_ISL_461769 | University College London, Great Ormond Street Hospital for Children NHS Foundation Trust, Imperial College Healthcare NHS Trust | COVID-19 Genomics UK (COG-UK) Consortium | Alison Holmes; Charlotte Williams; Helena Tutill; Jacqueline Findlay; James Price; Judith Breuer; Julianne Brown; Kathryn Harris; Leysa Forrest; Mark Kristiansen; Paola Niola; Paola Resende Silva; Patricia Dyal; Paul Randell; Rachel Williams; Sam Weeks; Sergi Castellano; Sunando Roy; Tony Brooks; Yasmin Panchbhaya |
| EPI_ISL_418280 | University Hospital Basel, Clinical Virology | University Hospital Basel, Clinical Bacteriology | Egli, A.; Hirsch, H.; Leuzinger, K.; Mari, A.; Roloff, T.; Seth-Smith, H. |
| EPI_ISL_710548, EPI_ISL_710549, EPI_ISL_710551, EPI_ISL_710555, EPI_ISL_710562, EPI_ISL_710564, EPI_ISL_710566, EPI_ISL_710567 | | | |
| see above | University Hospital Dubrava | Ruđer Boškovic Institute; Forensic Science Centre Ivan Vučetić; University of Zagreb Faculty of Science | Ana Livun; Antonela Blažeković; Boris Maček; Danilo Licastro; Dunja Glavaš; Fran Borovečki; Fuad Ćosović; Gordana Maravić Vlahoviček; Ivan Šamija; Ivana Ćelap; Jasna Kašman; Josipa Skelin; Katarina Marija Tupek; Kristian Vlahoviček; Kristina Gotovac Jerećić; Lidija Cvetko-Krajinović; Lucija Basić; Lucija Markulin; Maja Kuzman; Marina Korolija; Mario Štefanović; Mirjana Domazet-Lošo; Paula Štancl; Petra Vrabec; Robert Belužić; Rosa Karlić; Sanja Tadinac; Senčica Pejša; Tomislav Domazet-Lošo; Valentina Đumlijan-Combaj; Vjekoslav Tomaić; Vladimir Krajinović; Željka Maček Šafranko |
| EPI_ISL_483676 | University Hospital Zurich | Department of Biosystems Science and Engineering, ETH Zürich | Christian Beisel; Elodie Burcklen; Ina Nissen; Ivan Topolsky; Julia Martinez-Gomez; Mitch Levesque; Natascha Santacroce; Niko Beerenwinkel; Pedro Ferreira; Phil Cheng; Philipp Bosshard; Philipp Jablonski; Sarah Nadeau; Susana Posada-Céspedes; Tanja Stadler; Tobias Schär |
| EPI_ISL_454578, EPI_ISL_454583 | University Hospital for Infectious Diseases "Dr. Fran Mihaljević", Research Unit | University of Zagreb, Centre for research and knowledge transfer in biotechnology | Anamarija Slovic; Ivan-Christian Kurolt; Jelena Ivancic Jelecki |
| EPI_ISL_590944, EPI_ISL_708068, EPI_ISL_775445, EPI_ISL_796708 | University Hospital of Northern Norway, Department for Microbiology and Infectious Disease Control | Norwegian Institute of Public Health, Department of Virology | Atiya R Ali; Hilde Elshaug; Hilde Vollan; Kamilla Heddeland Instefjord; Karoline Bragstad; Kathrine Stene-Johansen; Marie Paulsen Madsen; Olav Hungnes; Rasmus Riis Kopperud |
| EPI_ISL_429220 | University Hospitals of Geneva Laboratory of Virology | University Hospitals of Geneva Laboratory of Virology | Laubscher F. |
| EPI_ISL_776007, EPI_ISL_776246, EPI_ISL_776359, EPI_ISL_776513, EPI_ISL_776524, EPI_ISL_776539, EPI_ISL_776542 | | | |
| see above | University Medical Center Hamburg Eppendorf | Heinrich Pette Institute, Leibniz Institute for Experimental Virology | Adam Grundhoff; Alexis Robitaille; Johannes Knobloch; Martin Aepfelbacher; Nicole Fischer; Thomas Günther |
| EPI_ISL_730652 | University of Bari, Valenzano, Italy | Dept. Food safety, Nutrition and Veterinary Public Health, Istituto superiore di sanità | C. Buonavoglia; C. Desario; D. Buonavoglia, V.; E. Lorusso; G. Elia; G. Vaccari; I. Di Bartolo; L. De Sabato; Martella; N. Decaro; U. Agrimi |
| EPI_ISL_476075, EPI_ISL_477010, EPI_ISL_477013, EPI_ISL_671444, EPI_ISL_671493 | University of Debrecen, Department of Medical Microbiology | National Laboratory of Virology, Szentágothai Research Centre | Balázs Somogyi; Brigitta Zana; Endre Gábor Tóth; Eszter Csoma; Ferenc Jakab; Gábor Kemenesi |
| EPI_ISL_455682 | University of Florida | University of Florida | C.-Y.; J.A.; J.G.; Lauzardo; Lednicky; M. and Morris; Wu |
| EPI_ISL_495593 | University of Michigan Clinical Microbiology Laboratory | Lauring Lab, University of Michigan, Department of Microbiology and Immunology | Valesano et al. |
| EPI_ISL_477615, EPI_ISL_677744, EPI_ISL_677795, EPI_ISL_677815 | University of Szeged, Institute of Clinical Microbiology | National Laboratory of Virology, Szentágothai Research Centre | Balázs Somogyi; Brigitta; Brigitta Zana; Endre Gábor Tóth; Ferenc Jakab; Gabriella Terhes; Gábor Kemenesi; Terhes Gabriella |
| EPI_ISL_485391, EPI_ISL_485392 | University of Ulsan College of Medicine and Asan Medical Center | University of Ulsan College of Medicine and Asan Medical Center | Heungsup Sung; Jaewoong Lee; Jina Lee; Jiwon Jung; Jongsik Chun; Kihyun Lee; Kuenyoul Park; Kyu-Hwa Hur; Mauricio Chalita; Mi-Na Kim; Seok-Hwan Yoon; Sung-Han Kim; and Hae Kyung Lee |
| EPI_ISL_677600, EPI_ISL_728040, EPI_ISL_791123, EPI_ISL_791159 | University of Wisconsin-Madison AIDS Vaccine Research Laboratories | University of Wisconsin-Madison AIDS Vaccine Research Laboratories | Gage Moreno; Katarina Braun; et al. AIDS Vaccine Research Laboratories |
| EPI_ISL_693234 | Upa Vereador Jose Da Rocha Goncalves | Instituto Adolfo Lutz, Interdiciplinary Procedures Center, Strategic Laboratory | Claudia Regina Gonçalves; Claudio Tavares Sacchi; Erica Valessa Ramos Gomes; Karoline Rodrigues Campos |
| EPI_ISL_450812, EPI_ISL_475517 | Uppsala Narakut Aleris | The Public Health Agency of Sweden | Anna Risberg; Anna-Malin Linde; Annika Nilsson; Karin Tegmark-Wisell; Maria Lind Karlberg; Mattias Haukland; Mia Brytting; Olov Svartstrom; Oskar Karlsson Lindsjo; Reza Advani; Sandra Broddesson; Theresa Enkirch |
| EPI_ISL_582779, EPI_ISL_676528 | Uppsala klinisk mikrobiologi | The Public Health Agency of Sweden | Anna Risberg; Anna-Malin Linde; Department of Microbiology; Karin Tegmark-Wisell; Maria Lind Karlberg; Mattias Haukland; Mia Brytting; Olov Svartstrom; Oskar Karlsson Lindsjo; Petra Edquist; Reza Advani; Sandra Broddesson; The Public Health Agency of Sweden |
| EPI_ISL_708815, | Urban Institute for Disease Prevention and Control | National Institute of Health, Department of Medical Sciences, Ministry of Public Health, Thailand | Malinee Chittaganpitch; Pakorn Piromtong; Pilailuk Okada; Siripaporn Phuygun; Sittiporn Parnmen; Sunthareeya Waicharoen; Thanutsapa Thanadachakul; Warawan Wongboot |

| | | | |
|---|---|---|---|
| EPI_ISL_708816, EPI_ISL_708817 | | | |
| EPI_ISL_582527, EPI_ISL_582528, EPI_ISL_582533, EPI_ISL_644572 | Veterinary Specialized Institute "Kraljevo", Serbia | Veterinary Specialized Institute "Kraljevo", Serbia | Afonso, C.; Banovic Djeri, B.; Jankovic, M.; Jovanovic, T.; Knezevic, A.; Petrovic, T.; Sekler, M.; Tesovic, B.; Vidanovic, D.; Volkening, J. |
| EPI_ISL_678486 | Veterinary Specialized Institute "Sabac", Serbia | Veterinary Specialized Institute "Kraljevo", Serbia | Afonso, C.; Banovic Djeri, B.; Jankovic, M.; Jovanovic, T.; Knezevic, A.; Mrkovacki, S.; Petrovic, T.; Sekler, M.; Tesovic, B.; Vidanovic, D.; Volkening, J. |
| EPI_ISL_708823 | Vibharam Hospital | National Institute of Health, Department of Medical Sciences, Ministry of Public Health, Thailand | Malinee Chittaganpitch; Pakorn Piromtong; Pilailuk Okada; Siripaporn Phuygun; Sittiporn Parnmen; Sunthareeya Waicharoen; Thanutsapa Thanadachakul; Warawan Wongboot |
| EPI_ISL_456429, EPI_ISL_456538, EPI_ISL_456550, EPI_ISL_456581 | Victorian Infectious Diseases Reference Laboratory (VIDRL) | Microbiological Diagnostic Unit Public Health Laboratory and Victorian Infectious Diseases Reference Laboratory, Doherty Institute | Caly L.; Druce J.; Sait, M.; Schultz M.; Seemann T.; Sherry, N. |
| EPI_ISL_480665, EPI_ISL_480740, EPI_ISL_521879, EPI_ISL_562705 | Victorian Infectious Diseases Reference Laboratory (VIDRL) | VIDRL and MDU-PHL | Caly L.; Caly, L.; Druce J.; M. B.; Sait, M.; Schultz; Schultz M.; Seemann T.; Seemann, T.; Sherry, N. |
| EPI_ISL_416411, EPI_ISL_419749, EPI_ISL_419857 | Victorian Infectious Diseases Reference Laboratory (VIDRL) | Victorian Infectious Diseases Reference Laboratory and Microbiological Diagnostic Unit Public Health Laboratory, Doherty Institute | Caly L.; Druce J.; Sait, M.; Schultz M.; Seemann T.; Sherry, N.; Taiaroa, G. |
| EPI_ISL_468313, EPI_ISL_468319, EPI_ISL_603033 | Vigilancia Epidemiologica de São Bernardo do Campo | Instituto Adolfo Lutz, Interdiciplinary Procedures Center, Strategic Laboratory | Claudia Regina Gonçalves; Claudio Tavares Sacchi; Erica Valessa Ramos Gomes; Karoline Rodrigues Campos |
| EPI_ISL_515542 | Vigilância Epidemiológica de Leme | Instituto Adolfo Lutz, Interdiciplinary Procedures Center, Strategic Laboratory | Claudia Regina Gonçalves; Claudio Tavares Sacchi; Erica Valessa Ramos Gomes |
| EPI_ISL_603023 | Vigilância em Saúde Visa Sul | Instituto Adolfo Lutz, Interdiciplinary Procedures Center, Strategic Laboratory | Claudia Regina Gonçalves; Claudio Tavares Sacchi; Erica Valessa Ramos Gomes; Karoline Rodrigues Campos |
| EPI_ISL_547573, EPI_ISL_583493 | Vigilância em Saúde de Cajamar | Instituto Adolfo Lutz, Interdiciplinary Procedures Center, Strategic Laboratory | Claudia Regina Gonçalves; Claudio Tavares Sacchi; Erica Valessa Ramos Gomes; Karoline Rodrigues Campos |
| EPI_ISL_802852, EPI_ISL_802853, EPI_ISL_802857, EPI_ISL_802860, EPI_ISL_802861 | Vilnius University Hospital Santaros Klinikos, Vilnius University | Institute of Biotechnology, Life Sciences Center, Vilnius University | Albertas Timinskas; Alma Gedvilaite; Aurelija Zvirbliene; Daniel Naumovas; Emilija Vasiliunaite; Laimonas Griskevicius; Milda Norkiene |
| EPI_ISL_560396, EPI_ISL_560399, EPI_ISL_560405 | Vilnius University Hospital Santaros Klinikos, Vilnius University | Institute of Biotechnology, Life Sciences Center, Vilnius University and Thermo Fisher Scientific | Albertas Timinskas; Alma Gedvilaite; Aurelija Zvirbliene; Daniel Naumovas; Justinas Slikas; Laimonas Griskevicius; Ligita Jancioriene; Mindaugas Paulauskas |
| EPI_ISL_451749, EPI_ISL_466939, EPI_ISL_466960, EPI_ISL_467010, EPI_ISL_468232, EPI_ISL_476084, EPI_ISL_476093, EPI_ISL_476094, EPI_ISL_476112, EPI_ISL_476123, EPI_ISL_483657, EPI_ISL_486510, EPI_ISL_486534, EPI_ISL_486538, EPI_ISL_498626, EPI_ISL_510720, EPI_ISL_510762, EPI_ISL_510783, EPI_ISL_516588, EPI_ISL_523858, EPI_ISL_535615, EPI_ISL_535630, EPI_ISL_539468, EPI_ISL_560440, EPI_ISL_560513, EPI_ISL_603288, EPI_ISL_603338, EPI_ISL_603549, EPI_ISL_614904, EPI_ISL_693806, EPI_ISL_693889, EPI_ISL_693982, EPI_ISL_721748, EPI_ISL_721836, EPI_ISL_721923, EPI_ISL_721971, EPI_ISL_737676, EPI_ISL_767984, EPI_ISL_796400, EPI_ISL_796469 | | | |
| see above | Viollier AG | Department of Biosystems Science and Engineering, ETH Zürich | Andrea Patrignani; Andreia Cabral de Gouvea; Catharine Aquino; Chaoran Chen; Christian Beisel; Christiane Beckmann; Christoph Noppen; David Dreifuss; Doris Popovic; Elodie Burcklen; Griffin White; Ina Nissen; Ivan Topolsky; Jay Tracy; Katharina Jahn; Lara Fuhrmann; Laura Neff; Lennart Opitz; Maria Domenica Moccia; Maurice Redondo; Natascha Santacroce; Niko Beerenwinkel; Noemie Santamaria de Souza; Olivier Kobel; Pedro Ferreira; Philipp Jablonski; Ralph Schlapbach; Sarah Nadeau; Simon Grüter; Sophie Seidel; Susana Posada-Céspedes; Tanja Stadler; Timothy Sykes; Tobias Schär |
| EPI_ISL_417441, EPI_ISL_420035, EPI_ISL_420849, EPI_ISL_429258, EPI_ISL_435163, EPI_ISL_437340, EPI_ISL_447249, EPI_ISL_513610, EPI_ISL_513613, EPI_ISL_514122, EPI_ISL_527552, EPI_ISL_591086 | | | |
| see above | Viral Respiratory Lab, National Institute for Biomedical Research (INRB) | Pathogen Sequencing Lab, National Institute for Biomedical Research (INRB) | Allison Black; Amuri Aziza; Andrew Rambaut; Catherine Pratt; Eddy Kinganda-Lusamaki; Edith Nkwembe; Emmanuel Lokilo Lofiko; Francisca Muyembe Mawete; Ian Goodfellow; James Hadfield; Jean Claude Makangara; Jean-Jacques Muyembe Tamfum; Josh Quick; Kristian Andersen; Matthias Pauthner; Michael Wiley; Nick Loman; Placide Mbala-Kingebeni; Steve Ahuka-Mundeke; Trevor Bedford |
| EPI_ISL_416488 | ViroGenetics - BSL3 Laboratory of Virology; Human Genome Variation Research Group & Genomics Centre MCB; Bioinformatics Research Group Department of Virology | ViroGenetics - BSL3 Laboratory of Virology; Human Genome Variation Research Group & Genomics Centre MCB; Bioinformatics Research Group Department of Virology | Adrianna Klajmon; Agata Jarosz; Agnieszka Kołakowska-Kulesza; Aleksandra Milewska; Ewelina Hallman-Szelińska; Ewelina Pośpiech; Kamila Marszałek; Katarzyna Pancer; Katarzyna Zacharczuk; Krzysztof Pyrć; Magdalena Rzeczkowska; Natalia Wolaniuk; Paweł P Łabaj; Tomasz Wołkowicz; Wojciech Branicki |
| EPI_ISL_428927 | ViroGenetics - BSL3 Laboratory of Virology; Human Genome Variation Research Group & Genomics Centre MCB; Bioinformatics Research Group; Wojewódzka Stacja Sanitarno-Epidemiologiczna w Krakowie | ViroGenetics - BSL3 Laboratory of Virology; Human Genome Variation Research Group & Genomics Centre MCB; Bioinformatics Research Group | Adrianna Klajmon; Agnieszka Starowicz; Aleksandra Pisarek; Aleksandra Synowiec; Danuta Piniewska-Róg; Ewelina Pośpiech; Inga Drebot; Jarosław Foremny; Kamila Marszałek; Katarzyna Dudek; Katarzyna Guła; Katarzyna Kopera; Krzysztof Pyrć; Magda Pachota; Marek Sanak; Michał Kowalski; Paweł P Łabaj; Tomasz Gromowski; Wojciech Branicki |
| EPI_ISL_418183, EPI_ISL_435417 | Virological Research Group, Szentágothai Research Centre | Bioinformatics Research Group, Szentágothai Research Centre | Attila Gyenesei; Endre Gábor Tóth; Ferenc Jakab; Gábor Kemenesi; Péter Urbán; Róbert Herczeg |
| EPI_ISL_487766, EPI_ISL_532994 | Virology Department, Royal Infirmary of Edinburgh, NHS Lothian / School of Biological Sciences, University of Edinburgh | Wellcome Sanger Institute for the COVID-19 Genomics UK (COG-UK) consortium | Colquhoun R; Cordelia Langford; David K. Jackson; Dewar R; Dominic Kwiatkowski; Ewan Harrison; Hill V; Ian Johnston; Jackson B; John Sillitoe; John Sillitoe on behalf of the Wellcome Sanger Institute COVID-19 Surveillance Team (http://www.sanger.ac.uk/covid-team); McCrone JT; McHugh M; O'Toole Á; Rambaut A; Roberto Amato; Rooke S; Scher E; Sonia Goncalves; Templeton K and Alex Alderton; Yu X |
| EPI_ISL_425836, EPI_ISL_439267, EPI_ISL_453144, EPI_ISL_473833, EPI_ISL_473910, EPI_ISL_473947, EPI_ISL_493970, EPI_ISL_514555, EPI_ISL_526482, EPI_ISL_652006, EPI_ISL_652026, EPI_ISL_652062, EPI_ISL_680461, EPI_ISL_724242, EPI_ISL_764363 | | | |
| see above | Virology Department, Royal Infirmary of Edinburgh, NHS Lothian / School of Biological Sciences, University of Edinburgh / Institute of Genetics and Molecular Medicine, University of Edinburgh | COVID-19 Genomics UK (COG-UK) Consortium | Balcaza C; Colquhoun R; Dewar R; Gallagher M; Hill V; Jackson B; McCrone JT; McHugh M; O'Toole A; O'Toole Á; O,ÄôToole √Ä; Rambaut A; Rooke S; Scher E; Templeton K; Williams TC; Yu X |
| EPI_ISL_453772, EPI_ISL_540722, EPI_ISL_573763 | Virology Department, Sheffield Teaching Hospitals NHS Foundation Trust/Department of Infection, Immunity and Cardiovascular Disease, The Medical School, University of Sheffield | COVID-19 Genomics UK (COG-UK) Consortium | Adri Angyal; Alex Keeley; Benjamin Lindsey; Cariad Evans; Danielle Groves; Dave Partridge; Katie Johnson; Laura Carrilero; Luke Green; Matthew Parker; Matthew Wyles; Mehmet Yavuz; Mohammad Raza; Nikki Smith; Paul Parsons; Rachel Tucker; Rebecca Brown; Thushan de Silva |
| EPI_ISL_493206 | Virology Lab,Department of Pathology, National Cheng Kung University Hospital | Virology Lab,Department of Pathology, National Cheng Kung University Hospital | Huey-Pin Tsai; et al |
| EPI_ISL_508863, EPI_ISL_625456, EPI_ISL_677634, EPI_ISL_677635 | Virology Unit, Institut Pasteur de Madagascar | Virology Unit, Institut Pasteur de Madagascar | Cara Brook; Cara E. Brook; Christian Ranaivoson; Cristina M. Tato; Helisoa Razafimanjato; Jean-Michel Heraud; Joseph L. DeRisi; Michelle Tan; Norosoa Razanajatovo; Philippe Dussart; Soa Fy Andriamandimby; Tsiry Randriambolamanantsoa; Vida Ahyong; Vololoniaina Raharinosy |
| EPI_ISL_411902 | Virology Unit, Institut Pasteur du Cambodge. | Virology Unit, Institut Pasteur du Cambodge (Sequencing done by: Jessica E Manning/Jennifer A Bohl at Malaria and Vector Research Research Laboratory, National Institute of Allergy and Infectious Diseases and Vida Ahyong from Chan-Zuckerberg Biohub) | Erik A Karlsson; Jennifer A Bohl; Jessica E Manning.; Philippe Dussart; Veasna Duong; Vida Ahyong |
| EPI_ISL_447627 | Virology, Wageningen Bioveterinary Research | Virology, Wageningen Bioveterinary Research | Hakze van der Honing; Harders, F.; R.W.; W.H.M.; van der Poel |
| EPI_ISL_745189 | Vredenburg Hospital wc VBG | National Health Laboratory Service (NHLS), Tygerberg | Bronwyn Kleinhans; Eduan Wilkindon; Gert van Zyl; Houriiyah Tegally; Kayla Delaney; Susan Engelbrecht; Tulio de Oliveira; Wolfgang Preiser |
| EPI_ISL_430070, EPI_ISL_430110, EPI_ISL_450283, EPI_ISL_524000, EPI_ISL_524045, EPI_ISL_596230, EPI_ISL_596267, EPI_ISL_732985, EPI_ISL_733284, EPI_ISL_733466 | | | |
| see above | WHO National Influenza Centre Russian Federation | WHO National Influenza Centre Russian Federation | Andrey Komissarov; Anna Ivanova; Artem Fadeev; Daria Danilenko; Dmitry Bazhenov; Dmitry Lioznov; Elena Nabieva; Georgii Bazykin; Ksenia Komissarova; Ksenia Safina; Kseniya Komissarova; Mariia Pisareva; Mariia Sergeeva; Mariia Timofeeva; Tamila Musaeva; Veronica Eder |
| EPI_ISL_676843, EPI_ISL_767470 | Wadsworth Center, New York State Department.of Health | Wadsworth Center, New York State Department.of Health | Alexis Russel; Daryl M. Lamson; Erasmus Schneider; Erica Lasek-Nesselquist; John Kelly; Jonathan Plitnick; Kirsten St. George; Matthew Shudt; Melissa A Leisner; Navjot Singh; Sara Griesemer |
| EPI_ISL_649124 | Waikato Hospital | Institute of Environmental Science and Research (ESR) | Anja Werno; Antje van der Linden; Arlo Upton; Chris Mansell; David Hammer; Dragana Drinkovic; Erasmus Smit; Gary McAuliffe; Hana Sofia Andersson; Hermes Perez; James Ussher; Jill Sherwood; Jing Wang; Joep de Ligt; Josh Freeman; Julia Howard; Juliet Elvy; Lauren Jelly; Mary DeAlmeida; Matt Blakiston; Matt Storey; Matthew Rogers; Max Bloomfield; Michael Addidle; Michelle Balm; Muhammad Faisal; Nikki Freed; Olin Silander; Sally Roberts; Sarah Jefferies; Sharmini Muttaiyah; Susan Morpeth; Susan Taylor; Timothy Blackmore; Vani Sathyendran; Veronica Playle; Virginia Hope; Xiaoyun Ren |
| EPI_ISL_479863 | Wakayama Prefectural Research Center of Environment and Public Health | Pathogen Genomics Center, National Institute of Infectious Diseases | Fumio Terasoma; Hajime Kamiya; Kentaro Itokawa; Makoto Kuroda; Masanori Hashino; Motoi Suzuki; Rina Tanaka; Tsuyoshi Sekizuka; Yosuke Hamajima |
| EPI_ISL_413555, EPI_ISL_413556, EPI_ISL_418158, EPI_ISL_420970, EPI_ISL_422097, EPI_ISL_445639 | Wales Specialist Virology Centre | Public Health Wales Microbiology Cardiff | Alec Birchley; Alexander Adams; Amy Gaskin; Bree Gatica-Wilcox; Catherine Moore; Cen Sabu; Jason Coombes; Joanne Watkins; Johnathan Evans; Laura Gifford; Lauren Gilbert; Lee Graham; Malorie Perry; Matt Bull; Matthew Bull; Nicole Pacchiarini; Sally Corden; Sara Kumziene-Summerhayes; Sara Rey; Sarah Taylor; Simon Cottrell; Simon Cottrell Sara Rey; Sophie Jones; Tim Jones; Tom Connor |
| EPI_ISL_472851, EPI_ISL_472889, EPI_ISL_473039, EPI_ISL_473088, EPI_ISL_473149, EPI_ISL_474009, EPI_ISL_474156, EPI_ISL_474265, EPI_ISL_474360, EPI_ISL_490733, EPI_ISL_499381, EPI_ISL_499425, EPI_ISL_512519, EPI_ISL_512533, EPI_ISL_535197, EPI_ISL_574239, EPI_ISL_577436, EPI_ISL_595470, EPI_ISL_612926, EPI_ISL_639075, EPI_ISL_639596, EPI_ISL_680768, EPI_ISL_680923, EPI_ISL_727010, EPI_ISL_727453, EPI_ISL_742484, EPI_ISL_743564, EPI_ISL_764726 | | | |

| | | | |
|---|---|---|---|
| see above | Wales Specialist Virology Centre Sequencing lab: Pathogen Genomics Unit | COVID-19 Genomics UK (COG-UK) Consortium | Alec Birchley; Alexander Adams; Amy Gaskin; Angela Marchbank; Bree Gatica-Wilcox; Catherine Moore; Jason Coombes; Joanne Watkins; Joel Southgate; Johnathan Evans; Laura Gifford; Lauren Gilbert; Lee Graham; Malorie Perry; Matthew Bull; Nicole Pacchiarini; Sally Corden; Sara Kumziene-Summerhayes; Sara Rey; Sarah Taylor; Simon Cottrell; Sophie Jones; Tom Connor |
| EPI_ISL_417096, EPI_ISL_496213 | Washington State Department of Health | Seattle Flu Study | Amanda Adler; Barry R. Lutz; Benjamin Pelle; Brian Hiatt; Caitlin R. Wolf; Chris D. Frazar; Chu etl al; Deborah A. Nickerson; Elisabeth Brandstetter; Geoff Melly; Helen Y. Chu; Janet A. Englund; Jay Shendure; Jover Lee; Kairsten Fay; Kirsten Lacombe; Lea M. Starita; Mark J. Rieder; Matthew Richardson; Matthew Thompson; Melissa Truong; Michael Boeckh; Michael Famulare; Misja Ilcisin; Peter D. Han; Philip Dykema; Romesh Gautom; Scott Lindquist; Thomas R. Sibley; Trevor Bedford |
| EPI_ISL_451318, EPI_ISL_451322, EPI_ISL_451371, EPI_ISL_451387, EPI_ISL_451393 | West China Hospital of Sichuan University | State Key Laboratory of Biotherapy of Sichuan University | Baowen Du; Binwu Ying; Chao Tang; Chuan Chen; Hancheng Wei; Jia Geng; Jing-wen Lin; Lu Chen; Mingxia Yu; Minjin Wang; Weimin Li; Yongzhao Zhou |
| EPI_ISL_425795, EPI_ISL_425812, EPI_ISL_433657, EPI_ISL_453074, EPI_ISL_490697, EPI_ISL_490709, EPI_ISL_499354, EPI_ISL_705731 | | | |
| see above | West of Scotland Specialist Virology Centre, NHSGGC / MRC-University of Glasgow Centre for Virus Research | COVID-19 Genomics UK (COG-UK) Consortium | Alasdair MacLean; Alice Broos; Ana da Silva Filipe; Antonia Ho; Daniel Mair; David L Robertson; Elihu Aranday-Cortes; Emma Thomson; Guy Mollett; James Shepherd; Jenna Nichols; Joseph Hughes; Kathy Li; Kathy Smollett; Kirstyn Brunker; Kyriaki Nomikou; Lily Tong; Marc Niebel; Matthew Holden; Natasha Jesudason; Natasha Johnson; Patawee Asamaphan; Rachel Blacow; Rajiv Shah; Richard Orton; Rory Gunson; Rory Gunson.; Sarah McDonald; Sharif Shaaban; Sreenu Vattipally; Stephen Carmichael; Yasmin Parr |
| EPI_ISL_566036, EPI_ISL_566041 | Wildlife Conservation Society, Bronx Zoo | Diagnostic Virology Laboratory, United States Department of Agriculture, National Veterinary Services Laboratories, Population Medicine and Diagnostic Sciences, Cornell University | B.D.; Bartlett; Calle; Cardia Caserta, L.; Caserta; Cronk; D.C.; D.G.; Delaney; Diel; Elvinger, F.; Fang, Y.; Farnzen, K.; Frederikson, R.; Goodman, L.; Goodrich, E.; Hines Bergeson, N.; Ingerman, K.; Ivancic, M.; J.M.; Jenkins-Moore, M.; Killian; Kuzmin, I.; L.C.; Laverack, M.; M.A.; M.K.; M.L.; M.R.; Mauldin; McAloose, D.; McCann, C.; Mitchell; Mozingo, K.; Olmstead, C.; P.K.; P.P.; Queen, K.; S.L.; Slavinski, S.; Smith; Stokol, T.; Sykes; Terio, K.; Thomas, P.; Tong, S.; Torchetti; Wang, H.; Wang, L.; Yuan, F.; Zec, S. |
| EPI_ISL_408670 | Wisconsin Department of Health Services | Pathogen Discovery, Respiratory Viruses Branch, Division of Viral Diseases, Centers for Dieases Control and Prevention | Anna Uehara; Brett L. Whitaker; Brian Lynch; Clinton R. Paden; Janna' R. Murray; Jing Zhang; Krista Queen; Lijuan Wang; Senthil Kumar K. Sakthivel; Shifaq Kamili; Stephen Lindstrom; Susan I. Gerber; Suxiang Tong; Xiaoyan Lu; Yan Li; Ying Tao |
| EPI_ISL_803342, EPI_ISL_803576, EPI_ISL_803837 | Wisconsin State Laboratory of Hygiene Communicable Disease Division | Wisconsin State Laboratory of Hygiene Communicable Disease Division | Abigail C. Shockey; Kelsey R. Florek |
| EPI_ISL_708812 | World Medical Hospital | National Institute of Health, Department of Medical Sciences, Ministry of Public Health, Thailand | Malinee Chittaganpitch; Pakorn Piromtong; Pilailuk Okada; Siripaporn Phuygun; Sittiporn Parnmen; Sunthareeya Waicharoen; Thanutsapa Thanadachakul; Warawan Wongboot |
| EPI_ISL_454918, EPI_ISL_454919, EPI_ISL_454921, EPI_ISL_454931, EPI_ISL_454949, EPI_ISL_454956, EPI_ISL_454976, EPI_ISL_455381, EPI_ISL_455386 | | | |
| see above | Wuhan Chain Medical Labs (CMLabs) | State Key Laboratory of Biotherapy of Sichuan University | Baowen Du; Binwu Ying; Chao Tang; Chuan Chen; Hancheng Wei; Jia Geng; Jing-wen Lin; Lu Chen; Mingxia Yu; Minjin Wang; Weimin Li; Yongzhao Zhou |
| EPI_ISL_514753 | Yaftabad Hospital, COVID Lab Center | University of Tabriz | Hashemian; Hosseinzadeh Gharajeh, N.; S.M. and Barati, O.; Shahabzadeh, Z. |
| EPI_ISL_408478 | Yongchuan District Center for Disease Control and Prevention | Chongqing Municipal Center for Disease Control and Prevention | Chen Shuang; Li Qing; Ling Hua; Rong Rong; Su Kun; Tan ZhangPing; Tang Wenge; Tang Yun; Ye Sheng; Yu zhen |
| EPI_ISL_539577, EPI_ISL_631307, EPI_ISL_631375, EPI_ISL_718073, EPI_ISL_733511 | ZOTZ KLIMAS MVZ Düsseldorf-Centrum GbR ÜBAG für Labormedizin, Genetik, Zytologie, Pathologie | Center of Medical Microbiology, Virology, and Hospital Hygiene, University of Duesseldorf | Alexander Dilthey; Andreas Walker; Ashley-Jane Duplessis; Daniel Strelow; Jessica Nicolai; Jörg Timm; Katrin Hoffmann; Klaus Pfeffer; Lisanna Hülse; Malte Kohns Vasconcelos; Marek Korencak; Maximilian Damagnez; Nadine Lübke; Patrick Finzer; Rainer Zotz; Tobias Wienemann; Torsten Houwaart |
| EPI_ISL_404228 | Zhejiang Provincial Center for Disease Control and Prevention | Department of Microbiology, Zhejiang Provincial Center for Disease Control and Prevention | Chen Chen; Chonggao Hu; Enfu Chen; Fang Xu; Haiyan Mao; Hanping Zhu; Hao Yan; Jian Gao; Jianmin Jiang; Junhang Pan; Juying Yan; Liming Gong; Qiong Ge; Wen Shi; Wenwu Yao; Xinying Wang; Xiuyu Lou; Yan Feng; Yanjun Zhang; Yi Sun; Yin Chen; Yisheng Sun; Yiyu Lu; Zhangnv Yang; Zhen Li; Zhen Wang; Zhiping Chen |
| EPI_ISL_408479 | Zhongxian Center for Disease Control and Prevention | Chongqing Municipal Center for Disease Control and Prevention | Chen Shuang; Li Qin; Ling Hua; Rong Rong; Su Kun; Tan ZhangPing; Tang Wenge; Tang Yun; Ye Sheng; Yu zhen; Zhang Hong |
| EPI_ISL_467430 | Zoonotic and Exotic infection Diseases Division | Zoontic and Exotic infection Diseases Division | Jinlang Wang; Zhigao Bu |
| EPI_ISL_459909 | Zoonotic and Exotic infection Diseases Division, Harbin Veterinary Research Institute, CAAS | Zoonotic and Exotic infection Diseases Division, Harbin Veterinary Research Institute, CAAS | Jinliang Wang; Zhigao Bu |
| EPI_ISL_660069, EPI_ISL_660070, EPI_ISL_681703 | Zurita & Zurita Laboratorios | Zurita & Zurita Laboratorios | Camilo Zurita-Salinas; Gabriela Sevillano; Gabriela Sevillano Camilo Zurita-Salinas Karen Loaiza David Ortega-Paredes Jeannete Zurita; Jeannete Zurita |
| EPI_ISL_468135, EPI_ISL_468144 | [Romania, Bucharest] National Institute for Infectious Diseases “Prof. Dr. Matei Balş” | [Romania, Bucharest] National Institute for Infectious Diseases “Prof. Dr. Matei Balş” | Corina Casangiu; Leontina Banica; Marius Cotic; Marius Surleac; Simona Paraschiv |
| EPI_ISL_447913, EPI_ISL_447920 | n/a | National Institute of Health. Department of medical Sciences, Ministry of Public Health, Thailand | Chittaganpitch; Malinee; Okada; Parnmen; Phuygun; Pilailuk; Siripaporn; Sittiporn; Sunthareeya; Thanadachakul; Thanutsapa; Waicharoen; Warawan; Wongboot |
| EPI_ISL_450500 | unknown | CAS Key Laboratory of Special Pathogens and Biosafety and Center for Emerging Infectious Diseases | Lin, H.; Shi, Z.; Si, H.; Xie, S.; Zhou, P.; Zhu, Y. |
| EPI_ISL_486427, EPI_ISL_486429 | unknown | Clinical Laboratory, Hospital Israelita Albert Einstein | Amgarteb, D.; Amgarten, D.; C.L. and Pinho; F.G.; Guedes; J.R.; Malta, F.; Mangueira; R.A.; R.L.; Santana; de Menezes |
| EPI_ISL_463005 | unknown | Clinical virology | Fares, W.; Triki, H. |
| EPI_ISL_487272, EPI_ISL_487273 | unknown | Communicable Disease Laboratory, Public Health Directorate | AlWasti, H.; Altaif, Z.; Altaif, z.; Shehab, F.; Zaed, A. |
| EPI_ISL_455687, EPI_ISL_455689 | unknown | Department of Microbiology | Bao, L.; Cai, F.; Deng, W.; Gao, H.; Gao, Q.; Ge, X.; Gong, X.; Hu, Y.; Jiang, D.; Kan, B.; Li, C.; Li, J.; Li, Y.; Liu, J.; Lou, X.; Lu, J.; Lv, Z.; Mao, H.; Qin, C.; Shi, W.; Sun, Y.; Wang, L.; Wang, N.; Wang, X.; Wu, D.; Xu, K.; Yang, M.; Yin, W.; Yin, Y.; Zhang, H.; Zhang, Y.; Zhu, L. |
| EPI_ISL_449481 | unknown | Department of Respiratory and Critical Care | Bai, X.; Chang, Y.; Deng, T.; Gao, Y.; He, Y.; Jiang, N.; Liang, L.; Liu, L.; Liu, W.; Ma; Ma, L.; Ma, X.; Pan, T.; Shi, H.; W.L. and Gao, Z.; Wang, G.; Wang, X.; Wei, X.; Xu, Y.; Yang, B.; Yang, D.; Zhai, B.; Zhang, J.; Zhang, Z.; Zheng, Y.; Zhou, Q. |
| EPI_ISL_437616, EPI_ISL_437621 | unknown | Faculty of Medicine | Buathong, R.; Bunprakob, S.; Ghai, S.; Joyjinda, Y.; Mungaomklang, A.; Petcharat, S.; Plipat; Prasithsirikul, W.; Rodpan, A.; Sirichan, N.; T. and Hemachudha, T.; Wacharapluesadee, S. |
| EPI_ISL_476559 | unknown | Laboratoire Sciences et Technologies de la Santé (STS) Institut Supérieur des Sciences de la Santé Université Hassan 1er, Settat, Morocco | Abderraouf Hilali; Amal Souiri; Hajar Lemriss; Jawad Bouzid; Mustapha Mouallif; Narjis Amar; Sanaâ Lemriss; Saâd EL Kabbaj; Touria Essayagh |
| EPI_ISL_462436 | unknown | Laboratory Diagnostic | Afonso; Banovic Djeri, B.; C.L.; Dmitric, M.; Jankovic, M.; Knezevic, A.; Petrovic, T.; Sekler, M.; Tesovic, B.; Vidanovic, D.; Volkening, J. |
| EPI_ISL_450408 | unknown | Microbiology | K.-Y.; K.K.W.; To; Yuen |
| EPI_ISL_434697 | unknown | National Institute of Health. Department of medical Sciences, Ministry of Public Health, Thailand | Chittaganpitch; Malinee; Okada; Parnmen; Phuygun; Pilailuk; Siripaporn; Sittiporn; Sunthareeya; Thanadachakul; Thanutsapa; Waicharoen; Warawan; Wongboot |
| EPI_ISL_413611 | unknown | Pathogen Discovery, Respiratory Viruses Branch, Division of Viral Diseases, Centers for Dieases Control and Prevention | Alison S Laufer Halpin; Anna Uehara; Christopher A. Elkins; Clinton R. Paden; Haibin Wang; Jasmine Padilla; Jing Zhang; Justin Lee; Krista Queen; Mary S. Keckler; Susan I. Gerber; Suxiang Tong; Yan Li; Ying Tao |
| EPI_ISL_414479 | unknown | Pathogen Discovery, Respiratory Viruses Branch, Division of Viral Diseases, Centers for Disease Control and Prevention | Alison S. Laufer Halpin; Anna Uehara; Christopher A. Elkins; Clinton R. Paden; Haibin Wang; Jasmine Padilla; Jing Zhang; Justin Lee; Krista Queen; Mary S. Keckler; Susan I. Gerber; Suxiang Tong; Yan Li; Ying Tao |
| EPI_ISL_450404 | unknown | School of Public Health, The University of Hong Kong | Brackman; C.J.; Chu; D.K.W.; D.N.C.; L.D.; L.L.M. and Peiris, M.; Perera; Poon; R.A.P.M.; Sims; Sit; T.H.S.; Tsang |
| EPI_ISL_483063 | unknown | Virology, Ecole Nationale Veterinaire de Toulouse | Bessiere, P.; Cadiergues; Croville, G.; Dubois, M.; Guerin; Izopet, J.; J.-L.; M.-C.; Walch, M. |
| EPI_ISL_458138, EPI_ISL_458139, EPI_ISL_458140, EPI_ISL_458141, EPI_ISL_458142, EPI_ISL_458143, EPI_ISL_458144, EPI_ISL_458145, EPI_ISL_458146, EPI_ISL_458147, EPI_ISL_458148, EPI_ISL_458149, EPI_ISL_524783, EPI_ISL_524784, EPI_ISL_524785, EPI_ISL_524786, EPI_ISL_524787, EPI_ISL_524788, EPI_ISL_524789, EPI_ISL_524790, EPI_ISL_524791, EPI_ISL_524792, EPI_ISL_524793, EPI_ISL_524794, EPI_ISL_524795, EPI_ISL_524796, EPI_ISL_524797, EPI_ISL_524798, EPI_ISL_524799, EPI_ISL_524800, EPI_ISL_524801 | | | |
| see above | Evandro Chagas Institute | Evandro Chagas Institute | A.M.; Barbagelata; E.C.; E.M.A.; Ferreira; G.M.R; H.R; J.A.; Junior; L.C.; L.S.; M.C.; Martins; P.S.; Resque; Santos; Silva; Sousa; Sousa Junior; Viana; W.D.C.; da Silva |
